# Safety and Immunogenicity of XBB.1.5-Containing mRNA Vaccines

**DOI:** 10.1101/2023.08.22.23293434

**Authors:** Spyros Chalkias, Nichole McGhee, Jordan L. Whatley, Brandon Essink, Adam Brosz, Joanne E. Tomassini, Bethany Girard, Kai Wu, Darin K. Edwards, Arshan Nasir, Diana Lee, Laura E. Avena, Jing Feng, Weiping Deng, David C. Montefiori, Lindsey R. Baden, Jacqueline M. Miller, Rituparna Das

## Abstract

**Background:** Subvariants of the severe acute respiratory syndrome coronavirus (SARS-CoV-2) omicron XBB-lineage have the potential to escape immunity provided by prior infection or vaccination. For Covid-19 immunizations beginning in the Fall 2023, the U.S. FDA has recommended updating to a monovalent omicron XBB.1.5-containing vaccine.

**Methods:** In this ongoing, phase 2/3 study participants were randomized 1:1 to receive 50-µg doses of mRNA-1273.815 monovalent (50-µg omicron XBB.1.5 spike mRNA) or mRNA-1273.231 bivalent (25-µg omicron XBB.1.5 and 25-µg omicron BA.4/BA.5 spike mRNAs) vaccines, administered as 5th doses, to adults who previously received a primary series and 3rd dose of an original mRNA coronavirus disease 2019 (Covid-19) vaccine, and a 4th dose of a bivalent (omicron BA.4/BA.5 and original SARS-CoV-2) vaccine. Interim safety and immunogenicity data 15 days post-vaccination are presented.

**Results:** In April 2023, participants received mRNA-1273.815 (n=50) and mRNA-1273.231 (n=51). The median intervals from the prior dose of BA.4/BA.5-containing bivalent vaccine were 8.2 and 8.3 months for the mRNA-1273.815 and mRNA-1273.231 groups, respectively. Both vaccines increased neutralizing antibody (nAb) geometric mean titers against all variants tested at day 15 post-booster nAb compared to pre-booster levels. Geometric mean fold-rises from pre-booster titers after the monovalent booster were numerically higher against XBB.1.5, XBB.1.16 and SARS-CoV-2 (D614G) than those of the bivalent booster and were comparable against BA.4/BA.5 and BQ1.1 variants for both vaccines. The monovalent vaccine also elicited nAb responses against omicron XBB.2.3.2, EG.5.1, FL.1.5.1 and BA.2.86 that were similar to those against XBB.1.5 in a subset (n=20) of participants. The occurrence of solicited adverse reactions and unsolicited adverse events were overall similar to those previously reported for the original mRNA-1273 50-µg and omicron BA.4/BA.5-containing bivalent mRNA-1273 vaccines.

**Conclusion:** In this interim analysis, XBB.1.5-containing monovalent and bivalent vaccines elicited potent neutralizing responses against variants of the omicron XBB-lineage (XBB.1.5, XBB.1.6, XBB.2.3.2, EG.5.1, and FL.1.5.1) as well as the recently emerged BA.2.86 variant. The safety profile of the XBB.1.5-containing vaccine was consistent with those of prior vaccines. These results overall indicate that the XBB.1.5-containing mRNA-1273.815 vaccine has the potential to provide protection against these emerging variants and support the Covid-19 vaccine update in 2023-2024 to a monovalent XBB.1.5-containing vaccine.

## Introduction

As severe acute respiratory syndrome coronavirus (SARS-CoV-2) XBB-subvariants evolved to escape immunity provided by prior infection or vaccination, the U.S. FDA has recommended updating to a monovalent omicron XBB.1.5-containing vaccine for immunizations beginning in the Fall 2023.^1^ Our ongoing, phase 2/3 open-label study randomized participants 1:1 to receive 50-µg doses of mRNA-1273.815 monovalent (50-µg omicron XBB.1.5 spike mRNA) or mRNA-1273.231 bivalent (25-µg omicron XBB.1.5 and 25-µg omicron BA.4/BA.5) spike mRNAs vaccines, administered as 5th doses, to adults who previously received a two-dose primary series, a 3rd dose of an original mRNA coronavirus disease 2019 (Covid-19) vaccine, and a 4th dose of a bivalent (omicron BA.4/BA.5 plus original strain) vaccine. Interim safety and immunogenicity data, 15 days post-vaccination with the updated investigational vaccines, are presented.

## Methods

### Study design and participants

This open-label, ongoing phase 2/3 study (clinicaltrials.gov: NCT04927065) evaluates the safety and immunogenicity of severe acute respiratory syndrome coronavirus (SARS-CoV-2) variant-containing vaccine candidates against Coronavirus disease 2019 (Covid-19). Part J of the study evaluates the safety, reactogenicity and immunogenicity of 50-µg doses of the monovalent mRNA-1273.815 vaccine (50-µg omicron XBB.1.5 spike protein mRNA) and bivalent mRNA-1273.231 vaccine (25-µg omicron XBB.1.5 and 25-µg omicron BA.4/BA.5 spike protein mRNAs). The vaccines were administered as fifth doses to adults who previously received a two-dose primary series and a booster dose of an original Covid-19 vaccine and a booster dose of a bivalent (Original + omicron BA.4/BA.5) vaccine. Interim 15-day analysis results are reported (data cutoff date May 16, 2023). Additional details of the study design and analysis are provided in the Supplementary protocol and statistical analysis plan.

Adults with a known history of SARS-CoV-2 infection ≤3 months from screening were excluded from the study. Additional details of inclusion/exclusion criteria are provided in the Supplement.

### Trial Oversight

The trial is being conducted across 9 U.S. sites, in accordance with the International Council for Harmonisation of Technical Requirements for Registration of Pharmaceuticals for Human Use, Good Clinical Practice guidelines. The central Institutional Review Board (Advarra, Inc., 6100 Merriweather Drive, Columbia, MD 21044) approved the protocol and consent forms. All participants provided written informed consent.

### Trial Vaccine

The monovalent mRNA-1273.815 50-µg vaccine contains 50-µg of mRNA encoding the prefusion-stabilized spike glycoprotein of the SARS-CoV-2 subvariants XBB.1.5/XBB.1.9.1. The bivalent mRNA-1273.231 50-µg contains 25-µg each of mRNAs encoding the prefusion-stabilized spike glycoproteins of the SARS-CoV-2 omicron variant (BA.4/BA.5) and the SARS-CoV-2 subvariants XBB.1.5/XBB.1.9.1, formulated in a 1:1 ratio. In both vaccines, mRNAs were encapsulated in lipid nanoparticles as described previously.^2^ The booster doses of mRNA-1273.815 and mRNA-1273.231 were each administered by intramuscular injection at 50-µg of mRNA.

### Randomization

Approximately 100 participants were planned to be randomized in a 1:1 ratio to receive a single booster dose of mRNA-1273.815 (50-µg) or mRNA-1273.231 (50-µg). The two groups were randomized 1:1 in an open-label manner. No statistical hypothesis testing was performed between the two randomized groups and all results are descriptive. As prespecified, the exploratory endpoint of surveillance for Covid-19 events begins 14 days after the booster dose and is therefore not applicable to the day 15 interim analysis.

### Study Objectives

The primary objectives of the study are evaluation of the safety and reactogenicity of mRNA-1273.815 and mRNA-1273.231 when administered as a fifth (third booster) dose. The evaluation of immunogenicity based on neutralizing antibody (nAb) responses at day 15 against the variants contained in the vaccines was also a primary objective.

### Statistical Analysis

Safety assessments in the safety set included solicited local and systemic adverse reactions ≤7 days and unsolicited adverse events ≤28 days post-booster administrations, and serious adverse events, adverse events leading to discontinuation from study vaccine and/or participation, medically-attended adverse events, and adverse events of special interest from day 1 through the interim study period.

Immunogenicity was assessed in participants in the Per-Protocol Set for Immunogenicity at pre-booster and day 15 post-booster dose by lentivirus-based pseudovirus neutralizing assays as described in the Supplementary Methods. The BA.4/BA.5 and ancestral SARS-CoV-2 (D614G) were assessed using validated pseudovirus assays, and those for XBB.1.5, XBB.1.16, and BQ.1.1, were qualified assays (Duke University).^3^ In a separate analysis, ancestral SARS-CoV2 (Wuhan-Hu-1, D614G) and BA.4/BA.5, XBB.1.5, XBB.1.16, XBB.2.3.2, EG.5.1, FL.1.5.1 and BA.2.86 variants were assessed using a research grade Vesicular Stomatitis Virus (VSV)-based SARS-CoV-2 Pseudovirus Neutralization Assay in a randomly-selected subgroup (n=20) of participants who received mRNA-1273.815. Hypothesis testing was not pre-specified with respect to the immune responses and results are descriptive. Geometric mean levels, and the geometric mean fold-rises (GMFRs) from pre-booster baseline levels and the corresponding 95% CI after the mRNA-1273.815 and mRNA-1273.231 doses are provided.

Interim day 15 analysis results are presented (data cutoff date May 16, 2023). All analyses were conducted using SAS Version 9.4 or higher.

## Results

In April (25^th^ to 27^th^) 2023, 101 participants were enrolled and received mRNA-1273.815 (n=50) and mRNA-1273.231 (n=51). Baseline characteristics were generally balanced for the groups (Table 1) including intervals between prior doses and percentages of participants with evidence of SARS-CoV-2 infection pre-booster. The median intervals (interquartile ranges) from the prior dose of the BA.4/BA.5-containing bivalent vaccine were 8.2 (8.1-8.3) and 8.3 (8.1-8.4) months for the mRNA-1273.815 and mRNA-1273.231 vaccines, respectively.^4^

**Table 1.**
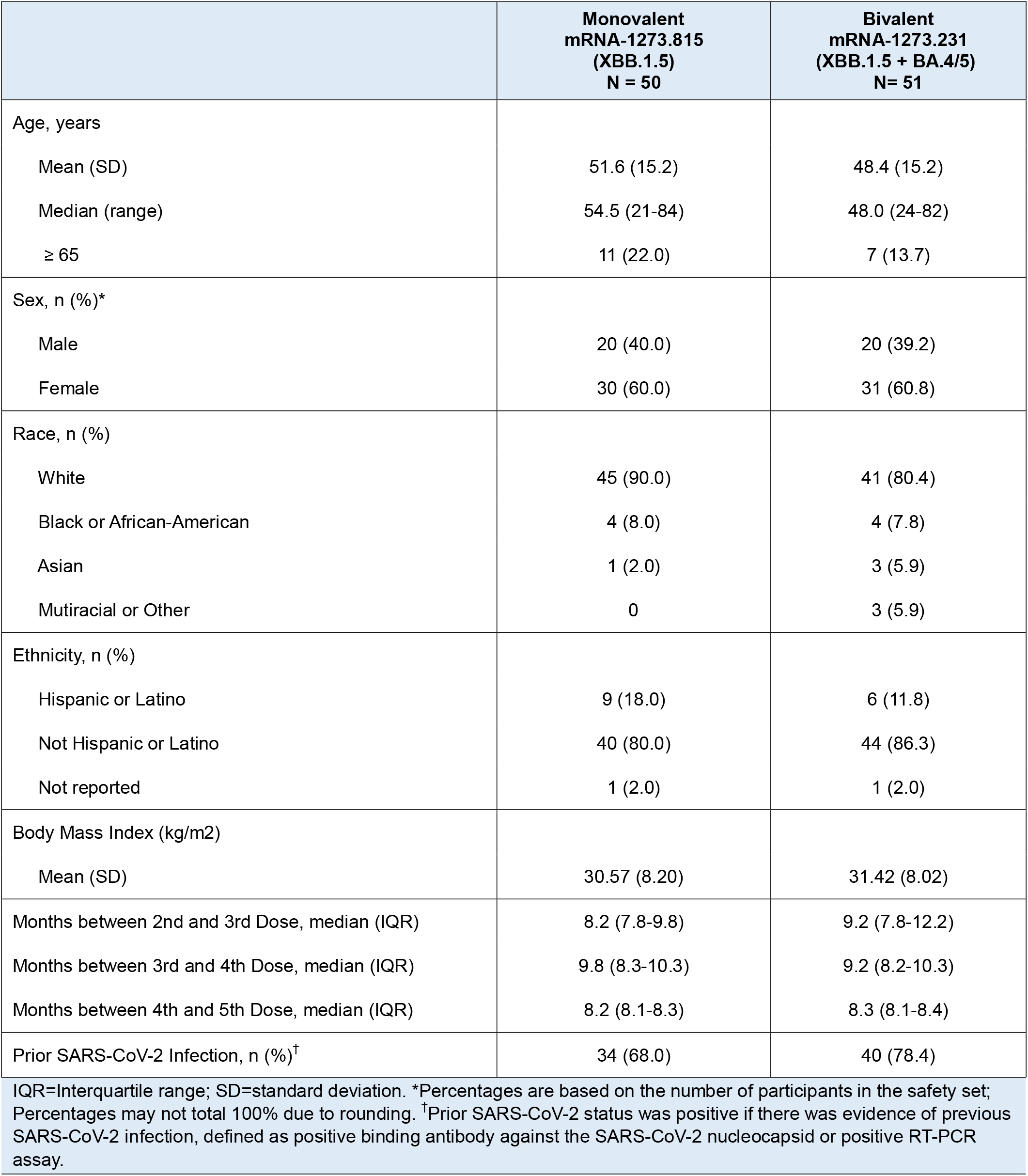
Demographics and Characteristics of Study Participants (Safety Set)

The pre-booster and day 15 titers for both vaccines, mRNA-1273.815 and mRNA-1273.231, are shown in Figures 1 and S1 and Tables S1-S2 for XBB.1.5, XBB.1.6, BQ.1.1, BA.4/BA.5 and ancestral SARS-CoV-2 (D614G). Both vaccines increased the neutralizing antibody responses as measured by a lentivirus reporter assay at day 15 in all participants in the Per-Protocol Set for Immunogenicity and those with and without prior SARS-CoV-2 infection. The nAb geometric mean levels and geometric mean fold-rises from pre-booster levels after the monovalent booster were numerically higher against XBB.1.5, XBB.1.16 and ancestral SARS-CoV-2 (D614G) than those of the bivalent booster and were comparable against BA.4/BA.5 and BQ1.1 variants for both vaccines. The mRNA-1273.815 vaccine also elicited similar nAb responses using a research grade VSV assay against XBB.1.5, XBB.1.16, XBB.2.3.2, EG.5.1, FL.1.5.1 and BA.2.86 variants in a subset of randomly-selected participants regardless of prior SARS-CoV-2 infection (Figure 2).

**Figure 1.**
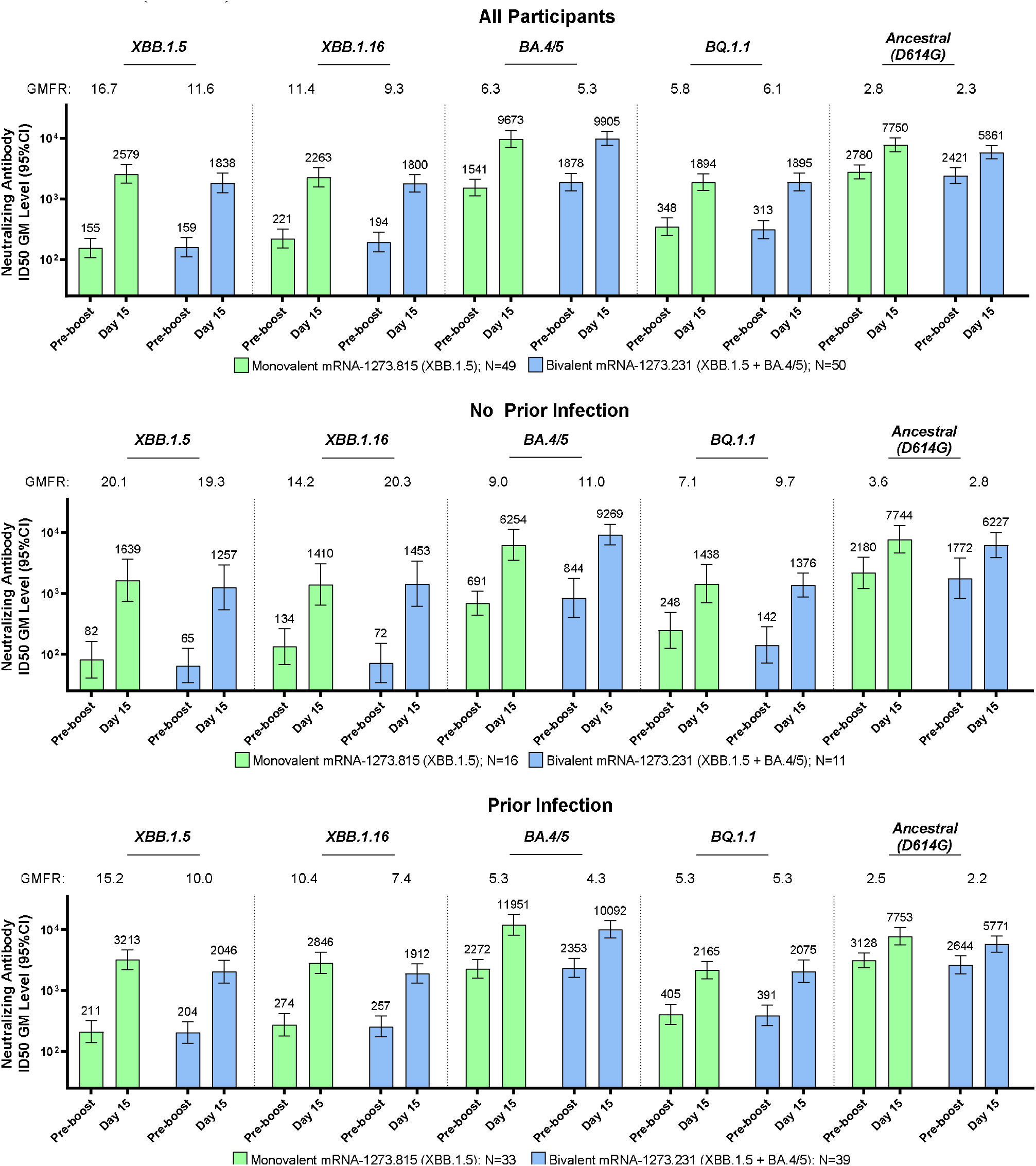
Neutralizing Antibodies After a Booster Dose of XBB.1.5-Containing Monovalent and Bivalent Vaccines Against XBB.1.5, XBB.1.16, BQ.1.1 and BA.4/5 Variants and Ancestral SARS-CoV-2 (D614G) GM=geometric mean; GMFR=geometric mean fold rise at day 15 relative to pre-booster. GM-levels of neutralizing antibody titers were assessed at pre-booster and at day 15 post-vaccination for the monovalent (mRNA-1273.815) and bivalent (mRNA-1273.231) vaccines using a lentivirus-based pseudovirus assay as described previously^3^ and in the Supplementary Methods. Analyses were performed in all participants in the Per-protocol Set for Immunogenicity (those with and without prior SARS-CoV-2 infection), and in those with prior infection and without prior infection. Antibody values reported as below the lower limit of quantification (LLOQ: 18.5 [1.3 log_10_] for ancestral SARS-CoV-2 [D614G] and 36.7 [1.6 log_10_] for Omicron BA.4/BA.5) are replaced by 0.5 x LLOQ. Values greater than the upper limit of quantification (ULOQ: 45,118 [4.7 log_10_] for ancestral SARS-CoV-2 [D614G] and 13,705 [4.1 log_10_) for Omicron BA.4/BA.5) are replaced by the ULOQ if actual values are not available. Antibody values reported as below the lower limit of detection (LOD: 10 for XBB.1.5, XBB.1.16 and BQ.1.1) are replaced by 0.5 x LOD.

**Figure 2.**
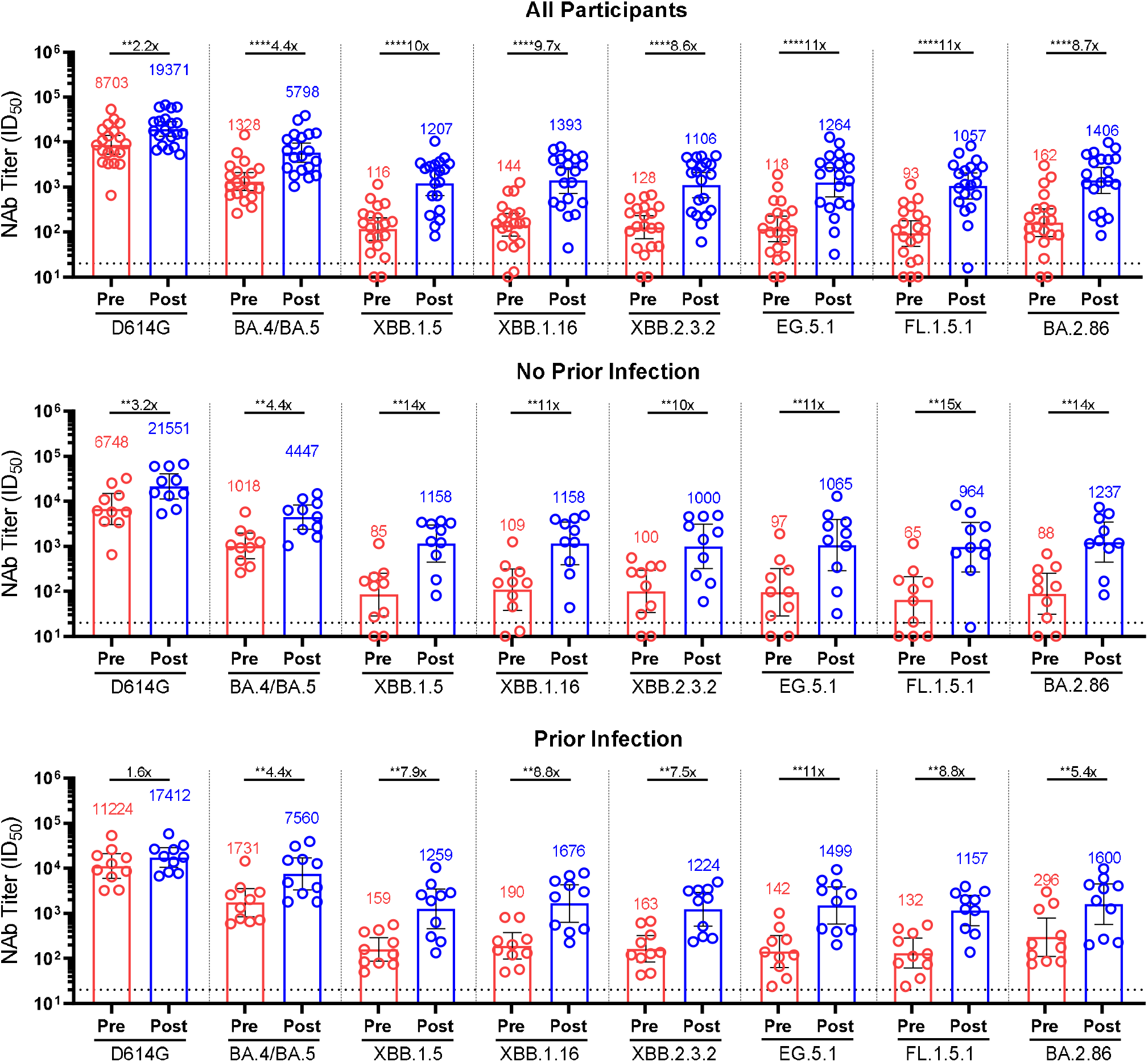
Analysis of Neutralizing Antibody Titers Against Ancestral SARS-CoV-2 (D614G) and BA.4/BA.5, XBB.1.5, XBB.1.16, XBB.2.3.2, EG.5.1, FL.1.5.1 and BA.2.86 Variants in a Randomly-selected Subset of Participants Who Received Monovalent mRNA-1273.815. Analysis of neutralizing antibody titers against variants in randomly-selected all participants (n=20) and in those with (n=10) and without (n=10) SARS-CoV-2 infection from mRNA-1273.815 arm using a research grade VSV-based pseudovirus assay (supplementary methods). **p≤0.01 and ****p<0.0001 by the Wilcoxon t-test. Dotted line denotes the limit of detection (20) of the assay.

During a median follow-up (interquartile range) of 20 (20-22) days for both vaccine groups, the occurrence of solicited local and systemic adverse reactions and unsolicited adverse events (AEs) were generally similar to those previously reported for the original mRNA-1273 50-µg and omicron BA.1- and BA.4/BA.5-containing bivalent mRNA-1273 vaccines^4-6^ (Figure 3 and Tables S3-S4).

**Figure 3.**
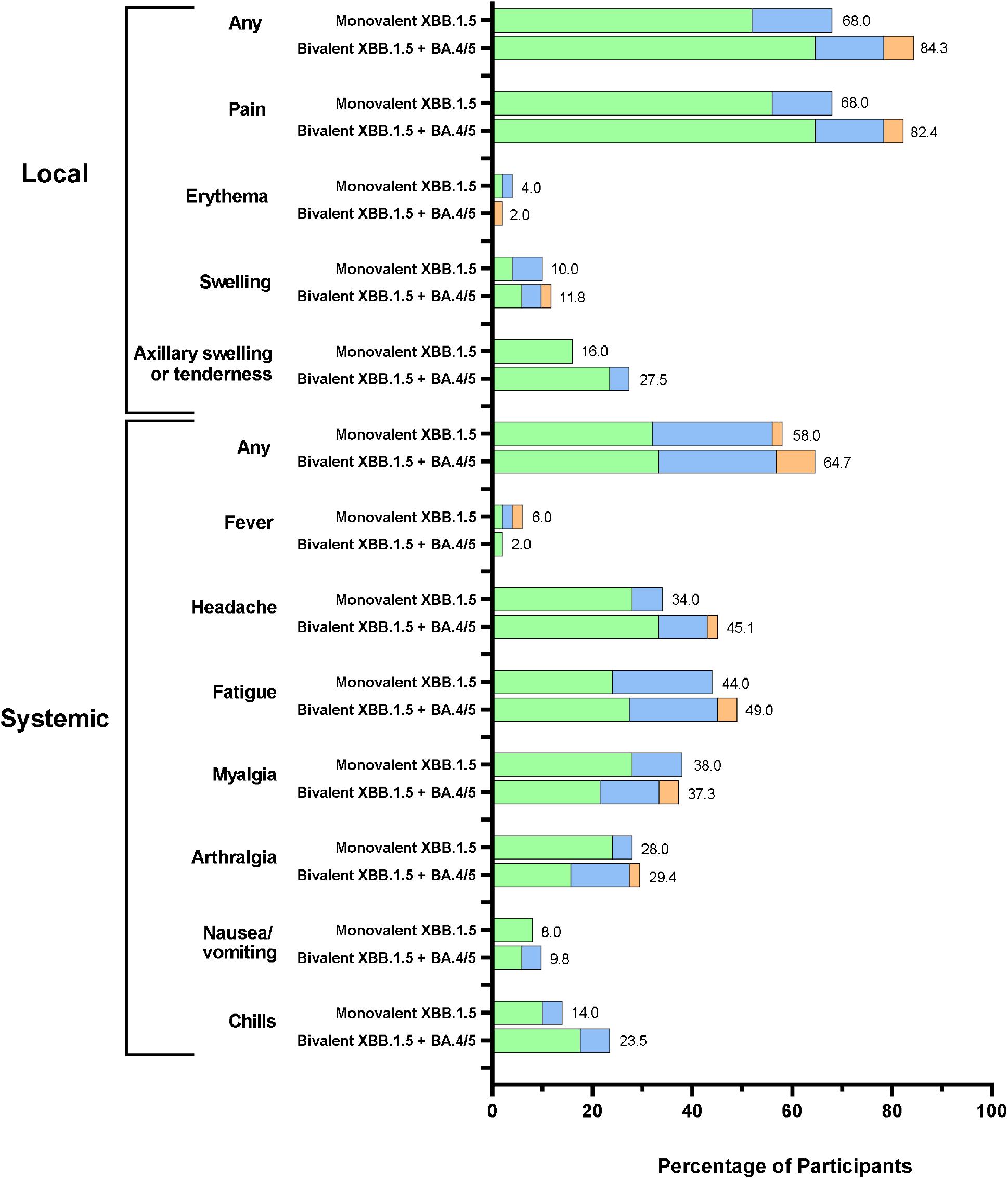
Solicited Local and Systemic Adverse Reactions. Shown are the percentages of participants in whom solicited local or systemic adverse reactions occurred within 7 days after the booster dose in the solicited safety set (n=50 in the monovalent mRNA-1273.815 group and n=51 in the bivalent mRNA-1273.231 group).

There were no serious or fatal AEs reported.

## Discussion

In this interim analysis, the XBB.1.5-containing monovalent and bivalent vaccines had tolerability similar to prior vaccinations^4-6^ and elicited potent neutralizing responses against Omicron variants of the XBB-lineage including XBB.1.5, XBB.1.16 and XBB.2.3.2, consistent with the limited antigenic differences measured between these variants.^7,8^ Antibody responses were numerically higher with the monovalent vaccine against the XBB-lineage variants, compared to the bivalent vaccine, and generally the responses were higher against the XBB-lineage variants, compared to previous variants.

The XBB.1.5 monovalent mRNA-1273.815 vaccine also cross-neutralized the more recent EG.5.1 (Eris), FL.1.5.1 (Fornax) and BA.2.86 (Pirola) variants. EG.5.1 and FL.1.5.1 are descendants of the Omicron lineage and similar in genetic makeup to XBB.1.5, and contain additional spike mutations including the F456L mutation detected in both variants.^7,8^ EG.5.1 is considered a global variant of interest that is now dominant in the US and the FL.1.5.1 variant is beginning to surge in parts of the U.S.^8,9^ The vaccine also neutralized the BA.2.86 (Pirola) variant which has recently been detected globally as well as in the US. This variant has multiple additional spike mutations (>30) beyond those seen in the Omicron BA.5 and XBB.1.5 spike proteins.^8-12^ While these variants may be capable of evading immunity from prior infection or vaccination, overall these data reassuringly suggest that the XBB.1.5-containing mRNA-1273.815 vaccine has the potential to provide protection against these emerging variants as well as others anticipated to circulate during the upcoming 2023-2024 vaccination season.

This study was not powered for a statistical comparison of the immune responses between the vaccine groups or vaccine efficacy. The study design enabled a rapid assessment of the antibody responses and the results support the strategy of updating Covid-19 vaccines to more closely match circulating variants. These clinical data also informed the selection of XBB.1.5-containing monovalent Covid-19 vaccines as recommended by the U.S. FDA for use in 2023-2024 immunizations.

## Supporting information

CONSORT Checklist

Protocol and SAP

## Data Availability

As the trial is ongoing, access to patient-level data and supporting clinical documents with qualified external researchers may be available upon request and subject to review once the trial is complete.

## Acknowledgements

We thank the participants in the trial and the members of the mRNA-1273 trial team (listed in the Supplement) for their dedication and contributions to the trial, the Immune Assay Team at Duke University Medical Center, Durham, NC for PsVNA analyses, Daniela Montes Berrueta, and Matthew Koch from Moderna for the VSV SARS-CoV-2 pseudovirus analyses and data, Michael Whitt and Rita Kansal from (University of Tennessee Health Science Center, Memphis, TN) for generating the VSV-based SARS-CoV-2 pseudoviruses, and Frank J. Dutko (Moderna consultant) for figure development and editorial support.

