## Supplementary material for "Safety and Immunogenicity of XBB.1.5-Containing mRNA Vaccines": Protocol and SAP

This supplement contains the following items:

1. Original protocol, final protocol and summary of protocol changes integrated in summary of changes in final protocol.

2. Original statistical analysis plan, final statistical analysis plan and summary of changes for statistical analysis plan integrated in summary of changes in final statistical analysis plan.

### CLINICAL STUDY PROTOCOL

| | |
|---|---|
| **Protocol Title:** | A Phase 2/3 Study to Evaluate the Immunogenicity and Safety of mRNA Vaccine Boosters for SARS-CoV-2 Variants |
| **Protocol Number:** | mRNA-1273-P205 |
| **Sponsor Name:** | ModernaTX, Inc. |
| **Legal Registered Address:** | 200 Technology Square<br>Cambridge, MA 02139 |
| **Sponsor Contact and Medical Monitor:** | Spyros Chalkias, MD<br>ModernaTX, Inc.<br>200 Technology Square<br>Cambridge, MA 02139<br>Telephone: 1-617-355-0744<br> |
| **Regulatory Agency Identifier Number(s):** | IND: 19745 |
| **Amendment Number** | 10 |
| **Date of Amendment 10** | 23 Mar 2023 |
| **Date of Amendment 9** | 30 Sep 2022 |
| **Date of Amendment 8** | 01 Aug 2022 |
| **Date of Amendment 7** | 26 Apr 2022 |
| **Date of Amendment 6** | 17 Mar 2022 |
| **Date of Amendment 5** | 10 Feb 2022 |
| **Date of Amendment 4** | 04 Jan 2022 |
| **Date of Amendment 3** | 15 Sep 2021 |
| **Date of Amendment 2** | 26 Jul 2021 |
| **Date of Amendment 1** | 23 Jun 2021 |
| **Date of Original Protocol** | 21 May 2021 |

**CONFIDENTIAL**

All financial and nonfinancial support for this study will be provided by ModernaTX, Inc. The concepts and information contained in this document or generated during the study are considered proprietary and may not be disclosed in whole or in part without the expressed written consent of ModernaTX, Inc. The study will be conducted according to the *International Council for Harmonisation (ICH) of Technical Requirements for Pharmaceuticals for Human Use, E6(R2) Good Clinical Practice (GCP) Guidance.*

### PROTOCOL APPROVAL – SPONSOR SIGNATORY

| | |
|---|---|
| **Study Title:** | A Phase 2/3 Study to Evaluate the Immunogenicity and Safety of mRNA Vaccine Boosters for SARS-CoV-2 Variants |
| **Protocol Number:** | mRNA-1273-P205 |
| **Amendment Number:** | 10 |
| **Date of Amendment:** | 23 Mar 2023 |

Protocol accepted and approved by:

**See eSignature and date signed on last page of the document**

| | |
|---|---|
| Rituparna Das, MD, PhD<br>Vice President Clinical Development,<br>Infectious Diseases<br>ModernaTX, Inc.<br>200 Technology Square<br>Cambridge, MA 02139<br>Telephone: 1-617-710-9794 | Date |

### DECLARATION OF INVESTIGATOR

I have read and understood all sections of the protocol entitled "A Phase 2/3 Study to Evaluate the Immunogenicity and Safety of mRNA Vaccine Boosters for SARS-CoV-2 Variants" and the most recent version of the Investigator's Brochure (IB).

I agree to supervise all aspects of the protocol and to conduct the clinical investigation in accordance with the current Protocol, the *International Council for Harmonisation (ICH) of Technical Requirements for Pharmaceuticals for Human Use, E6(R2) Good Clinical Practice (GCP) Guidance*, and all applicable government regulations. I will not make changes to the protocol before consulting with ModernaTX, Inc. or implement protocol changes without Institutional Review Board (IRB) approval except to eliminate an immediate risk to participants.

I agree to administer study treatment only to participants under my personal supervision or the supervision of a sub-investigator. I will not supply study treatment to any person not authorized to receive it. I also agree that persons debarred from conducting or working on clinical studies by any court or regulatory agency will not be allowed to conduct or work on studies for the Sponsor or a partnership in which the Sponsor is involved. I will immediately disclose it in writing to the Sponsor if any person who is involved in the study is debarred, or if any proceeding for debarment is pending, or, to the best of my knowledge, threatened.

I will not disclose confidential information contained in this document including participant information, to anyone other than the recipient, study staffs and members of the IRB. I agree to ensure that this information will not be used for any purpose other than the evaluation or conduct of the clinical investigation without the prior written consent from ModernaTX, Inc. I will not disclose information regarding this clinical investigation or publish results of the investigation without authorization from ModernaTX, Inc.

The signature below provides the necessary assurance that this study will be conducted according to all stipulations of the protocol, including statements regarding confidentiality, and according to local legal and regulatory requirements, US federal regulations, and ICH E6(R2) GCP guidelines.

________________________________________ ________________________

Signature of Principal Investigator Date

____________________________________

Printed Name of Principal Investigator

### PROTOCOL AMENDMENT SUMMARY OF CHANGES

| DOCUMENT HISTORY | |
|---|---|
| **Document** | **Date** |
| Amendment 10 | 23 Mar 2023 |
| Amendment 9 | 30 Sep 2022 |
| Amendment 8 | 01 Aug 2022 |
| Amendment 7 | 26 Apr 2022 |
| Amendment 6 | 17 Mar 2022 |
| Amendment 5 | 10 Feb 2022 |
| Amendment 4 | 04 Jan 2022 |
| Amendment 3 | 15 Sep 2021 |
| Amendment 2 | 26 Jul 2021 |
| Amendment 1 | 23 Jun 2021 |
| Original Protocol | 21 May 2021 |

**Amendment 10, 23 Mar 2023: Current Amendment**

**Main Rationale for the Amendment**

The main purpose of this amendment is to add a new part to the study, Part J, to evaluate the safety and immunogenicity of vaccine candidates to target antigenically divergent variants Omicron BA.4/BA.5, and Omicron XBB.1.5.

The summary of changes table provided below describes the major changes made to Amendment 10 relative to Amendment 9, including the sections modified and corresponding rationales. The synopsis of Amendment 10 has been modified to correspond to changes the body of the protocol. Minor copy edits and administrative updates were made throughout the protocol to align with new content and/or accuracy.

| Section # and Name | Description of Change | Brief Rationale |
|---|---|---|
| Title Page, Protocol Approval Page, and Protocol Amendment Summary of Changes | Updated protocol version and date. Added Summary of Changes. | Updated to reflect the new protocol version. Summary of Changes added to be in line with Moderna guidelines for protocol amendments. |
| Section 1.1 Study Rationale | Added rationale for Part J.<br>Added Omicron XBB.1.5 as a new variant to be targeted and descriptions of new mRNA-1273.815 and mRNA-1273.231 vaccine candidates. | Updated to include the rationale for evaluating the immunogenicity and safety of new vaccine candidates against emerging Omicron variants.<br>To provide background information on new vaccine candidates and their targeted |

| Section # and Name | Description of Change | Brief Rationale |
|---|---|---|
| | | severe acute respiratory syndrome coronavirus 2 (SARS-CoV-2) variants. |
| Section 1.2 Nonclinical Studies | Added language informing that additional animal studies are currently being conducted with mRNA vaccines that target different Omicron variants. | To update with developments in the ongoing mRNA-1273 nonclinical program |
| Section 1.4 Benefit/Risk Assessment | Added new Omicron-targeting vaccine candidates.<br>Clarified that the pivotal study on which the safety profile of mRNA-1273 is based is Study mRNA-1273-P301 (COVE). | To update Benefit/Risk section with the enhanced protection conferred by new Omicron-targeting vaccine candidates against coronavirus disease 2019 (COVID-19) caused by emergent SARS-CoV-2 variants.<br>To clearly reference the study on which the safety profile of mRNA-1273 is based. |
| Section 2 Objectives and Endpoints | Added objectives and endpoints of Part J of study. | To ensure the immunogenicity and safety objectives and endpoints for the new Part of study (Part J) are clearly described. |
| Section 3 Study Design | Removed Day 8 from the list of additional safety and immunogenicity visits from all study parts.<br>Added new vaccine candidates and description of study design of Part J. Added Part J to table describing treatment arms. Added information on number of visits conducted by participants in Part J. Added information on timing for blood draws from participants in Part J. Added information on dosing for participants in Part J. Added information on impact of Part J on timing for interim and final analysis. | To accurately capture that Day 8 is a safety call in which on site safety and immunogenicity procedures cannot be performed.<br>To capture the study design of Part J and the overall impact on the design, and analysis of the entire study. |
| Section 3.2 Scientific Rationale for Study Design | Updated with timing of blood and nasopharyngeal (NP) swab sample collection in Part J. | To clarify the timing for blood and NP swab sampling is aligned with the timing of SARS-CoV-2 circulation during the study. |

| Section # and Name | Description of Change | Brief Rationale |
|---|---|---|
| Section 3.3 Justification for Dose, Control Product, and Choice of Study Population | Updated dose of new multivalent booster candidates (50 µg). Added information on study population for Part J. | To clarify the dose of new multivalent booster candidates will remain similar to prior booster candidates. To describe prior study participation and prior vaccination of Part J participants to enhance understanding of eligibility requirements. |
| Section 3.4 End of Study Definition | Added participation in Part J in the description of End of Study. | To clarify prerequisites for study completion in participants who roll-over from Part H to Part J and general requirements for participants in Part J to complete all procedures in Part J schedule of events (SoE). |
| Section 4 Study Population | Added Part J information which includes number of participants, prior vaccination, and booster dose to be administered. | To include Part J population sample size and prior vaccination. |
| Section 4.1 Inclusion Criteria | Added eligibility criteria for participants in Part J of study. | To ensure Part J-specific eligibility criteria are accurately described. |
| Section 5.1 Investigational Products Administered | Added new Omicron-targeting vaccine candidates. | To ensure all investigational product (IP) used in this study is captured. |
| Section 5.2 Randomization and Blinding | Clarified that while randomization will occur for Part J participants, no blinding will occur in the study. | To accurately capture randomization in Part J but clarify that study remains open-label |
| Section 5.3. Preparation/Handling/ Storage/Accountability | Added relevant information on new Omicron-targeting vaccine candidates | To ensure completeness of Chemistry, Manufacturing, and Controls and Clinical Supply information on IP used in the study |
| Section 5.4 Study Intervention Compliance | Added link to Part J SoE in text describing visit windows | To accurately capture |
| Section 5.5.2 Concomitant Medications and Therapies | Clarified that non-study vaccinations and medications which are not used to treat COVID-19 or it symptoms will be recorded if taken through 28 days after study vaccination. | To distinguish between the timing for recording non-study vaccinations and concomitant medications depending on whether or not |

| Section # and Name | Description of Change | Brief Rationale |
|---|---|---|
| | Clarified that concomitant medications relevant to or for the treatment of an serious adverse event (SAE) or a medically attended adverse event (MAAE) or to prevent or treat COVID-19 or its symptoms will be recorded at any timepoint during the study. | they are used for the treatment of COVID-19 or it symptoms. |
| Section 6.1.1 Individual Participant Criteria for Delay of Study Vaccination | Added link to Part J SoE. | To ensure Part J procedures are captured when describing delay of vaccination. |
| Section 7 Study Assessments and Procedures | Added links to Part J SoE.<br>Revised polymerase chain reaction (PCR) sampling in Section 7.2 Immunogenicity Assessments, to reverse transcriptase PCR (RT-PCR) and added Day 181 as sampling timepoint for RT-PCR for all study parts.<br>Removed SAE Hotline and all references to the SAE Hotline, updated SAE Mailbox e-mail address, and SAE Fax Line | To ensure safety monitoring, eDiary use, timing for NP swab sampling, and timing for blood collection are captured for Part J participants.<br>To ensure RT-PCR sampling is consistent with the SoE.<br>To ensure SAE reporting mechanisms are up to date. |
| Section 8.1 Blinding and Responsibility for Analyses | Stated that randomization will occur for Part J participants. | To clarify that while randomization will occur for Part J participants, treatment assignment will remain unblinded and the study will remain open-label. |
| Section 8.2 Statistical Hypotheses | Stated that no formal hypothesis testing will be conducted for Part J. | To ensure hypothesis testing is accurately captured for all Parts of the study. |
| Section 8.3 Sample Size Determination | Added information on sample size determination for Part J of study. | To ensure sample size determination for the entire study captures Part J. |
| Section 8.5.3.2.1 Analysis for the Primary Immunogenicity Objective for Part J | Added primary analysis information for Part J. | To clarify analysis set and statistical models to be used for primary immunogenicity analysis. |
| Section 8.5.6 | Stated that subgroup analysis may not be performed for Part J due to small sample size. | To ensure subgroup analyses are accurately described for the appropriate Parts of the study. |

| Section # and Name | Description of Change | Brief Rationale |
|---|---|---|
| Section 8.6.1, Interim Analysis and Section 8.6.2 Final Analyses | Added impact of Part J on timing for interim and final analyses. | To clarify that the timing of interim and final analyses is dependent on all parts of the study. |
| Section 10.1 Appendix 1: Schedule of Events | Added Table 20 Schedule of Events for Part J. | To ensure procedures and events related to Part J of the study are captured in the protocol. |

### PROTOCOL SYNOPSIS

**Name of Sponsor/Company:** ModernaTX, Inc.

**Name of Investigational Product:** mRNA-1273.211, mRNA-1273, mRNA-1273.617.2, mRNA-1273.213, mRNA-1273.529, mRNA-1273.214, mRNA-1273.222, mRNA-1273.815, and mRNA-1273.231 for injection

**Name of Active Ingredient:** mRNA-1273.211, mRNA-1273, mRNA-1273.617.2, mRNA-1273.213, mRNA-1273.529, mRNA-1273.214, mRNA-1273.222, mRNA-1273.815, and mRNA-1273.231

**Protocol Title:** A Phase 2/3 Study to Evaluate the Immunogenicity and Safety of mRNA Vaccine Boosters for SARS-CoV-2 Variants

**Protocol Number:** mRNA-1273-P205

**Study Period (years):** Approximately 12 months

**Phase of Development:** Phase 2/3

**Estimated Date First Participant Enrolled:** May 2021

**Estimated Date Last Participant Completed:** Dec 2023

**Total Number of Sites:** Approximately 25 sites in the United States or its territories

**Objectives and Endpoints:**

This study consists of 9 parts: A (1, 2), B, C, D, E, F, G, H, and J. The objectives and endpoints in each part are described in the tables below; Part E objectives and endpoints are described in a site-specific protocol amendment and are not covered in this global protocol amendment.

**Part A.1: 50 µg mRNA-1273.211 and 100 µg mRNA-1273.211**

| Objectives | Endpoints |
|---|---|
| **Primary** | |
| **To be assessed for each dose level of mRNA-1273.211**<br>• To demonstrate non-inferior immune response of a single booster dose of mRNA-1273.211 compared with 2 priming doses of mRNA-1273 in the pivotal Phase 3 efficacy trial (Study mRNA-1273-P301 [COVE]):<br>– To demonstrate non-inferiority based on geometric mean titer (GMT) ratio (mRNA-1273.211 vs. mRNA-1273) against ancestral SARS-CoV-2 with a | **To be assessed for each dose level of mRNA-1273.211**<br>• Geometric mean titer (GMT) ratio of GMT of mRNA-1273.211 against ancestral SARS-CoV-2 at Day 29 after the booster dose over GMT of mRNA-1273 against ancestral SARS-CoV-2 at Day 57 (historical control)<br>• Seroresponse rate (SRR) difference between mRNA-1273.211 against ancestral SARS-CoV-2 at Day 29 after the booster dose and mRNA-1273 against ancestral SARS-CoV-2 at Day 57 (historical control) |

| | |
|---|---|
| non-inferiority margin of 1.5<br>– To demonstrate non-inferiority based on the seroresponse rate (SRR) (mRNA-1273.211 - mRNA-1273) against ancestral SARS-CoV-2 with a non-inferiority margin of 10% | |
| **To be assessed for each dose level of mRNA-1273.211**<br>• To demonstrate non-inferior immune response based on GMT ratio of mRNA-1273.211 as a single booster dose against the variant B.1.351, compared to mRNA-1273 after 2 priming doses against ancestral SARS-CoV-2 with a non-inferiority margin of 1.5<br>• To demonstrate non-inferior immune response based on the SRR of a single booster dose of mRNA-1273.211 against the variant B.1.351 as compared to 2 priming doses of mRNA-1273 against ancestral SARS-CoV-2 with a non-inferiority margin of 10% | **To be assessed for each dose level of mRNA-1273.211**<br>• GMT ratio of GMT of mRNA-1273.211 against the variant B.1.351 at Day 29 after the booster dose over GMT of mRNA-1273 against ancestral SARS-CoV-2 at Day 57 (historical control).<br>• SRR difference between mRNA-1273.211 against the variant B.1.351 at Day 29 after the booster dose and mRNA-1273 against ancestral SARS-CoV-2 at Day 57 (historical control). |
| • To evaluate the safety and reactogenicity of mRNA-1273.211 | • Solicited local and systemic reactogenicity adverse reactions (ARs) during a 7-day follow-up period after vaccination<br>• Unsolicited adverse events (AEs) during the 28-day follow-up period after vaccination<br>• Serious AEs (SAEs), medically attended AEs (MAAEs), AEs leading to withdrawal and AEs of special interest (AESI) from Day 1 to end of study (EoS) |
| **Secondary** | |
| • To compare immune response of mRNA-1273.211 as booster against variants compared to the priming series of mRNA-1273 against ancestral SARS-CoV-2 | • GMT ratio and SRR difference of mRNA-1273.211 as a booster dose against variants compared to the priming series of mRNA-1273 against ancestral SARS-CoV-2 |
| **Exploratory** | |
| • To assess for symptomatic and asymptomatic SARS-CoV-2 infection | • Laboratory-confirmed symptomatic or asymptomatic SARS-CoV-2 infection will be defined in participants:<br>– Primary case definition per the mRNA-1273-P301 (COVE) study |

| | |
|---|---|
| | – Secondary case definition based on the Centers for Disease Control (CDC) criteria: the presence of one of the CDC-listed symptoms (https://www.cdc.gov/coronavirus/2019-ncov/symptoms-testing/symptoms.html) and a positive reverse transcriptase polymerase chain reaction (RT-PCR) test on a respiratory sample<br>– Asymptomatic SARS-CoV-2 infection is defined as a positive RT-PCR test on a respiratory sample in the absence of symptoms or a positive serologic test for anti-nucleocapsid antibody after a negative test at time of enrollment |
| • To evaluate the genetic and/or phenotypic relationships of isolated SARS-CoV-2 strains to the vaccine sequence | • Characterize the SARS-CoV-2 genomic sequence of viral isolates and compare with the vaccine sequence<br>• Characterize the immune responses to vaccine breakthrough isolates |
| • Exploratory analyses on the immune response to specific viral strains may be performed to compare different booster regimens<br>– To compare the immune response of 50 µg of mRNA-1273.211 with 100 µg of mRNA-1273.211 against the same viral strains | • GMT ratio and SRR difference of 50 µg mRNA-1273.211 compared to 100 µg against the same viral strains |
| • To compare immune response of mRNA-1273.211 as booster against variants compared to the priming series of mRNA-1273 against the variants | • GMT ratio and SRR difference of mRNA-1273.211 as a booster dose against variants compared to the priming series of mRNA-1273 against the variants |

**Part A.2: Second booster dose 50 µg mRNA-1273.214: Participants who received mRNA-1273.211 50 µg as a first booster dose in Part A.1**

| Objectives | Endpoints |
|---|---|
| **Primary** | |
| • To evaluate the immunogenicity of mRNA-1273.214 (50 µg) as a second booster dose in participants who previously received a first booster dose of mRNA-1273.211 (50 µg) | • GMT ratio and SRR difference of mRNA-1273.214 (50 µg) as a second booster dose against the ancestral SARS-CoV-2 and against SARS-CoV-2 variants (including Omicron) compared to mRNA-1273.211 (50 µg) against the ancestral SARS-CoV-2 and against SARS-CoV-2 variants as the first booster dose (Day 29, Day 181) |

| | |
|---|---|
| • To assess the safety and reactogenicity of the mRNA-1273.214 (50 µg) given as a second booster dose in participants who previously received a first booster dose of mRNA-1273.211 (50 µg) | • Solicited local and systemic reactogenicity ARs during a 7-day follow-up period after vaccination<br>• Unsolicited AEs during the 28-day follow-up period after vaccination<br>• SAEs, MAAEs, AEs leading to withdrawal and AESI from Day 1 to EoS |
| **Secondary** | |
| • To evaluate the immunogenicity of mRNA-1273.214 (50 µg) as a second booster dose in participants who previously received a first booster dose of mRNA-1273.211 (50 µg) | • Antibody response of the mRNA-1273.214 (50 µg) against the ancestral SARS-CoV-2 and against SARS-CoV-2 variants (including Omicron) by GMT and SRR at multiple time points after the mRNA-1273.214 booster dose |
| **Exploratory** | |
| • To assess for symptomatic and asymptomatic SARS-CoV-2 infection | • Laboratory-confirmed symptomatic or asymptomatic SARS-CoV-2 infection will be defined in participants:<br>– Primary case definition per the mRNA-1273-P301 (COVE) study<br>– Secondary case definition based on the CDC criteria: the presence of one of the CDC-listed symptoms (https://www.cdc.gov/coronavirus/2019-ncov/symptoms-testing/symptoms.html) and a positive RT-PCR test on a respiratory sample<br>– Asymptomatic SARS-CoV-2 infection is defined as a positive RT-PCR test on a respiratory sample in the absence of symptoms or a positive serologic test for anti-nucleocapsid antibody after a negative test at time of enrollment |
| • To evaluate the genetic and/or phenotypic relationships of isolated SARS-CoV-2 isolates to the vaccine sequence | • Characterize the SARS-CoV-2 genomic sequence of viral isolates and compare with the vaccine sequence<br>• Characterize the immune responses to vaccine breakthrough isolates |

**Part B: 100 µg mRNA-1273**

| Objectives | Endpoints |
|---|---|
| **Primary** | |
| • To demonstrate non-inferior immune response of a single booster dose of 100 µg mRNA-1273 compared with 2 priming doses of mRNA-1273 in the | • GMT ratio of 100 µg mRNA-1273 against ancestral SARS-CoV-2 at Day 29 after the booster dose over GMT of 100 µg mRNA-1273 against ancestral SARS-CoV-2 at Day 57 |

| | |
|---|---|
| pivotal Phase 3 efficacy trial (Study mRNA-1273-P301 [COVE]):<br>– To demonstrate non-inferiority based on GMT ratio (100 µg mRNA-1273 single booster dose vs. 100 µg primary series mRNA-1273) against ancestral SARS-CoV-2 with a non-inferiority margin of 1.5<br>– To demonstrate non-inferiority based on the seroresponse rate (SRR) (100 µg mRNA-1273 single booster dose vs. 100 µg primary series mRNA-1273) against ancestral SARS-CoV-2 with a non-inferiority margin of 10% | (historical control)<br>• SRR difference between 100 µg mRNA-1273 against ancestral SARS-CoV-2 at Day 29 after the booster dose and mRNA-1273 against ancestral SARS-CoV-2 at Day 57 (historical control) |
| • To evaluate the safety and reactogenicity of mRNA-1273 | • Solicited local and systemic reactogenicity ARs during a 7-day follow-up period after vaccination<br>• Unsolicited AEs during the 28-day follow-up period after vaccination<br>• SAEs, MAAEs, AEs leading to withdrawal and AESI from Day 1 to EoS |
| **Secondary** | |
| • To compare immune response of 100 µg mRNA-1273 as booster against variants compared to the priming series of mRNA-1273 against ancestral SARS-CoV-2 | • GMT ratio and SRR difference of 100 µg mRNA-1273 as a booster dose against variants compared to the priming series of mRNA-1273 against ancestral SARS-CoV-2 |
| **Exploratory** | |
| • To assess for symptomatic and asymptomatic SARS-CoV-2 infection | • Laboratory-confirmed symptomatic or asymptomatic SARS-CoV-2 infection will be defined in participants:<br>– Primary case definition per the mRNA-1273-P301 (COVE) study<br>– Secondary case definition based on the CDC criteria: the presence of one of the CDC-listed symptoms (https://www.cdc.gov/coronavirus/2019-ncov/symptoms-testing/symptoms.html) and a positive RT-PCR test on a respiratory sample<br>– Asymptomatic SARS-CoV-2 infection is defined as a positive RT-PCR test on a respiratory sample in the absence of symptoms or a positive serologic test for anti-nucleocapsid antibody after a negative test at time of enrollment |

| | |
|---|---|
| • To evaluate the genetic and/or phenotypic relationships of isolated SARS-CoV-2 strains to the vaccine sequence | • Characterize the SARS-CoV-2 genomic sequence of viral isolates and compare with the vaccine sequence<br>• Characterize the immune responses to vaccine breakthrough isolates |
| • Exploratory analyses on the immune response to specific viral strains may be performed to compare different booster regimens<br>– To compare the immune response to ancestral virus strain and variants after a single booster dose of 50 or 100 µg mRNA-1273.211 (Part A.1) vs. a single booster dose of 100 µg mRNA-1273 | • GMT ratio and SRR difference of 50 or 100 µg mRNA-1273.211 (Part A.1) as a booster dose against ancestral and variants compared to a booster dose of 100 µg mRNA-1273 against the ancestral and variants |
| • To compare immune response of mRNA-1273 as booster against variants compared to the priming series of mRNA-1273 against the variants | • GMT ratio and SRR difference of mRNA-1273 as a booster dose against variants compared to the priming series of mRNA-1273 against the variants |

**Part C: 50 µg mRNA-1273.617.2 and 100 µg mRNA-1273.617.2**

| Objectives | Endpoints |
|---|---|
| **Primary** | |
| **To be assessed for each dose level of mRNA-1273.617.2**<br>• To demonstrate non-inferior immune response of a single booster dose of mRNA-1273.617.2 compared with 2 priming doses of mRNA-1273 in the pivotal Phase 3 efficacy trial (Study mRNA-1273-P301 [COVE]):<br>– To demonstrate non-inferiority based on GMT ratio of mRNA-1273.617.2 as a single booster dose against the variant B.1.617.2 vs. 100 µg primary series mRNA-1273) against ancestral SARS-CoV-2 with a non-inferiority margin of 1.5<br>– To demonstrate non-inferiority based on the SRR of a single booster dose of mRNA1273.617.2 against the variant B.1.617.2 vs. 100 µg primary series mRNA1273 against ancestral SARS-CoV-2 | **To be assessed for each dose level of mRNA-1273.617.2**<br>• GMT ratio of mRNA-1273.617.2 against the variant B.1.617.2 at Day 29 after the booster dose over GMT of 100 µg mRNA-1273 against ancestral SARS-CoV-2 at Day 57 (historical control).<br>• SRR difference between mRNA-1273.617.2 against the variant at Day 29 after the booster dose and mRNA-1273 against ancestral SARS-CoV-2 at Day 57 (historical control) |

| | |
|---|---|
| with a non-inferiority margin of 10% | |
| • To evaluate the safety and reactogenicity of mRNA-1273.617.2 | • Solicited local and systemic reactogenicity ARs during a 7-day follow-up period after vaccination<br>• Unsolicited AEs during the 28-day follow-up period after vaccination<br>• SAEs, MAAEs, AEs leading to withdrawal and AESI from Day 1 to EoS |
| **Secondary** | |
| • To compare immune response of mRNA-1273.617.2 as booster against the ancestral viral strain and variants compared to the priming series of mRNA-1273 against ancestral SARS-CoV-2 | • GMT ratio and SRR difference of mRNA-1273.617.2 as a booster dose against ancestral SARS-CoV-2 and variants compared to the priming series of mRNA-1273 against ancestral SARS-CoV-2 |
| **Exploratory** | |
| • To assess for symptomatic and asymptomatic SARS-CoV-2 infection | • Laboratory-confirmed symptomatic or asymptomatic SARS-CoV-2 infection will be defined in participants:<br>– Primary case definition per the mRNA-1273-P301 (COVE) study<br>– Secondary case definition based on the CDC criteria: the presence of one of the CDC-listed symptoms (https://www.cdc.gov/coronavirus/2019-ncov/symptoms-testing/symptoms.html) and a positive RT-PCR test on a respiratory sample<br>– Asymptomatic SARS-CoV-2 infection is defined as a positive RT-PCR test on a respiratory sample in the absence of symptoms or a positive serologic test for anti-nucleocapsid antibody after a negative test at time of enrollment |
| • To evaluate the genetic and/or phenotypic relationships of isolated SARS-CoV-2 strains to the vaccine sequence | • Characterize the SARS-CoV-2 genomic sequence of viral isolates and compare with the vaccine sequence<br>• Characterize the immune responses to vaccine breakthrough isolates |
| • Exploratory analyses on the immune response to specific viral strains may be performed to compare different booster regimens<br>– To compare the immune response of 50 µg of mRNA-1273.617.2 with | • GMT ratio and SRR difference of 50 µg mRNA1273.617.2 compared to 100 µg against the same viral strains |

| | |
|---|---|
| 100 µg of mRNA-1273.617.2 against the same viral strains | |
| • Exploratory analyses on the immune response to specific viral strains may be performed to compare different booster regimens<br>– To compare immune response to ancestral virus and variants after a single booster dose of mRNA-1273.617.2 vs. a single booster dose of 100 µg mRNA-1273 (Part B) | • GMT ratio and SRR difference of mRNA-1273.617.2 as a booster dose against ancestral and variants compared to a booster dose of 100 µg mRNA-1273 (Part B) against ancestral SARS-CoV-2 and variant |
| • Exploratory analyses on the immune response to specific viral strains may be performed to compare different booster regimens:<br>– To compare immune response to the ancestral virus and variants after a single booster dose of mRNA-1273.617.2 vs. a single booster dose of 50 or 100 µg mRNA-1273.211 (Part A.1) | • GMT ratio and SRR difference of mRNA-1273.617.2 as a booster dose against ancestral SARS-CoV-2 and variants compared to a booster dose of 50 or 100 µg mRNA-1273.211 (Part A.1) against ancestral SARS-CoV-2 and variants |
| • To compare immune response of mRNA-1273.617.2 as a booster against variants compared to the priming series of mRNA-1273 against the variants | • GMT ratio and SRR difference of mRNA-1273.617.2 as a booster dose against variants compared to the priming series of mRNA-1273 against the variants |

**Part D: 50 µg mRNA-1273.213 and 100 µg mRNA-1273.213**

| Objectives | Endpoints |
|---|---|
| **Primary** | |
| **To be assessed for each dose level of mRNA-1273.213**<br>• To demonstrate non-inferior immune response of a single booster dose of mRNA-1273.213 compared with 2 priming doses of mRNA-1273 in the pivotal Phase 3 efficacy trial (Study mRNA-1273-P301 [COVE]):<br>– To demonstrate non-inferiority based on GMT ratio of mRNA-1273.213 as a single booster dose against the variant B.1.617.2 vs. 100 µg primary series mRNA-1273 against ancestral SARS-CoV-2 with a non-inferiority margin of 1.5 | **To be assessed for each dose level of mRNA-1273.213**<br>• GMT ratio of mRNA-1273.213 against the variant B.1.617.2 at Day 29 after the booster dose over GMT of 100 µg mRNA-1273 against ancestral SARS-CoV-2 at Day 57 (historical control)<br>• SRR difference between mRNA-1273.213 against the variant B.1.617.2 at Day 29 after the booster dose and mRNA-1273 against ancestral SARS-CoV-2 at Day 57 (historical control)<br>• GMT ratio of mRNA-1273.213 against the variant B.1.351 at Day 29 after the booster dose over GMT of 100 µg mRNA-1273 against ancestral SARS-CoV-2 at Day 57 (historical control) |

| | |
|---|---|
| – To demonstrate non-inferiority based on the SRR of a mRNA-1273.213 single booster dose against the variant B.1.617.2 vs. 100 µg primary series mRNA-1273 against ancestral SARS-CoV-2 with a non-inferiority margin of 10%<br>– To demonstrate non-inferiority based on GMT ratio of mRNA-1273.213 as a single booster dose against the variant B.1.351 vs. 100 µg primary series mRNA-1273 against ancestral SARS-CoV-2 with a non-inferiority margin of 1.5<br>– To demonstrate non-inferiority based on the SRR of a mRNA1273.213 single booster dose against the variant B.1.351 vs. 100 µg primary series mRNA1273 against ancestral SARS-CoV-2 with a non-inferiority margin of 10% | • SRR difference between mRNA-1273.213 against the variant B.1.351 at Day 29 after the booster dose and mRNA-1273 against ancestral SARS-CoV-2 at Day 57 (historical control) |
| • To evaluate the safety and reactogenicity of mRNA-1273.213 | • Solicited local and systemic reactogenicity ARs during a 7-day follow-up period after vaccination<br>• Unsolicited AEs during the 28-day follow-up period after vaccination<br>• SAEs, MAAEs, AEs leading to withdrawal and AESI from Day 1 to EoS |
| **Secondary** | |
| • To compare the immune response of mRNA-1273.213 as booster against the ancestral viral strain and other variants compared to the priming series of mRNA-1273 against ancestral SARS-CoV-2 | • GMT ratio and SRR difference of mRNA-1273.213 as a booster dose against ancestral SARS-CoV-2 and other variants compared to the priming series of mRNA-1273 against ancestral SARS-CoV-2 |
| **Exploratory** | |
| • To assess for symptomatic and asymptomatic SARS-CoV-2 infection | • Laboratory-confirmed symptomatic or asymptomatic SARS-CoV-2 infection will be defined in participants:<br>– Primary case definition per the mRNA-1273-P301 (COVE) study<br>– Secondary case definition based on the CDC criteria: the presence of one of the CDC-listed symptoms |

| | |
|---|---|
| | (https://www.cdc.gov/coronavirus/2019-ncov/symptoms-testing/symptoms.html) and a positive RT-PCR test on a respiratory sample<br>– Asymptomatic SARS-CoV-2 infection is defined as a positive RT-PCR test on a respiratory sample in the absence of symptoms or a positive serologic test for anti-nucleocapsid antibody after a negative test at time of enrollment |
| • To evaluate the genetic and/or phenotypic relationships of isolated SARS-CoV-2 strains to the vaccine sequence | • Characterize the SARS-CoV-2 genomic sequence of viral isolates and compare with the vaccine sequence<br>• Characterize the immune responses to vaccine breakthrough isolates |
| • Exploratory analyses on the immune response to specific viral strains may be performed to compare different booster regimens<br>– To compare the immune response of 50 µg of mRNA-1273.213 with 100 µg of mRNA-1273.213 against the same viral strains | • GMT ratio and SRR difference of 50 µg mRNA-1273.213 compared to 100 µg against the same viral strains |
| • Exploratory analyses on the immune response to specific viral strains may be performed to compare different booster regimens<br>– To compare immune response to ancestral SARS-CoV-2 and variants after a single booster dose of mRNA-1273.213 vs. a single booster dose of 50 or 100 µg mRNA-1273.211 (Part A.1)<br>– To compare immune response to ancestral SARS-CoV-2 and variants after a single booster dose of mRNA-1273.213 vs. a single booster dose of 100 µg mRNA-1273 (Part B)<br>– To compare immune response to ancestral SARS-CoV-2 and variants after a single booster dose of mRNA-1273.213 vs. a single booster dose of 50 or 100 µg mRNA-1273.617.2 (Part C) | • GMT ratio and SRR difference of mRNA-1273.213 as a booster dose against ancestral SARS-CoV-2 and variants compared to a booster dose of 50 or 100 µg mRNA-1273.211 (Part A.1) against ancestral SARS-CoV-2 and variants<br>• GMT ratio and SRR difference of mRNA-1273.213 as a booster dose against ancestral SARS-CoV-2 and variants compared to a booster dose of 100 µg mRNA-1273 (Part B) against ancestral SARS-CoV-2 and variant<br>• GMT ratio and SRR difference of mRNA-1273.213 as a booster dose against ancestral SARS-CoV-2 and variants compared to a booster dose of 50 or 100 µg mRNA-1273.617.2 (Part C) against ancestral SARS-CoV-2 and variant |
| • To compare immune response of mRNA-1273.213 as a booster against | • GMT ratio and SRR difference of mRNA-1273.213 as a booster dose against |

| | |
|---|---|
| variants compared to the priming series of mRNA-1273 against the variants | variants compared to the priming series of mRNA-1273 against the variants |

**Part F - Cohort 1: 50 µg mRNA-1273.529: Participants who previously received 100 µg mRNA-1273 primary series and have not received a mRNA-1273 booster dose previously.**

| Objectives | Endpoints |
|---|---|
| **Primary** | |
| • To demonstrate non-inferiority of the antibody response against the Omicron variant (B.1.1.529) of a first booster dose of mRNA-1273.529 compared to a first booster dose of mRNA-1273 (50 µg) based on GMT ratio and SRR difference<br>• To demonstrate superiority of the antibody response against the Omicron variant (B.1.1.529) of a first booster dose of mRNA-1273.529 compared to a first booster dose of mRNA-1273 (50 µg) based on GMT ratio. | Day 29 postboost GMT ratio of Omicron-specific GMT of mRNA-1273.529 over the Omicron-specific GMT of mRNA-1273 (historical mRNA-1273 booster dose control)<br>Day 29 SRR difference between mRNA-1273.529 against Omicron and mRNA-1273 against Omicron |
| • To evaluate the safety and reactogenicity of mRNA-1273.529 | • Solicited local and systemic reactogenicity ARs during a 7-day follow-up period after vaccination<br>• Unsolicited AEs during the 28-day follow-up period after vaccination<br>• SAEs, MAAEs, AEs leading to withdrawal and AESI from Day 1 to EoS |
| **Secondary** | |
| • To evaluate the immunogenicity of a mRNA-1273.529 dose compared to a mRNA-1273 administered as a first booster dose at all timepoints postboost | • GMT ratio of mRNA-1273.529 and mRNA-1273 against the Omicron variant at all timepoints postboost<br>• SRR difference between mRNA-1273.529 against the Omicron variant and mRNA-1273 against the Omicron variant<br>• GMT ratio of mRNA-1273.529 and mRNA-1273 against the ancestral SARS-CoV-2 and other variants at all timepoints postboost<br>• SRR difference between mRNA-1273.529 against the ancestral SARS-CoV-2 and other variants and mRNA-1273 against the ancestral SARS-CoV-2 and other variants |
| To compare the immune response of mRNA-1273.529 as a first booster dose against the Omicron variant compared to the priming series of mRNA-1273 | GMT ratio and SRR difference of mRNA-1273.529 as a booster dose against the Omicron variant compared to the priming series of mRNA-1273 |

| | |
|---|---|
| | against the ancestral SARS-CoV-2 (historical control) |
| **Exploratory** | |
| • To assess for symptomatic and asymptomatic SARS-CoV-2 infection | Laboratory-confirmed symptomatic or asymptomatic SARS-CoV-2 infection will be defined in participants:<br>– Primary case definition per the mRNA-1273-P301 (COVE) study<br>– Secondary case definition based on the CDC criteria: the presence of one of the CDC-listed symptoms (https://www.cdc.gov/coronavirus/2019-ncov/symptoms-testing/symptoms.html) and a positive RT-PCR test on a respiratory sample<br>– Asymptomatic SARS-CoV-2 infection is defined as a positive RT-PCR test on a respiratory sample in the absence of symptoms or a positive serologic test for anti-nucleocapsid antibody after a negative test at time of enrollment |
| • To evaluate the genetic and/or phenotypic relationships of isolated SARS-CoV-2 strains to the vaccine sequence | Characterize the SARS-CoV-2 genomic sequence of viral isolates and compare with the vaccine sequence<br>Characterize the immune responses to vaccine breakthrough isolates |
| • To characterize the cellular immune response of the mRNA-1273.529 booster dose against the ancestral SARS-CoV-2 and against variants | T-cell and B-cell response after the mRNA-1273.529 booster |

**Part F - Cohort 2: Second booster dose 50 µg mRNA-1273.529 or 50 µg mRNA-1273 dose: Participants who previously received 100 µg mRNA-1273 primary series and a booster dose of 50 µg mRNA-1273**

| Objectives | Endpoints |
|---|---|
| **Primary** | |
| • To demonstrate non-inferiority of the antibody response against the Omicron variant (B.1.1.529) of a second booster dose of mRNA-1273.529 compared to a second booster dose of mRNA-1273 (50 µg) based on GMT ratio and SRR difference<br>• To demonstrate superiority of the antibody response against the Omicron variant (B.1.1.529) of a second booster | • Day 29 postboost GMT ratio of Omicron-specific GMT of mRNA-1273.529 over the Omicron-specific GMT of mRNA-1273<br>• Day 29 SRR difference between mRNA-1273.529 against Omicron and mRNA-1273 against Omicron |

| | |
|---|---|
| dose of mRNA-1273.529 compared to a second booster dose of mRNA-1273 (50 µg) based on GMT ratio | |
| • To evaluate the safety and reactogenicity of mRNA-1273.529 | • Solicited local and systemic reactogenicity ARs during a 7-day follow-up period after vaccination<br>• Unsolicited AEs during the 28-day follow-up period after vaccination<br>• SAEs, MAAEs, AEs leading to withdrawal and AESI from Day 1 to EoS |
| **Secondary** | |
| • To evaluate the immunogenicity of mRNA-1273.529 booster compared to mRNA-1273 booster administered as a second booster dose at all timepoints postboost | • GMT ratio of mRNA-1273.529 and mRNA-1273 against the Omicron variant at all timepoints postboost<br>• SRR difference between mRNA-1273.529 against the Omicron variant and mRNA-1273 against the Omicron variant<br>• GMT ratio of mRNA-1273.529 and mRNA-1273 against the ancestral SARS-CoV-2 and other variants at all timepoints postboost<br>• SRR difference between mRNA-1273.529 against the ancestral SARS-CoV-2 and other variants and mRNA-1273 against the ancestral SARS-CoV-2 and other variants |
| • To compare the immune response of mRNA-1273.529 as a second booster dose against the Omicron variant compared to the priming series of mRNA-1273 | • GMT ratio and SRR difference of mRNA-1273.529 as a second booster dose against the Omicron variant compared to the priming series of mRNA-1273 against the ancestral SARS-CoV-2 (historical control) |
| **Exploratory** | |
| • To assess for symptomatic and asymptomatic SARS-CoV-2 infection | • Laboratory-confirmed symptomatic or asymptomatic SARS-CoV-2 infection will be defined in participants:<br>– Primary case definition per the mRNA-1273-P301 (COVE) study<br>– Secondary case definition based on the CDC criteria: the presence of one of the CDC-listed symptoms (https://www.cdc.gov/coronavirus/2019-ncov/symptoms-testing/symptoms.html) and a positive RT-PCR test on a respiratory sample<br>– Asymptomatic SARS-CoV-2 infection is defined as a positive RT-PCR test on a respiratory sample in the absence of |

| | |
|---|---|
| | symptoms or a positive serologic test for anti-nucleocapsid antibody after a negative test at time of enrollment |
| • To evaluate the genetic and/or phenotypic relationships of isolated SARS-CoV-2 strains to the vaccine sequence | • Characterize the SARS-CoV-2 genomic sequence of viral isolates and compare with the vaccine sequence<br>• Characterize the immune responses to vaccine breakthrough isolates |
| • To characterize the cellular immune response of mRNA-1273.529 as a booster against SARS-CoV-2 and other variants | • T-cell and B-cell response after the mRNA-1273.529 booster |

**Part G: Second booster dose 50 µg mRNA-1273.214: Participants who received 100 µg mRNA-1273 primary series and a booster dose of 50 µg mRNA-1273**

| Objectives | Endpoints |
|---|---|
| **Primary** | |
| • To demonstrate non-inferiority of the antibody response of a second booster dose of mRNA-1273.214 compared to mRNA-1273 (50 µg) when administered as a second booster dose against the Omicron variant (B.1.1.529) based on GMT ratio and SRR difference at Day 29 or Day 91<br>• To demonstrate superiority of the antibody response of a second booster dose of mRNA-1273.214 compared to mRNA-1273 (50 µg) administered as a second booster dose against the Omicron variant (B.1.1.529) based on GMT ratio at Day 29 or Day 91<br>• To demonstrate non-inferiority of the antibody response of a second booster dose of mRNA-1273.214 compared to mRNA-1273 (50 µg) when administered as a second booster dose against the ancestral SARS-CoV-2 based on GMT ratio at Day 29 or Day 91 | • GMT ratio of Omicron-specific GMT of mRNA-1273.214 over the Omicron-specific GMT of mRNA-1273 (Part F, Cohort 2, 50 µg mRNA-1273) at Day 29 and Day 91<br>• SRR difference between mRNA-1273.214 against Omicron variant and mRNA-1273 against Omicron variant at Day 29 and Day 91<br>• GMT ratio of ancestral SARS-CoV-2 GMT of mRNA-1273.214 over ancestral SARS-CoV-2 GMT of mRNA-1273 (Part F, Cohort 2, 50 µg mRNA-1273) at Day 29 and Day 91 |
| • To evaluate the safety and reactogenicity of mRNA-1273.214 | • Solicited local and systemic reactogenicity ARs during a 7-day follow-up period after vaccination<br>• Unsolicited AEs during the 28-day follow-up period after vaccination<br>• SAEs, MAAEs, AEs leading to withdrawal and AESI from Day 1 to EoS |

| Key Secondary | |
|---|---|
| • To demonstrate non-inferiority based on the SRR against ancestral SARS-CoV-2 of a second booster dose of mRNA-1273.214 compared to a second booster dose of mRNA-1273 (50 µg) at Day 29 or Day 91 | • SRR difference between mRNA-1273.214 against ancestral SARS-CoV-2 and mRNA-1273 against ancestral SARS-CoV-2 at Day 29 and Day 91 |
| **Secondary** | |
| • To evaluate the immunogenicity of mRNA-1273.214 booster compared to mRNA-1273 booster administered as a second booster dose at all timepoints postboost | • GMT ratio of mRNA-1273.214 and mRNA-1273 against the Omicron variant at all timepoints postboost<br>• SRR difference between mRNA-1273.214 against the Omicron variant and mRNA-1273 against the Omicron variant at all timepoints postboost<br>• GMT ratio of mRNA-1273.214 and mRNA-1273 against ancestral SARS-CoV-2 and other variants at all timepoints postboost<br>• SRR difference between mRNA-1273.214 against ancestral SARS-CoV-2 and other variants and mRNA-1273 against ancestral SARS-CoV-2 and other variants at all timepoints postboost |
| • To compare the immune response of mRNA-1273.214 as a second dose against the Omicron variant compared to the priming series of mRNA-1273 | • GMT ratio and SRR difference of mRNA-1273.214 as a second booster dose against the Omicron variant compared to the priming series of mRNA-1273 against the ancestral SARS-CoV-2 (historical control group) |
| **Exploratory** | |
| • To assess for symptomatic and asymptomatic SARS-CoV-2 infection | • Laboratory-confirmed symptomatic or asymptomatic SARS-CoV-2 infection will be defined in participants:<br>– Primary case definition per the mRNA-1273-P301 (COVE) study<br>– Secondary case definition based on the CDC criteria: the presence of one of the CDC-listed symptoms (https://www.cdc.gov/coronavirus/2019-ncov/symptoms-testing/symptoms.html) and a positive RT-PCR test on a respiratory sample<br>– Asymptomatic SARS-CoV-2 infection is defined as a positive RT-PCR test on a respiratory sample in the absence of symptoms or a positive serologic test for anti-nucleocapsid antibody after a negative |

| | |
|---|---|
| | test at time of enrollment |
| • To evaluate the genetic and/or phenotypic relationships of isolated SARS-CoV-2 strains to the vaccine sequence | • Characterize the SARS-CoV-2 genomic sequence of viral isolates and compare with the vaccine sequence<br>• Characterize the immune responses to vaccine breakthrough isolates |
| • To characterize the cellular immune response of mRNA-1273.214 as a booster against SARS-CoV-2 and other variants | • T-cell and B-cell response after the mRNA-1273.214 booster |

**Part H: Second booster dose of 50 µg mRNA-1273.222**

| Objectives | Endpoints |
|---|---|
| **Primary** | |
| • To demonstrate non-inferiority of the antibody response of a second booster dose of mRNA-1273.222 50 µg compared to mRNA-1273 50 µg when administered as a second booster dose against Omicron BA.4/5 based on GMT ratio and SRR difference at Day 29<br>• To demonstrate superiority of the antibody response of a second booster dose of mRNA-1273.222 compared to mRNA-1273 (50 µg) administered as a second booster dose against the Omicron BA.4/5 based on GMT ratio at Day 29<br>• To demonstrate non-inferiority of the antibody response of mRNA-1273.222 50 µg compared to mRNA-1273 50 µg when administered as a second booster dose against the ancestral SARS-CoV-2 D614G based on GMT ratio and SRR difference at Day 29 | • GMT ratio of Omicron BA.4/5 GMT of mRNA-1273.222 over the Omicron BA.4/5 GMT of mRNA-1273 (Part F, Cohort 2, 50 µg mRNA-1273) at Day 29<br>• SRR difference between mRNA-1273.222 against Omicron BA.4/5 and mRNA-1273 against Omicron BA.4/5 at Day 29<br>• GMT ratio of the ancestral SARS-CoV-2 D614G GMT of mRNA-1273.222 over the ancestral SARS-CoV-2 D614G GMT of mRNA-1273 at Day 29<br>• SRR difference between mRNA-1273.222 and mRNA-1273 against the ancestral SARS-CoV-2 D614G at Day 29 |
| • To evaluate the safety and reactogenicity of mRNA-1273.222 50 µg | • Solicited local and systemic reactogenicity ARs during a 7-day follow-up period after injection<br>• Unsolicited AEs during the 28-day follow-up period after injection<br>• SAEs, MAAEs, AEs leading to withdrawal, and AESI from Day 1 to EoS |
| **Secondary** | |
| • To evaluate the immunogenicity of mRNA-1273.222 50 µg as a second booster dose against the ancestral | • GMT ratio of mRNA-1273.222 50 µg and mRNA-1273 50 µg against ancestral |

| | |
|---|---|
| SARS-CoV-2 (and other variants) compared to a second booster dose of mRNA-1273 50 µg at all timepoints postbooster | SARS-CoV-2 (and other variants) at all timepoints postbooster<br>• SRR difference between mRNA-1273.222 50 µg and mRNA-1273 50 µg against ancestral SARS-CoV-2 and variants of concern at all timepoints postbooster |
| **Exploratory** | |
| • To assess for symptomatic and asymptomatic SARS-CoV-2 infection | • Laboratory-confirmed symptomatic or asymptomatic SARS-CoV-2 infection will be defined in participants:<br>– Primary case definition per the mRNA-1273-P301 (COVE) study<br>– Secondary case definition based on the CDC criteria: the presence of 1 of the CDC-listed symptoms (https://www.cdc.gov/coronavirus/2019-ncov/symptoms-testing/symptoms.html) and a positive RT-PCR test on a respiratory sample<br>– Asymptomatic SARS-CoV-2 infection is defined as a positive RT-PCR test on a respiratory sample in the absence of symptoms or a positive serologic test for antinucleocapsid antibody after a negative test at time of enrollment |
| • To evaluate the genetic and/or phenotypic relationships of isolated SARS-CoV-2 strains to the vaccine sequence | • Characterize the SARS-CoV-2 genomic sequence of viral isolates and compare with the vaccine sequence<br>• Characterize the immune responses to vaccine breakthrough isolates |

**Part J - Booster dose of mRNA-1273.815 (50 µg) or mRNA-1273.231 (50 µg): Participants who previously received a primary series of mRNA vaccine, a first booster dose of a monovalent mRNA vaccine, and a second booster dose of a bivalent mRNA vaccine against SARS-CoV-2**

| Objectives | Endpoints |
|---|---|
| **Primary** | |
| • To evaluate the immunogenicity of mRNA-1273.815 (50 µg) and mRNA-1273.231 (50 µg) against SARS-CoV-2 Omicron BA.4/BA.5, BQ.1.1, and XBB.1.5 subvariants at Day 15 and Day 29 | • Antibody response of mRNA-1273.815 (50 µg) and mRNA-1273.231(50 µg) against SARS-CoV-2 Omicron BA.4/BA.5, BQ.1.1, and XBB.1.5 subvariants by GMT, geometric mean fold rise (GMFR), and SRR at Day 15 and Day 29 |
| • To evaluate the safety and reactogenicity of mRNA-1273.815 | • Solicited local and systemic reactogenicity ARs during a 7-day follow-up period after injection |

| | |
|---|---|
| (50 µg) and mRNA-1273.231 (50 µg) | • Unsolicited AEs during the 28-day follow-up period after injection<br>• SAEs, MAAEs, AEs leading to withdrawal, and AESI from Day 1 to Day 181 (EoS) |
| **Exploratory** | |
| • To evaluate the immunogenicity of mRNA-1273.815 (50 µg) and mRNA-1273.231 (50 µg) against SARS-CoV-2 variants at any study timepoint | • Antibody response of mRNA-1273.815 (50 µg) and mRNA-1273.231(50 µg) against SARS-CoV-2 variants by GMT, GMFR, and SRR at any study timepoint |
| • To assess for symptomatic and asymptomatic SARS-CoV-2 infection | • Laboratory-confirmed symptomatic or asymptomatic SARS-CoV-2 infection will be defined in participants:<br>– Primary case definition per the mRNA-1273-P301 (COVE) study<br>– Secondary case definition based on the CDC criteria: the presence of 1 of the CDC-listed symptoms (https://www.cdc.gov/coronavirus/2019-ncov/symptoms-testing/symptoms.html) and a positive RT-PCR test on a respiratory sample<br>– Asymptomatic SARS-CoV-2 infection is defined as a positive RT-PCR test on a respiratory sample in the absence of symptoms or a positive serologic test for antinucleocapsid antibody after a negative test at time of enrollment |

**Overall Study Design:**

This is an open-label, Phase 2/3 study to evaluate the immunogenicity, safety, and reactogenicity of mRNA-1273.211, mRNA-1273, mRNA-1273.617.2, mRNA-1273.213, mRNA-1273.529, mRNA-1273.214, mRNA-1273.222, mRNA-1273.815, and mRNA-1273.231 administered as booster doses.

**Part A.1**

Part A.1 will evaluate the immunogenicity, safety, and reactogenicity of 2 dose levels of the mRNA-1273.211 vaccine candidate when administered as a single booster dose to adult participants of the mRNA-1273-P301 (COVE) study who have previously received 2 doses of mRNA-1273 as a primary series. Two dose levels (50 or 100 µg total mRNA content) of the mRNA-1273.211 booster will be evaluated in this study. Enrollment will begin with the mRNA-1273.211 50 µg dose arm, followed by the enrollment of the mRNA-1273.211 100 µg dose arm. The results of the mRNA-1273.211 vaccine candidate will be compared to the immunogenicity induced after a 2-dose primary series of mRNA-1273 in the mRNA-1273-P301 (COVE) study, which demonstrated the efficacy of mRNA-1273. See the Investigator's Brochure (IB) for further details.

**Part A.2**

Part A.2 will evaluate the immunogenicity, safety, and reactogenicity of the mRNA-1273.214 vaccine candidate when administered as a second booster dose to adult participants of the mRNA-1273-P205 study who have previously received 2 doses of mRNA-1273 as a primary series and a first booster dose of 50 µg of the mRNA-1273.211 in Part A.1 of this study.

**Part B**

Part B will evaluate the immunogenicity, safety, and reactogenicity of the mRNA-1273 vaccine when administered as a single booster dose to adult participants of the mRNA-1273-P301 (COVE) study who have previously received 2 doses of mRNA-1273 as a primary series. Enrollment of Part B will begin upon completion of enrollment of Part A.1 of the study. The results of the mRNA-1273 vaccine will be compared to the immunogenicity induced after a 2-dose primary series of mRNA-1273 in the mRNA-1273-P301 (COVE) study, which demonstrated the efficacy of mRNA-1273. See the IB for further details.

**Part C**

Part C will evaluate the immunogenicity, safety, and reactogenicity of 2 dose levels (50 or 100 µg) of the mRNA-1273.617.2 vaccine when administered as a single booster dose to adults who have previously received 2 doses of mRNA-1273 as a primary series in Study mRNA-1273-P301 (COVE) or under the Emergency Use Authorization (EUA). Enrollment of Part C 100 µg dose level will begin upon completion of enrollment of Part B of the study. Enrollment of the 50-µg dose arm will begin after completion of the 100 µg dose level arm in both Part C and Part D. The results of the mRNA-1273.617.2 vaccine will be compared to the immunogenicity induced after a 2-dose primary series of mRNA-1273 in the mRNA-1273-P301 (COVE) study, which demonstrated the efficacy of mRNA-1273. See the IB for further details.

**Part D**

Part D will evaluate the immunogenicity, safety, and reactogenicity of 2 dose levels (50 or 100 µg) of the mRNA-1273.213 vaccine when administered as a single booster dose to adults who have previously received 2 doses of mRNA-1273 as a primary series in Study mRNA-1273-P301 (COVE) or under the EUA. Enrollment of Part D 50 µg dose level arm will begin upon completion of enrollment of Part C 100µg dose level arm of the study. Part D 50 µg dose arm enrollment will begin after completion of the 100µg dose arm enrollment and may run in parallel with Part C 50 µg dose arm enrollment. The results of the mRNA-1273.213 vaccine will be compared to the immunogenicity induced after a 2-dose primary series of mRNA-1273 in the mRNA-1273-P301 (COVE) study, which demonstrated the efficacy of mRNA-1273. See the IB for further details.

**Part E**

Part E consists of a group described in a site-specific protocol amendment and will not be discussed in this global protocol amendment.

**Part F**

Part F will consist of 2 cohorts: Cohort 1 – adults who have previously received 2 doses of mRNA-1273 as primary series and Cohort 2- adults who have previously received 2 doses of 100 µg mRNA-1273 as a primary series and a single booster dose of 50 µg mRNA-1273, in Study mRNA-1273-P301 (COVE) or under the EUA.

Cohort 1 will evaluate the immunogenicity, safety, and reactogenicity of 50 µg of the mRNA1273.529 vaccine candidate when administered as a single booster dose to adults who have previously received 2 doses of mRNA-1273 as a primary series.

Cohort 2 will evaluate the immunogenicity, safety and reactogenicity of 50 µg of the mRNA-1273.529 and of 50 µg of the mRNA-1273 vaccine candidate when administered as a second booster dose to adults who have previously received 2 doses of 100 µg mRNA-1273 as a primary series and a single booster dose of 50 µg mRNA-1273.

Enrollment of the mRNA-1273.529 Cohort 1 will run in parallel with the mRNA-1273.529 in Cohort 2. Enrollment of the 50-µg mRNA-1273 arm in Cohort 2 will begin upon completion of enrollment of the mRNA-1273.529 Cohort 2 arm and may run in parallel with the enrollment of the mRNA-1273.529 Cohort 1 arm. For Cohort 1, the results of the mRNA1273.529 vaccine candidate administered as a booster will be compared to the immunogenicity induced after a booster dose of mRNA-1273 from the external historical comparator arm (details will be provided in statistical analysis plan [SAP]). For Cohort 2, the results of the mRNA-1273.529 vaccine candidate administered as the second booster dose will be compared to the immunogenicity induced after the second booster dose of mRNA-1273.

**Part G**

Enrollment of the mRNA-1273.214 50 µg second boost arm will begin upon completion of enrollment of the mRNA-1273 50 ug arm in Cohort 2 of Part F. Enrollment of the mRNA-1273.214 50 µg second boost arm may run in parallel with the enrollment of the mRNA-1273.529 Cohort 1 arm of Part F. Part G will evaluate the immunogenicity, safety, and reactogenicity of 50 µg of the mRNA1273.214 vaccine candidate when administered as a second booster dose to adults who have previously received 2 doses of 100 µg mRNA-1273 as a primary series and a single booster dose of 50 µg mRNA-1273. The results of the mRNA-1273.214 vaccine candidate administered as the second booster dose will be compared to the immunogenicity induced after the second booster dose of mRNA-1273 (Part F, Cohort 2, 50 µg mRNA-1273).

**Part H**

Part H will evaluate the immunogenicity, safety, and reactogenicity of 50 µg of the mRNA-1273.222 vaccine candidate when administered as a second booster dose to adults who have previously received 2 doses of 100 µg mRNA-1273 as a primary series and a single booster dose of 50 µg mRNA-1273. The immunogenicity of the mRNA-1273.222 candidate administered as the second booster dose will be compared to the immunogenicity of the second booster dose of mRNA-1273 (Part F, Cohort 2, 50 µg mRNA-1273).

**Part J**

Part J of the study will evaluate immunogenicity, safety, and reactogenicity of mRNA-1273.815 (50 µg) and mRNA-1273.231 (50 µg) administered as a booster to adults who previously received a primary series of mRNA vaccine, a first booster dose of a monovalent mRNA vaccine, and a second booster dose of a bivalent mRNA vaccine against SARS-CoV-2. Participants will be randomized in a 1:1 ratio to receive either mRNA-1273.815 (50 µg) or mRNA-1273.231 (50 µg).

Overall, study Parts A.1, B, C, and D will assess whether a single booster dose of the mRNA vaccines in each study part elicits antibody responses to ancestral SARS-CoV-2 and the variants similar to antibody responses to ancestral SARS-CoV-2 elicited by 2 doses of mRNA-1273 (100 µg) against ancestral SARS-CoV-2, using a historical control arm from the mRNA-1273-P301 (COVE) study.

Study Part F Cohort 1 will assess whether a single booster dose of the mRNA-1273.529 as the first booster dose elicits a similar or superior antibody response to the B.1.1.529 compared to a single booster dose of mRNA-1273, using an external historical comparator; Study Part F Cohort 2 will assess whether a single booster dose of the mRNA-1273.529 as a second booster dose elicits a similar or superior antibody response to the B.1.1.529 compared to a single booster dose of mRNA-1273 as a second booster dose.

Study Part G will assess whether a single booster dose of the mRNA-1273.214 as a second booster dose elicits a similar or superior antibody response to the B.1.1.529 compared to a single booster dose of mRNA-1273 as a second booster dose (Part F, Cohort 2, 50 µg mRNA-1273).

Study Part A.2 will evaluate the immunogenicity, safety, and reactogenicity of the mRNA-1273.214 vaccine candidate when administered as a second booster dose to participants who rolled over from Part A.1.

Study Part H will assess whether a single booster dose of the mRNA-1273.222 50 µg as a second booster dose elicits a superior antibody response against Omicron BA.4/5 compared to a single booster dose of mRNA-1273 as a second booster dose (Part F, Cohort 2, 50 µg mRNA-1273).

Part J will evaluate whether mRNA-1273.815 (50 µg) or mRNA-1273.231 (50 µg) administered as a booster will elicit antibody response against Omicron subvariants BA.4/BA.5, and XBB.1.5 in adults who previously received a primary series of mRNA vaccine, a first booster dose of a monovalent mRNA vaccine, and a second booster dose of a bivalent mRNA vaccine against SARS-CoV-2.

**Study Arms**

| Study Part | Study Arm | Dose[1] | N |
|---|---|---|---|
| Part A.1 | mRNA-1273.211 | 50 µg | ~300 |
| | mRNA-1273.211 | 100 µg | ~584 |
| Part A.2[2] | mRNA-1273.214 | 50 µg | ~300 |
| Part B | mRNA-1273 | 100 µg | ~300 |
| Part C | mRNA-1273.617.2 | 50 µg | ~584 |
| | mRNA-1273.617.2 | 100 µg | ~584 |
| Part D | mRNA-1273.213 | 50 µg | ~584 |
| | mRNA-1273.213 | 100 µg | ~584 |
| Part F (Cohort 1) | mRNA-1273.529 | 50 µg | ~375 |
| Part F (Cohort 2) | mRNA-1273.529 | 50 µg | ~375 |
| | mRNA-1273 | 50 µg | ~375 |
| Part G | mRNA-1273.214 | 50 µg | ~375 |
| Part H (2nd booster) | mRNA-1273.222 | 50 µg | ~500 |
| Part J (3rd booster)[3, 4] | mRNA-1273.815 | 50 µg | ~50 |
| | mRNA-1273.231 | 50 µg | ~50 |

1. Dose is total mRNA.
2. Participants rolled over from Part A.1 to Part A.2.

3. Participants may be rolled over from Part H.
4. Participants will be randomized in a 1:1 ratio to receive either mRNA-1273.815 or mRNA-1273.231.

Participants in Parts A.1, B, C, D, F, and G will have up to 7 visits; 6 visits if screening and dosing are performed on the same day. Study vaccine (mRNA-1273.211, mRNA-1273, mRNA-1273.617.2, mRNA-1273.213, mRNA-1273.529, or mRNA-1273.214) will be administered as a single dose on Day 1. Additional safety and/or immunogenicity study visits will occur on Days 15, 29, 91 (Parts F and G only), 181, and 366 (EoS). Study visits will include scheduled safety phone calls at Day 8, every 2 weeks from Day 43 to Day 169 and from Day 209 to Day 349 to collect AEs, MAAEs, AESI, AEs leading to withdrawal, SAEs, pregnancies, and information about concomitant medications and receipt of non-study vaccinations.

Participants in Part A.1 who choose to continue in Part A.2 will have 6 additional visits; 5 if screening and dosing are performed on the same day. Study vaccine (mRNA-1273.214 [50 µg]) will be administered as a single dose on Day 1. Additional safety and/or immunogenicity study visits will occur on Days 15, 29, 91, and 181 (EoS). Study visits will include scheduled safety phone calls at Day 8, and every 2 weeks from Day 43 to Day 169 to collect AEs, MAAEs, AESI, AEs leading to withdrawal, SAEs, pregnancies, and information about concomitant medications and receipt of non-study vaccinations.

Participants in Part H will have up to 6 visits; 5 visits if screening and dosing are performed on the same day. Study vaccine (mRNA-1273.222 [50 µg]) will be administered as a single dose on Day 1. Additional safety and/or immunogenicity study visits will occur on Days 15, 29, 91, and 181(EoS). Study visits will include scheduled safety phone calls at Day 8, and every 2 weeks from Day 43 to Day 169 to collect AEs, MAAEs, AESI, AEs leading to withdrawal, SAEs, pregnancies, and information about concomitant medications and receipt of non-study vaccinations.

Participants in Part J may include participants who have previously participated in Part H of this study. Participants in Part J will have up to 6 visits; 5 visits if screening and dosing are performed on the same day. Study vaccine (mRNA-1273.815 or mRNA-1273.231) will be administered as a single dose on Day 1. Additional safety and/or immunogenicity study visits will occur on Days 15, 29, 91, and 181 (EoS). Study visits will include scheduled safety phone calls at Day 8, and every 2 weeks from Day 43 to Day 169 to collect AEs, MAAEs, AESI, AEs leading to withdrawal, SAEs, pregnancies, and information about concomitant medications and receipt of non-study vaccinations. For participants who have previously participated in Part H of this study, the total number of visits completed in the study overall will be 10-12 depending on whether screening and dosing are performed on the same day.

At the dosing visit on Day 1, participants will be instructed how to document and report solicited ARs within a provided electronic diary (eDiary). Solicited ARs will be assessed for 7 days (the day of injection and the following 6 days), and unsolicited AEs will be assessed for 28 days after injection; SAEs, MAAEs, AEs leading to withdrawal, pregnancies, and AESI will be assessed throughout the study. All participants will be tested for the presence of SARS-CoV-2 antibodies at baseline and at Day 29 (primary immunogenicity endpoint). Participants in Part J will also be tested for the presence of SARS-CoV-2 antibodies at Day 15 (primary immunogenicity endpoint). Additional blood draws will be collected on Day 91 (Parts A.2, F, G, H, and J only), Day 181, and Day 366 (Parts A.1, B, C, D, F, and G only). In addition, active surveillance for intercurrent or breakthrough SARS-CoV-2 infection will occur throughout the study and reported as AEs (confirmed symptomatic infections will be reported as MAAEs if not SAEs). Participants with signs and symptoms meeting the CDC case definition for COVID-19 (21 Feb 2021 or most recent [https://www.cdc.gov/coronavirus/2019-ncov/symptoms-testing/symptoms.html]) will be asked to contact the site and undergo prompt assessment which will include RT-PCR testing (of a respiratory sample) to assess symptomatic COVID-19. Participants with any clinical or radiographic evidence of pneumonia will also undergo RT-PCR testing. Suspected COVID-19 cases will also be tested using a multiplex assay to assess for non–SARS-CoV-2 causes of upper or lower respiratory tract infection. Participants will have blood samples collected at scheduled study site visits during the study for immunogenicity assessments or other medical concerns according to the investigator's judgment.

Participants may experience AEs, to include symptoms of COVID-19, that necessitate an unscheduled visit. There may also be situations in which the investigator asks a participant to report for an unscheduled visit following the report of an AE. Additional examinations may be conducted at these visits as necessary to ensure the safety and well-being of participants during the study. Electronic case report forms should be completed for each unscheduled visit. In addition, participants may have blood samples collected at unscheduled visits for acute respiratory symptoms.

Peripheral blood mononuclear cells (PBMCs) may be collected for a subset of participants at selected sites at baseline (Day 1) and at Days 15, 29, and 91 to characterize the T-cell and B-cell responses against SARS-CoV-2 and variants.

Participants will be enrolled to receive 50 or 100 µg dose of mRNA-1273.211 (Part A.1), 100 µg mRNA-1273 (Part B), 50 or 100 µg mRNA-1273.617.2 (Part C), 50 or 100 µg mRNA-1273.213 (Part D), 50 µg mRNA-1273.529 or 50 µg mRNA-1273 (Part F), 50 µg mRNA-1273.214 (Part G and Part A.2), 50 µg mRNA-1273.222 (Part H), or 50 µg mRNA-1273.815 or 50 µg mRNA-1273.231 (Part J). The interim analysis will be conducted based on safety and immunogenicity data collected through Day 15 or Day 29. The interim analysis may be conducted either after all participants in Parts A.1, A.2, B, C, D, F, G, H, or J have completed their Day 15 or Day 29 visit assessments and/or subsequent timepoint visits (eg, Day 91 for Parts F, G, and H) or combined after the last participant in any of the study parts (Parts A.1, A.2, B, C, D, F, G, H, or J) or dose arm, or pre-specified subset of dose arm has completed their Day 15 or Day 29 visit assessments. The final study analysis after 12 months of follow-up will be completed for all participants in Parts A.1, B, C, D, F, and G. The final study analysis after 6 months of follow-up will be completed for all participants in Parts A.2, H, and J.

**Safety Oversight:** No safety monitoring committee or data safety monitoring board is planned for this study.

Safety monitoring for this study will include study team members, inclusive of, at a minimum, the Sponsor medical monitor, Sponsor safety physician (from Pharmacovigilance), and contract research organization medical monitor. The study team will conduct ongoing safety reviews during the study and will be responsible to monitor for safety concerns during the study as described in the Safety Management Plan.

An independent cardiac event adjudication committee that includes pediatric and adult cardiologists will review suspected cases of myocarditis and pericarditis to determine if they meet CDC criteria of "probable" or "confirmed" events, and to assess severity.

**Study Duration:** Approximately 12 months for each participant in Parts A.1, B, C, D, F, and G. Approximately 6 months for each participant in Parts A.2, H, and J.

**Number of Participants:**

**Part A.1:**

Approximately 300 participants will receive a single booster dose of mRNA-1273.211 50 µg, to achieve 270 evaluable participants in the 50-µg dose study arm. Approximately 584 participants will receive a single booster dose of mRNA-1273.211 100 µg, to achieve 526 evaluable participants in the 100-µg study arm.

**Part A.2**

Approximately 300 participants will receive a second booster dose of mRNA-1273.214 50 µg.

**Part B:**

Approximately 300 participants will receive a single booster dose of mRNA-1273 100 µg, to achieve 270 evaluable participants in Part B of the study.

**Part C:**

Approximately 584 participants will receive a single booster dose of mRNA-.1273-617.2 50 µg, to achieve 526 evaluable participants in the 50 µg dose study arm. Approximately 584 participants will receive a single booster dose of mRNA-1273.617.2 100 µg, to achieve 526 evaluable participants in the 100 µg dose study arm.

**Part D:**

Approximately 584 participants will receive a single booster dose of mRNA-1273.213 50 µg, to achieve 526 evaluable participants in the 50-µg dose study arm. Approximately 584 participants will receive a single booster dose of mRNA-1273.213 100 µg, to achieve 526 evaluable participants in the 100-µg dose study arm.

**Part F:**

Cohort 1:

Approximately 375 participants will receive a single booster dose of mRNA-1273.529 50 µg, to achieve 300 evaluable participants in the 50-µg dose study arm.

Cohort 2:

Approximately 375 participants will receive a second booster dose of mRNA-1273.529 50 µg. In addition, approximately 375 participants will receive a second booster dose of mRNA-1273 50 µg to achieve 300 evaluable participants in each arm (sequential enrollment for the 2 groups in Cohort 2).

**Part G:**

Approximately 375 participants will receive a second booster dose of mRNA-1273.214 50 µg, to achieve 300 evaluable participants in mRNA-1273.214 50 µg arm.

**Part H:**

Approximately 500 participants will receive a second booster dose of mRNA-1273.222 50 µg to achieve 300 evaluable participants.

**Part J:**

Approximately 100 participants will be randomized in a 1:1 ratio to receive a single booster dose of mRNA-1273.815 (50 µg) or mRNA-1273.231 (50 µg).

**Study Eligibility Criteria**

**Inclusion Criteria:**

Each participant must meet all of the following criteria to be enrolled in this study:

1. Male or female, at least 18 years of age at the time of consent (Screening Visit).
2. Investigator's assessment that participant understands and is willing and physically able to comply with protocol-mandated follow-up, including all procedures.
3. Participant has provided written informed consent for participation in this study, including all evaluations and procedures as specified in this protocol.
4. Female participants of nonchildbearing potential may be enrolled in the study. Nonchildbearing potential is defined as surgically sterile (history of bilateral tubal ligation, bilateral oophorectomy, hysterectomy) or postmenopausal (defined as amenorrhea for ≥ 12 consecutive months prior to screening [Day 0] without an alternative medical cause). A follicle-stimulating hormone level may be measured at the discretion of the investigator to confirm postmenopausal status.
5. Female participants of childbearing potential may be enrolled in the study if the participant fulfills all of the following criteria:
   - Has a negative pregnancy test on the day of vaccination (Day 1).
   - Has practiced adequate contraception or has abstained from all activities that could result in pregnancy for at least 28 days prior to Day 1.
   - Has agreed to continue adequate contraception through 3 months following vaccination.
   - Is not currently breastfeeding.

   Adequate female contraception is defined as consistent and correct use of a Food and Drug Administration (FDA)-approved contraceptive method in accordance with the product label.
6. Participant must have been either previously enrolled in the mRNA-1273-P301 (COVE) study, must have received 2 doses of mRNA-1273 in that study, with his/her second dose at least 6 months prior to enrollment in mRNA-1273-P205, and must be currently enrolled and compliant in that study (ie, has not withdrawn or discontinued early); or participant must have received 2 doses of mRNA-1273 under the EUA with their second dose at least 6 months prior to enrollment in mRNA-1273 P205; or have received a 2 dose primary series of mRNA-1273 followed by a 50 µg booster dose of mRNA-1273 in the mRNA-1273-P301 (COVE) study or under EUA at least 3 months prior to enrollment in mRNA-1273-P205; and able to provide proof of vaccination status at the time of screening (Day 1); or for enrollment in Part A.2, participant must be currently enrolled and compliant in Part A.1 of the mRNA-1273-P205 study and must have received their first booster dose of mRNA-1273.211 50 µg; or for enrollment in Part J, participant must meet at least one of the following criteria:
   - Completed enrollment in Part H of the mRNA-1273-P205 study; or
   - Received a 2-dose primary series of mRNA-1273 (100 µg) followed by a 50 µg booster dose of mRNA-1273 in the mRNA-1273-P301 (COVE) study or under EUA, followed by a 50 µg booster dose of mRNA-1273.222 under EUA at least 3 months prior to enrollment in Part J of mRNA-1273-P205; or

- Previously received a 2-dose primary series of mRNA vaccine against SARS-CoV-2 followed by a booster dose of a monovalent mRNA vaccine, followed by a second booster dose of a bivalent mRNA vaccine.

Participants in Part J must also provide proof of vaccination status at the time of screening (Day 0 or Day 1).

**Exclusion Criteria:**

Participants meeting any of the following criteria at the Screening Visit, unless noted otherwise, will be excluded from the study:

1. Had significant exposure to someone with SARS-CoV-2 infection or coronavirus disease 2019 (COVID-19) in the past 14 days, as defined by the CDC as a close contact of someone who has COVID-19).
2. Has known history of SARS-CoV-2 infection within 3 months prior to enrollment.
3. Is acutely ill or febrile (temperature ≥ 38.0°C [100.4°F]) less than 72 hours prior to or at the Screening Visit or Day 1. Participants meeting this criterion may be rescheduled and will retain their initially assigned participant number.
4. Currently has symptomatic acute or unstable chronic disease requiring medical or surgical care, to include significant change in therapy or hospitalization for worsening disease, at the discretion of the investigator.
5. Has a medical, psychiatric, or occupational condition that may pose additional risk as a result of participation, or that could interfere with safety assessments or interpretation of results according to the investigator's judgment.
6. Has a current or previous diagnosis of immunocompromising condition to include human immunodeficiency virus, immune-mediated disease requiring immunosuppressive treatment, or other immunosuppressive condition.
7. Has received systemic immunosuppressants or immune-modifying drugs for > 14 days in total within 6 months prior to screening (for corticosteroids ≥ 10 mg/day of prednisone equivalent) or is anticipating the need for immunosuppressive treatment at any time during participation in the study.
8. Has known or suspected allergy or history of anaphylaxis, urticaria, or other significant AR to the vaccine or its excipients.
9. Has a documented history of myocarditis or pericarditis within 2 months prior to Screening Visit (Day 0).
10. Coagulopathy or bleeding disorder considered a contraindication to intramuscular (IM) injection or phlebotomy.
11. Has received or plans to receive any licensed vaccine ≤ 28 days prior to the injection (Day 1) or a licensed vaccine within 28 days before or after the study injection, with the exception of influenza vaccines, which may be given 14 days before or after receipt of a study vaccine.
12. Has received systemic immunoglobulins or blood products within 3 months prior to the Screening Visit (Day 0) or plans for receipt during the study.
13. Has donated ≥ 450 mL of blood products within 28 days prior to the Screening Visit or plans to donate blood products during the study.
14. Plans to participate in an interventional clinical trial of an investigational vaccine or drug while participating in this study.

15. Is an immediate family member or household member of study personnel, study site staff, or Sponsor personnel.
16. Is currently experiencing an SAE in Study mRNA-1273-P301 (COVE) at the time of screening for this study.

**Study Treatments**

**Investigational Product, Dosage, and Mode of Administration:**

**Part A.1 (mRNA-1273.211)**

mRNA-1273.211 is a multivalent product that contains 2 mRNAs: CX-024414 encoding for the prefusion stabilized spike protein (S-2P) of Wuhan-Hu-1 and CX-027367 encoding for the S-2P of B.1.351, in a 1:1 ratio. mRNA-1273.211 consists of the mRNA formulated in a mixture of 4 lipids common to the Sponsor's mRNA vaccine platform: SM-102, cholesterol, 1,2-distearoyl-sn-glycero-3-phosphocholine (DSPC), and 1-monomethoxypolyethyleneglycol-2,3-dimyristylglycerol with polyethylene glycol of average molecular weight 2000 (PEG-2000-DMG).

mRNA-1273.211 injection will be provided as a sterile liquid for injection, white to off-white dispersion in appearance, at a concentration of 0.2 mg/mL in 20 mM Tris buffer containing 87 mg/mL sucrose and 4.3 mM sodium acetate at pH 7.5.

mRNA-1273.211 will be administered at 50 and 100 µg dose levels. Doses will be administered by IM injection according to the procedures specified in the mRNA-1273-P205 Pharmacy Manual. Preferably, the vaccine should be administered into the nondominant arm.

**Part A.2 (mRNA-1273.214)**

mRNA-1273.214 contains CX-024414, the mRNA that encodes for the prefusion stabilized spike glycoprotein (S-2P) of the Wuhan-Hu-1 isolate of SARS-CoV-2 and CX-031302, the mRNA that encodes for the S-2P of the SARS-CoV-2 B.1.1.529 variant. mRNA-1273.214 consists of each mRNA formulated in a mixture of 4 lipids common to the Sponsor's mRNA vaccine platform: SM-102, cholesterol, DSPC, and PEG2000-DMG.

The formulated mRNA are mixed in a 1:1 ratio. mRNA-1273.214 injection will be provided as a sterile liquid for injection, white to off-white dispersion in appearance, at a concentration of 0.1 mg/ML in 20 mM Tris buffer containing 87 mg/mL sucrose and 2.1 mM acetate at pH 7.5.

mRNA-1273 will be administered at a 50 µg dose level. Doses will be administered by IM injection according to the procedures specified in the mRNA-1273-P205 Pharmacy Manual. Preferably, the vaccine should be administered into the nondominant arm.

**Part B (mRNA-1273)**

mRNA-1273 contains mRNA CX-024414 encoding for the S-2P of Wuhan-Hu-1. mRNA-1273 consists of the mRNA formulated in a mixture of 4 lipids common to the Sponsor's mRNA vaccine platform: SM-102, cholesterol, DSPC and 1 PEG-2000-DMG.

mRNA-1273 injection will be provided as a sterile liquid for injection, white to off-white dispersion in appearance, at a concentration of 0.2 mg/mL in 20 mM Tris buffer containing 87 mg/mL sucrose and 4.3 mM sodium acetate at pH 7.5.

mRNA-1273 will be administered at a 100 µg dose level. Doses will be administered by IM injection according to the procedures specified in the mRNA-1273-P205 Pharmacy Manual. Preferably, the vaccine should be administered into the nondominant arm.

**Part C (mRNA-1273.617.2)**

mRNA-1273.617.2 contains mRNA CX-029444 encoding for the S-2P of B.1.617.2. mRNA-1273.617.2 consists of the mRNA formulated in a mixture of 4 lipids common to the Sponsor's mRNA vaccine platform: SM-102, cholesterol, DSPC and PEG-2000-DMG.

mRNA-1273.617.2 injection will be provided as a sterile liquid for injection, white to off-white dispersion in appearance, at a concentration of 0.2 mg/mL in 20 mM Tris buffer containing 87 mg/mL sucrose and 4.3 mM sodium acetate at pH 7.5.

mRNA-1273.617.2 will be administered at 50 µg and 100 µg dose level. Doses will be administered by IM injection according to the procedures specified in the mRNA-1273-P205 Pharmacy Manual. Preferably, the vaccine should be administered into the nondominant arm.

**Part D (mRNA-1273.213)**

mRNA-1273.213 is a multivalent product that contains 2 mRNAs: CX-029444 encoding for the S-2P of B.1.617.2 and CX-027367 encoding for the S-2P of B.1.351, in a 1:1 ratio. mRNA-1273.213 consists of the mRNA formulated in a mixture of 4 lipids common to the Sponsor's mRNA vaccine platform: SM-102, cholesterol, DSPC and PEG-2000-DMG.

mRNA-1273.213 injection will be provided as a sterile liquid for injection, white to off-white dispersion in appearance, at a concentration of 0.2 mg/mL in 20 mM Tris buffer containing 87 mg/mL sucrose and 4.3 mM sodium acetate at pH 7.5.

mRNA-1273.213 will be administered at 50 µg and 100 µg dose level. Doses will be administered by IM injection according to the procedures specified in the mRNA-1273-P205 Pharmacy Manual. Preferably, the vaccine should be administered into the nondominant arm.

**Part F (mRNA-1273.529)**

mRNA-1273.529 contains mRNA CX-031302 encoding for the S-2P of B.1.1.529. mRNA-1273.529 consists of the mRNA formulated in a mixture of 4 lipids common to the Sponsor's mRNA vaccine platform: SM-102, cholesterol, DSPC and PEG-2000-DMG.

mRNA-1273.529 injection will be provided as a sterile liquid for injection, white to off-white dispersion in appearance, at a concentration of 0.2 mg/mL in 20 mM Tris buffer containing 87 mg/mL sucrose and 4.3 mM sodium acetate at pH 7.5.

mRNA-1273.529 will be administered at a 50 µg dose level. Doses will be administered by IM injection according to the procedures specified in the mRNA-1273-P205 Pharmacy Manual. Preferably, the vaccine should be administered into the nondominant arm.

**Part G (mRNA-1273.214)**

mRNA-1273.214 contains CX-024414, the mRNA that encodes for the prefusion stabilized spike glycoprotein (S-2P) of the Wuhan-Hu-1 isolate of SARS-CoV-2 and CX-031302, the mRNA that encodes for the S-2P of the SARS-CoV-2 B.1.1.529 variant. mRNA-1273.214 consists of each mRNA formulated in a mixture of 4 lipids common to the Sponsor's mRNA vaccine platform: SM-102, cholesterol, DSPC, and PEG2000-DMG.

The formulated mRNA are mixed in a 1:1 ratio. mRNA-1273.214 injection will be provided as a sterile liquid for injection, white to off-white dispersion in appearance, at a concentration of 0.1 mg/ML in 20 mM Tris buffer containing 87 mg/mL sucrose and 2.1 mM acetate at pH 7.5.

mRNA-1273 will be administered at a 50 µg dose level. Doses will be administered by IM injection according to the procedures specified in the mRNA-1273-P205 Pharmacy Manual. Preferably, the vaccine should be administered into the nondominant arm.

**Part H (mRNA-1273.222)**

mRNA-1273.222 contains CX-024414, the mRNA that encodes for the prefusion stabilized spike glycoprotein (S-2P) of the Wuhan-Hu-1 isolate of SARS-CoV-2 and CX-034476, the mRNA that encodes for the S-2P of the SARS-CoV-2 B.1.1.529 subvariants BA.4/BA.5. mRNA-1273.222 consists of each mRNA formulated in a mixture of 4 lipids common to the Sponsor's mRNA vaccine platform: SM-102, cholesterol, DSPC, and PEG2000-DMG.

The formulated mRNA are mixed in a 1:1 ratio. mRNA-1273.222 injection will be provided as a sterile liquid for injection, white to off-white dispersion in appearance, at a concentration of 0.1 mg/mL in 20 mM Tris buffer containing 87 mg/mL sucrose and 2.1 mM acetate at pH 7.5.

mRNA-1273.222 will be administered at a 50 µg dose level. Doses will be administered by IM injection according to the procedures specified in the mRNA-1273-P205 Pharmacy Manual. Preferably, the vaccine should be administered into the nondominant arm.

**Part J (mRNA-1273.815 and mRNA-1273.231)**

mRNA-1273.815 contains CX-038839, the monovalent mRNA that encodes for the S-2P of the SARS-CoV-2 Omicron subvariants XBB.1.5/XBB.1.9.1. mRNA-1273.231 contains CX-034476, the mRNA that encodes for the S-2P of the SARS-CoV-2 B.1.1.529 Omicron subvariants BA.4/BA.5, and CX-038839, the mRNA that encodes for the S-2P of the SARS-CoV-2 subvariants XBB.1.5/XBB.1.9.1. The formulated mRNA in mRNA-1273.231 are mixed in a 1:1 ratio. Both mRNA-1273.815 and mRNA-1273.231 consist of mRNA formulated in a mixture of 4 lipids common to the Sponsor's mRNA vaccine platform: SM-102, cholesterol, DSPC, and PEG2000-DMG.

mRNA-1273.815 and mRNA-1273.231 will be administered at a 50 µg dose level. Doses will be administered by IM injection according to the procedures specified in the mRNA-1273-P205 Pharmacy Manual. Preferably, the vaccine should be administered into the nondominant arm.

mRNA-1273.231 and mRNA-1273.815 injections will be provided as a sterile liquid for injection, white to off-white dispersion in appearance, at a concentration of 0.1 mg/mL in 20 mM Tris buffer containing 87 mg/mL sucrose and 2.1 mM acetate at pH 7.5.

**Procedures and Assessments:**

**Safety Assessments:**

Safety assessments will include monitoring and recording of the following for each participant:

- Solicited local and systemic ARs that occur during the 7 days following vaccination (ie, the day of injection and 6 subsequent days). Solicited ARs will be recorded daily using eDiaries.
- Unsolicited AEs observed or reported during the 28 days following vaccination (ie, the day of injection and 27 subsequent days).
- AEs leading to withdrawal from Day 1 through EoS or withdrawal from the study.
- MAAEs from vaccination on Day 1 through EoS or withdrawal from the study.
- AESI from vaccination on Day 1 through EoS or withdrawal from the study.
- SAEs from vaccination on Day 1 through EoS or withdrawal from the study.
- Vital sign measurements before and after vaccination.
- Physical examination findings (if performed).

- Assessments for SARS-CoV-2 infection from Day 1 through study completion.
- Details of all pregnancies in female participants will be collected after the start of study treatment and until the end of their participation in the study.

The incidence and severity of the above events will be monitored by an independent safety team (IST) on a regular basis.

**Immunogenicity Assessments:**

Blood samples for immunogenicity assessments will be collected at the time points indicated in the SoE. The following immunogenicity assessments will be measured contemporaneously with Day 57 samples from participants in the mRNA-1273-P301 (COVE) study (historical control samples; further details will be provided in the SAP):

- Serum binding antibody (bAb) level against SARS-CoV-2 as measured by ligand binding assay specific to the SARS-CoV-2 S protein and the S protein receptor-binding domain (RBD).
- Serum neutralizing antibody (nAb) level against SARS-CoV-2 as measured by pseudovirus neutralization assays.
- Testing for serologic markers for SARS-CoV-2 infection as measured by anti-nucleocapsid antibodies detected by immunoassay (Day 1, Day 29, Day 91 [Parts A.2, Part F and Part G only], Day 181, and Day 366 [Parts A.1, B, C, D, F and G only]). Prior results from the mRNA-1273-P301 (COVE) study, specifically Day 1 and 57 samples, tested in validated ligand binding and neutralizing antibody assays, will be used for comparison in primary and secondary endpoints in this trial.
- Sequencing of SARS-CoV-2 genome in samples positive for viral infection by PCR testing.
- PBMCs may be collected for a subset of participants at selected sites to characterize T-cell and B-cell response against SARS-CoV-2 and variants.

**Statistical Methods:**

**Part A.1:**

Each dose level of mRNA-1273.211 booster dose (50 µg, 100 µg) will be assessed with respect to mRNA-1273 primary series historical control.

**Hypotheses:** For the primary immunogenicity objective, there are 4 null hypotheses to be tested for each arm.

A. $H_0^1$: 50 or 100 µg mRNA-1273.211, as a single booster dose, is inferior to the primary series (100 µg) of mRNA-1273 based on GMT ratio against ancestral SARS-CoV-2 with a non-inferiority margin of 1.5.

B. $H_0^2$: 50 or 100 µg mRNA-1273.211, as a single booster dose, is inferior to the primary series of (100 µg) mRNA-1273 based on difference in SRR against ancestral SARS-CoV-2 with a non-inferiority margin of 10%.

C. $H_0^3$: 50 or 100 µg mRNA-1273.211 against the variant B.1.351 is inferior to the primary series of (100 µg) mRNA-1273 against ancestral SARS-CoV-2 based on the GMT ratio of mRNA-1273.211 against the variant B.1.351 at Day 29 compared to mRNA-1273 against ancestral SARS-CoV-2 at Day 57 with a non-inferiority margin of 1.5.

D. $H_0^4$: 50 or 100 µg mRNA-1273.211 against the variant B.1.351 is inferior to the primary series of (100 µg) mRNA-1273 against ancestral SARS-CoV-2 based on the difference in SRR (SRR of mRNA-1273.211 against the variant B.1.351 at Day 29 – SRR of mRNA-1273 against ancestral SARS-CoV-2 at Day 57) with a non-inferiority margin of 10%.

**Part A.2:**

No statistical hypothesis testing will be performed for Part A.2.

**Part B:**

For the primary immunogenicity objective for a single booster dose of 100 µg mRNA-1273.

A. $H_0^1$: 100 µg mRNA-1273, as a single booster dose, is inferior to the primary series of (100 µg) mRNA-1273 based on GMT ratio against ancestral SARS-CoV-2 with a non-inferiority margin of 1.5.

B. $H_0^2$: 100 µg mRNA-1273, as a single booster dose, is inferior to the primary series of (100 µg) mRNA-1273 based on difference in SRR against ancestral SARS-CoV-2 with a non-inferiority margin of 10%.

**Part C:**

Each dose level of mRNA-1273.617.2 booster dose (50 µg, 100 µg) will be assessed with respect to mRNA-1273 primary series historical control.

For the primary immunogenicity objective, there are 2 null hypotheses to be tested for each arm.

A. $H_0^1$: 50 µg or 100 µg mRNA-1273.617.2, as a single booster dose, against the variant B.1.617.2 is inferior to the primary series of (100 µg) mRNA-1273 against ancestral SARS-CoV-2 based on the GMT ratio of mRNA-1273.617.2 against the variant B.1.617.2 at Day 29 compared to mRNA-1273 against ancestral SARS-CoV-2 at Day 57 with a non-inferiority margin of 1.5.

B. $H_0^2$: 50 µg or 100 µg mRNA-1273.617.2, as a single booster dose, against the variant B.1.617.2 is inferior to the primary series of (100 µg) mRNA-1273 against ancestral SARS-CoV-2 based on the difference in SRR (SRR of mRNA-1273.617.2 against the variant B.1.617.2 at Day 29 – SRR of mRNA-1273 against ancestral SARS-CoV-2 at Day 57) with a non-inferiority margin of 10%.

**Part D:**

Each dose level of mRNA-1273.213 booster dose (50 µg, 100 µg) will be assessed with respect to mRNA-1273 primary series historical control.

For the primary immunogenicity objective, there are 4 null hypotheses to be tested for each arm.

A. $H_0^1$: 50 µg or 100 µg mRNA-1273.213, as a single booster dose, against the variant B.1.617.2 is inferior to the primary series of (100 µg) mRNA-1273 against ancestral SARS-CoV-2 based on the GMT ratio of mRNA-1273.213 against the variant B.1.617.2 at Day 29 compared to mRNA-1273 against ancestral SARS-CoV-2 at Day 57 with a non-inferiority margin of 1.5.

B. $H_0^2$: 50 µg or 100 µg mRNA-1273.213, as a single booster dose, against the variant B.1.617.2 is inferior to the primary series of (100 µg) mRNA-1273 against ancestral SARS-CoV-2 based on the difference in SRR (SRR of mRNA-1273.213 against the variant B.1.617.2 at Day 29 – SRR of mRNA-1273 against ancestral SARS-CoV-2 at Day 57) with a non-inferiority margin of 10%.

C. $H_0^3$: 50 µg or100 µg mRNA-1273.213, as a single booster dose, against the variant B.1.351 is inferior to the primary series of (100 µg) mRNA-1273 against ancestral SARS-CoV-2 based on the GMT ratio of mRNA-1273.213 against the variant B.1.351 at Day 29 compared to mRNA-1273 against ancestral SARS-CoV-2 at Day 57 with a non-inferiority margin of 1.5.

D. $H_0^4$: 50 µg or100 µg mRNA-1273.213, as a single booster dose, against the variant B.1.351 is inferior to the primary series of (100 µg) mRNA-1273 against ancestral SARS-CoV-2 based on the difference in SRR (SRR of mRNA-1273.213 against the variant B.1.351 at Day 29 – SRR of mRNA-1273 against ancestral SARS-CoV-2 at Day 57) with a non-inferiority margin of 10%.

**Part F:**

Cohort 1:

50 µg mRNA-1273.529 booster dose (as a first booster dose) will be assessed with respect to mRNA-1273 booster dose (as a first booster dose) using an external comparator (details regarding the external historical comparator will be included in the SAP).

For the primary immunogenicity objectives, there are 3 hypotheses to be tested:

A. $H_1^1$: 50 µg mRNA-1273.529, as a single booster dose, against the variant B.1.1.529 is non-inferior to the booster dose of (50 µg) mRNA-1273 against B.1.1.529 based on the GMT ratio of mRNA-1273.529 against the variant B.1.1.529 at Day 29 compared to mRNA-1273 against the variant B.1.1.529 at Day 29 with a non-inferiority margin of 1.5.

B. $H_1^2$: 50 µg mRNA-1273.529, as a single booster dose, against the variant B.1.1.529 is non-inferior to the booster dose of (50 µg) mRNA-1273 against B.1.1.529 based on the difference in SRR at Day 29 with a non-inferiority margin of 10%.

C. $H_1^3$: 50 µg mRNA-1273.529, as a single booster dose, against the variant B.1.1.529 is superior to the booster dose of (50 µg) mRNA-1273 against B.1.1.529 based on the GMT ratio of mRNA-1273.529 against the variant B.1.1.529 at Day 29 compared to mRNA-1273 against the variant B.1.1.529 at Day 29.

The primary immunogenicity objective is considered met if non-inferiority against B.1.1.529 (based on GMR and SRR difference) is demonstrated.

Cohort 2:

50 µg mRNA-1273.529 as the second booster dose will be compared to 50 µg mRNA-1273 as the second booster dose.

For the primary immunogenicity objective, there are 3 hypotheses to be tested:

A. $H_1^1$: 50 µg mRNA-1273.529, as a second booster dose, against the variant B.1.1.529 is non-inferior to the second booster dose of (50 µg) mRNA-1273 against B.1.1.529 based on the GMT ratio of mRNA-1273.529 against the variant B.1.1.529 at Day 29 compared to mRNA-1273 against the variant B.1.1.529 at Day 29 with a non-inferiority margin of 1.5.

B. $H_1^2$: 50 µg mRNA-1273.529, as a second booster dose, against the variant B.1.1.529 is non-inferior to the second booster dose of (50 µg) mRNA-1273 against B.1.1.529 based on the difference in SRR at Day 29 with a non-inferiority margin of 10%.

C. $H_1^3$: 50 µg mRNA-1273.529, as a second booster dose, against the variant B.1.1.529 is superior to the second booster dose of (50 µg) mRNA-1273 against B.1.1.529 based on the GMT ratio of mRNA-1273.529 against the variant B.1.1.529 at Day 29 compared to mRNA-1273 against the variant B.1.1.529 at Day 29.

The primary immunogenicity objective is considered met if non-inferiority against B.1.1.529 (based on GMR and SRR difference) is demonstrated.

**Part G:**

50 µg mRNA-1273.214 as the second booster dose will be compared to 50 µg mRNA-1273 as the second booster dose (active control arm in Part F, Cohort 2)

For the primary immunogenicity objective, there are 8 hypotheses (4 hypotheses at Day 29 and 4 hypotheses at Day 91).

A. $H_1^1$: 50 µg mRNA-1273.214, as a second booster dose, against the variant B.1.1.529 is non-inferior to the second booster dose of (50 µg) mRNA-1273 against B.1.1.529 based on the GMT ratio of mRNA-1273.214 against the variant B.1.1.529 at Day 29 compared to mRNA-1273 against the variant B.1.1.529 at Day 29 with a non-inferiority margin of 1.5.

B. $H_1^2$: 50 µg mRNA-1273.214, as a second booster dose, against the variant B.1.1.529 is non-inferior to the second booster dose of (50 µg) mRNA-1273 against B.1.1.529 based on the difference in SRR at Day 29 with a non-inferiority margin of 10%.

C. $H_1^3$: 50 µg mRNA-1273.214, as a second booster dose, against ancestral SARS-CoV-2 is non-inferior to the second booster dose of (50 µg) mRNA-1273 against ancestral SARS-CoV-2 based on the GMT ratio of mRNA-1273.214 against ancestral SARS-CoV-2 at Day 29 compared to mRNA-1273 against ancestral SARS-CoV-2 at Day 29 with a non-inferiority margin of 1.5.

D. $H_1^4$: 50 µg mRNA-1273.214, as a second booster dose, against the variant B.1.1.529 is superior to the second booster dose of (50 µg) mRNA-1273 against B.1.1.529 based on the GMT ratio of mRNA-1273.214 against the variant B.1.1.529 at Day 29 compared to mRNA-1273 against the variant B.1.1.529 at Day 29.

E. $H_1^5$: 50 µg mRNA-1273.214, as a second booster dose, against the variant B.1.1.529 is non-inferior to the second booster dose of (50 µg) mRNA-1273 against B.1.1.529 based on the GMT ratio of mRNA-1273.214 against the variant B.1.1.529 at Day 91 compared to mRNA-1273 against the variant B.1.1.529 at Day 91 with a non-inferiority margin of 1.5.

F. $H_1^6$: 50 µg mRNA-1273.214, as a second booster dose, against the variant B.1.1.529 is non-inferior to the second booster dose of (50 µg) mRNA-1273 against B.1.1.529 based on the difference in SRR at Day 91 with a non-inferiority margin of 10%.

G. $H_1^7$: 50 µg mRNA-1273.214, as a second booster dose, against the ancestral SARS-CoV-2 is non-inferior to the second booster dose of (50 µg) mRNA-1273 against ancestral SARS-CoV-2 based on the GMT ratio of mRNA-1273.214 against ancestral SARS-CoV-2 at Day 91 compared to mRNA-1273 against ancestral SARS-CoV-2 at Day 91 with a non-inferiority margin of 1.5.

H. $H_1^8$: 50 µg mRNA-1273.214, as a second booster dose, against the variant B.1.1.529 is superior to the second booster dose of (50 µg) mRNA-1273 against B.1.1.529 based on the GMT ratio of mRNA-1273.214 against the variant B.1.1.529 at Day 91 compared to mRNA-1273 against the variant B.1.1.529 at Day 91.

For the primary immunogenicity objective, an alpha of 0.05 (two-sided) will be allocated to the 2 time points (Day 29 and Day 91). Day 29 and Day 91 will each have an alpha of 0.025 (two-sided) for hypotheses testing.

The primary immunogenicity objective is considered met if non-inferiority against B.1.1.529 based on GMR, SRR difference, and non-inferiority against the ancestral SARS-CoV-2 based on GMR are demonstrated at Day 29 or Day 91.

For the key secondary immunogenicity objective, there are 2 hypotheses to be tested (Day 29 and 91 will each have an alpha of 0.025 [two-sided] for hypotheses testing):

A. $H_1^9$: 50 µg mRNA-1273.214, as a second booster dose, against ancestral SARS-CoV-2 is non-inferior to the booster dose of (50 µg) mRNA-1273 against ancestral SARS-CoV-2 based on the difference in SRR at Day 29 with a non-inferiority margin of 10%.
B. $H_1^{10}$: 50 µg mRNA-1273.214, as a second booster dose, against ancestral SARS-CoV-2 is non-inferior to the booster dose of (50 µg) mRNA-1273 against ancestral SARS-CoV-2 based on the difference in SRR at Day 91 with a non-inferiority margin of 10%.

**Part H**

50 µg mRNA-1273.222 given as a second booster dose will be compared to 50 µg mRNA-1273 given as a second booster dose (in Part F, Cohort 2).

For the primary immunogenicity objective, there are 5 hypotheses to be tested as shown below:

A. $H_1^1$: 50 µg mRNA-1273.222, as a second booster dose, against Omicron BA.4/5 is non-inferior to the second booster dose of (50 µg) mRNA-1273 against Omicron BA.4/5 based on the GMT ratio of mRNA-1273.222 against Omicron BA.4/5 at Day 29 compared to mRNA-1273 against Omicron BA.4/5 at Day 29 with a non-inferiority margin of 1.5.
B. $H_1^2$: 50 µg mRNA-1273.222, as a second booster dose, against Omicron BA.4/5 is non-inferior to the second booster dose of (50 µg) mRNA-1273 against Omicron BA.4/5 based on the difference in SRR at Day 29 with a non-inferiority margin of 5%.
C. $H_1^3$: 50 µg mRNA-1273.222, as a second booster dose, against ancestral SARS-CoV-2 D614G is non-inferior to the second booster dose of (50 µg) mRNA-1273 against ancestral SARS-CoV-2 D614G based on the GMT ratio of mRNA-1273.222 against ancestral SARS-CoV-2 D614G at Day 29 compared to mRNA-1273 against ancestral SARS-CoV-2 D614G at Day 29 with a non-inferiority margin of 1.5.
D. $H_1^4$: 50 µg mRNA-1273.222, as a second booster dose, against ancestral SARS-CoV-2 D614G is non-inferior to the second booster dose of (50 µg) mRNA-1273 against ancestral SARS-CoV-2 D614G based on the difference in SRR at Day 29 with a non-inferiority margin of 10%.
E. $H_1^5$: 50 µg mRNA-1273.222, as a second booster dose, against Omicron BA.4/5 is superior to the second booster dose of (50 µg) mRNA-1273 against Omicron BA.4/5 based on the GMT ratio of mRNA-1273.222 against Omicron BA.4/5 at Day 29 compared to mRNA-1273 against Omicron BA.4/5 at Day 29.

**Sample Size:**

**Part A.1**

Each dose level of mRNA-1273.211 will be assessed at a 2-sided type I error rate of 5%

The target enrollment of the 50-µg mRNA-1273.211 arm is approximately 300 participants. Assuming 10% of participants will be excluded from the Per-Protocol (PP) Set for Immunogenicity, with approximately 270 participants in the mRNA-1273.211 50 µg study arm and from the mRNA-1273 primary series historical control arm in the PP Set for Immunogenicity, there is approximately 75% power to reject all null hypotheses for the primary objectives. With approximately 270 participants in the mRNA-1273.211 50 µg study arm and 526 participants from the mRNA-1273 primary series historical control arm, there is approximately 90% power to reject all null hypotheses for the primary objectives based on ratio of geometric mean titers (GMR) and the difference in SRR against ancestral SARS-CoV-2 and variant (B.1.351) virus strain at 2-sided alpha of 5.0%. The assumptions are: the true GMR (50 µg mRNA-1273.211 vs. 100 µg mRNA-1273) against ancestral SARS-CoV-2 and variant virus strain is 1, the standard deviation of the log-transformed titer is 1.5, with a non-inferiority margin of 1.5; the true SRR against ancestral SARS-CoV-2 and variant virus strain after a single booster dose of mRNA-1273.211 is 90%, SRR against ancestral SARS-CoV-2 after mRNA-1273 primary series is 90%, and non-inferiority margin for SRR difference is 10%.

The target enrollment of 100 µg mRNA-1273.211 arm is approximately 584 participants. Assuming 10% of participants will be excluded from the PP Set for Immunogenicity, with approximately 526 participants in mRNA-1273.211 100 µg and mRNA-1273 primary series historical control arm respectively in the PP Set for Immunogenicity, there is approximately 80% power to reject all null hypotheses for the primary objectives based on GMR and difference in SRR against ancestral SARS-CoV-2 at 2-sided alpha of 5.0%. The assumptions are: the true GMR against ancestral SARS-CoV-2 and the variant (B.1.351) is 0.9, the standard deviation of the log-transformed titer is 1.5, non-inferiority margin for GMR is 1.5; the true SRR against ancestral SARS-CoV-2 and variant after a single booster dose of mRNA-1273.211 is 90%, and SRR against ancestral SARS-CoV-2 and variant after mRNA-1273 primary series is 90%, and non-inferiority margin for SRR difference is 10%.

Approximately 584 participants in the mRNA-1273-P301 (COVE) study will be selected as the mRNA-1273 primary series historical control. The same historical comparator arm will be used for the subsequent Parts B, C, and D.

With approximately 300 and 584 participants exposed to 50 and 100 µg of mRNA-1273.211, respectively, there is at least 90% probability to observe one participant at each dose level reporting an AE if the true rate of AEs is 1%.

**Part A.2**

We anticipate approximately 300 participants will be enrolled in Part A.2, there is no statistical hypothesis testing in Part A.2.

**Part B**

The target enrollment of the 100 µg mRNA-1273 arm is approximately 300 participants. Assuming 10% of participants will be excluded from the PP Set for Immunogenicity, with approximately 270 participants in the 100 µg mRNA-1273 study arm and approximately 526 from the mRNA-1273 (primary series) historical control arm in the PP Set for Immunogenicity, there is >90% power to reject both null hypotheses for the primary objective based on GMR and the difference in SRR against ancestral SARS-CoV-2 (mRNA-1273) at 2-sided alpha of 5.0%. The assumptions are: the true GMR (100 µg mRNA-1273 booster vs. 100 µg mRNA-1273 primary series historical control) against ancestral SARS-CoV-2 is 1.5, the standard deviation of the log-transformed titer is 1.5, with a non-inferiority margin of 1.5; the true SRR against ancestral SARS-CoV-2 after a single booster dose of mRNA-1273 is 90%, SRR against ancestral SARS-CoV-2 after mRNA-1273 primary series is 90%, and non-inferiority margin for SRR difference is 10%.

With approximately 300 participants exposed to 100 µg of mRNA-1273, there is at least 90% probability to observe one participant reporting an AE if the true rate of AEs is 1%.

**Part C**

Each dose level of mRNA-1273.617.2 will be assessed at a 2-sided type I error rate of 5%.

The target enrollment is approximately 584 participants for each dose level of mRNA-1273.617.2. Assuming 10% of participants will be excluded from the PP Set for Immunogenicity, with approximately 526 participants in each dose level of mRNA-1273.617.2 and mRNA-1273 primary series historical control arm respectively in the PP Set for Immunogenicity, there is approximately 90% power to reject both null hypotheses for the primary objectives based on GMR and difference in SRR. The assumptions are: the true GMR (mRNA-1273.617.2 booster vs. 100 µg mRNA-1273 primary series historical control) against the variant (B.1.617.2) is 0.9 compared to 100 ug mRNA-1273 against ancestral SARS-CoV-2, the standard deviation of the log-transformed titer is 1.5, non-inferiority margin for GMR is 1.5; the true SRR against variant after a single booster dose of mRNA-1273.617.2 is 90%, and SRR against ancestral SARS-CoV-2 and variant after mRNA-1273 primary series is 90%, and non-inferiority margin for SRR difference is 10%. The immune response assumptions are the same for 50 µg and 100 µg mRNA-1273.617.2.

With approximately 584 participants exposed to each dose of mRNA-1273.617.2, there is at least 90% probability in each group to observe one participant reporting an AE if the true rate of AEs is 1%.

**Part D**

Each dose level of mRNA-1273.213 will be assessed at a 2-sided type I error rate of 5%.

The target enrollment is approximately 584 participants for each dose level of mRNA-1273.213. Assuming 10% of participants will be excluded from the PP Set for Immunogenicity, with approximately 526 participants in each dose level of mRNA-1273.213 and mRNA-1273 primary series historical control arm respectively in the PP Set for Immunogenicity, there is approximately 80% power to reject all null hypotheses for the primary objectives based on GMR and difference in SRR. The assumptions are: the true GMR (mRNA-1273.213 vs. 100 µg mRNA-1273 primary series historical control) against the 2 variants (B.1.617.2, B.1.351) is 0.9 compared to 100 ug mRNA-1273 against ancestral SARS-CoV-2, the standard deviation of the log-transformed titer is 1.5, non-inferiority margin for GMR is 1.5; the true SRR against variants (B.1.617.2, B.1.351) after a single booster dose of mRNA-1273.213 is 90%, and SRR against ancestral SARS-CoV-2 and variant after mRNA-1273 primary series is 90%, and non-inferiority margin for SRR difference is 10%. The immune response assumptions are the same for 50 µg and 100 µg mRNA-1273.617.2.

With approximately 584 participants exposed to each dose of mRNA-1273.213, there is at least 90% probability in each group to observe one participant reporting an AE if the true rate of AEs is 1%.

**Part F**

mRNA-1273.529 in each cohort will be assessed at a 2-sided type I error rate of 5%.

Cohort 1:

The target enrollment is approximately 375 participants for 50 µg mRNA-1273.529. Assuming 20% of participants will be excluded from the PP Set for Immunogenicity-SARS-CoV2 negative, with approximately 300 participants in 50 µg mRNA-1273.529 and 300 participants in 50 µg mRNA-1273 (external comparator) in the PP Set for Immunogenicity-SARS-CoV-2 negative, there is approximately 89% global power for the primary immunogenicity objectives with alpha level of 0.05 (2-sided). The assumptions are: the true GMR (mRNA-1273.529 booster vs. 50 µg mRNA-1273 booster) against the variant (B.1.1.529) is 1.5, the standard deviation of the log-transformed titer is 1.5, and the non-inferiority margin for GMR is 1.5; the true SRR against B.1.1.529 after a single booster dose of 50 µg mRNA-1273.529 is 90% (same assumption for 50 µg mRNA-1273), and non-inferiority margin for SRR difference is 10%. With approximately 375 participants exposed to 50 µg mRNA-1273.529, there is at least 90% probability in this group to observe 1 participant reporting an AE if the true rate of AEs is 1%.

There may be an urgency to perform the Day 29 analysis as early as possible and depending on the testing capability of assays of antibodies against B.1.1.529, the Sponsor may decide using an external arm with less than 300 participants. Such decision will be documented in SAP prior to the planned Day 29 analysis.

Cohort 2:

The target enrollment is approximately 750 participants for 50 µg mRNA-1273.529 and 50 µg mRNA-1273 (1:1 ratio). Assuming 20% of participants will be excluded from the PP Set for Immunogenicity – SARS-CoV-2 negative, with approximately 300 participants each in 50 µg mRNA-1273.529 and 50 µg mRNA-1273 in the PP Set for Immunogenicity and SARS-CoV-2 negative, there is approximately 89% global power to demonstrate the primary immunogenicity objectives of alpha level of 0.05 (2-sided). The assumptions are: the true GMR (mRNA-1273.529 as the second booster vs 50 µg mRNA-1273 as the second booster) against the variant (B.1.1.529) is 1.5, the standard deviation of the log-transformed titer is 1.5, and the non-inferiority margin for GMR is 1.5; the true SRR against B.1.1.529 after mRNA-1273 as a second booster dose is 90% (same assumption for both 50 µg mRNA-1273.529 and 50 µg mRNA-1273), and non-inferiority margin for SRR difference is 10%.

With approximately 375 participants exposed to each group, there is at least 90% probability in each group to observe 1 participant reporting an AE if the true rate of AEs is 1%.

**Part G**

The target enrollment is approximately 375 participants for 50 µg mRNA-1273.214. Hypotheses testing will be performed at Day 29 and Day 91, alpha of 0.025 (2-sided) will be allocated equally to each one of the 2 time points. Assuming 20% of participants will be excluded from the PP Set for Immunogenicity – SARS-CoV-2 negative, with approximately 300 participants in 50 µg mRNA-1273.214 and 300 participants in 50 µg mRNA-1273 (Part F, Cohort 2-50 µg mRNA-1273) in the PP Set for Immunogenicity and SARS-CoV-2 negative, there is approximately 71% global power to demonstrate the primary immunogenicity objectives with alpha of 0.025 (2-sided) at each time point. The assumptions are: the true GMR (mRNA-1273.214 second booster vs 50 µg mRNA-1273 second booster) against the variant (B.1.1.529) is 1.5, the true GMR against ancestral SARS-CoV-2 is 1, the standard deviation of the log-transformed titer is 1.5, and the non-inferiority margin for GMR is 1.5, the true SRR against B.1.1.529 after mRNA-1273.214 as a second booster dose is 90% (same assumption for both 50 µg mRNA-1273.214 and 50 µg mRNA-1273), and non-inferiority margin for SRR difference is 10%.

With approximately 375 participants exposed to 50 µg mRNA-1273.214, there is at least 90% probability in this group to observe 1 participant reporting an AE if the true rate of AEs is 1%.

**Part H**

The target enrollment is approximately 500 participants for 50 µg mRNA-1273.222. Assuming 40% of participants will be excluded from the PP Set for Immunogenicity – SARS-CoV-2 negative (due to a SARS-CoV-2 infection prebooster), with approximately 300 participants in 50 µg mRNA-1273.222 and 260 participants in 50 µg mRNA-1273 (Part F, Cohort 2-50 µg mRNA-1273) in the PP Set for Immunogenicity and SARS-CoV-2 negative, there is approximately 60% power to demonstrate the primary immunogenicity objectives with an alpha of 0.05 (2-sided) at Day 29. The assumptions are: the true GMR (mRNA-1273.222 second booster vs. mRNA-1273 second booster) against Omicron BA.4/5 is 1.5, GMR (mRNA-1273.222 second booster vs. mRNA-1273 second booster) against ancestral SARS-CoV-2 D614G is 1, the standard deviation of the log-transformed titer is 1.5, and the non-inferiority margin for GMR is 1.5. The true SRR against Omicron BA.4/5 after mRNA-1273.222 as a second booster dose is 95% (same assumption for 50 µg mRNA-1273), and non-inferiority margin for SRR difference against Omicron BA.4/5 is 5%. The true SRR against ancestral SARS-CoV-2 D614G after mRNA-1273.222 as a second booster dose is 95% (same assumption for 50 µg mRNA-1273), and the non-inferiority margin for SRR difference against ancestral SARS-CoV-2 D614G is 10%.

With approximately 500 participants exposed to 50 µg mRNA-1273.222, there is at least 90% probability in this group to observe 1 participant reporting an AE if the true rate of AEs is 1%.

**Part J**

The sample size for Part J is not driven by statistical assumptions for formal hypothesis testing. The number of proposed participants is considered sufficient to provide a descriptive summary of the safety and immunogenicity of each treatment arm.

The target enrollment is approximately 100 participants who will be randomized in a 1:1 ratio to receive either mRNA-1273.815 (50 µg) or mRNA-1273.231 (50 µg).

**Analysis Sets:**

Analysis sets are described below (same definitions across Part A [1, 2], B, C, D, F, G, H, and J when applicable)**:**

| Set | Description |
|---|---|
| Full Analysis Set (FAS) | The FAS consists of all participants who receive IP. |
| Modified Intent-to-Treat (mITT) Set | The mITT Set consists of all participants in the FAS who have no serologic or virologic evidence of prior SARS-CoV-2 infection (both negative RT-PCR test for SARS-CoV-2 and negative serology test based on bAb specific to SARS-CoV-2 nucleocapsid) prebooster, ie, all FAS participants with baseline SARS-CoV-2 negative status prebooster. |
| Per-Protocol (PP) Set for Immunogenicity | The PP Set for Immunogenicity consists of all participants in the FAS who received the planned dose of study vaccination and no major protocol deviations that impact key or critical data.<br>The PP Set will be used as the primary analysis set for analyses of immunogenicity for immunobridging. |

| | |
|---|---|
| PP Set for Immunogenicity – SARS-CoV-2 negative (PPSI-Neg) | Participants in the PPSI who have no serologic or virologic evidence of SARS-CoV-2 infection at baseline, ie, who are SARS-CoV-2 negative, defined by both negative RT-PCR test for SARS-CoV-2 and negative serology test based on bAb specific to SARS-CoV-2 nucleocapsid<br>PPSI-Neg will be the primary analysis set for analyses of immunogenicity for between booster comparisons. |
| Solicited Safety Set | The Solicited Safety Set consists of all participants who receive IP and contribute any solicited AR data.<br>The Solicited Safety Set will be used for the analyses of solicited ARs. Participants will be included in the study arm corresponding to the dose of IP that they actually received. |
| Safety Set | The Safety Set consists of all participants who receive IP. The Safety Set will be used for all analyses of safety except for the solicited ARs. Participants will be included in the study arm corresponding to the dose of IP that they actually received. |
| Per-Protocol Set for Efficacy | The PP Set for Efficacy consists of all participants in the FAS who receive the planned dose of study vaccination, who are SARS-CoV-2 negative at baseline (ie, have a negative RT-PCR test for SARS-CoV-2 and a negative serology test based on bAb specific to SARS-CoV-2 nucleocapsid at baseline), and have no major protocol deviations that impact key or critical data. |

**Safety Analyses:**

All safety analyses will be based on the Safety Set, except summaries of solicited ARs, which will be based on the Solicited Safety Set. All safety analyses will be provided by study arm.

Safety and reactogenicity will be assessed by clinical review of all relevant parameters including solicited ARs (local and systemic ARs), unsolicited AEs, treatment-related AEs, severe AEs, SAEs, MAAEs, AESI, AEs leading to withdrawal, vital sign measurements, and physical examination findings.

The number and percentage of participants with any solicited local AR, with any solicited systemic AR, with any solicited AR during the 7-day follow-up period after vaccination, and with Grade 3 or higher solicited AR will be provided. A 2-sided 95% exact confidence interval (CI) using the Clopper-Pearson method will be provided for the percentage of participants with any solicited AR.

The number and percentage of participants with unsolicited AEs, treatment-related AEs, severe AEs, SAEs, MAAEs, AESI, and AEs leading to withdrawal will be summarized. Unsolicited AEs will be presented by Medical Dictionary for Regulatory Activities (MedDRA) system organ class and preferred term. Unsolicited AEs will be coded according to the MedDRA Dictionary for Adverse Reaction Terminology.

The number of events of solicited ARs, unsolicited AEs/SAEs, MAAEs, AEs leading to withdrawal, and AESI will be reported in summary tables accordingly. Pregnancy outcomes will also be summarized.

**Immunogenicity Analyses:**

**Part A.1:**

Each dose level of mRNA-1273.211 (50 and 100 µg) will be assessed with respect to mRNA-1273 primary series historical control separately. For the primary objective on immune response, there are 4 null hypotheses to be tested for each arm.

A. $H_0^1$: 50 or 100 µg mRNA-1273.211, as a single booster dose, is inferior to the primary series (100 µg) of mRNA-1273 based on GMT ratio against ancestral SARS-CoV-2 with a non-inferiority margin of 1.5.
B. $H_0^2$: 50 or 100 µg mRNA-1273.211, as a single booster dose, is inferior to the primary series (100 µg) mRNA-1273 based on difference in SRR against ancestral SARS-CoV-2 with a non-inferiority margin of 10%.
C. $H_0^3$: 50 or 100 µg mRNA-1273.211 against the variant B.1.351 is inferior to the primary series of (100 µg) mRNA-1273 against ancestral SARS-CoV-2 based on the GMT ratio of mRNA-1273.211 against the variant B.1.351 at Day 29 compared to mRNA-1273 against ancestral SARS-CoV-2 at Day 57 with a non-inferiority margin of 1.5.
D. $H_0^4$: 50 or 100 µg mRNA-1273.211 against the variant B.1.351 is inferior to the primary series of (100 µg) mRNA-1273 against ancestral SARS-CoV-2 based on the difference in SRR (SRR of mRNA-1273.211 against the variant B.1.351 at Day 29 – SRR of mRNA-1273 against ancestral SARS-CoV-2 at Day 57) with a non-inferiority margin of 10%.

Pseudotyped virus neutralizing antibody will be used as the basis to assess non-inferiority in immune response. The assays that will be used to assess the immune response to vaccination will be described in the SAP.

Each dose level of mRNA-1273.211 will be assessed with respect to mRNA-1273 primary series (historical control) separately. An analysis of covariance (ANCOVA) model will be carried out with antibody titers (Day 29 on 50 or 100 µg mRNA-1273.211 and Day 57 on mRNA-1273 primary series historical control) against ancestral SARS-CoV-2 as a dependent variable and a group variable (50 or 100 µg mRNA-1273.211 and mRNA-1273 primary series historical control) as the fixed effect, adjusting for age groups (<65, ≥ 65). The GMT will be estimated by the geometric least square mean (GLSM) from the model for each group and corresponding 95% CI will be provided for each group. The GMR (ratio of GMTs) for each dose level of mRNA-1273.211 with respect to mRNA-1273 will be estimated by the ratio of GLSM from the model, the estimated between group differences along with the corresponding 95% CI. The 95% CI for the ratio of GLSM will be provided to assess the between group difference in immune response against ancestral SARS-CoV-2 for each dose level of mRNA-1273.211 at Day 29 compared to mRNA-1273 primary series historical control at Day 57.

For each dose level of mRNA-1273.211, the non-inferiority of immune response to mRNA-1273 will be considered demonstrated if the lower bound of the corresponding CI of the GMR against ancestral SARS-CoV-2 is > 0.667 based on the non-inferiority margin of 1.5.

The primary definition of seroresponse is defined as ≥ 4 × LLOQ for those with pre-dose 1 of primary series baseline < LLOQ; ≥ 4-foldrise for those with pre-dose 1 of primary series baseline ≥ LLOQ. This definition will be used as the basis for non-inferiority hypothesis testing. The number and percentage (rate) of participants achieving seroresponse at Day 29 will be summarized with 95% CI calculated using the Clopper-Pearson method for each group. The difference of SRR between each dose level of mRNA-1273.211 at Day 29 and mRNA-1273 primary series historical control at Day 57 in mRNA-1273-P301 against ancestral SARS-CoV-2 will be calculated with 95% CI. The non-inferiority in SRR of each dose level of mRNA-1273.211 compared to mRNA-1273 primary series historical control will be considered demonstrated if the lower bound of the 95% of the SRR difference is > -10% based on the non-inferiority margin of 10%.

The same analysis methods for immune response against variant B.1.351 will be used for $H_0^3$ and $H_0^4$ hypotheses testing.

For each dose level of mRNA-1273.211, the primary immunogenicity objective (against ancestral SARS-CoV-2 and variant B.1.351) is considered met if non-inferiority is demonstrated based on both GMR and SRR difference, specifically:

- If the lower bound of the 95% CI of the GMT ratio against ancestral SARS-CoV-2 between mRNA-1273.211 and mRNA-1273 is > 0.667 based on the non-inferiority margin of 1.5, and
- The lower bound of the 95% CI of the SRR difference (mRNA-1273.211 – mRNA-1273) against ancestral SARS-CoV-2 is ≥ -10%, and
- The lower bound of the 95% CI of the GMT ratio between mRNA-1273.211 against the variant (B.1.351) as compared to 100 µg mRNA-1273 at Day 29 against ancestral SARS-CoV-2 at Day 57 is > 0.667 based on the non-inferiority margin of 1.5, and
- The lower bound of the 95% CI of the SRR difference (mRNA1273.211 against the variant [B.1.351] at Day 29 – 100 µg mRNA1273 against ancestral SARS-CoV-2 at Day 57) is > -10%

**Part A.2**

There is no hypothesis testing for Part A.2.

For Part A.2 participants, Day 29 and Day 181 immune response after mRNA-1273.214 (50 µg) as a second booster dose will be compared with their own Day 29 and Day 181 immune response of mRNA-1273.211 (50 µg) received as the first booster dose. GMT ratios will be calculated by back transforming the mean of paired differences of antibody titer data on the logarithmic scale between Day 29 and Day 181 post mRNA-1273.214 and Day 29 and Day 181 of antibody titer data post mRNA-1273.211. CIs for the GMT ratio will be based on t-distribution of the log-transformed values then back-transformed to the original scale for presentation. Seroresponse rates at Day 29 and Day 181 post mRNA-1273.214 will be compared with their seroresponse rates at Day 29 and Day 181 post mRNA-1273.211. The difference in seroresponse rates and its corresponding 95% CI based on adjusted Wald method will be provided.

**Part B:**

For the primary objective on immune response for a single booster dose of 100 µg mRNA-1273, there are 2 null hypotheses to be tested.

A. $H_0^1$: 100 µg mRNA-1273, as a single booster dose, is inferior to the primary series of (100 µg) mRNA-1273 based on GMT ratio against ancestral SARS-CoV-2 with a non-inferiority margin of 1.5.

B. $H_0^2$: 100 µg mRNA-1273, as a single booster dose, is inferior to the primary series of (100 µg) mRNA-1273 based on difference in SRR against ancestral SARS-CoV-2 with a non-inferiority margin of 10%.

The same analysis methods described in Part A.1 will be used for the primary immunogenicity objective for Part B.

The primary immunogenicity objective (against ancestral SARS-CoV-2) is considered met if non-inferiority is demonstrated based on both GMR and SRR difference, specifically:

- The lower bound of the 95% CI of the GMT ratio against ancestral SARS-CoV-2 between 100 µg mRNA-1273 booster dose and 100 µg mRNA-1273 primary series is > 0.667 based on the non-inferiority margin of 1.5, and
- The lower bound of the 95% CI of the SRR difference (100 µg mRNA-1273 booster dose – 100 µg mRNA-1273 primary series) against ancestral SARS-CoV-2 > -10%

**Part C:**

Each dose level of mRNA-1273.617.2 (50 and 100 µg) will be assessed with respect to mRNA-1273 primary series historical control separately. For the primary objective on immune response, there are 2 null hypotheses to be tested for each arm.

A. $H_0^1$: 50 µg or 100 µg mRNA-1273.617.2, as a single booster dose, against the variant B.1.617.2 is inferior to the primary series of (100 µg) mRNA-1273 against ancestral SARS-CoV-2 based on the GMT ratio of mRNA-1273.617 against the variant B.1.617.2 at Day 29 compared to mRNA-1273 against ancestral SARS-CoV-2 at Day 57 with a non-inferiority margin of 1.5

B. $H_0^2$: 50 µg or 100 µg mRNA-1273.617.2, as a single booster dose, against the variant B.1.617.2 is inferior to the primary series of (100 µg) mRNA-1273 against ancestral SARS-CoV-2 based on the difference in SRR (SRR of mRNA-1273.617.2 against the variant B.1.617.2 at Day 29 – SRR of mRNA-1273 against ancestral SARS-CoV-2 at Day 57) with a non-inferiority margin of 10%

The same analysis methods described in Part A.1 will be used for the primary immunogenicity objective for Part C.

The primary immunogenicity objective (against the variant) is considered met if non-inferiority is demonstrated based on both GMR and SRR difference, specifically:

- The lower bound of the 95% CI of the GMT ratio between mRNA1273.617.2 against the variant (B.1.617.2) at Day 29 as compared to 100 µg mRNA1273 against ancestral SARS-CoV-2 at Day 57 is > 0.667 based on the non-inferiority margin of 1.5, and
- The lower bound of the 95% CI of the SRR difference (mRNA-1273.617.2 against the variant [B.1.617.2] at Day 29 – 100 µg mRNA-1273 against ancestral SARS-CoV-2 at Day 57 is > -10%)

**Part D**

Each dose level of mRNA-1273.213 (50 and 100 µg) will be assessed with respect to mRNA-1273 primary series historical control separately. For the primary objective on immune response, there are 4 null hypotheses to be tested for each arm.

A. $H_0^1$: 50 µg or 100 µg mRNA-1273.213, as a single booster dose, against the variant B.1.617.2 is inferior to the primary series of (100 µg) mRNA-1273 against ancestral SARS-CoV-2 based on the GMT ratio of mRNA-1273.617.2 against the variant

B.1.617.2 at Day 29 compared to mRNA-1273 against ancestral SARS-CoV-2 at Day 57 with a non-inferiority margin of 1.5

B. $H_0^2$: 50 µg or 100 µg mRNA-1273.213, as a single booster dose, against the variant B.1.617.2 is inferior to the primary series of (100 µg) mRNA-1273 against ancestral SARS-CoV-2 based on the difference in SRR (SRR of mRNA-1273.617.2 against the variant B.1.617.2 at Day 29 – SRR of mRNA-1273 against ancestral SARS-CoV-2 at Day 57) with a non-inferiority margin of 10%

C. $H_0^3$: 50 µg or 100 µg mRNA-1273.213, as a single booster dose, against the variant B.1.351 is inferior to the primary series of (100 µg) mRNA-1273 against ancestral SARS-CoV-2 based on the GMT ratio of mRNA-1273.213 against the variant B.1.351 at Day 29 compared to mRNA-1273 against ancestral SARS-CoV-2 at Day 57 with a non-inferiority margin of 1.5

D. $H_0^4$: 50 µg or 100 µg mRNA-1273.213, as a single booster dose, against the variant B.1.351 is inferior to the primary series of (100 µg) mRNA-1273 against ancestral SARS-CoV-2 based on the difference in SRR (SRR of mRNA-1273.213 against the variant B.1.351 at Day 29 – SRR of mRNA-1273 against ancestral SARS-CoV-2 at Day 57) with a non-inferiority margin of 10%

The same analysis methods described in Part A.1 will be used for the primary immunogenicity objective for Part D.

The primary immunogenicity objective (against the variant) is considered met if non-inferiority is demonstrated based on both GMR and SRR difference, specifically:

- The lower bound of the 95% CI of the GMT ratio between mRNA-1273.213 against the variant (B.1.617.2) at Day 29 as compared to 100 µg mRNA-1273 against ancestral SARS-CoV-2 at Day 57 is > 0.667 based on the non-inferiority margin of 1.5, and
- The lower bound of the 95% CI of the SRR difference (mRNA-1273.213 against the variant [B.1.351] at Day 29 – 100 µg mRNA-1273 against ancestral SARS-CoV-2 at Day 57 is > -10%), and
- The lower bound of the 95% CI of the GMT ratio between mRNA-1273.213 against the variant (B.1.351) at Day 29 as compared to 100 µg mRNA-1273 against ancestral SARS-CoV-2 at Day 57 is > 0.667 based on the non-inferiority margin of 1.5, and
- The lower bound of the 95% CI of the SRR difference (mRNA-1273.213 against the variant [B.1.351] at Day 29 – 100 µg mRNA-1273 against ancestral SARS-CoV-2 at Day 57 is > -10%)

**Part F:**

Cohort 1:

50 µg mRNA-1273.529 booster dose (first booster dose) will be assessed with respect to mRNA-1273 booster dose (first booster dose) using an external comparator (details regarding the external historical control will be included in the SAP). For the primary immunogenicity objectives, there are 3 hypotheses to be tested.

A. $H_1^1$: 50 µg mRNA-1273.529, as a single booster dose, against the variant B.1.1.529 is non-inferior to the booster dose of (50 µg) mRNA-1273 against B.1.1.529 at Day 29 compared to mRNA-1273 against the variant B.1.1.529 at Day 29 with a non-inferiority margin of 1.5.

B. $H_1^2$: 50 µg mRNA-1273.529, as a single booster dose, against the variant B.1.1.529 is non-inferior to the booster dose of (50 µg) mRNA-1273 against B.1.1.529 based on the difference in SRR at Day 29 with a non-inferiority margin of 10%.

C. $H_1^3$: 50 µg mRNA-1273.529, as a single booster dose, against the variant B.1.1.529 is superior to the booster dose of (50 µg) mRNA-1273 against B.1.1.529 based on the GMT ratio of mRNA-1273.529 against the variant B.1.1.529 at Day 29 compared to mRNA-1273 against the variant B.1.1.529 at Day 29.

An ANCOVA model will be performed to assess the difference in immune response between mRNA-1273.529 and mRNA-1273 (using external comparator) as the first booster dose. For immune response against the B.1.1.529 strain, in the ANCOVA model, antibody titers at Day 29 postbooster against the B.1.1.529 strain will be a dependent variable, and a group variable (mRNA-1273.529 and mRNA-1273) will be the fixed effect, adjusting for age groups (< 65, ≥ 65 years) and prebooster antibody titer level, if applicable.

The GMT will be estimated by the GLSM from the model and its corresponding 95% will be provided for each group. The GMR (ratio of GMTs) for mRNA-1273.529 with respect to mRNA-1273 will be estimated by the ratio of GLSM from the model and the corresponding 95% CIs will be provided. The 95% CI for GMR will be used to assess the between group difference in immune response against the B.1.1.529 strain for mRNA-1273.529 at Day 29 compared to the mRNA-1273.

The number and percentage (rate) of participants achieving seroresponse at Day 29 will be summarized with 95% CI calculated using the Clopper-Pearson method for each group. The difference of SRR between mRNA-1273.529 and mRNA-1273 will be calculated with 95% CI based on Miettinen-Nurminen method. The non-inferiority in SRR of mRNA-1273.529 compared to mRNA-1273 will be considered demonstrated if the lower bound of the 95% of the SRR difference is > -10% based on the non-inferiority margin of 10%.

The primary immunogenicity objective (against the B.1.1.529) is considered met if non-inferiority is demonstrated based on GMR and SRR difference, specifically:

- The lower bound of the 95% CI of the GMT ratio between mRNA1273.529 against the variant (B.1.1.529) at Day 29 as compared to 50 µg mRNA1273 against B.1.1.529 at Day 29 is > 0.667 based on the non-inferiority margin of 1.5
- The lower bound of the 95% CI of the SRR difference (50 µg mRNA-1273.529 against the variant B.1.1.529 at Day 29–- 50 µg mRNA-1273 against B.1.1.529 at Day 29) is >-10%
- If non-inferiority is demonstrated (based on GMT ratio and SRR difference), the lower bound of 95% CI of the GMT ratio will be compared to 1, if it's greater than 1, then superiority is demonstrated.

Cohort 2:

The same analyses methods described in Part F Cohort 1 will be used for Cohort 2.

A. $H_1^1$: 50 µg mRNA-1273.529, as a second booster dose, against the variant B.1.1.529 is non-inferior to the second booster dose of (50 µg) mRNA-1273 against B.1.1.529 based on the GMT ratio of mRNA-1273.529 against the variant B.1.1.529 at Day 29 compared to mRNA-1273 against the variant B.1.1.529 at Day 29 with a non-inferiority margin of 1.5.

B. $H_1^2$: 50 µg mRNA-1273.529, as a second booster dose, against the variant B.1.1.529 is non-inferior to the second booster dose of (50 µg) mRNA-1273 against B.1.1.529 based on the difference in SRR at Day 29 with a non-inferiority margin of 10%.
C. $H_1^3$: 50 µg mRNA-1273.529, as a second booster dose, against the variant B.1.1.529 is superior to the second booster dose of (50 µg) mRNA-1273 against B.1.1.529 based on the GMT ratio of mRNA-1273.529 against the variant B.1.1.529 at Day 29 compared to mRNA-1273 against the variant B.1.1.529 at Day 29.

The primary immunogenicity objective (against the B.1.1.529) is considered met if non-inferiority is demonstrated based on GMR and SRR difference, specifically:

- The lower bound of the 95% CI of the GMT ratio between mRNA1273.529 against the variant (B.1.1.529) at Day 29 as compared to 50 µg mRNA1273 against B.1.1.529 at Day 29 is > 0.667 based on the non-inferiority margin of 1.5
- The lower bound of the 95% CI of the SRR difference (50 µg mRNA-1273.529 against the variant B.1.1.529 at Day 29–- 50 µg mRNA-1273 against B.1.1.529 at Day 29) is >-10%
- If non-inferiority is demonstrated (based on GMT ratio and SRR difference), the lower bound of 95% CI will be compared to 1, if it's greater than 1, then superiority is also demonstrated.

**Part G:**

50 µg mRNA-1273.214 as the second booster dose will be compared to 50 µg mRNA-1273 as the second booster dose (active control arm in Part F, Cohort 2)

For the primary immunogenicity objective, there are 4 hypotheses to be tested at Day 29 and 4 hypotheses to be tested at Day 91.

A. $H_1^1$: 50 µg mRNA-1273.214, as a second booster dose, against the variant B.1.1.529 is non-inferior to the second booster dose of (50 µg) mRNA-1273 against B.1.1.529 based on the GMT ratio of mRNA-1273.529 against the variant B.1.1.529 at Day 29 compared to mRNA-1273 against the variant B.1.1.529 at Day 29 with a non-inferiority margin of 1.5.
B. $H_1^2$: 50 µg mRNA-1273.214, as a second booster dose, against the variant B.1.1.529 is non-inferior to the second booster dose of (50 µg) mRNA-1273 against B.1.1.529 based on the difference in SRR at Day 29 with a non-inferiority margin of 10%.
C. $H_1^3$: 50 µg mRNA-1273.214, as a second booster dose, against ancestral SARS-CoV-2 is non-inferior to the second booster dose of (50 µg) mRNA-1273 against ancestral SARS-CoV-2based on the GMT ratio of mRNA-1273.214 against ancestral SARS-CoV-2 at Day 29 compared to mRNA-1273 against ancestral SARS-CoV-2 at Day 29 with a non-inferiority margin of 1.5.
D. $H_1^4$: 50 µg mRNA-1273.214, as a second booster dose, against the variant B.1.1.529 is superior to the second booster dose of (50 µg) mRNA-1273 against B.1.1.529 based on the GMT ratio of mRNA-1273.214 against the variant B.1.1.529 at Day 29 compared to mRNA-1273 against the variant B.1.1.529 at Day 29.
E. $H_1^5$: 50 µg mRNA-1273.214, as a second booster dose, against the variant B.1.1.529 is non-inferior to the second booster dose of (50 µg) mRNA-1273 against B.1.1.529 based on the GMT ratio of mRNA-1273.214against the variant B.1.1.529 at Day 91

compared to mRNA-1273 against the variant B.1.1.529 at Day 91 with a non-inferiority margin of 1.5.

F. $H_1^6$: 50 µg mRNA-1273.214, as a second booster dose, against the variant B.1.1.529 is non-inferior to the second booster dose of (50 µg) mRNA-1273 against B.1.1.529 based on the difference in SRR at Day 91 with a non-inferiority margin of 10%.
G. $H_1^7$: 50 µg mRNA-1273.214, as a second booster dose, against ancestral SARS-CoV-2 is non-inferior to the second booster dose of (50 µg) mRNA-1273 against ancestral SARS-CoV-2 based on the GMT ratio of mRNA-1273.214 against ancestral SARS-CoV-2 at Day 91 compared to mRNA-1273 against ancestral SARS-CoV-2 at Day 91 with a non-inferiority margin of 1.5.
H. $H_1^8$: 50 µg mRNA-1273.214, as a second booster dose, against the variant B.1.1.529 is superior to the second booster dose of (50 µg) mRNA-1273 against B.1.1.529 based on the GMT ratio of mRNA-1273.214 against the variant B.1.1.529 at Day 91 compared to mRNA-1273 against the variant B.1.1.529 at Day 91.

The analyses method described in Part F Cohort 1 will be used for Part G.

The primary immunogenicity objective is considered met if non-inferiority against B.1.1.529 based on GMR and SRR difference, and non-inferiority against ancestral SARS-CoV-2 based on GMR are demonstrated at Day 29 or at Day 91.

For the primary immunogenicity objective, the non-inferiority of mRNA-1273.214 as compared to mRNA-1273 against the B.1.1.529 and the non-inferiority of mRNA-1273.214 as compared to mRNA-1273 against ancestral SARS-CoV-2 at Day 29 will be assessed using a non-inferiority margin of 1.5 at 2-sided alpha of 0.025, and the non-inferiority of mRNA-1273.214 as compared to mRNA-1273 against B.1.1.529 based on SRR using a non-inferiority margin of 10% at 2-sided alpha of 0.025.

Superiority of mRNA-1273.214 as compared to mRNA-1273 against the B.1.1.529 strain will be evaluated at Day 29. Once the non-inferiority of mRNA-1273.214 as compared to mRNA-1273 against B.1.1.529 and against ancestral SARS-CoV-2 at Day 29 is demonstrated, the 97.5% CI of GMR (mRNA-1273.214 vs mRNA-1273) will be used to assess superiority of mRNA-1273.214 as compared to mRNA-1273. If the lower bound of the GMR > 1 at Day 29, superiority of mRNA-1273.214 compared to mRNA-1273 against B.1.1.529 will be considered demonstrated.

Hypotheses testing at Day 91 will be performed in the same manner; first test 3 non-inferiority hypotheses (2 against B.1.1.529 and 1 against ancestral SARS-CoV-2) at alpha of 0.025 level (two-sided). Once non-inferiority is demonstrated for both B.1.1529 and the ancestral SARS-CoV-2, then superiority testing against B.1.1.529 at alpha of 0.025 level (two-sided) will also be performed.

For the key secondary objective, there are 2 hypotheses to be tested (Day 29 and Day 91 each with alpha level of 0.025, 2-sided):

A. $H_1^9$: 50 µg mRNA-1273.214, as a second booster dose, against ancestral SARS-CoV-2 is non-inferior to the booster dose of (50 µg) mRNA-1273 against ancestral SARS-CoV-2 based on the difference in SRR at Day 29 with a non-inferiority margin of 10%.
B. $H_1^{10}$: 50 µg mRNA-1273.214, as a second booster dose, against ancestral SARS-CoV-2 is non-inferior to the booster dose of (50 µg) mRNA-1273 against ancestral SARS-CoV-2 based on the difference in SRR at Day 91 with a non-inferiority margin of 10%.

If the lower bound of the 97.5% CI of the SRR difference (50 µg mRNA-1273.214 against ancestral SARS-CoV-2–- 50 µg mRNA-1273 against ancestral SARS-CoV-2) is >-10% at Day 29 or Day 91, then the key secondary objective will be considered met.

In the event that an early assessment of the 1273.214 data is needed due to public health concerns, a two-staged approach might be used. Specifically, a subset of participants' (ie, 50 first enrolled participants) serum samples will first be tested against ancestral SARS-CoV-2 and variants of concern. For the Day 29 and Day 91 immunogenicity analyses, all participants' immune data will be used in the formal interim analysis to evaluate the primary immunogenicity objective.

**Part H:**

The primary immunogenicity objective will be assessed in the per-protocol immunogenicity, SARS-CoV-2 negative set.

The analyses methods described in Part F Cohort 1 will be used for Part H.

The primary immunogenicity objective is considered met if non-inferiority against Omicron BA.4/5 and ancestral SARS-CoV-2 D614G based on GMR and SRR difference at Day 29 is demonstrated.

Non-inferiority against Omicron BA.4/5:

- The lower bound of the 95% CI of the GMT ratio between mRNA-1273.222 against Omicron BA.4/5 at Day 29 as compared to 50 µg mRNA-1273 against BA.4/5 at Day 29 is > 0.667 based on the non-inferiority margin of 1.5.
- The lower bound of the 95% CI of the SRR difference (50 µg mRNA-1273.222 against Omicron BA. 4/5 at Day 29–- 50 µg mRNA-1273 against BA.4/5 at Day 29) is >-5%.

Non-inferiority against ancestral SARS-CoV-2 D614G:

- The lower bound of the 95% CI of the GMT ratio between mRNA-1273.222 against ancestral SARS-CoV-2 D614G at Day 29 as compared to 50 µg mRNA-1273 against ancestral SARS-CoV-2 D614G at Day 29 is > 0.667 based on the non-inferiority margin of 1.5.
- The lower bound of the 95% CI of the SRR difference (50 µg mRNA-1273.222 against ancestral SARS-CoV-2 D614G at Day 29–- 50 µg mRNA-1273 against ancestral SARS-CoV-2 D614Gat Day 29) is >-10%.

If non-inferiority is demonstrated based on GMT ratio and SRR difference against Omicron BA. 4/5 and ancestral SARS-CoV-2 D614G then the lower bound of 95% CI of the GMT ratio will be compared to 1, and if it is higher than 1 then superiority will also be demonstrated.

**Part J:**

No formal hypothesis testing will be performed for Part J and all safety and immunogenicity analyses will be descriptive. The primary immunogenicity objective will be assessed using the Per-Protocol Set for Immunogenicity. Comparisons on immunogenicity response between the mRNA-1273.815 and mRNA-1273.231 treatment arms may be performed.

An ANCOVA model will be used to compare antibody response between treatment arms. Day 15 and Day 29 antibody titers (against SARS-CoV-2 variants) will be included in the model as a dependent variable and treatment group variables (mRNA-1273.815, mRNA-1273.231) will be included as fixed effect. The model may adjust for other covariates as appropriate. Geometric mean ratio and the corresponding 95% CI will be estimated from the model.

Seroresponse rate difference at Day 15 and Day 29 will be compared and 95% CI will be provided using Miettinen-Nurminen method.

**Immunogenicity endpoints applicable to all study parts:**

SARS-CoV-2-specific bAb and nAb are assessed at multiple timepoints in each part of this study.

For each of the antibodies of interest, eg, levels of SARS-CoV-2–specific bAb and SARS-CoV-2–specific nAb, the GMT or level with corresponding 95% CI at each time point, and GMFR of post-baseline/baseline titers or levels with corresponding 95% CI at each post-baseline time point will be provided for each arm. The 95% CIs will be calculated based on the t-distribution of the log-transformed values then back-transformed to the original scale for presentation. The following descriptive statistics will also be provided at each time point: number of participants (n), median, minimum, and maximum.

The mixed effect model repeated measure (MMRM) will be used to analyze all postbooster measures for between booster comparison when applicable, the model will include treatment group, analysis visit, treatment by visit interaction, and adjusting for age groups and prebooster titer levels. An unstructured covariance structure will be used to model the within-participant errors. The GMT will be estimated from the model and its corresponding 95% CI will be provided for each group at each postboost timepoint. The GMR (ratio of GMTs) will be estimated from the model and the corresponding 95% CI will be provided at each postboost timepoint.

The SRR of each arm against ancestral SARS-CoV-2 and variants, defined as the percentage of participants achieving seroresponse against ancestral SARS-CoV-2 and variants respectively, will be provided for each arm with the 95% CI calculated using the Clopper-Pearson method.

The primary definition of seroresponse is defined as ≥ 4 × LLOQ for those with pre-dose 1 of primary series baseline < LLOQ; ≥ 4-foldrise for those with pre-dose 1 of primary series baseline ≥ LLOQ. The secondary definition of seroresponse is defined as ≥ 4 × LLOQ for those with prebooster baseline < LLOQ; ≥ 4-foldrise for those with prebooster ≥ LLOQ. SRR will be summarized using both definitions for all the study parts.

**Efficacy Analysis**

Descriptive summaries of symptomatic COVID-19 disease, asymptomatic SARS-CoV-2 infection, as well as COVID-19 regardless of symptoms will be provided for each arm. Vaccine efficacy may be estimated if the number of cases accrued is deemed to be sufficient.

**Planned Analyses**

**Interim Analyses**

The interim analysis will be conducted based on safety and immunogenicity data collected through Day 15 or Day 29. The interim analysis may be conducted either after all participants in Parts A.1, A.2, B, C, D, F, G, H, or J have completed their Day 15 or Day 29 visit assessments and/or subsequent timepoint visits (eg, Day 91 for Parts F, G, H, and J) or combined after the last participant of each study part (Parts A.1, A.2, B, C, D, F, G, H, or J), dose arm, or pre-specified subset of dose arm has completed their Day 15 or Day 29 visit assessments.

**Final Analysis**

The final analysis of all endpoints will be performed after all participants have completed all planned study procedures. Results of this analysis will be presented in a final clinical study

| |
|---|
| report (CSR), including individual listings. The final CSR will include full analyses of all safety and immunogenicity through Day 366 (Month 12) for Parts A.1, B, C, D, F, and G and through D181 (Month 6) for Parts A.2, H, and J. |

### LIST OF FIGURES

Figure 1: Statistical Hypotheses Testing Strategy for Part G.................................................154

Figure 2: Statistical Hypotheses Testing for Part H .............................................................156

### LIST OF ABBREVIATIONS AND DEFINITION OF TERMS

The following abbreviations and terms are used in this study protocol.

| Abbreviation or Specialist Term | Definition |
|---|---|
| AE | adverse event |
| AESI | adverse event of special interest |
| ALT | alanine aminotransferase |
| ANCOVA | analysis of covariance |
| AR | adverse reaction |
| ARDS | acute respiratory distress syndrome |
| AST | aspartate aminotransferase |
| bAb | binding antibody |
| CDC | US Centers for Disease Control and Prevention |
| CEAC | Cardiac Event Adjudication Committee |
| CFR | Code of Federal Regulations |
| CI | confidence interval |
| CoV | coronavirus |
| COVID-19 | coronavirus disease 2019 |
| CRO | contract research organization |
| CSR | clinical study report |
| DSPC | 1,2-distearoyl-sn-glycero-3-phosphocholine |
| ECG or EKG | electrocardiogram |
| eCRF | electronic case report form |
| EDC | electronic data capture |
| eDiary | electronic diary |
| EoS | end of study |
| EUA | Emergency Use Authorization |
| FAS | Full Analysis Set |
| FDA | United States Food and Drug Administration |
| FSH | follicle-stimulating hormone |
| GCP | Good Clinical Practice |

| Abbreviation or Specialist Term | Definition |
|---|---|
| GLSM | geometric least square mean |
| GMFR | geometric mean fold rise |
| GMR | ratio of geometric mean titers |
| GMT | geometric mean titer |
| HCP | healthcare practitioner |
| HIPAA | Health Insurance Portability and Accountability Act |
| HRT | hormone replacement therapy |
| IB | Investigator's Brochure |
| ICF | informed consent form |
| ICH | International Council for Harmonisation |
| IM | intramuscular |
| IP | investigational product |
| IRB | institutional review board |
| LLOQ | lower limit of quantitation |
| LNP | lipid nanoparticle |
| LTFU | lost to follow-up |
| MAAE | medically attended adverse event |
| MedDRA | Medical Dictionary for Regulatory Activities |
| MMRM | mixed effect model repeated measure |
| mRNA | messenger RNA |
| nAb | neutralizing antibody |
| NP | nasopharyngeal |
| PBMC | peripheral blood mononuclear cells |
| PEG-2000-DMG | 1-monomethoxypolyethyleneglycol-2,3-dimyristylglycerol with polyethylene glycol of average molecular weight 2000 |
| PP | per-protocol |
| RBD | receptor-binding domain |
| RT-PCR | reverse transcriptase polymerase chain reaction |
| S | spike |

| Abbreviation or Specialist Term | Definition |
|---|---|
| S-2P | prefusion stabilized spike protein |
| SAE | serious adverse event |
| SAP | statistical analysis plan |
| SARS | severe acute respiratory syndrome |
| SARS-CoV-2 | severe acute respiratory syndrome coronavirus 2 |
| SoE | schedule of events |
| SRR | seroresponse rate |
| USP | United States Pharmacopoeia |
| VOC | variants of concern |
| WHO | World Health Organization |

### GLOSSARY OF TERMS

| Term/Concept | Definition |
|---|---|
| Adequate female contraception | Consistent and correct use of a FDA-approved contraceptive method in accordance with the product label. |
| Adverse event (AE) | Any untoward medical occurrence associated with the use of a drug in humans, whether or not considered drug related. |
| Adverse event of special interest (AESI) | An AE (serious or nonserious) of scientific and medical concern specific to the Sponsor's product or program for which ongoing monitoring and immediate notification by the investigator to the Sponsor is required.<br>A list of the AESI pertinent to this study is provided in Section 10.4 (Appendix 4). |
| Adverse reaction (AR) | Any AE for which there is a reasonable possibility that the vaccine caused the AE. For the purposes of investigational new drug safety reporting, "reasonable possibility" means that there is evidence to suggest a causal relationship between the vaccine and the AE.<br>Solicited ARs are defined in Section 7.4.3. |
| Anaphylaxis | An acute hypersensitivity reaction with multi-organ system involvement that can present as, or rapidly progress to, a severe life-threatening reaction. It may occur following exposure to allergens from a variety of sources.<br>Characteristics of anaphylaxis are provided in Section 7.4.4. |
| Asymptomatic SARS-CoV-2 infection | Asymptomatic SARS-CoV-2 infection is defined as a positive RT-PCR test on a respiratory sample in the absence of symptoms or a positive serologic test for anti-nucleocapsid antibody after a negative test at time of enrollment. |
| COVID-19 symptoms | • Fever (temperature ≥ 38.0°C [100.4°F]) or chills<br>• Cough<br>• Shortness of breath and/or difficulty<br>• Fatigue<br>• Muscle or body aches<br>• Headache<br>• New loss of taste and/or smell<br>• Sore throat, congestion, or runny nose<br>• Nausea or vomiting<br>• Diarrhea |
| Phase 3 Study (mRNA-1273-P301) definition of COVID-19 | The participant must have experienced at least TWO of the following systemic symptoms: Fever (≥ 38ºC), chills, myalgia, headache, sore throat, new olfactory and taste disorder(s),<br>OR<br>The participant must have experienced at least ONE of the following respiratory signs/symptoms: cough, shortness of breath or difficulty |

| Term/Concept | Definition |
|---|---|
| | breathing, OR clinical or radiographical evidence of pneumonia; AND The participant must have at least one NP swab, nasal swab, or saliva sample (or respiratory sample, if hospitalized) positive for SARS-CoV-2 by RT-PCR. |
| End of Study | Completion of the last visit of the last participant in the study or last scheduled procedure, as shown in the SoE (Table 17, Table 18, Table 19, and Table 20) for the last participant in the study. |
| Lost to follow-up | A participant who repeatedly fails to return for scheduled visits without stating an intention to withdraw consent and is unable to be contacted by the study site. |
| Medically attended adverse event (MAAE) | An AE that leads to an unscheduled visit to a healthcare provider. |
| Nonchildbearing potential | Surgically sterile (history of bilateral tubal ligation, bilateral oophorectomy, hysterectomy) or postmenopausal (defined as amenorrhea for ≥ 12 consecutive months prior to screening without an alternative medical cause). |
| Screen failures | Participants who consent to participate in the clinical study but are not subsequently assigned to treatment. |
| Serious adverse event (SAE) | An AE is considered an SAE, if, in the view of either the investigator or Sponsor, it results in any of the following outcomes (see Section 7.4.2 for further details of each criterion):<br>• Death<br>• Is life-threatening<br>• Inpatient hospitalization or prolongation of existing hospitalization<br>• Persistent or significant incapacity or substantial disruption of the ability to conduct normal life functions<br>• Congenital anomaly or birth defect<br>• Medically important event. |
| Symptomatic COVID-19 | The presence of one of the CDC-listed symptoms (CDC 2022) and a positive RT-PCR test on a respiratory sample. |
| Unsolicited AE | Any AE reported by the participant that is not specified as a solicited AR in the protocol or is specified as a solicited AR but starts outside the protocol-defined period for reporting solicited ARs (ie, 7 days after vaccination). |
| Women of childbearing potential | Women of childbearing potential are those who are considered fertile following menarche and until becoming postmenopausal unless permanently sterile (see Section 10.3, Appendix 3). |

Abbreviations: AE = adverse event; AESI = adverse event of special interest; AR = adverse reaction; MAAE = medically attended adverse event; RT-PCR = reverse transcriptase polymerase chain reaction; SAE = serious adverse event.

### 1. INTRODUCTION

#### 1.1. Study Rationale

Coronaviruses (CoVs) are a large family of viruses that cause illness ranging from the common cold to more severe diseases, such as Middle East Respiratory Syndrome and severe acute respiratory syndrome (SARS). An outbreak of a novel coronavirus (COVID-19, later designated SARS-CoV-2) initially emerged in Wuhan, Hubei Province, China in December 2019. The World Health Organization (WHO) declared COVID-19 a pandemic on 11 Mar 2020 with more than 157 million cases and 3.2 million deaths by 09 May 2021 (WHO 2021).

ModernaTX, Inc. (the Sponsor)'s scalable messenger RNA (mRNA)/lipid nanoparticle (LNP) technology platform allowed for a rapid response to the pandemic and was used to develop mRNA-1273, a novel LNP-encapsulated mRNA-based vaccine against SARS-CoV-2. mRNA-1273 contains a single mRNA (CX-024414) that encodes for the full-length SARS-CoV-2 spike (S) protein of the Wuhan-Hu-1 SARS-CoV-2 virus, modified with 2 proline substitutions within the heptad repeat 1 domain (S-2P) to stabilize the spike protein into a prefusion conformation. Having achieved the primary endpoint in a pivotal Phase 3 study conducted in persons at high risk for SARS-CoV-2 infection, in December 2020, mRNA-1273 was granted EUA for the prevention of COVID-19 for individuals 18 years of age and older based on the demonstration of efficacy and safety in a Phase 3 pivotal trial and was subsequently licensed in the US (31 January 2022). (Baden et al 2021).

Over the course of the pandemic, SARS-CoV-2 variants have emerged and are likely to continue to emerge, some of which may prove to have some level of escape from immunity associated with previous infection or vaccination. Recently, newer variants have raised concern, due to reports of increased infectivity or reduction in the ability of convalescent sera or sera from vaccinated participants to neutralize these emergent strain variants. Mutations occurring in the receptor-binding domain (RBD) are of particular concern, as this site includes the dominant neutralization epitopes on the S protein and these mutations could impact the effectiveness of antibodies elicited by infection or vaccination in neutralizing the virus (Greaney et al 2021).

These recent evolutionary events indicate that SARS-CoV-2 has the capacity to develop more efficient transmission between human hosts (Martin et al 2021) and vaccination strategies to control the virus need to be responsive to this evolution. A SARS-CoV-2 variant, B.1.1.7, rapidly spread from southeast England around the globe. Relative to the Wuhan viral isolate, B.1.1.7 includes 8 mutations located in the S protein, including the N501Y mutation occurring in the RBD. Early analyses indicate that B.1.1.7 has a substantial fitness advantage over other currently circulating lineages. The B.1.351 variant emerged in South Africa, and the P.1 lineage has recently been reported in Brazil. There are at least 11 mutations located in the S protein, 3 of which (K417N, E484K, and N501Y) are found in the RBD. More recently SARS-CoV-2 variants emerged in India, and the B.1.617.2 variant (Delta variant) containing 2 mutations in the RBD (L452R and T478K) is currently circulating globally. In vitro characterization of sera from individuals recently vaccinated with the 2-dose regimen of the Moderna COVID-19 Vaccine at the 100 µg dose showed that the Moderna COVID-19 Vaccine produced neutralizing titers against key emerging variants tested, including B.1.1.7, B.1.351, and B.1.617.2 (Wang et al 2021, Wu et al 2021a, Choi A et al 2021). The studies showed no significant reduction in neutralizing titers against the B.1.1.7 relative to the ancestral Wuhan-Hu-1 strain and a 2.1-fold reduction versus B.1.617.2; however, a greater than six-fold reduction in neutralizing titers was observed against the B.1.351 variant relative to the Wuhan-Hu-1. Evidence from adenovirus vector SARS-CoV-2 vaccines based on the Wuhan-Hu-1 sequence suggests reduced vaccine efficacy against moderate to severe COVID-19 in South Africa where the B.1.351 variant is circulating (Madhi et al 2021, Sadoff et al 2021).

In November 2021, the SARS-CoV-2 Omicron variant (B.1.1.529; BA.1) was detected in South Africa and currently epidemiological information about its spread in other regions is being evaluated. The Omicron variant has significant antigenic change with a potential growth advantage. In addition, it contains potential antibody escape site mutations (such as K417N, T478K, E484A, and N501Y). After the emergency of the BA.1 sublineage, other Omicron sublineages (BA.2, BA.4, BA.5, and XBB.1.5) with additional potential antibody escape site mutations have been detected in multiple geographies.

There is an urgent need for vaccination strategies that induce broader protection against variants of concern (VOC), to decrease morbidity and mortality. In addition, it is currently not known whether breakthrough infections could occur long-term due to waning antibody titers (Doria-Rose et al 2021, Pegu A et al). Based on the experience of mRNA-1273 and leveraging the flexible nature of the mRNA technology, Moderna is evaluating multiple mRNA vaccines to address emerging variants.

Overall, study Parts A.1, B, C, and D will assess whether a single booster dose of the mRNA vaccines in each study part elicits antibody responses to ancestral SARS-CoV-2 and the variants similarly to antibody responses to ancestral SARS-CoV-2 elicited by 2 doses of mRNA-1273 (100 µg) against ancestral SARS-CoV-2, using a historical control arm from the mRNA-1273-P301 (COVE) study. Study Part F Cohort 1 will assess whether a single booster dose of the mRNA-1273.529 as the first booster dose will elicit a similar or superior antibody response to the B.1.1.529 compared to a single booster dose of mRNA-1273, using an external comparator; Study Part F Cohort 2 will assess whether a single booster dose of the mRNA-1273.529 as a second booster dose will elicit a similar or superior antibody response to the B.1.1.529 compared to a single booster dose of mRNA-1273 as a second booster dose. Study Part G will assess whether a single booster dose of the mRNA-1273.214 as a second booster dose will elicit a similar or superior antibody response to the B.1.1.529 compared to a single booster dose of mRNA-1273 as a second booster dose (Part F, Cohort 2, 50 µg mRNA-1273). Study Part A.2 will assess whether a second booster dose with mRNA-1273.214 (50 µg) in individuals who have previously received the mRNA-1273.211 (50 µg) booster will elicit a robust immune response against SARS-CoV-2. Study Part H will assess whether a single booster dose of the mRNA-1273.222 50 µg as a second booster dose elicits a superior antibody response against Omicron BA.4/5 compared to a single booster dose of mRNA-1273 as a second booster dose (Part F, Cohort 2, 50 µg mRNA-1273). Part J will assess whether a booster dose of mRNA-1273.815 or mRNA-1273.231 will elicit a robust immune response against SARS-CoV-2 Omicron BA.4/BA.5, BQ.1.1, and XBB.1.5 variants in individuals who previously received a primary series of mRNA vaccine, a first booster dose of a monovalent mRNA vaccine, and a second booster dose of a bivalent mRNA vaccine against SARS-CoV-2.

**Background and Overview**

The Sponsor has developed a rapid response, proprietary vaccine platform based on mRNA delivery system. The platform is based on the principle and observations that cells in vivo can take up mRNA, translate it, and then express protein viral antigen(s) on the cell surface. The delivered mRNA does not enter the cellular nucleus or interact with the genome, is nonreplicating, and is expressed transiently.

##### 1.1.1. mRNA-1273

The Sponsor is using its mRNA-based platform to develop a novel LNP-encapsulated mRNA-based vaccine against SARS-CoV-2 (mRNA-1273). mRNA-1273 encodes for the full-length S protein of SARS-CoV-2, modified to introduce 2 proline residues to stabilize the S protein (S-2P) in a prefusion conformation. The CoV-S protein mediates attachment and entry of the virus into host cells (by fusion), making it a primary target for neutralizing antibodies (nAbs) that prevent infection (Corbett et al 2020). It has been confirmed that the stabilized SARS-CoV-2 S2P antigen presents in the correct prefusion conformation (Wrapp et al 2020).

##### 1.1.2. mRNA-1273.211

mRNA-1273.211 is a multivalent product that contains 2 mRNAs: CX-024414 encoding for the S-2P of Wuhan-Hu-1 and CX-027367 encoding for the S-2P of B.1.351, in a 1:1 ratio.

##### 1.1.3. mRNA-1273.617.2

mRNA-1273.617.2 contains mRNA CX-029444 encoding for the S-2P of B.1.617.2.

##### 1.1.4. mRNA-1273.213

mRNA-1273.213 is a multivalent product that contains 2 mRNAs: CX-029444 encoding for the S-2P of B.1.617.2 and CX-027367 encoding for the S-2P of B.1.351, in a 1:1 ratio.

##### 1.1.5. mRNA-1273.529

mRNA-1273.529 contains mRNA CX-031302 encoding for the S-2P of B.1.1.529.

##### 1.1.6. mRNA-1273.214

mRNA-1273.214 contains CX-024414, the mRNA that encodes for the prefusion stabilized spike glycoprotein (S-2P) of the Wuhan-Hu-1 isolate of SARS-CoV-2 and CX-031302, the mRNA that encodes for the S-2P of the SARS-CoV-2 B.1.1.529 variant.

##### 1.1.7. mRNA-1273.222

mRNA-1273.222 contains CX-024414, the mRNA that encodes for the prefusion stabilized spike glycoprotein (S-2P) of the Wuhan-Hu-1 isolate of SARS-CoV-2 and CX-034476, the mRNA that encodes for the S-2P of the SARS-CoV-2 B.1.1.529 subvariants BA.4/BA.5.

##### 1.1.8. mRNA-1273.815

mRNA-1273.815 contains CX-038839, the monovalent mRNA that encodes for the S-2P of the SARS-CoV-2 Omicron subvariants XBB.1.5/XBB.1.9.1.

##### 1.1.9. mRNA-1273.231

mRNA-1273.231 contains CX-034476 and CX-038839, the bivalent mRNA that encodes for the S-2P of the SARS-CoV-2 B.1.1.529 Omicron subvariants BA.4/BA.5 and XBB.1.5/XBB.1.9.1.

#### 1.2. Nonclinical Studies

The Sponsor has conducted preclinical studies to evaluate modified vaccines. The immunogenicity of mRNA-1273.351 (a monovalent product containing mRNA CX-027367) and a 1:1 mix of mRNA-1273 and mRNA-1273.351 was evaluated in BALB/c mice. Both vaccines were evaluated as a 2-dose primary series in mice, and mRNA-1273.351 was also evaluated as a booster dose in animals previously vaccinated with 2 doses of mRNA-1273. The results demonstrated that a primary vaccination series of mRNA-1273.351 was effective at increasing nAb titers against the B.1.351 variant, while a 1:1 mix of mRNA-1273 and mRNA-1273.351 was most effective at providing broad cross-variant neutralization. Studies also demonstrated that a third dose of mRNA-1273.351 significantly increased both Wuhan-Hu-1 and B.1.351-specific neutralization titers (Wu et al 2021b).

Additional studies are being performed in mice, golden Syrian hamsters, and rhesus macaques to further evaluate mRNA-1273, mRNA-1273.351, and mRNA-1273.211 as both a primary series vaccine and as a single booster dose in animals previously vaccinated with a primary mRNA-1273 vaccine series. These studies are to determine the immunogenicity of the primary series or booster vaccines and to evaluate protection from challenge with wild-type and variant viruses. These data will be reported once studies have been completed. A vaccine that contains the spike protein sequence of the B.1.617.2 vaccine has been developed, mRNA-1273.617.2. The immunogenicity of this vaccine is being evaluated as a monovalent product and as a component in a multivalent formulation, mixed with mRNA-1273 or mRNA-1273.351. Additional animal studies are currently being conducted with mRNA vaccines that target different Omicron variants.

#### 1.3. Clinical Studies

The Sponsor has previously evaluated modified Beta variant-matched booster vaccines in the completed Phase 2a clinical study (NCT04405076) and the Sponsor is currently evaluating several modified vaccines (containing spike sequences of the Beta, Delta, and Omicron variants) in the present study, mRNA-1273-P205.

#### 1.4. Benefit/Risk Assessment

##### 1.4.1. Known Potential Benefits

The following benefits may accrue to participants that will receive the mRNA-1273.211, mRNA-1273, mRNA-1273.617.2, mRNA-1273.213, mRNA-1273.529, mRNA-1273.214, mRNA-1273.222, mRNA-1273.815, or mRNA-1273.231 booster vaccine candidates:

- The mRNA-1273.211 vaccine candidate may be an effective vaccine against COVID-19 VOC and may provide an effective immune response against ancestral SARS-CoV-2.

- The mRNA-1273.617.2 vaccine candidate may be an effective vaccine against COVID-19 VOC.
- The mRNA-1273.213 vaccine candidate may be an effective vaccine against COVID-19 VOC.
- The mRNA-1273.529 vaccine candidate may be an effective vaccine against COVID-19 VOC.
- The mRNA-1273.214 vaccine candidate may be an effective vaccine against COVID-19 VOC.
- The mRNA-1273.222 vaccine candidate may be an effective vaccine against COVID-19 VOC.
- The mRNA-1273.815 vaccine candidate may be an effective vaccine against COVID-19 VOC.
- The mRNA-1273.231 vaccine candidate may be an effective vaccine against COVID-19 VOC.
- Participants will have a baseline (Day 1) evaluation for SARS-CoV-2 infection and ongoing surveillance for COVID-19 throughout the study.
- The study will contribute to the development of a vaccine against COVID-19 VOC, a current pandemic disease.

##### 1.4.2. Risks from Study Participation and Their Mitigation

The safety profile of mRNA-1273 is largely based on data from the pivotal Phase 3 Study mRNA-1273-P301 (COVE).

Solicited ARs were reported more frequently among vaccine participants than among placebo participants. The most frequently reported ARs after any dose in the vaccine group were pain at the injection site, fatigue, headache, myalgia and chills. The most common solicited local AR was pain. Solicited systemic ARs were reported more frequently by vaccine participants after dose 2 (fatigue, 65.3%, headache, 58.6%, myalgia, 58% and arthralgia, 42.8%) than after dose 1 (fatigue, 37,2%, headache, 32.7%, myalgia, 22.7% and arthralgia,16.6%). Grade 3 systemic ARs were also reported more frequently after dose 2 than after dose 1. The majority of local and systemic ARs had a median duration of 1 to 3 days.

Overall, there was a higher reported rate of some ARs in younger age groups: the incidence of axillary swelling/tenderness, fatigue, headache, myalgia, arthralgia, chills, nausea/vomiting, and fever was higher in adults aged 18 to < 65 years than in those aged 65 years and above.

Grade 3 solicited local ARs were more frequently reported after dose 2 than after dose 1.

Unsolicited AEs that occurred within 28 days following each vaccination were reported by 23.9% of participants who received mRNA-1273 and 21.6% of participants who received placebo. Unsolicited AEs that occurred in ≥ 1% of study participants who received mRNA-1273 and at a rate at least 1.5-fold higher rate than placebo, were lymphadenopathy related events (1.1% vs. 0.6%). All of the lymphadenopathy events are similar to the axillary swelling/tenderness in the injected arm reported as solicited ARs. Several participants reported injection site reactions after Day 7 that were characterized by erythema, induration and often pruritus. Consultation with a dermatopathologist suggested that these were most likely dermal hypersensitivity and were unlikely to represent a long-term safety concern.

Hypersensitivity AEs were reported in 1.5% of vaccine recipients and 1.1% of placebo recipients. Hypersensitivity events in the vaccine group included injection site rash and injection site urticaria, which are likely related to vaccination. There have been no cases of severe hypersensitivity or anaphylactic reactions reported immediately after vaccination in the trial to date.

There were 3 reports of Bell's palsy in the mRNA-1273 vaccine group (one of which was an SAE), which occurred 22, 28, and 32 days after vaccination, and one in the placebo group which occurred 17 days after vaccination. Currently available information on Bell's palsy is insufficient to determine a causal relationship with the vaccine.

SAEs were reported at the same rates in participants who received mRNA-1273 and placebo from the first dose until the last observation. There were 2 SAEs of facial swelling in vaccine recipients with a history of injection of dermatological fillers. The onset of swelling was reported 1 and 2 days, respectively, after vaccination and was likely related to vaccination. There was 1 SAE of intractable nausea and vomiting in a participant with prior history of severe headache and nausea requiring hospitalization. This event occurred 1 day after vaccination and was likely related to vaccination.

There were no other notable patterns or numerical imbalances between study arms for specific categories of AEs (including other neurologic, neuro-inflammatory, and thrombotic events) that would suggest a causal relationship to mRNA-1273.

In the Post-Authorization setting, anaphylaxis has been reported following mRNA-1273 administration. In addition, there have been very rare reports of myocarditis and pericarditis occurring after vaccination with Moderna COVID-19 Vaccine. Although causality has not been established, the majority of the cases have been reported in young males shortly after the second dose of the vaccine. These are typically mild cases and individuals tend to recover within a short time following standard treatment and rest. Healthcare professionals should be alert to the signs and symptoms of myocarditis and pericarditis. Safety will be monitored throughout the study (Section 7.4).

##### 1.4.3. Overall Benefit/Risk Conclusion

The evolving antigenic variation of SARS-CoV-2 underscores the urgent need for vaccination strategies that induce broader protection, specifically against VOC with attendant risk of viral escape. ModernaTX, Inc. is developing monovalent and multivalent mRNA vaccines (including mRNA-1273.214, mRNA-1273.222, mRNA-1273.815, and mRNA-1273.231) that are similar to the mRNA-1273 vaccine, but in which the mRNA encodes for mutations included in the S protein of both ancestral SARS-CoV-2 (Wuhan-Hu-1) and VOC to enhance protection against COVID-19 caused by SARS-CoV-2 variants.

### 2. OBJECTIVES AND ENDPOINTS

This study consists of 9 parts: A (1, 2), B, C D, E, F, G, H, and J. The objectives and endpoints in each part are described in the tables below, Part E objectives and endpoints are described in a site-specific protocol amendment and are not covered in this global protocol amendment. The objectives and endpoints for Parts A (1, 2), B, C, and D are described below in Table 1, Table 2, Table 3, Table 4, and Table 5, respectively, the objectives and endpoints for Part F (Cohorts 1 and 2) are described in Table 6 and Table 7, the objectives and endpoints for Part G are described in Table 8, the objectives and endpoints for Part H are described in Table 9, and the objectives and endpoints for Part J are described in Table 10.

**Table 1: Part A.1 – 50 µg mRNA-1273.211 and 100 µg mRNA-1273.211**

| Objectives | Endpoints |
|---|---|
| **Primary** | |
| **To be assessed for each dose level of mRNA-1273.211**<br>• To demonstrate non-inferior immune response of a single booster dose of mRNA-1273.211 compared with 2 priming doses of mRNA-1273 in the pivotal Phase 3 efficacy trial (Study mRNA-1273-P301 [COVE]):<br>– To demonstrate non-inferiority based on GMT ratio (mRNA-1273.211 vs. mRNA-1273) against ancestral SARS-CoV-2 with a non-inferiority margin of 1.5<br>– To demonstrate non-inferiority based on the seroresponse rate (SRR) (mRNA-1273.211 – mRNA-1273) against ancestral SARS-CoV-2 with a non-inferiority margin of 10% | **To be assessed for each dose level of mRNA-1273.211**<br>• GMT ratio of GMT of mRNA-1273.211 against ancestral SARS-CoV-2 at Day 29 after the booster dose over GMT of mRNA-1273 against ancestral SARS-CoV-2 at Day 57 (historical control)<br>• SRR difference between mRNA-1273.211 against ancestral SARS-CoV-2 at Day 29 after the booster dose and mRNA-1273 against ancestral SARS-CoV-2 at Day 57 (historical control) |
| **To be assessed for each dose level of mRNA-1273.211**<br>• To demonstrate non-inferior immune response based on GMT ratio of mRNA-1273.211 as a single booster dose against the variant B.1.351, compared to mRNA-1273 after 2 priming doses against ancestral SARS-CoV-2 with a non-inferiority margin of 1.5<br>• To demonstrate non-inferior immune response based on the SRR of a single booster dose of mRNA-1273.211 against | **To be assessed for each dose level of mRNA-1273.211**<br>• GMT ratio of GMT of mRNA-1273.211 against the variant B.1.351 at Day 29 after the booster dose over GMT of mRNA-1273 against ancestral SARS-CoV-2 at Day 57 (historical control).<br>• SRR difference between mRNA-1273.211 against the variant B.1.351 at Day 29 after the booster dose and mRNA-1273 against ancestral SARS-CoV-2 at Day 57 (historical control) |

| Objectives | Endpoints |
|---|---|
| the variant B.1.351 as compared to 2 priming doses of mRNA-1273 against ancestral SARS-CoV-2 with a non-inferiority margin of 10% | |
| • To evaluate the safety and reactogenicity of mRNA-1273.211 | • Solicited local and systemic reactogenicity ARs during a 7-day follow-up period after vaccination<br>• Unsolicited AEs during the 28-day follow-up period after vaccination<br>• SAEs, MAAEs, AEs leading to withdrawal and AESI from Day 1 to EoS |
| **Secondary** | |
| • To compare immune response of mRNA-1273.211 as booster against variants compared to the priming series of mRNA-1273 against ancestral SARS-CoV-2 | • GMT ratio and SRR difference of mRNA-1273.211 as a booster dose against variants compared to the priming series of mRNA-1273 against ancestral SARS-CoV-2 |
| **Exploratory** | |
| • To assess for symptomatic and asymptomatic SARS-CoV-2 infection | • Laboratory-confirmed symptomatic or asymptomatic SARS-CoV-2 infection will be defined in participants:<br>– Primary case definition per the mRNA-1273-P301 (COVE) study<br>– Secondary case definition based on the CDC criteria: the presence of one of the CDC-listed symptoms (CDC 2022) and a positive RT-PCR test on a respiratory sample<br>– Asymptomatic SARS-CoV-2 infection is defined as a positive RT-PCR test on a respiratory sample in the absence of symptoms or a positive serologic test for anti-nucleocapsid antibody after a negative test at time of enrollment |
| • To evaluate the genetic and/or phenotypic relationships of isolated SARS-CoV-2 strains to the vaccine sequence | • Characterize the SARS-CoV-2 genomic sequence of viral isolates and compare with the vaccine sequence<br>• Characterize the immune responses to vaccine breakthrough isolates |
| • Exploratory analyses on the immune response to specific viral strains may be performed to compare different booster regimens<br>– To compare the immune response of 50 µg of mRNA-1273.211 with 100 µg of mRNA-1273.211 against the same viral strains | • GMT ratio and SRR difference of 50 µg mRNA-1273.211 compared to 100 µg against the same viral strains |

| Objectives | Endpoints |
|---|---|
| • To compare immune response of mRNA-1273.211 as booster against variants compared to the priming series of mRNA-1273 against the variants | • GMT ratio and SRR difference of mRNA-1273.211 as a booster dose against variants compared to the priming series of mRNA-1273 against the variants |

**Table 2: Part A.2 – Second booster dose 50 µg mRNA-1273.214: Participants who received an mRNA-1273.211 50 µg as a first booster dose in Part A.1**

| Objectives | Endpoints |
|---|---|
| **Primary** | |
| • To evaluate the immunogenicity of mRNA-1273.214 (50 µg) as a second booster dose in participants who previously received a first booster dose of mRNA-1273.211 (50 µg) | GMT ratio and SRR difference of mRNA-1273.214 (50 µg) as a second booster dose against the ancestral SARS-CoV-2 and against SARS-CoV-2 variants (including Omicron) compared to mRNA-1273.211 (50 µg) against the ancestral SARS-CoV-2 and against SARS-CoV-2 variants as the first booster dose (Day 29, Day 181) |
| • To assess the safety and reactogenicity of the mRNA-1273.214 (50 µg) given as a second booster dose in participants who previously received a first booster dose of mRNA-1273.211 (50 µg) | • Solicited local and systemic reactogenicity ARs during a 7-day follow-up period after vaccination<br>• Unsolicited AEs during the 28-day follow-up period after vaccination<br>• SAEs, MAAEs, AEs leading to withdrawal and AESI from Day 1 to EoS |
| **Secondary** | |
| • To evaluate the immunogenicity of mRNA-1273.214 (50 µg) as a second booster dose in participants who previously received a first booster dose of mRNA-1273.211 (50 µg) | • Antibody response of the mRNA-1273.214 (50 µg) against the ancestral SARS-CoV-2 and against SARS-CoV-2 variants (including Omicron) by GMT and SRR at multiple time points after the mRNA-1273.214 booster dose |
| **Exploratory** | |
| • To assess for symptomatic and asymptomatic SARS-CoV-2 infection | • Laboratory-confirmed symptomatic or asymptomatic SARS-CoV-2 infection will be defined in participants:<br>– Primary case definition per the mRNA-1273-P301 (COVE) study<br>– Secondary case definition based on the CDC criteria: the presence of one of the CDC-listed symptoms (CDC 2022) and a positive RT-PCR test on a respiratory sample<br>– Asymptomatic SARS-CoV-2 infection is defined as a positive RT-PCR test on a respiratory sample in the absence of symptoms or a positive serologic test for anti-nucleocapsid antibody after a negative test at time of enrollment |

| Objectives | Endpoints |
|---|---|
| • To evaluate the genetic and/or phenotypic relationships of isolated SARS-CoV-2 isolates to the vaccine sequence | • Characterize the SARS-CoV-2 genomic sequence of viral isolates and compare with the vaccine sequence<br>• Characterize the immune responses to vaccine breakthrough isolates |

**Table 3: Part B – 100 µg mRNA-1273**

| Objectives | Endpoints |
|---|---|
| **Primary** | |
| • To demonstrate non-inferior immune response of a single booster dose of 100 µg mRNA-1273 compared with 2 priming doses of mRNA-1273 in the pivotal Phase 3 efficacy trial (Study mRNA-1273-P301 [COVE]):<br>– To demonstrate non-inferiority based on GMT ratio (100 µg mRNA-1273 single booster dose vs. 100 µg primary series mRNA-1273) against ancestral SARS-CoV-2 with a non-inferiority margin of 1.5<br>– To demonstrate non-inferiority based on the SRR (100 µg mRNA-1273 single booster dose vs. 100 µg primary series mRNA-1273) against ancestral SARS-CoV-2 with a non-inferiority margin of 10% | • GMT ratio of 100 µg mRNA-1273 against ancestral SARS-CoV-2 at Day 29 after the booster dose over GMT of 100 µg mRNA-1273 against ancestral SARS-CoV-2 at Day 57 (historical control)<br>• SRR difference between 100 µg mRNA-1273 against ancestral SARS-CoV-2 at Day 29 after the booster dose and mRNA-1273 against ancestral SARS-CoV-2 at Day 57 (historical control) |
| • To evaluate the safety and reactogenicity of mRNA-1273 | • Solicited local and systemic reactogenicity ARs during a 7-day follow-up period after vaccination<br>• Unsolicited AEs during the 28-day follow-up period after vaccination<br>• SAEs, MAAEs, AEs leading to withdrawal and AESI from Day 1 to EoS |
| **Secondary** | |
| • To compare immune response of 100 µg mRNA-1273 as booster against variants compared to the priming series of mRNA-1273 against ancestral SARS-CoV-2 | • GMT ratio and SRR difference of 100 µg mRNA-1273 as a booster dose against variants compared to the priming series of mRNA-1273 against ancestral SARS-CoV-2 |
| **Exploratory** | |
| • To assess for symptomatic and asymptomatic SARS-CoV-2 infection | • Laboratory-confirmed symptomatic or asymptomatic SARS-CoV-2 infection will be defined in participants: |

| Objectives | Endpoints |
|---|---|
| | – Primary case definition per the mRNA-1273-P301 (COVE) study<br>– Secondary case definition based on the CDC criteria: the presence of one of the CDC-listed symptoms (CDC 2022) and a positive RT-PCR test on a respiratory sample<br>– Asymptomatic SARS-CoV-2 infection is defined as a positive RT-PCR test on a respiratory sample in the absence of symptoms or a positive serologic test for anti-nucleocapsid antibody after a negative test at time of enrollment |
| • To evaluate the genetic and/or phenotypic relationships of isolated SARS-CoV-2 strains to the vaccine sequence | • Characterize the SARS-CoV-2 genomic sequence of viral isolates and compare with the vaccine sequence<br>• Characterize the immune responses to vaccine breakthrough isolates |
| • Exploratory analyses on the immune response to specific viral strains may be performed to compare different booster regimens<br>– To compare the immune response to ancestral SARS-CoV-2 and variants after a single booster dose of 50 or 100 µg mRNA-1273.211 (Part A.1) vs. a single booster dose of 100 µg mRNA-1273 | • GMT ratio and SRR difference of 50 or 100 µg mRNA-1273.211 (Part A.1) as a booster dose against ancestral SARS-CoV-2 and variants compared to a booster dose of 100 µg mRNA-1273 against ancestral SARS-CoV-2 and variants |
| • To compare immune response of mRNA-1273 as booster against variants compared to the priming series of mRNA-1273 against the variants | • GMT ratio and SRR difference of mRNA-1273 as a booster dose against variants compared to the priming series of mRNA-1273 against the variants |

**Table 4: Part C – 50 µg mRNA-1273.617.2 and 100 µg mRNA-1273.617.2**

| Objectives | Endpoints |
|---|---|
| **Primary** | |
| **To be assessed for each dose level of mRNA-1273.617.2**<br>• To demonstrate non-inferior immune response of a single booster dose of mRNA-1273.617.2 compared with 2 priming doses of mRNA-1273 in the pivotal Phase 3 efficacy trial (Study mRNA-1273-P301 [COVE]): | **To be assessed for each dose level of mRNA-1273.617.2**<br>• GMT ratio of mRNA-1273.617.2 against the variant B.1.617.2 at Day 29 after the booster dose over GMT of 100 µg mRNA-1273 against ancestral SARS-CoV-2 at Day 57 (historical control)<br>• SRR difference between mRNA-1273.617.2 against the variant at Day 29 after the booster |

| Objectives | Endpoints |
|---|---|
| – To demonstrate non-inferiority based on GMT ratio of mRNA1273.617.2 as a single booster dose against the variant B.1.617.2 vs. 100 µg primary series mRNA1273) against ancestral SARS-CoV-2 with a non-inferiority margin of 1.5<br>– To demonstrate non-inferiority based on the SRR of a single booster dose of mRNA1273.617.2 against the variant B.1.617.2 vs. 100 µg primary series mRNA1273 against ancestral SARS-CoV-2 with a non-inferiority margin of 10% | dose and mRNA-1273 against ancestral SARS-CoV-2 at Day 57 (historical control) |
| • To evaluate the safety and reactogenicity of mRNA-1273.617.2 | • Solicited local and systemic reactogenicity ARs during a 7-day follow-up period after vaccination<br>• Unsolicited AEs during the 28-day follow-up period after vaccination<br>• SAEs, MAAEs, AEs leading to withdrawal and AESI from Day 1 to EoS |
| **Secondary** | |
| • To compare immune response of mRNA-1273.617.2 as booster against ancestral SARS-CoV-2 and variants compared to the priming series of mRNA-1273 against ancestral SARS-CoV-2 | • GMT ratio and SRR difference of mRNA-1273.617.2 as a booster dose ancestral SARS-CoV-2 and variants compared to the priming series of mRNA-1273 against ancestral SARS-CoV-2 |
| **Exploratory** | |
| • To assess for symptomatic and asymptomatic SARS-CoV-2 infection | • Laboratory-confirmed symptomatic or asymptomatic SARS-CoV-2 infection will be defined in participants:<br>– Primary case definition per the mRNA-1273-P301 (COVE) study<br>– Secondary case definition based on the CDC criteria: the presence of one of the CDC-listed symptoms (CDC 2022) and a positive RT-PCR test on a respiratory sample<br>– Asymptomatic SARS-CoV-2 infection is defined as a positive RT-PCR test on a respiratory sample in the absence of symptoms or a positive serologic test for anti-nucleocapsid antibody after a negative test at time of enrollment |

| Objectives | Endpoints |
|---|---|
| • To evaluate the genetic and/or phenotypic relationships of isolated SARS-CoV-2 strains to the vaccine sequence | • Characterize the SARS-CoV-2 genomic sequence of viral isolates and compare with the vaccine sequence<br>• Characterize the immune responses to vaccine breakthrough isolates |
| • Exploratory analyses on the immune response to specific viral strains may be performed to compare different booster regimens<br>– To compare the immune response of 50 µg of mRNA-1273.617.2 with 100 µg of mRNA-1273.617.2 against the same viral strains | • GMT ratio and SRR difference of 50 µg mRNA-1273.617.2 compared to 100 µg against the same viral strains |
| • Exploratory analyses on the immune response to specific viral strains may be performed to compare different booster regimens<br>– To compare immune response to ancestral SARS-CoV-2 and variants after a single booster dose of mRNA-1273.617.2 vs. a single booster dose of 100 µg mRNA-1273 (Part B) | • GMT ratio and SRR difference of mRNA-1273.617.2 as a booster dose against ancestral SARS-CoV-2 and variants compared to a booster dose of 100 µg mRNA-1273 (Part B) against the ancestral SARS-CoV-2 and variant |
| • Exploratory analyses on the immune response to specific viral strains may be performed to compare different booster regimens:<br>– To compare immune response to ancestral SARS-CoV-2 and variants after a single booster dose of mRNA-1273.617.2 vs. a single booster dose of 50 or 100 µg mRNA-1273.211 (Part A.1) | • GMT ratio and SRR difference of mRNA-1273.617.2 as a booster dose against ancestral SARS-CoV-2 and variants compared to a booster dose of 50 or 100 µg mRNA-1273.211 (Part A.1) against ancestral SARS-CoV-2 e and variants |
| • To compare immune response of mRNA-1273.617.2 as a booster against variants compared to the priming series of mRNA-1273 against the variants | • GMT ratio and SRR difference of mRNA-1273.617.2 as a booster dose against variants compared to the priming series of mRNA-1273 against the variants |

**Table 5: Part D – 50 µg mRNA-1273.213 and 100 µg mRNA-1273.213**

| Objectives | Endpoints |
|---|---|
| **Primary** | |
| **To be assessed for each dose level of mRNA-1273.213** | **To be assessed for each dose level of mRNA-1273.213** |

| Objectives | Endpoints |
|---|---|
| • To demonstrate non-inferior immune response of a single booster dose of mRNA-1273.213 compared with 2 priming doses of mRNA-1273 in the pivotal Phase 3 efficacy trial (Study mRNA-1273-P301 [COVE]):<br>– To demonstrate non-inferiority based on GMT ratio of mRNA-1273.213 as a single booster dose against the variant B.1.617.2 vs. 100 µg primary series mRNA-1273 against ancestral SARS-CoV-2 with a non-inferiority margin of 1.5<br>– To demonstrate non-inferiority based on the SRR of a mRNA-1273.213 single booster dose against the variant B.1.617.2 vs. 100 µg primary series mRNA-1273 against ancestral SARS-CoV-2 with a non-inferiority margin of 10%<br>– To demonstrate non-inferiority based on GMT ratio of mRNA1273.213 as a single booster dose against the variant B.1.351 vs. 100 µg primary series mRNA1273 against ancestral SARS-CoV-2 with a non-inferiority margin of 1.5<br>– To demonstrate non-inferiority based on the SRR of a mRNA1273.213 single booster dose against the variant B.1.351 vs. 100 µg primary series mRNA-1273 against ancestral SARS-CoV-2 with a non-inferiority margin of 10% | • GMT ratio of mRNA-1273.213 against the variant B.1.617.2 at Day 29 after the booster dose over GMT of 100 µg mRNA-1273 against ancestral SARS-CoV-2 at Day 57 (historical control)<br>• SRR difference between mRNA-1273.213 against the variant B.1.617.2 at Day 29 after the booster dose and mRNA-1273 against ancestral SARS-CoV-2 at Day 57 (historical control)<br>• GMT ratio of mRNA-1273.213 against the variant B.1.351 at Day 29 after the booster dose over GMT of 100 µg mRNA-1273 against ancestral SARS-CoV-2 at Day 57 (historical control)<br>• SRR difference between mRNA-1273.213 against the variant B.1.351 at Day 29 after the booster dose and mRNA-1273 against ancestral SARS-CoV-2 at Day 57 (historical control) |
| • To evaluate the safety and reactogenicity of mRNA-1273.213 | • Solicited local and systemic reactogenicity ARs during a 7-day follow-up period after vaccination<br>• Unsolicited AEs during the 28-day follow-up period after vaccination<br>• SAEs, MAAEs, AEs leading to withdrawal and AESI from Day 1 to EoS |
| **Secondary** | |
| • To compare the immune response of mRNA-1273.213 as booster ancestral SARS-CoV-2 and other variants compared to the priming series of mRNA-1273 against ancestral SARS-CoV-2 | • GMT ratio and SRR difference of mRNA-1273.213 as a booster dose against ancestral SARS-CoV-2 and other variants compared to the priming series of mRNA-1273 against ancestral SARS-CoV-2 |

| Objectives | Endpoints |
|---|---|
| **Exploratory** | |
| • To assess for symptomatic and asymptomatic SARS-CoV-2 infection | • Laboratory-confirmed symptomatic or asymptomatic SARS-CoV-2 infection will be defined in participants:<br>– Primary case definition per the mRNA-1273-P301 (COVE) study<br>– Secondary case definition based on the CDC criteria: the presence of one of the CDC-listed symptoms (CDC 2022) and a positive RT-PCR test on a respiratory sample<br>• Asymptomatic SARS-CoV-2 infection is defined as a positive RT-PCR test on a respiratory sample in the absence of symptoms or a positive serologic test for anti-nucleocapsid antibody after a negative test at time of enrollment |
| • To evaluate the genetic and/or phenotypic relationships of isolated SARS-CoV-2 strains to the vaccine sequence | • Characterize the SARS-CoV-2 genomic sequence of viral isolates and compare with the vaccine sequence<br>• Characterize the immune responses to vaccine breakthrough isolates |
| • Exploratory analyses on the immune response to specific viral strains may be performed to compare different booster regimens<br>• To compare the immune response of 50 µg of mRNA-1273.213 with 100 µg of mRNA-1273.213 against the same viral strains | • GMT ratio and SRR difference of 50 µg mRNA-1273.213 compared to 100 µg against the same viral strains |
| • Exploratory analyses on the immune response to specific viral strains may be performed to compare different booster regimens<br>– To compare immune response to ancestral SARS-CoV-2 and variants after a single booster dose of mRNA-1273.213 vs. a single booster dose of 50 or 100 µg mRNA-1273.211 (Part A.1)<br>– To compare immune response to ancestral SARS-CoV-2 and variants after a single booster dose of mRNA-1273.213 vs. a single booster dose of 100 µg mRNA-1273 (Part B)<br>• To compare immune response to ancestral SARS-CoV-2 and variants after a single | • GMT ratio and SRR difference of mRNA-1273.213 as a booster dose against ancestral SARS-CoV-2 and variants compared to a booster dose of 50 or 100 µg mRNA-1273.211 (Part A.1) against ancestral SARS-CoV-2 and variants<br>• GMT ratio and SRR difference of mRNA-1273.213 as a booster dose against ancestral SARS-CoV-2 and variants compared to a booster dose of 100 µg mRNA-1273 (Part B) against ancestral SARS-CoV-2 and variant<br>• GMT ratio and SRR difference of mRNA-1273.213 as a booster dose against ancestral SARS-CoV-2 and variants compared to a booster dose of 50 or 100 µg mRNA-1273.617.2 (Part C) against ancestral SARS-CoV-2 and variant |

| Objectives | Endpoints |
|---|---|
| booster dose of mRNA-1273.213 vs. a single booster dose of 50 or 100 µg mRNA-1273.617.2 (Part C) | |
| • To compare immune response of mRNA-1273.213 as a booster against variants compared to the priming series of mRNA-1273 against the variants | • GMT ratio and SRR difference of mRNA-1273.213 as a booster dose against variants compared to the priming series of mRNA-1273 against the variants |

**Table 6: Part F – Cohort 1 (first booster dose): Participants Who Previously Received 100 µg mRNA-1273 Primary Series and Have Not Received a Booster Dose Previously (50 and 100 µg mRNA-1273.529)**

| Objectives | Endpoints |
|---|---|
| **Primary** | |
| • To demonstrate non-inferiority of the antibody response against the Omicron variant (B.1.1.529) of a first booster dose of mRNA-1273.529 compared to a first booster dose of mRNA-1273 (50 µg) based on GMT ratio and SRR difference<br>• To demonstrate superiority of the antibody response against the Omicron variant (B.1.1.529) of a first booster dose of mRNA-1273.529 compared a first booster dose of mRNA-1273 (50 µg) based on GMT ratio | • Day 29 postboost GMT ratio of Omicron-specific GMT of mRNA-1273.529 over the Omicron-specific GMT of mRNA-1273 (historical mRNA-1273 booster dose control)<br>• Day 29 SRR difference between mRNA-1273.529 against Omicron and mRNA-1273 against Omicron |
| • To evaluate the safety and reactogenicity of mRNA-1273.529 | • Solicited local and systemic reactogenicity ARs during a 7-day follow-up period after vaccination<br>• Unsolicited AEs during the 28-day follow-up period after vaccination<br>• SAEs, MAAEs, AEs leading to withdrawal and AESI from Day 1 to EoS |
| **Secondary** | |
| • To evaluate the immunogenicity of a mRNA-1273.529 dose compared to a mRNA-1273 administered as a first booster dose at all timepoints postboost | • GMT ratio of mRNA-1273.529 and mRNA-1273 against the Omicron variant at all timepoints postboost<br>• SRR difference between mRNA-1273.529 against the Omicron variant and mRNA-1273 against the Omicron variant<br>• GMT ratio of mRNA-1273.529 and mRNA-1273 against the ancestral SARS-CoV-2 and other variants at all timepoints postboost<br>• SRR difference between mRNA-1273.529 against the ancestral SARS-CoV-2 and other |

| Objectives | Endpoints |
|---|---|
| | variants and mRNA-1273 against the ancestral SARS-CoV-2 and other variants |
| • To compare the immune response of mRNA-1273.529 as a first booster dose against the Omicron variant compared to the priming series of mRNA-1273 | • GMT ratio and SRR difference of mRNA-1273.529 as a booster dose against the Omicron variant compared to the priming series of mRNA-1273 against the ancestral SARS-CoV-2 (historical control) |
| **Exploratory** | |
| • To assess for symptomatic and asymptomatic SARS-CoV-2 infection | • Laboratory-confirmed symptomatic or asymptomatic SARS-CoV-2 infection will be defined in participants:<br>– Primary case definition per the mRNA-1273-P301 (COVE) study<br>– Secondary case definition based on the CDC criteria: the presence of one of the CDC-listed symptoms (CDC 2022) and a positive RT-PCR test on a respiratory sample<br>– Asymptomatic SARS-CoV-2 infection is defined as a positive RT-PCR test on a respiratory sample in the absence of symptoms or a positive serologic test for anti-nucleocapsid antibody after a negative test at time of enrollment |
| • To evaluate the genetic and/or phenotypic relationships of isolated SARS-CoV-2 strains to the vaccine sequence | • Characterize the SARS-CoV-2 genomic sequence of viral isolates and compare with the vaccine sequence<br>• Characterize the immune responses to vaccine breakthrough isolates |
| • To characterize the cellular immune response of the mRNA-1273.529 booster dose against the ancestral SARS-CoV-2 and against variants | • T-cell and B-cell response after the mRNA-1273.529 booster |

**Table 7: Part F–- Cohort 2 (second booster dose): Participants Who Previously Received 100 µg mRNA-1273 Primary Series Plus 1 Booster Dose of 50 µg mRNA-1273 (50 µg mRNA-1273.529 and 50 µg mRNA-1273)**

| Objectives | Endpoints |
|---|---|
| **Primary** | |
| • To demonstrate non-inferiority of the antibody response against the Omicron variant (B.1.1.529) of a second booster dose of mRNA-1273.529 compared to a second booster dose of mRNA-1273 (50 µg) based on GMT ratio and SRR difference | • Day 29 postboost GMT ratio of Omicron-specific GMT of mRNA-1273.529 over the Omicron-specific GMT of mRNA-1273 |

| Objectives | Endpoints |
|---|---|
| • To demonstrate superiority of the antibody response against the Omicron variant (B.1.1.529) of a second booster dose of mRNA-1273.529 compared to a second booster dose of mRNA-1273 (50 µg) based on GMT ratio | • Day 29 SRR difference between mRNA-1273.529 against Omicron and mRNA-1273 against Omicron |
| • To evaluate the safety and reactogenicity of mRNA-1273.529 | • Solicited local and systemic reactogenicity ARs during a 7-day follow-up period after vaccination<br>• Unsolicited AEs during the 28-day follow-up period after vaccination<br>• SAEs, MAAEs, AEs leading to withdrawal and AESI from Day 1 to EoS |
| **Secondary** | |
| • To evaluate the immunogenicity of mRNA-1273.529 booster compared to mRNA-1273 booster administered as a second booster dose at all timepoints postboost | • GMT ratio of mRNA-1273.529 and mRNA-1273 against the Omicron variant at all timepoints postboost<br>• SRR difference between mRNA-1273.529 against the Omicron variant and mRNA-1273 against the Omicron variant<br>• GMT ratio of mRNA-1273.529 and mRNA-1273 against the ancestral SARS-CoV-2 and other variants at all timepoints postboost<br>• SRR difference between mRNA-1273.529 against the ancestral SARS-CoV-2 and other variants and mRNA-1273 against the ancestral SARS-CoV-2 and other variants |
| • To compare the immune response of mRNA-1273.529 as a second booster dose against the Omicron variant compared to the priming series of mRNA-1273 | • GMT ratio and SRR difference of mRNA-1273.529 as a second booster dose against the Omicron variant compared to the priming series of mRNA-1273 against the ancestral SARS-CoV-2 (historical control). |
| **Exploratory** | |
| • To assess for symptomatic and asymptomatic SARS-CoV-2 infection | • Laboratory-confirmed symptomatic or asymptomatic SARS-CoV-2 infection will be defined in participants:<br>– Primary case definition per the mRNA-1273-P301 (COVE) study<br>– Secondary case definition based on the CDC criteria: the presence of one of the CDC-listed symptoms (CDC 2022) ) and a positive RT-PCR test on a respiratory sample<br>– Asymptomatic SARS-CoV-2 infection is defined as a positive RT-PCR test on a |

| Objectives | Endpoints |
|---|---|
| | respiratory sample in the absence of symptoms or a positive serologic test for anti-nucleocapsid antibody after a negative test at time of enrollment |
| • To evaluate the genetic and/or phenotypic relationships of isolated SARS-CoV-2 strains to the vaccine sequence | • Characterize the SARS-CoV-2 genomic sequence of viral isolates and compare with the vaccine sequence<br>• Characterize the immune responses to vaccine breakthrough isolates |
| • To characterize the cellular immune response of mRNA-1273.529 as a booster against SARS-CoV-2 and other variants | • T-cell and B-cell response after the mRNA-1273.529 booster |

**Table 8: Part G – Second Booster Dose 50 µg mRNA-1273.214: Participants Who Received 100 µg mRNA-1273 Primary Series and a Booster Dose of 50 µg mRNA-1273**

| Objectives | Endpoints |
|---|---|
| **Primary (**see Figure 1**)** | |
| • To demonstrate non-inferiority of the antibody response of a second booster dose of mRNA-1273.214 compared to mRNA-1273 (50 µg) when administered as a second booster dose against the Omicron variant (B.1.1.529) at Day 29 or Day 91 based on GMT ratio and SRR difference at Day 29 or Day 91<br>• To demonstrate superiority of the antibody response of a second booster dose of mRNA-1273.214 compared to mRNA-1273 (50 µg) administered as a second booster dose against the Omicron variant (B.1.1.529) based on GMT ratio at Day 29 or Day 91<br>• To demonstrate non-inferiority of the antibody response of a second booster dose of mRNA-1273.214 compared to mRNA-1273 (50 µg) when administered as a second booster dose against ancestral SARS-CoV-2 based on GMT ratio at Day 29 or Day 91 | • GMT ratio of Omicron-specific GMT of mRNA-1273.214 over the Omicron-specific GMT of mRNA-1273 (Part F, Cohort 2, 50 µg mRNA-1273) at Day 29 and Day 91<br>• SRR difference between mRNA-1273.214 against Omicron variant and mRNA-1273 against Omicron variant at Day 29 and Day 91<br>• GMT ratio of ancestral SARS-CoV-2 GMT of mRNA-1273.214 over ancestral SARS-CoV-2 GMT of mRNA-1273 (Part F, Cohort 2, 50 µg mRNA-1273) at Day 29 or Day 91 |
| • To evaluate the safety and reactogenicity of mRNA-1273.214 | • Solicited local and systemic reactogenicity ARs during a 7-day follow-up period after vaccination<br>• Unsolicited AEs during the 28-day follow-up period after vaccination<br>• SAEs, MAAEs, AEs leading to withdrawal and |

| Objectives | Endpoints |
|---|---|
| | AESI from Day 1 to EoS |
| **Key Secondary** | |
| • To demonstrate non-inferiority based on the SRR against ancestral SARS-CoV-2 of a second booster dose of mRNA-1273.214 compared to a second booster dose of mRNA-1273 (50 µg) at Day 29 or Day 91 | • SRR difference between mRNA-1273.214 against ancestral SARS-CoV-2 and mRNA-1273 against ancestral SARS-CoV-2 at Day 29 and Day 91 |
| **Secondary** | |
| • To evaluate the immunogenicity of mRNA-1273.214 booster compared to mRNA-1273 booster administered as a second booster dose at all timepoints postboost | • GMT ratio of mRNA-1273.214 and mRNA-1273 against the Omicron variant at all timepoints postboost<br>• SRR difference between mRNA-1273.214 against the Omicron variant and mRNA-1273 against the Omicron variant at all timepoints postboost<br>• GMT ratio of mRNA-1273.214 and mRNA-1273 against ancestral SARS-CoV-2 and other variants at all timepoints postboost<br>• SRR difference between mRNA-1273.214 against ancestral SARS-CoV-2 and other variants and mRNA-1273 against ancestral SARS-CoV-2 and other variants at all timepoints postboost |
| • To compare the immune response of mRNA-1273.214 as a second dose against the Omicron variant compared to the priming series of mRNA-1273 | • GMT ratio and SRR difference of mRNA-1273.214 as a second booster dose against the Omicron variant compared to the priming series of mRNA-1273 against ancestral SARS-CoV-2 (historical control group) |
| **Exploratory** | |
| • To assess for symptomatic and asymptomatic SARS-CoV-2 infection | • Laboratory-confirmed symptomatic or asymptomatic SARS-CoV-2 infection will be defined in participants:<br>– Primary case definition per the mRNA-1273-P301 (COVE) study<br>– Secondary case definition based on the CDC criteria: the presence of one of the CDC-listed symptoms (CDC 2022) ) and a positive RT-PCRtest on a respiratory sample<br>• Asymptomatic SARS-CoV-2 infection is defined as a positive RT-PCR test on a respiratory sample in the absence of symptoms or a positive serologic test for anti-nucleocapsid antibody after a negative test at time of enrollment |

| Objectives | Endpoints |
|---|---|
| • To evaluate the genetic and/or phenotypic relationships of isolated SARS-CoV-2 strains to the vaccine sequence | • Characterize the SARS-CoV-2 genomic sequence of viral isolates and compare with the vaccine sequence<br>• Characterize the immune responses to vaccine breakthrough isolates |
| • To characterize the cellular immune response of mRNA-1273.214 as a booster against SARS-CoV-2 and other variants | • T-cell and B-cell response after the mRNA-1273.214 booster |

**Table 9: Part H - Second Booster Dose 50 µg mRNA-1273.222: Participants Who Received 100 µg mRNA-1273 Primary Series and a Booster Dose of 50 µg mRNA-1273**

| Objectives | Endpoints |
|---|---|
| **Primary** | |
| • To demonstrate non-inferiority of the antibody response of a second booster dose of mRNA-1273.222 50 µg compared to mRNA-1273 50 µg when administered as a second booster dose against Omicron BA.4/5 based on GMT ratio and SRR difference at Day 29<br>• To demonstrate superiority of the antibody response of a second booster dose of mRNA-1273.222 compared to mRNA-1273 (50 µg) administered as a second booster dose against the Omicron BA.4/5 based on GMT ratio at Day 29<br>• To demonstrate non-inferiority of the antibody response of mRNA-1273.222 50 µg compared to mRNA-1273 50 µg when administered as a second booster dose against the ancestral SARS-CoV-2 D614G based on GMT ratio and SRR difference at Day 29 | • GMT ratio of Omicron BA.4/5 GMT of mRNA-1273.222 over the Omicron BA.4/5 GMT of mRNA-1273 (Part F, Cohort 2, 50 µg mRNA-1273) at Day 29<br>• SRR difference between mRNA-1273.222 against Omicron BA.4/5 and mRNA-1273 against Omicron BA.4/5 at Day 29<br>• GMT ratio of the ancestral SARS-CoV-2 D614G GMT of mRNA-1273.222 over the ancestral SARS-CoV-2 D614G GMT of mRNA-1273 at Day 29<br>• SRR difference between mRNA-1273.222 and mRNA-1273 against the ancestral SARS-CoV-2 D614G at Day 29 |
| • To evaluate the safety and reactogenicity of mRNA-1273.222 50 µg | • Solicited local and systemic reactogenicity ARs during a 7-day follow-up period after injection<br>• Unsolicited AEs during the 28-day follow-up period after injection<br>• SAEs, MAAEs, AEs leading to withdrawal, and AESI from Day 1 to EoS |
| **Secondary** | |
| • To evaluate the immunogenicity of mRNA-1273.222 50 µg as a second booster dose against the ancestral SARS-CoV-2 (and other variants) compared to a second booster | • GMT ratio of mRNA-1273.222 50 µg and mRNA-1273 50 µg against ancestral SARS-CoV-2 (and other variants) at all timepoints postbooster |

| Objectives | Endpoints |
|---|---|
| dose of mRNA-1273 50 µg at all timepoints postbooster | • SRR difference between mRNA-1273.222 50 µg and mRNA-1273 50 µg against ancestral SARS-CoV-2 and variants of concern at all timepoints postbooster |
| **Exploratory** | |
| • To assess for symptomatic and asymptomatic SARS-CoV-2 infection | • Laboratory-confirmed symptomatic or asymptomatic SARS-CoV-2 infection will be defined in participants:<br>– Primary case definition per the mRNA-1273-P301 (COVE) study<br>– Secondary case definition based on the CDC criteria: the presence of 1 of the CDC-listed symptoms (CDC 2022) ) and a positive RT-PCR test on a respiratory sample<br>– Asymptomatic SARS-CoV-2 infection is defined as a positive RT-PCR test on a respiratory sample in the absence of symptoms or a positive serologic test for antinucleocapsid antibody after a negative test at time of enrollment |
| • To evaluate the genetic and/or phenotypic relationships of isolated SARS-CoV-2 strains to the vaccine sequence | • Characterize the SARS-CoV-2 genomic sequence of viral isolates and compare with the vaccine sequence<br>• Characterize the immune responses to vaccine breakthrough isolates |

**Table 10: Part J –- Booster dose of mRNA-1273.815 (50 µg) or mRNA-1273.231 (50 µg): Participants who previously received a primary series of mRNA vaccine, a first booster dose of a monovalent mRNA vaccine, and a second booster dose of a bivalent mRNA vaccine against SARS-CoV-2**

| Objectives | Endpoints |
|---|---|
| **Primary** | |
| • To evaluate the immunogenicity of mRNA-1273.815 (50 µg) and mRNA-1273.231 (50 µg) against SARS-CoV-2 Omicron BA.4/BA.5, BQ.1.1, and XBB.1.5 subvariants at Day 15 and Day 29 | • Antibody response of mRNA-1273.815 (50 µg) and mRNA-1273.231 (50 µg) against SARS-CoV-2 Omicron BA.4/BA.5, BQ.1.1, and XBB.1.5 subvariants by GMT, GMFR, and SRR at Day 15 and Day 29 |
| • To evaluate the safety and reactogenicity of mRNA-1273.815 (50 µg) and mRNA-1273.231 (50 µg) | • Solicited local and systemic reactogenicity ARs during a 7-day follow-up period after injection<br>• Unsolicited AEs during the 28-day follow-up period after injection<br>• SAEs, MAAEs, AEs leading to withdrawal, and AESI from Day 1 to Day 181 (EoS) |
| **Exploratory** | |
| • To evaluate the immunogenicity of mRNA-1273.815 (50 µg) and mRNA-1273.231 | • Antibody response of mRNA-1273.815 (50 µg) and mRNA-1273.231 (50 µg) against |

| Objectives | Endpoints |
|---|---|
| (50 µg) against SARS-CoV-2 variants at any study timepoint | SARS-CoV-2 variants by GMT, GMFR, and SRR at any study timepoint |
| • To assess for symptomatic and asymptomatic SARS-CoV-2 infection | • Laboratory-confirmed symptomatic or asymptomatic SARS-CoV-2 infection will be defined in participants:<br>– Primary case definition per the mRNA-1273-P301 (COVE) study<br>– Secondary case definition based on the CDC criteria: the presence of 1 of the CDC-listed symptoms (CDC 2022) and a positive RT-PCR test on a respiratory sample<br>– Asymptomatic SARS-CoV-2 infection is defined as a positive RT-PCR test on a respiratory sample in the absence of symptoms or a positive serologic test for antinucleocapsid antibody after a negative test at time of enrollment |

### 3. STUDY DESIGN

#### 3.1. General Design

This is an open-label, Phase 2/3 study to evaluate the immunogenicity, safety, and reactogenicity of mRNA-1273.211, mRNA-1273, mRNA-1273.617.2, mRNA-1273.213, mRNA-1273.529, mRNA-1273.214, mRNA-1273.222, mRNA-1273.815, and mRNA-1273.231, administered as booster doses.

**Part A.1**

Part A.1 will evaluate the immunogenicity, safety, and reactogenicity of 2 dose levels of the mRNA-1273.211 vaccine candidate when administered as a single booster dose to adult participants of the mRNA-1273-P301 (COVE) study who have previously received 2 doses of mRNA-1273 as a primary series. Two dose levels (50 or 100 µg total mRNA content) of the mRNA-1273.211 booster will be evaluated in this study. Enrollment will begin with the mRNA-1273.211 50 µg dose arm, followed by the enrollment of the mRNA-1273.211 100 µg dose arm. The results of the mRNA-1273.211 vaccine candidate will be compared to the immunogenicity induced after a 2-dose primary series of mRNA-1273 in the mRNA-1273-P301 (COVE) study, which demonstrated the efficacy of mRNA-1273. See the IB for further details.

**Part A.2**

Part A.2 will evaluate the immunogenicity, safety, and reactogenicity of the mRNA-1273.214 vaccine candidate when administered as a second booster dose to adult participants of the mRNA-1273-P205 study who have previously received 2 doses of mRNA-1273 as a primary series and a first booster dose (50 µg total mRNA content) of the mRNA-1273.211 in Part A.1 of this study.

**Part B**

Part B will evaluate the immunogenicity, safety, and reactogenicity of the mRNA-1273 vaccine when administered as a single booster dose to adult participants of the mRNA-1273-P301 (COVE) study who have previously received 2 doses of mRNA-1273 as a primary series. Enrollment of Part B will begin upon completion of enrollment of Part A.1 of the study. The results of the mRNA-1273 vaccine will be compared to the immunogenicity induced after a 2-dose primary series of mRNA-1273 in the mRNA-1273-P301 (COVE) study, which demonstrated the efficacy of mRNA-1273. IB for further details.

**Part C**

Part C will evaluate the immunogenicity, safety, and reactogenicity of 2 dose levels (50 or 100 µg) of the mRNA-1273.617.2 vaccine when administered as a single booster dose to adults who have previously received 2 doses of mRNA-1273 as a primary series in Study mRNA-1273-P301 (COVE) or under the EUA. Enrollment of Part C 100 µg dose level will begin upon completion of enrollment of Part B of the study. Enrollment of the 50-µg dose arm will begin after completion of the 100 µg dose level arm in both Part C and Part D. The results of the mRNA-1273.617.2 vaccine will be compared to the immunogenicity induced after a 2-dose primary series of mRNA-1273 in the mRNA-1273-P301 (COVE) study, which demonstrated the efficacy of mRNA-1273. See the IB for further details.

**Part D**

Part D will evaluate the immunogenicity, safety, and reactogenicity of 2 dose levels (50 or 100 µg) of the mRNA-1273.213 vaccine when administered as a single booster dose to adults who have previously received 2 doses of mRNA-1273 as a primary series in Study mRNA-1273-P301 (COVE) or under the EUA. Enrollment of Part D 100µg dose level arm will begin upon completion of enrollment of Part C 100µg dose level arm of the study. Part D 50 µg dose arm enrollment will begin after completion of the 100-µg dose arm enrollment and may run in parallel with Part C 50 µg dose arm enrollment. The results of the mRNA-1273.213 vaccine will be compared to the immunogenicity induced after a 2-dose primary series of mRNA-1273 in the mRNA-1273-P301 (COVE) study, which demonstrated the efficacy of mRNA-1273. See the IB for further details.

**Part E**

Part E consisted of a group described in a site-specific protocol amendment and will not be discussed in this global protocol amendment.

**Part F**

Part F will consist of 2 cohorts: Cohort 1–- adults who have previously received 2 doses of mRNA-1273 as primary series and Cohort 2- adults who have previously received 2 doses of 100 µg mRNA-1273 as a primary series and a single booster dose of 50 µg mRNA-1273, in Study mRNA-1273-P301 (COVE) or under the EUA.

Cohort 1 will evaluate the immunogenicity, safety, and reactogenicity of 50 µg of the mRNA1273.529 vaccine candidate when administered as a single booster dose to adults who have previously received 2 doses of mRNA-1273 as a primary series.

Cohort 2 will evaluate the immunogenicity, safety and reactogenicity of 50 µg of the mRNA-1273.529 and of 50 µg of the mRNA-1273 vaccine candidate when administered as a second booster dose to adults who have previously received 2 doses of 100 µg mRNA-1273 as a primary series and a single booster dose of 50 µg mRNA-1273.

Enrollment of the mRNA-1273.529 Cohort 1 will run in parallel with the mRNA-1273.529 in Cohort 2. Enrollment of the 50-µg mRNA-1273 arm in Cohort 2 will begin upon completion of enrollment of the mRNA-1273.529 Cohort 2 arm and may run in parallel with the enrollment of the mRNA-1273.529 Cohort 1 arm. For Cohort 1, the results of the mRNA1273.529 vaccine candidate administered as a booster will be compared to the immunogenicity induced after a booster dose of mRNA1273 from the external historical comparator arm. (details will be provided in SAP). For Cohort 2, the results of the mRNA1273.529 vaccine candidate administered as the second booster dose will be compared to the immunogenicity induced after the second booster dose of mRNA-1273.

**Part G**

Enrollment of the mRNA-1273.214 50 µg second boost arm will begin upon completion of enrollment of the mRNA-1273 50 µg arm in Cohort 2 of Part F. Enrollment of the mRNA-1273.214 50 µg second boost arm may run in parallel with the enrollment of the mRNA-1273.529 Cohort 1 arm of Part F. Part G will evaluate the immunogenicity, safety, and reactogenicity of 50 µg of the mRNA1273.214 vaccine candidate when administered as a second booster dose to adults who have previously received 2 doses of 100 µg mRNA-1273 as a primary series and a single booster dose of 50 µg mRNA-1273. The results of the mRNA-1273.214 vaccine candidate administered as the second booster dose will be compared to the immunogenicity induced after the second booster dose of mRNA-1273 (Part F, Cohort 2, 50 µg mRNA-1273).

**Part H**

Part H will evaluate the immunogenicity, safety, and reactogenicity of 50 µg of the mRNA-1273.222 vaccine candidate when administered as a second booster dose to adults who have previously received 2 doses of 100 µg mRNA-1273 as a primary series and a single booster dose of 50 µg mRNA-1273. The immunogenicity of the mRNA-1273.222 candidate administered as the second booster dose will be compared to the immunogenicity of the second booster dose of mRNA-1273 (Part F, Cohort 2, 50 µg mRNA-1273).

**Part J**

Part J of the study will evaluate immunogenicity, safety, and reactogenicity of mRNA-1273.815 (50 µg) and mRNA-1273.231 (50 µg) administered as a booster to adults who previously received a primary series of mRNA vaccine, a first booster dose of a monovalent mRNA vaccine, and a second booster dose of a bivalent mRNA vaccine against SARS-CoV-2. Participants will be randomized in a 1:1 ratio to receive either mRNA-1273.815 (50 µg) or mRNA-1273.231 (50 µg).

Overall, study Parts A.1, B, C, and D will assess whether a single booster dose of the mRNA vaccines in each study part elicits antibody responses to ancestral SARS-CoV-2 and the variants similar to antibody responses to ancestral SARS-CoV-2 elicited by 2 doses of mRNA-1273 (100 µg) against ancestral SARS-CoV-2, using a historical control arm from the mRNA-1273-P301 (COVE) study. Study Part F Cohort 1 will assess whether a single booster dose of the mRNA-1273.529 as the first booster dose elicits a similar or superior antibody response to the B.1.1.529 compared to a single booster dose of mRNA-1273, using an external historical comparator; Study Part F Cohort 2 will assess whether a single booster dose of the mRNA-1273.529 as a second booster dose elicits a similar or superior antibody response to the B.1.1.529 compared to a single booster dose of mRNA-1273 as a second booster dose. Study Part G will assess whether a single booster dose of the mRNA-1273.214 as a second booster dose elicits a similar or superior antibody response to the B.1.1.529 compared to a single booster dose of mRNA-1273 as a second booster dose (Part F, Cohort 2, 50 µg mRNA-1273).

Study Part A.2 will evaluate the immunogenicity, safety, and reactogenicity of the mRNA-1273.214 vaccine candidate when administered as a second booster dose to participants who rolled over from Part A.1.

Study Part H will assess whether a single booster dose of the mRNA-1273.222 50 µg as a second booster dose elicits a superior antibody response against Omicron BA.4/5 compared to a single booster dose of mRNA-1273 as a second booster dose (Part F, Cohort 2, 50 µg mRNA-1273).

Part J will evaluate whether mRNA-1273.815 (50 µg) or mRNA-1273.231 (50 µg) administered as a booster will elicit antibody response against Omicron subvariants BA.4/BA.5, BQ.1.1, and XBB.1.5 in adults who previously received a primary series of mRNA vaccine, a first booster dose of a monovalent mRNA vaccine, and a second booster dose of a bivalent mRNA vaccine against SARS-CoV-2.

**Table 11: Study Arms**

| Study Part | Study Arm | Dose[1] | N |
|---|---|---|---|
| Part A.1 | mRNA-1273.211 | 50 µg | ~300 |
| | mRNA-1273.211 | 100 µg | ~584 |
| Part A.2[2] | mRNA-1273.214 | 50 µg | ~300 |
| Part B | mRNA-1273 | 100 µg | ~300 |
| Part C | mRNA-1273.617.2 | 50 µg | ~584 |
| | mRNA-1273.617.2 | 100 µg | ~584 |
| Part D | mRNA-1273.213 | 50 µg | ~584 |
| | mRNA-1273.213 | 100 µg | ~584 |
| Part F (Cohort 1) | mRNA-1273.529 | 50 µg | ~375 |
| Part F (Cohort 2) | mRNA-1273.529 | 50 µg | ~375 |
| | mRNA-1273 | 50 µg | ~375 |
| Part G | mRNA-1273.214 | 50 µg | ~375 |
| Part H | mRNA-1273.222 | 50 µg | ~500 |
| Part J (3rd booster)[3, 4] | mRNA-1273.815 | 50 µg | ~50 |
| | mRNA-1273.231 | 50 µg | ~50 |

1. Dose is total mRNA.
2. Participants rolled over from Part A.1 to Part A.2.
3. Participants may be rolled over from Part H.
4. Participants will be randomized in a 1:1 ratio to receive either mRNA-1273.815 or mRNA-1273.231.

The SoEs are provided in Table 17, Table 18, Table 19, and Table 20. Participants in Parts A.1, B, C, D, F, and G will have up to 7 visits; 6 visits if screening and dosing are performed on the same day. Study vaccine (mRNA-1273.211, mRNA-1273, mRNA-1273.617.2, mRNA-1273.213, mRNA-1273.529, or mRNA-1273.214) will be administered as a single dose on Day 1. Additional safety and/or immunogenicity study visits will occur on Days 15, 29, 91 (Part F and G only), 181, and 366 (EoS). Study visits will include scheduled safety phone calls at Day 8, every 2 weeks from Day 43 to Day 169 and from Day 209 to Day 349 to collect AEs, MAAEs, AESI, AEs leading to withdrawal, SAEs, pregnancies, and information about concomitant medications and receipt of non-study vaccinations.

Participants in Part A.1 who choose to continue in Part A.2 will have 6 additional visits; 5 if screening and dosing are performed on the same day. Study vaccine (mRNA-1273.214 [50 µg]) will be administered as a single dose on Day 1. Additional safety and/or immunogenicity study visits will occur on Days 15, 29, 91, and 181 (EoS). Study visits will include scheduled safety phone calls at Day 8, and every 2 weeks from Day 43 to Day 169 to collect AEs, MAAEs, AESI, AEs leading to withdrawal, SAEs, pregnancies, and information about concomitant medications and receipt of non-study vaccinations.

Participants in Part H will have up to 6 visits; 5 visits if screening and dosing are performed on the same day. Study vaccine (mRNA-1273.222 50 µg) will be administered as a single dose on Day 1. Additional safety and/or immunogenicity study visits will occur on Days 15, 29, 91, and 181 (EoS). Study visits will include scheduled safety phone calls at Day 8, and every 2 weeks from Day 43 to Day 169 to collect AEs, MAAEs, AESI, AEs leading to withdrawal, SAEs, pregnancies, and information about concomitant medications and receipt of non-study vaccinations.

Participants in Part J may include participants who have previously participated in Part H of this study. Participants in Part J will have up to 6 visits; 5 visits if screening and dosing are performed on the same day. Study vaccine (mRNA-1273.815 or mRNA-1273.231) will be administered as a single dose on Day 1. Additional safety and/or immunogenicity study visits will occur on Days 15, 29, 91, and 181 (EoS). Study visits will include scheduled safety phone calls at Day 8, and every 2 weeks from Day 43 to Day 169 to collect AEs, MAAEs, AESI, AEs leading to withdrawal, SAEs, pregnancies, and information about concomitant medications and receipt of non-study vaccinations. For participants who have previously participated in Part H of this study, the total number of visits completed in the study overall will be 10-12 depending on whether screening and dosing are performed on the same day.

At the dosing visit on Day 1, participants will be instructed how to document and report solicited ARs within a provided electronic diary (eDiary). Solicited ARs will be assessed for 7 days (the day of injection and the following 6 days), and unsolicited AEs will be assessed for 28 days after injection; SAEs, MAAEs, AEs leading to withdrawal, pregnancies and AESI will be assessed throughout the study. All participants will be tested for the presence of SARS-CoV-2 antibodies at baseline and at Day 29 (primary immunogenicity endpoint). Participants in Part J will also be tested for the presence of SARS-CoV-2 antibodies at Day 15 (primary immunogenicity endpoint). Additional blood draws will be collected on Day 91 (Parts A.2, F, G, H, and J only), Day 181, and Day 366 (Parts A.1, B, C, D, F, and G only). In addition, active surveillance for intercurrent or breakthrough SARS-CoV-2 infection will occur throughout the study and reported as AEs (confirmed symptomatic infections will be reported as MAAEs if not SAEs). Participants with signs and symptoms meeting the CDC case definition for COVID-19 (21 February 2021 or most recent [CDC 2020]) will be asked to contact the site and undergo prompt assessment which will include RT-PCR testing (of a respiratory sample) to assess symptomatic COVID-19. Participants with any clinical or radiographic evidence of pneumonia will also undergo RT-PCR testing. Suspected COVID-19 cases will also be tested using a multiplex assay to assess for non–SARS-CoV-2 causes of upper or lower respiratory tract infection (Table 12 and Table 13). Participants will have blood samples collected at scheduled study site visits during the study for immunogenicity assessments or other medical concerns according to the investigator's judgment.

Participants may experience AEs, to include symptoms of COVID-19, that necessitate an unscheduled visit. There may also be situations in which the investigator asks a participant to report for an unscheduled visit following the report of an AE. Additional examinations may be conducted at these visits as necessary to ensure the safety and well-being of participants during the study. Electronic case report forms should be completed for each unscheduled visit. In addition, participants may have blood samples collected at unscheduled visits for acute respiratory symptoms.

Peripheral blood mononuclear cells (PBMCs) may be collected for a subset of participants at selected sites at baseline (Day 1) and at Days 15, 29, and 91 to characterize the T-cell and B-cell responses against SARS-CoV-2 and variants.

Participants will be enrolled to receive 50 or 100 µg dose of mRNA-1273.211 (Part A.1), 100 µg mRNA-1273 (Part B), 50 µg or 100 µg mRNA-1273.617.2 (Part C), 50 µg or 100 µg mRNA-1273.213 (Part D), 50 µg mRNA-1273.529 or 50 µg mRNA-1273 (Part F), 50 µg mRNA-1273.214 (Part G and Part A.2), 50 µg mRNA-1273.222 (Part H), or 50 µg mRNA-1273.815 or 50 µg mRNA-1273.231 (Part J). The interim analysis will be conducted based on safety and immunogenicity data collected through Day 15 or Day 29. The interim analysis may be conducted either after all participants in Parts A.1, A.2, B, C, D, F, G, H, or J have completed their Day 15 or Day 29 visit assessments and/or subsequent timepoint visits (eg, Day 91 for Parts F and G, and H) or combined after the last participant in any of the study parts (Parts A.1, A.2, B, C, D, F, G, H or J), dose arm or pre-specified subset of dose arm has completed their Day 15 or Day 29 visit assessments. The final study analysis after 12 months of follow-up will be completed for all participants in Parts A.1, B, C, D, F, and G. The final study analysis after 6 months of follow-up will be completed for all participants in Parts A.2, H, and J.

#### 3.2. Scientific Rationale for Study Design

This study is designed as an open-label study.

With SARS-CoV-2 expected to be circulating in the general population during the study, all participants will provide pre-injection and post-injection blood samples for analysis of antibodies to non-vaccine antigens through 12 months after study injection, or 6 months after study injection for Parts A.2, H, and J. In addition, participants will have NP swab samples collected before vaccination on Day 1, on Day 29, and also on Day 91 (Parts A.2, F, G, H, and J only), Day 181, and Day 366 (EoS) (Parts A.1, B, C, D, F, and G only). Furthermore, in case of any signs or symptoms or MAAEs suggesting SARS-CoV-2 infection in a participant, an additional NP swab sample and blood sample will be collected to confirm the diagnosis of SARS-CoV-2 via serology and RT-PCR. Additionally, clinical information will be carefully collected to evaluate the severity of the clinical case.

Since it is possible that participants are naturally exposed to SARS-CoV-2 through community exposure, the NP swab samples collected before study injection and the serologic assays performed for antibody responses to non-vaccine antigen(s), may help to discriminate between natural infection and vaccine-induced antibody responses, should such discrimination be needed.

#### 3.3. Justification for Dose, Control Product, and Choice of Study Population

The dose of the ancestral SARS-CoV-2 vaccine (mRNA-1273) was clinically evaluated at dose levels of 25, 50, 100, and 250 µg in a 2-dose series in a Phase 1 dose-ranging study, with the 100-µg dose level selected for the pivotal Phase 3 trial. Therefore, the dose of 100 µg will be used as the booster dose in Part B of this study (monovalent mRNA1273 booster). In addition, the dose of 50 µg and100 µg will be used for the monovalent mRNA1273.617.2 booster (Part C). In Part D, the bivalent mRNA-1273-213 50 µg and 100µg dose will be used as a booster. In Part F, the monovalent mRNA-1273.529 50 µg will be used as a first booster dose after mRNA-1273 primary series at least 6 months post-second dose, or as a second booster dose at least 3 months after mRNA-1273 50 µg booster. In Part G and Part A.2, the multivalent mRNA-1273.214 booster candidate will be administered at the 50-µg dose level as a second booster dose. In Part H the multivalent mRNA-1273.222 booster candidate will be administered at the 50-µg dose level as a second booster dose. In Part J, the multivalent booster candidates mRNA-1273.815 and mRNA-1273.231 will be administered at a dose of 50 µg.

In the mRNA-1273-P201 study, mRNA-1273 administered as a single booster dose of 50 µg was well-tolerated and demonstrated significant boosting of neutralizing antibody responses to SARS-CoV-2 in a pseudovirus neutralization assay (Wu et al 2021b).

This study will screen and enroll healthy adults, 18 years of age and above, who have previously received 2 doses of mRNA-1273 in the mRNA-1273-P301 (COVE) study or under EUA (Parts A.1, B, C, and D); or who have previously received 2 doses of mRNA-1273 and a booster dose of mRNA-1273 in the mRNA-1273-P301 (COVE) study or under EUA (Parts F, G, and H); or who have previously received 2 doses of mRNA-1273 in the mRNA-1273-P301 (COVE) study or under EUA and have received a booster dose of mRNA-1273.211 50 µg in Part A.1 of this study (Part A.2); or who previously received a primary series of mRNA vaccine, a first booster dose of a monovalent mRNA vaccine, and a second booster dose of a bivalent mRNA vaccine against SARS-CoV-2 (Part J).

#### 3.4. End of Study Definition

A participant is considered to have completed the study if he or she has completed all phases of the study including the last scheduled procedure as shown in the SoE (Table 17) for Parts A.1, B, C, D, F and G), unless they choose to participate in Part A.2. A participant who chooses to enroll in Part A.2 is considered to have completed the study if he or she has completed all phases of the study including the last scheduled procedure as shown in in the SoE (Table 18). A participant in Part H is considered to have completed the study if he or she has completed all phases of the study including the last scheduled procedure as shown in the SoE (Table 19), unless they choose to participate in Part J. A participant who chooses to enroll in Part J is considered to have completed the study if he or she has completed all phases of the study including the last scheduled procedure as shown in in the SoE (Table 20).

The EoS is defined as completion of the last visit of the last participant in the study or last scheduled procedure, as shown in the SoE (Table 17, Table 18, Table 19, and Table 20, whichever is later), for the last participant in the study.

### 4. STUDY POPULATION

**Part A.1:**

Approximately 300 participants will receive a single booster dose of mRNA-1273.211 50 µg, to achieve 270 evaluable participants in the 50-µg dose study arm. Approximately 584 participants will receive a single booster dose of mRNA-1273.211 100 µg, to achieve 526 evaluable participants in the 100 µg dose study arm.

**Part A.2:**

Approximately 300 participants will receive a second booster dose of mRNA-1273.214 50 µg.

**Part B:**

Approximately 300 participants will receive a single booster dose of mRNA-1273 100 µg, to achieve 270 evaluable participants in Part B of the study.

**Part C:**

Approximately 584 participants will receive a single booster dose of mRNA-1273.617.2 50 µg, to achieve 526 evaluable participants in the 50-µg dose study arm. Approximately 584 participants will receive a single booster dose of mRNA-1273.617.2 100 µg, to achieve 526 evaluable participants in the 100-µg dose study arm.

**Part D:**

Approximately 584 participants will receive a single booster dose of mRNA-213 50 µg, to achieve 526 evaluable participants in the 50-µg dose study arm. Approximately 584 participants will receive a single booster dose of mRNA-1273.213 100 µg, to achieve 526 evaluable participants in the 100-µg dose study arm.

**Part F:**

In Cohort 1, approximately 375 participants will receive a single booster dose of mRNA-1273.529 50 µg after receiving a primary series of mRNA-1273 vaccine to achieve 300 evaluable participants in the mRNA-1273.529 50 µg dose study arm.

In Cohort 2, approximately 375 participants will receive a single booster dose of mRNA-1273.529 50 µg after receiving a primary series of mRNA-1273 vaccine and a single booster dose of mRNA-1273 50 µg to achieve 300 evaluable participants in the mRNA-1273.529 50 µg study arm. Approximately 375 participants will receive a single booster dose of mRNA-1273 50 µg after receiving a primary series of mRNA-1273 vaccine and a single booster dose of mRNA1273 50 µg to achieve 300 evaluable participants in the mRNA-1273 50 µg study arm.

Prospective approval of protocol deviations to recruitment and enrollment criteria, also known as protocol waivers or exemptions, is not permitted.

**Part G:**

Approximately 375 participants will receive a single booster dose of mRNA-1273.214 50 µg after receiving a primary series of mRNA-1273 vaccine and a single booster dose of mRNA-1273 50 µg, to achieve 300 evaluable participants.

**Part H:**

Approximately 500 participants will receive a second booster dose of mRNA1273.222 50 µg to achieve 300 evaluable participants.

**Part J:**

Approximately 100 participants will be randomized in a 1:1 ratio to receive a single booster dose of mRNA-1273.815 (50 µg) or mRNA-1273.231 (50 µg).

#### 4.1. Inclusion Criteria

Each participant must meet all of the following criteria to be enrolled in this study:

1. Male or female, at least 18 years of age at the time of consent (Screening Visit).
2. Investigator's assessment that participant understands and is willing and physically able to comply with protocol-mandated follow-up, including all procedures.
3. Participant has provided written informed consent for participation in this study, including all evaluations and procedures as specified in this protocol.
4. Female participants of nonchildbearing potential may be enrolled in the study. Nonchildbearing potential is defined as surgically sterile (history of bilateral tubal ligation, bilateral oophorectomy, hysterectomy) or postmenopausal (defined as amenorrhea for ≥ 12 consecutive months prior to screening [Day 0] without an alternative medical cause). A follicle-stimulating hormone (FSH) level may be measured at the discretion of the investigator to confirm postmenopausal status.
5. Female participants of childbearing potential may be enrolled in the study if the participant fulfills all of the following criteria:
   - Has a negative pregnancy test on the day of vaccination (Day 1).
   - Has practiced adequate contraception (Section 10.3) or has abstained from all activities that could result in pregnancy for at least 28 days prior to Day 1.
   - Has agreed to continue adequate contraception through 3 months following vaccination.
   - Is not currently breastfeeding.

   Adequate female contraception is defined as consistent and correct use of a United States FDA-approved contraceptive method in accordance with the product label (Section 10.3).
6. Participant must have been either previously enrolled in the mRNA-1273-P301 (COVE) study, must have received 2 doses of mRNA-1273 in that study, with his/her second dose at least 6 months prior to enrollment in mRNA-1273-P205, and must be currently enrolled and compliant in that study (ie, has not withdrawn or discontinued early); or participant must have received 2 doses of mRNA-1273 under the EUA with their second dose at least 6 months prior to enrollment in mRNA-1273-P205; or have received a 2 dose primary series of mRNA-1273 followed by a 50 µg booster dose of mRNA-1273 in the mRNA-1273-P301 (COVE) study or under EUA at least 3 months prior to enrollment in mRNA-1273-P205; and able to provide proof of vaccination status at the time of

screening (Day 1); or for enrollment in Part A.2, participant must be currently enrolled and compliant in Part A.1 of the mRNA-1273-P205 study and must have received their first booster dose of mRNA-1273.211 50 µg; or for enrollment in Part J, participant must meet at least one of the following criteria:

- Completed enrollment in Part H of the mRNA-1273-P205 study; or
- Received a 2-dose primary series of mRNA-1273 (100 µg) followed by a 50 µg booster dose of mRNA-1273 in the mRNA-1273-P301 (COVE) study or under EUA, followed by a 50 µg booster dose of mRNA-1273.222 under EUA at least 3 months prior to enrollment in Part J of mRNA-1273-P205; or
- Previously received a 2-dose primary series of mRNA vaccine against SARS-CoV-2 followed by a booster dose of a monovalent mRNA vaccine, followed by a second booster dose of a bivalent mRNA vaccine

Participants in Part J must also provide proof of vaccination status at the time of screening (Day 0 or Day 1).

#### 4.2. Exclusion Criteria

Participants meeting any of the following criteria at the Screening Visit, unless noted otherwise, will be excluded from the study:

1. Had significant exposure to someone with SARS-CoV-2 infection or coronavirus disease 2019 (COVID-19) in the past 14 days, as defined by the CDC as a close contact of someone who has had COVID-19.
2. Has known history of SARS-CoV-2 infection within 3 months prior to enrollment.
3. Is acutely ill or febrile (temperature ≥ 38.0°C/[100.4°F]) less than 72 hours prior to or at the Screening Visit or Day 1. Participants meeting this criterion may be rescheduled and will retain their initially assigned participant number.
4. Currently has symptomatic acute or unstable chronic disease requiring medical or surgical care, to include significant change in therapy or hospitalization for worsening disease, at the discretion of the investigator.
5. Has a medical, psychiatric, or occupational condition that may pose additional risk as a result of participation, or that could interfere with safety assessments or interpretation of results according to the investigator's judgment.
6. Has a current or previous diagnosis of immunocompromising condition to include human immunodeficiency virus, immune-mediated disease requiring immunosuppressive treatment, or other immunosuppressive condition.
7. Has received systemic immunosuppressants or immune-modifying drugs for > 14 days in total within 6 months prior to screening (for corticosteroids ≥ 10 mg/day of prednisone equivalent) or is anticipating the need for immunosuppressive treatment at any time during participation in the study.
8. Has known or suspected allergy or history of anaphylaxis, urticaria, or other significant AR to the vaccine or its excipients.

9. Has a documented history of myocarditis or pericarditis within 2 months prior to Screening Visit (Day 0).
10. Coagulopathy or bleeding disorder considered a contraindication to intramuscular (IM) injection or phlebotomy.
11. Has received or plans to receive any licensed vaccine ≤ 28 days prior to the injection (Day 1) or a licensed vaccine within 28 days before or after the study injection, with the exception of influenza vaccines, which may be given 14 days before or after receipt of a study vaccine.
12. Has received systemic immunoglobulins or blood products within 3 months prior to the Screening Visit (Day 0) or plans for receipt during the study.
13. Has donated ≥ 450 mL of blood products within 28 days prior to the Screening Visit or plans to donate blood products during the study.
14. Plans to participate in an interventional clinical trial of an investigational vaccine or drug while participating in this study.
15. Is an immediate family member or household member of study personnel, study site staff, or Sponsor personnel.
16. Is currently experiencing an SAE in Study mRNA-1273-P301 (COVE) at the time of screening for this study.

#### 4.3. Lifestyle Restrictions

Participants must not eat or drink anything hot or cold within 10 minutes before oral temperature is taken.

#### 4.4. Screen Failures

Screen failures are defined as participants who consent to participate in the clinical study but are not subsequently assigned to treatment. A minimum set of screen failure information is required to ensure transparent reporting of screen failures to meet the Consolidated Standards of Reporting Trials publishing requirements and to respond to queries from regulatory authorities. Minimum information includes date of informed consent, demography, reason(s) for screen failure, eligibility criteria, and information on any SAE that may have occurred from the time informed consent was obtained to the time of withdrawal.

### 5. STUDY TREATMENT

#### 5.1. Investigational Products Administered

The term “investigational product (IP)” refers to mRNA-1273, mRNA-1273.211, mRNA-1273.617.2, mRNA-1273.213, mRNA-1273.529, mRNA-1273.214, mRNA-1273.222, mRNA-1273.815, or mRNA-1273.231 vaccine administered in this study.

**Part A.1 (mRNA-1273.211)**

mRNA-1273.211 is a multivalent product that contains 2 mRNAs: CX-024414 encoding for the S-2P of Wuhan-Hu-1 and CX-027367 encoding for the S-2P of B.1.351, in a 1:1 ratio. mRNA-1273.211 consists of the mRNA formulated in a mixture of 4 lipids common to the Sponsor’s mRNA vaccine platform: SM-102, cholesterol, 1,2-distearoyl-sn-glycero-3-phosphocholine (DSPC), and 1-monomethoxypolyethyleneglycol-2,3-dimyristylglycerol with polyethylene glycol of average molecular weight 2000 (PEG-2000-DMG).

mRNA-1273.211 injection will be provided as a sterile liquid for injection, white to off-white dispersion in appearance, at a concentration of 0.2 mg/mL (0.8 mL fill volume) in 20 mM Tris buffer containing 87 mg/mL sucrose and 4.3 mM sodium acetate at pH 7.5. mRNA-1273.211 will be administered at a 50 and 100 µg dose levels.

**Part A.2 (mRNA-1273.214)**

mRNA-1273.214 contains CX-024414, the mRNA that encodes for the prefusion stabilized spike glycoprotein (S-2P) of the Wuhan-Hu-1 isolate of SARS-CoV-2 and CX-031302, the mRNA that encodes for the S-2P of the SARS-CoV-2 B.1.1.529 variant. mRNA-1273.214 consists of each mRNA formulated in a mixture of 4 lipids common to the Sponsor’s mRNA vaccine platform: SM-102, cholesterol, DSPC, and PEG2000-DMG.

The formulated mRNAs are mixed in a 1:1 ratio. mRNA-1273.214 injection will be provided as a sterile liquid for injection, white to off-white dispersion in appearance, at a concentration of 0.1 mg/ML in 20 mM Tris buffer containing 87 mg/mL sucrose and 2.1 mM acetate at pH 7.5.

mRNA-1273 will be administered at a 50-µg dose level. Doses will be administered by IM injection according to the procedures specified in the mRNA-1273-P205 Pharmacy Manual. Preferably, the vaccine should be administered into the nondominant arm.

**Part B (mRNA-1273)**

mRNA-1273 contains mRNA CX-024414 encoding for the S-2P of Wuhan-Hu-1. mRNA-1273 consists of the mRNA formulated in a mixture of 4 lipids common to the Sponsor’s mRNA vaccine platform: SM-102, cholesterol, DSPC, and PEG-2000-DMG.

mRNA-1273 injection will be provided as a sterile liquid for injection, white to off-white dispersion in appearance, at a concentration of 0.2 mg/mL (0.8 mL fill volume) in 20 mM Tris buffer containing 87 mg/mL sucrose and 4.3 mM sodium acetate at pH 7.5. mRNA-1273 will be administered at a 100-µg dose level.

**Part C (mRNA-1273.617.2)**

mRNA-1273.617.2 contains mRNA CX-029444 encoding for the S-2P of B.1.617.2. mRNA-1273.617.2 consists of the mRNA formulated in a mixture of 4 lipids common to the Sponsor's mRNA vaccine platform: SM-102, cholesterol, DSPC, and PEG-2000-DMG.

mRNA-1273.617.2 injection will be provided as a sterile liquid for injection, white to off-white dispersion in appearance, at a concentration of 0.2 mg/mL (0.8 mL fill volume) in 20 mM Tris buffer containing 87 mg/mL sucrose and 4.3 mM sodium acetate at pH 7.5. mRNA-1273.617.2 will be administered at 50 µg and 100 µg dose level.

**Part D (mRNA-1273.213)**

mRNA-1273.213 is a multivalent product that contains 2 mRNAs: CX-029444 encoding for the S-2P of B.1.617.2 and CX-027367 encoding for the S 2P of B.1.351, in a 1:1 ratio. mRNA-1273.213 consists of the mRNA formulated in a mixture of 4 lipids common to the Sponsor's mRNA vaccine platform: SM-102, cholesterol, DSPC and PEG 2000 DMG.

mRNA-1273.213 injection will be provided as a sterile liquid for injection, white to off-white dispersion in appearance, at a concentration of 0.2 mg/mL in 20 mM Tris buffer containing 87 mg/mL sucrose and 4.3 mM sodium acetate at pH 7.5. mRNA-1273.213 will be administered at 50 µg and 100 µg dose level.

**Part F (mRNA-1273.529 and mRNA-1273)**

mRNA-1273.529 contains mRNA CX-031302 encoding for the S-2P of B.1.1.529. mRNA-1273.529 consists of the mRNA formulated in a mixture of 4 lipids common to the Sponsor's mRNA vaccine platform: SM-102, cholesterol, DSPC and PEG 2000 DMG.

mRNA-1273.529 injection will be provided as a sterile liquid for injection, white to off-white dispersion in appearance, at a concentration of 0.2 mg/mL in 20 mM Tris buffer containing 87 mg/mL sucrose and 4.3 mM sodium acetate at pH 7.5.

mRNA-1273.529 will be administered at 50 µg dose level. Doses will be administered by IM injection according to the procedures specified in the mRNA-1273 P205 Pharmacy Manual. Preferably, the vaccine should be administered into the nondominant arm.

**Part G (mRNA-1273.214)**

mRNA-1273.214 contains CX-024414, the mRNA that encodes for the prefusion stabilized spike glycoprotein (S-2P) of the Wuhan-Hu-1 isolate of SARS-CoV-2 and CX-031302, the mRNA that encodes for the S-2P of the SARS-CoV-2 B.1.1.529 variant. mRNA-1273.214 consists of each mRNA formulated in a mixture of 4 lipids common to the Sponsor's mRNA vaccine platform: SM-102, cholesterol, DSPC, and PEG2000-DMG.

The formulated mRNAs are mixed in a 1:1 ratio. mRNA-1273.214 injection will be provided as a sterile liquid for injection, white to off-white dispersion in appearance, at a concentration of 0.1 mg/ML in 20 mM Tris buffer containing 87 mg/mL sucrose and 2.1 mM acetate at pH 7.5.

mRNA-1273.214 will be administered at a 50-µg dose level. Doses will be administered by IM injection according to the procedures specified in the mRNA-1273 P205 Pharmacy Manual. Preferably, the vaccine should be administered into the nondominant arm.

**Part H (mRNA-1273.222)**

mRNA-1273.222 contains CX-024414, the mRNA that encodes for the prefusion stabilized spike glycoprotein (S-2P) of the Wuhan-Hu-1 isolate of SARS-CoV-2 and CX-034476, the mRNA that encodes for the S-2P of the SARS-CoV-2 B.1.1.529 subvariants BA.4/BA.5. mRNA-1273.222 consists of each mRNA formulated in a mixture of 4 lipids common to the Sponsor's mRNA vaccine platform: SM-102, cholesterol, DSPC, and PEG2000-DMG.

The formulated mRNA are mixed in a 1:1 ratio. mRNA-1273.222 injection will be provided as a sterile liquid for injection, white to off-white dispersion in appearance, at a concentration of 0.1 mg/mL in 20 mM Tris buffer containing 87 mg/mL sucrose and 2.1 mM acetate at pH 7.5.

mRNA-1273.222 will be administered at a 50-µg dose level. Doses will be administered by IM injection according to the procedures specified in the mRNA-1273-P205 Pharmacy Manual. Preferably, the vaccine should be administered into the nondominant arm.

**Part J (mRNA-1273.815 and mRNA-1273.231)**

mRNA-1273.815 contains CX-038839, the mRNA that encodes for the S-2P of the SARS-CoV-2 subvariants XBB.1.5/XBB.1.9.1 mRNA-1273.231 contains CX-034476, the mRNA that encodes for the S-2P of the SARS-CoV-2 B.1.1.529 subvariants BA.4/BA.5, and CX-038839, the mRNA that encodes for the S-2P of the SARS-CoV-2 subvariants XBB.1.5/XBB.1.9.1. The formulated mRNA in mRNA-1273.231 are mixed in a 1:1 ratio. Both mRNA-1273.815 and mRNA-1273.231 consist of mRNA formulated in a mixture of 4 lipids common to the Sponsor's mRNA vaccine platform: SM-102, cholesterol, DSPC, and PEG2000-DMG.

mRNA-1273.231 and mRNA-1273.815 injections will be provided as a sterile liquid for injection, white to off-white dispersion in appearance, at a concentration of 0.1 mg/mL in 20 mM Tris buffer containing 87 mg/mL sucrose and 2.1 mM acetate at pH 7.5.

mRNA-1273.815 and mRNA-1273.231 will be administered at a 50 µg dose level. Doses will be administered by IM injection according to the procedures specified in the mRNA-1273-P205 Pharmacy Manual.

#### 5.2. Randomization and Blinding

This is an open-label study, no blinding will be performed. Randomization will be performed for participants in Part J.

#### 5.3. Preparation/Handling/Storage/Accountability

##### 5.3.1. Preparation of Study Vaccine

**Part A.1**

The mRNA-1273.211 vaccine candidate will have a fill volume of 0.8 mL and contain mRNA-1273.211 at the dose of 50 and 100 µg (dose volume 0.5 mL). Vaccine preparation instructions are detailed in the mRNA-1273-P205 Pharmacy Manual.

**Part A.2**

The mRNA-1273.214 vaccine will have a fill volume of 0.8 mL and contains mRNA-1273.214 at the dose of 50 µg (dose volume 0.5 mL). Vaccine preparation instructions are detailed in the mRNA-1273-P205 Pharmacy Manual.

**Part B**

The mRNA-1273 vaccine will have a fill volume of 6.3 mL and contains mRNA-1273 at the dose of 100 µg (dose volume 0.5 mL). Vaccine preparation instructions are detailed in the mRNA-1273-P205 Pharmacy Manual.

**Part C**

The mRNA-1273.617.2 vaccine will have a fill volume of 0.8 mL and contains mRNA-1273.617.2 at the dose of 50 µg (dose volume 0.25 mL) and 100 µg (dose volume 0.5 mL). Vaccine preparation instructions are detailed in the mRNA-1273-P205 Pharmacy Manual.

**Part D**

The mRNA-1273.213 vaccine will have a fill volume of 0.8 mL and contains mRNA-1273.213 at the dose of 50 µg (dose volume 0.25 mL) and 100 µg (dose volume 0.5 mL). Vaccine preparation instructions are detailed in the mRNA-1273-P205 Pharmacy Manual.

**Part F**

The mRNA-1273.529 vaccine will have a fill volume of 0.8 mL and contains mRNA-1273.529 at the dose of 50 µg (dose volume 0.25 mL). The mRNA-1273 vaccine will have a fill volume of 6.3 mL and contains mRNA-1273 at the dose of 50 ug (dose volume 0.25 mL). Vaccine preparation instructions are detailed in the mRNA-1273-P205 Pharmacy Manual.

**Part G**

The mRNA-1273.214 vaccine will have a fill volume of 0.8 mL and contains mRNA-1273.214 at the dose of 50 µg (dose volume 0.5 mL). Vaccine preparation instructions are detailed in the mRNA-1273-P205 Pharmacy Manual.

**Part H**

The mRNA-1273.222 vaccine will have a fill volume of 0.8 mL and contains mRNA-1273.222 at the dose of 50 µg (dose volume 0.5 mL). Vaccine preparation instructions are detailed in the mRNA-1273-P205 Pharmacy Manual.

**Part J**

The mRNA-1273.815 vaccine will have a fill volume of 0.8 mL and contains mRNA-1273.815 at the dose of 50 µg (dose volume 0.5 mL). Vaccine preparation instructions are detailed in the mRNA-1273-P205 Pharmacy Manual.

The mRNA-1273.231 vaccine will have a fill volume of 0.8 mL and contains mRNA-1273.231 at the dose of 50 µg (dose volume 0.5 mL). Vaccine preparation instructions are detailed in the mRNA-1273-P205 Pharmacy Manual.

##### 5.3.2. Study Vaccine Administration

mRNA-1273.211, mRNA-1273, mRNA-1273.617.2, mRNA-1273.213, mRNA-1273.529, mRNA-1273.214, mRNA-1273.222, mRNA-1273.815, or mRNA-1273.231 will be administered as an IM injection into the deltoid muscle on Day 1. Preferably, vaccine should be administered into the nondominant arm.

On Day 1, participants will be monitored for a minimum of 30 minutes after vaccination. Assessments will include vital sign measurements and monitoring for local or systemic ARs as shown in the SoE (Table 17, Table 18, Table 19, and Table 20).

The study site will be appropriately staffed with individuals with basic cardiopulmonary resuscitation training/certification. Either onsite resuscitation equipment and personnel or appropriate protocols for the rapid transport of a participant to a resuscitation area or facility are required.

##### 5.3.3. Study Vaccine Delivery and Receipt

The Sponsor or designee is responsible for the following:

- Supplying the IP
- Confirming the appropriate labeling of the IP, so that it complies with the legal requirements of the United States

The investigator is responsible for acknowledging the receipt of the IP by a designated staff member at the site, which includes the following:

- Confirming that the IP was received in good condition
- Confirming that the temperature during shipment from the Sponsor to the investigator's designated storage location was appropriate
- Confirming that the Sponsor has authorized the IP for use
- Ensuring the appropriate dose of IP is properly prepared using aseptic technique

Further description of the IP and instructions for the receipt, storage, preparation, administration, accountability, and destruction of IP are described in the mRNA-1273-P205 Pharmacy Manual.

##### 5.3.4. Study Vaccine Packaging and Labeling

**Part A.1**

The Sponsor will provide the investigator (via the study site pharmacy) with adequate quantities of mRNA-1273.211. Sterile mRNA-1273.211 is packaged in 2R glass vials with a 0.8-mL fill volume. The IP will have all required labeling per regulations and will be supplied to the pharmacy in an unblinded manner.

The Sponsor or Sponsor's designee will supply the 0.9% sodium chloride injection for use as a diluent to mRNA-1273.211. The 0.9% sodium chloride bears a commercial label and does not contain study-specific identification.

The IP will be packaged and labeled in accordance with the standard operating procedures of the Sponsor or of its designee, Code of Federal Regulations (CFR) Title 21 Good Manufacturing Practice guidelines, International Council for Harmonisation (ICH) GCP Guideline, guidelines for Quality System Regulations, and applicable regulations.

**Part A.2**

The Sponsor will provide the investigator (via the study site pharmacy) with adequate quantities of mRNA-1273.214. Sterile mRNA-1273.214 is packaged in 2R glass vials with a 0.8-mL fill volume. The IP will have all required labeling per regulations and will be supplied to the pharmacy in an unblinded manner.

The IP will be packaged and labeled in accordance with the standard operating procedures of the Sponsor or of its designee, CFR Title 21 Good Manufacturing Practice guidelines, ICH GCP Guideline, guidelines for Quality System Regulations, and applicable regulations.

**Part B**

The Sponsor will provide the investigator (via the study site pharmacy) with adequate quantities of mRNA-1273. Sterile mRNA-1273 is packaged in 10R glass vials with a 6.3-mL fill volume. The IP will have all required labeling per regulations and will be supplied to the pharmacy in an unblinded manner.

The IP will be packaged and labeled in accordance with the standard operating procedures of the Sponsor or of its designee, CFR Title 21 Good Manufacturing Practice guidelines, ICH GCP Guideline, guidelines for Quality System Regulations, and applicable regulations.

**Part C**

The Sponsor will provide the investigator (via the study site pharmacy) with adequate quantities of mRNA-1273.617.2. Sterile mRNA-1273.617.2 is packaged in 2R glass vials with a 0.8-mL fill volume. The IP will have all required labeling per regulations and will be supplied to the pharmacy in an unblinded manner.

The IP will be packaged and labeled in accordance with the standard operating procedures of the Sponsor or of its designee, CFR Title 21 Good Manufacturing Practice guidelines, ICH GCP Guideline, guidelines for Quality System Regulations, and applicable regulations.

**Part D**

The Sponsor will provide the investigator (via the study site pharmacy) with adequate quantities of mRNA-1273.213. Sterile mRNA-1273.213 is packaged in 2R glass vials with a 0.8-mL fill volume. The IP will have all required labeling per regulations and will be supplied to the pharmacy in an unblinded manner.

The IP will be packaged and labeled in accordance with the standard operating procedures of the Sponsor or of its designee, CFR Title 21 Good Manufacturing Practice guidelines, ICH GCP Guideline, guidelines for Quality System Regulations, and applicable regulations.

**Part F**

The Sponsor will provide the investigator (via the study site pharmacy) with adequate quantities of mRNA-1273.529 and mRNA-1273. Sterile mRNA-1273.529 is packaged in 2R glass vials with a 0.8-mL fill volume. Sterile mRNA-1273 is packaged in 10R glass vials with a 6.3-mL fill volume. The IP will have all required labeling per regulations and will be supplied to the pharmacy in an unblinded manner.

The IP will be packaged and labeled in accordance with the standard operating procedures of the Sponsor or of its designee, CFR Title 21 Good Manufacturing Practice guidelines, ICH GCP Guideline, guidelines for Quality System Regulations, and applicable regulations.

**Part G**

The Sponsor will provide the investigator (via the study site pharmacy) with adequate quantities of mRNA-1273.214. Sterile mRNA-1273.214 is packaged in 2R glass vials with a 0.8-mL fill volume. The IP will have all required labeling per regulations and will be supplied to the pharmacy in an unblinded manner.

The IP will be packaged and labeled in accordance with the standard operating procedures of the Sponsor or of its designee, CFR Title 21 Good Manufacturing Practice guidelines, ICH GCP Guideline, guidelines for Quality System Regulations, and applicable regulations.

**Part H**

The Sponsor will provide the investigator (via the study site pharmacy) with adequate quantities of mRNA-1273.222. Sterile mRNA-1273.222 is packaged in 2R glass vials with a 0.8-mL fill volume. The IP will have all required labeling per regulations and will be supplied to the pharmacy in an unblinded manner.

The IP will be packaged and labeled in accordance with the standard operating procedures of the Sponsor or of its designee, CFR Title 21 Good Manufacturing Practice guidelines, ICH GCP Guideline, guidelines for Quality System Regulations, and applicable regulations.

**Part J**

The Sponsor will provide the investigator (via the study site pharmacy) with adequate quantities of mRNA-1273.815 and mRNA-1273.231. Sterile mRNA-1273.815 and mRNA-1273.231 will be packaged in 2R glass vials with a 0.8-mL fill volume. The IP will have all required labeling per regulations and will be supplied to the pharmacy in an unblinded manner.

The IP will be packaged and labeled in accordance with the standard operating procedures of the Sponsor or of its designee, CFR Title 21 Good Manufacturing Practice guidelines, ICH GCP Guideline, guidelines for Quality System Regulations, and applicable regulations.

##### 5.3.5. Study Vaccine Storage

mRNA-1273.211, mRNA-1273, mRNA-1273.617.2, mRNA-1273.213, mRNA-1273.529, mRNA-1273.214, and mRNA-1273.222, mRNA-1273.815, and mRNA-1273.231 must be stored at -60°C to -90°C (-76°F to -130°F), and mRNA-1273 must be stored at -25°C to -15°C (-13°F to 5°F). All study vaccines must be stored in a secure area with limited access and protected from moisture and light until it is prepared for administration (Section 5.3.1). The freezer should have automated temperature recording and a 24-hour alert system in place that allows for rapid response in case of or refrigerator malfunction. There must be an available backup freezer. The freezer must be connected to a backup generator. In addition, IP accountability study staff are required to keep a temperature log to establish a record of compliance with these storage conditions. The site is responsible for reporting any IP that was not temperature-controlled during shipment or storage. Such IP will be retained for inspection by the monitor and disposed of according to approved methods.

The 0.9% sodium chloride injection (USP) should be stored at 20ºC to 25ºC (68ºF to 77ºF) in a restricted access area.

##### 5.3.6. Study Vaccine Accountability

It is the investigator's responsibility that the IP accountability study staff maintain accurate records in an IP accountability log of receipt of all IP, site IP inventory, IP dispensing, IP injections, and return to the Sponsor or alternative disposition of used and unused IP vials.

A site monitor will review the inventory and accountability log during site visits and at the completion of the study. Additional details are found in the mRNA-1273-P205 Pharmacy Manual.

##### 5.3.7. Study Vaccine Handling and Disposal

A site monitor will reconcile the IP inventory during the conduct and at the end of the study for compliance. Once fully reconciled at the site at the end of the study, the IP should be destroyed on site, if site procedures allow, or returned to a destruction depot per instruction of the Sponsor. Additional details are found in the mRNA-1273-P205 Pharmacy Manual.

##### 5.3.8. Unblinding

This is an open-label study.

#### 5.4. Study Intervention Compliance

All doses of IP will be administered at the study site under direct observation of medically qualified study staff and appropriately recorded (date and time) in the eCRF. Qualified staff will confirm that the participant has received the entire dose of IP. If a participant does not receive IP or does not receive all of the planned dose, the reason for the missed dose will be recorded. Data will be reconciled with site accountability records to assess compliance.

The study site staff are responsible for ensuring that participants comply with the allowed study visit windows. If a participant misses a visit, every effort should be made to contact the participant and complete a visit within the defined visit window specified in the SoE (Table 17, Table 18, Table 19, and Table 20). If a participant does not complete a visit within the time window, that visit will be classified as a missed visit and the participant will continue with subsequent scheduled study visits. All safety requirements of the missed visit will be captured and included in the subsequent visit.

#### 5.5. Prior and Concomitant Medications

##### 5.5.1. Prior Medications and Therapies

Information about prior medications (including any prescription or over-the-counter medications, vaccines, or blood products) taken by the participant within the 28 days before providing informed consent (or as designated in the inclusion/exclusion requirements) will be recorded in the participant's eCRF.

##### 5.5.2. Concomitant Medications and Therapies

At study site, study staff must question the participant regarding any medications taken and non-study vaccinations received by the participant and record the following information in the eCRF:

- All non-study vaccinations administered within the period starting 28 days before the study injection.
- Seasonal influenza vaccine administered for the current influenza season (typically October through April in the Northern Hemisphere).
- All concomitant medications and non-study vaccinations taken through 28 days after vaccination. Antipyretics and analgesics taken prophylactically (ie, taken in the absence of any symptoms in anticipation of an injection reaction) will be recorded as such.
- Any concomitant medications used to prevent or treat COVID-19 or its symptoms.
- Any concomitant medications relevant to or for the treatment of an SAE or an MAAE.
- The participant will be asked in the eDiary if they have taken any antipyretic or analgesic to treat or prevent fever or pain within 7 days after vaccination, including the day of injection. Reported antipyretic or analgesic medications should be recorded in the source document by the study site staff during the post-injection study visits or via other participant interactions (eg, telephone calls).

Concomitant medications (including vaccinations) will be coded using the WHO Drug Dictionary. If a participant takes a prohibited drug therapy, the investigator and the contract research organization (CRO)'s medical monitor will make a joint decision about continuing or withholding further injection of the participant based on the time the medication was administered, the drug's pharmacology and pharmacokinetics, and whether use of the medication will compromise the participant's safety or interpretation of the data. It is the investigator's responsibility to ensure that details regarding the concomitant medications are adequately recorded in the eCRF.

##### 5.5.3. Concomitant Medications and Vaccines that May Lead to the Elimination of a Participant from Per-Protocol Analyses

The use of the following concomitant medications and/or vaccines will not require withdrawal of the participant from the study but may determine a participant's evaluability in the per-protocol (PP) analysis (analysis sets are described in Section 8.4):

- Any investigational or nonregistered product (drug or vaccine) other than the IP used during the study period.
- Immunosuppressants or other immune-modifying drugs administered chronically (ie, more than 14 days in total) during the study period. For corticosteroids, this will mean

that prednisone ≥ 10 mg/day or the equivalent is not permitted. Inhaled, nasal, and topical steroids are allowed.

- Long-acting immune-modifying drugs administered at any time during the study period (eg, infliximab).
- An authorized or licensed vaccine administered during the period from 28 days before through 28 days after vaccination, except for any licensed influenza vaccine that was administered 14 days before or after vaccination.
- Immunoglobulins and/or any blood products administered during the study period.

In addition, any participant confirmed to have received or plans to receive a non-study COVID-19 vaccine, either licensed or under EUA, may also not be included in the PP analysis.

#### 5.6. Intervention After the End of the Study

Any SAE occurring after the end of the study and considered to be caused by the study vaccine must be reported to the Sponsor.

### 6. DELAY OR DISCONTINUATION OF STUDY INTERVENTION AND PARTICIPANT DISCONTINUATION/WITHDRAWAL

#### 6.1. Criteria for Delay of Vaccine Administration

##### 6.1.1. Individual Participant Criteria for Delay of Study Vaccination

Body temperature must be measured before vaccination. The following events constitute criteria for delay of injection, and, if either of these events occur at the time scheduled for dosing, the participant may be injected at a later date within the time window specified in the SoE (Table 17, Table 18, Table 19, and Table 20), or the participant may be discontinued from dosing at the discretion of the investigator (Section 6.2):

- Acute moderate or severe infection with or without fever at the time of dosing
- Fever, defined as body temperature ≥ 38.0°C (100.4°F) at the time of dosing

Afebrile participants with minor illnesses can be vaccinated at the discretion of the investigator. Participants with a fever of 38.0°C (100.4°F) or higher will be contacted within the time window acceptable for participation and re-evaluated for eligibility. If the investigator determines that the participant's health on the day of dosing temporarily precludes injection, the visit should be rescheduled within the allowed interval for that visit.

If a participant takes a prohibited drug therapy, an injection could be delayed within the visit window based on the joint decision of the investigator and the CRO's medical monitor (Section 5.5.3).

#### 6.2. Participant Discontinuation/Withdrawal from the Study

Participants who withdraw or are withdrawn from the study will not be replaced.

Participants can withdraw consent and withdraw from the study at any time, for any reason, without prejudice to further treatment the participant may need to receive. The investigator will request that the participant complete all study procedures pending at the time of withdrawal.

A participant who chooses not to receive the second booster dose of mRNA-1273.214 in Part A.2 of the study will NOT be considered to have discontinued treatment and can remain in the study in Part A.1 and will complete all scheduled visits and assessments.

Participants receiving a second or subsequent non-study COVID-19 booster vaccine can remain in the study for safety evaluation. The investigator will request that the participant complete all scheduled study visits. Blood and NP samples will not be collected in participants who have received subsequent non-study COVID-19 booster vaccines. If a participant meets the criteria for an illness visit (Section 7.1.6), NP samples will be collected during the illness visit per protocol.

If participant desires to withdraw from the study because of an AE, the investigator will attempt to obtain agreement to follow-up with the participant until the event is considered resolved or stable and will then complete the EoS eCRF.

Information related to the withdrawal will be documented in the eCRF. The investigator will document whether the decision to withdraw a participant from the study was made by the participant or by the investigator, as well as which of the following possible reasons was responsible for withdrawal:

- AE (specify)
- AESI (specify)
- SAE (specify)
- Death
- Lost to follow-up (LTFU)
- Physician decision (specify)
- Pregnancy
- Protocol deviation
- Study terminated by Sponsor
- Withdrawal of consent by participant (specify)
- Other (specify)

Participants who are withdrawn from the study because of AEs (including SAEs and AESI) must be clearly distinguished from participants who are withdrawn for other reasons. Investigators will follow-up with participants who are withdrawn from the study as result of an SAE or AE until resolution of the event.

A participant withdrawing from the study may request destruction of any samples taken and not tested, and the investigator must document this in the site study records.

If the participant withdraws consent for disclosure of future information, the Sponsor may retain and continue to use any data collected before such a withdrawal of consent (Section 10.2.6).

The Sponsor will continue to retain and use all research results that have already been collected for the study evaluation, unless the participant has requested destruction of these samples. All biological samples that have already been collected may be retained and analyzed at a later date (or as permitted by local regulations).

#### 6.3. Lost to Follow-up

A participant will be considered LTFU if he or she repeatedly fails to return for scheduled visits without stating an intention to withdraw consent and is unable to be contacted by the study site. The following actions must be taken if a participant fails to return to the clinic for a required study visit:

- The site must attempt to contact the participant and reschedule the missed visit as soon as possible, counsel the participant on the importance of maintaining the assigned visit schedule and ascertain whether the participant wishes to and/or should continue in the study.
- Before a participant is deemed LTFU, the investigator or designee must make every effort to regain contact with the participant (where possible, 3 telephone calls and, if

necessary, a certified letter to the participant's last known mailing address or local equivalent methods). These contact attempts (eg, dates of telephone calls and registered letters) should be documented in the participant's medical record.

- A participant who continues to be unreachable or continues to be noncompliant with study visits or procedures will be considered to have withdrawn from the study.
- A participant should not be considered LTFU until due diligence has been completed.

### 7. STUDY ASSESSMENTS AND PROCEDURES

Before performing any study procedures, all potential participants will sign an informed consent form (ICF) (as detailed in Section 10.2.6). Participants will undergo study procedures at the time points specified in the SoE (Table 17, Table 18, Table 19, and Table 20). A participant can also be seen for an unscheduled visit at any time during the study. An unscheduled visit may be prompted by reactogenicity issues, illness visit criteria for COVID-19, or new or ongoing AEs. The site also has the discretion to make reminder telephone calls or send text messages to inform the participant about visits, review eDiary requirements, or follow-up on ongoing or outstanding issues.

In accordance with “FDA Guidance on Conduct of Clinical Trials of Medical Products during COVID-19 Public Health Emergency” (DHHS 2020), investigators may convert study site visits to home visits or telemedicine visits with the approval of the Sponsor. Such action should be taken to protect the safety and well-being of participants and study site staff or to comply with state or municipal mandates.

General considerations for study assessments and procedures include the following:

- Protocol waivers or exemptions are not allowed. The study procedures and their timing must be followed as presented in the SoE (Table 17, Table 18, Table 19, and Table 20). Adherence to the study design requirements is essential and required for study conduct.
- Immediate safety concerns should be discussed with the Sponsor immediately upon occurrence or awareness to determine if the participant should continue participation in the study.
- All screening evaluations must be completed and reviewed to confirm that potential participants meet all eligibility criteria. The investigator will maintain a screening log to record details of all participants screened and to confirm eligibility or record reasons for screening failure, as applicable.
- Procedures conducted as a part of the participant’s routine clinical management and obtained before signing of the ICF may be utilized for screening or baseline purposes provided that the procedures meet the protocol-specified criteria and are performed within the time frame defined in the SoE (Table 17, Table 18, Table 19, and Table 20).
- The Screening Visit and Day 1 visit may be completed on the same day.

#### 7.1. Safety Assessments and Procedures

Safety assessments will include monitoring and recording of the following for each participant, according to the SoE (Table 17, Table 18, Table 19, and Table 20):

- Solicited local and systemic ARs (Section 7.4.3) that occur during the 7 days following vaccination (ie, the day of injection and 6 subsequent days). Solicited ARs will be recorded daily using eDiaries (Section 7.1.1).
- Unsolicited AEs observed or reported during the 28 days following vaccination (ie, the day of injection and 27 subsequent days). Unsolicited AEs are defined in Section 7.4.1.
- AEs leading to withdrawal from Day 1 through EoS.
- MAAEs from vaccination on Day 1 through EoS or withdrawal from the study.
- AESI from vaccination on Day 1 through EoS or withdrawal from the study.
- SAEs from vaccination on Day 1 through EoS or withdrawal from the study.
- Vital sign measurements before and after vaccination (Section 7.1.4).
- Physical examination findings (if performed) (Section 7.1.5).
- Assessments for SARS-CoV 2 infection from Day 1 through study completion.
- Details of all pregnancies in female participants will be collected after the start of study treatment and until the end of their participation in the study (Section 7.4.6).

The incidence and severity of the above events will be monitored by an IST on a regular basis.

Participants who receive a non-study second booster dose (or any subsequent doses, if available in the future), after having received all prior booster doses in the study, can remain in the study for safety follow-up.

##### 7.1.1. Use of Electronic Diaries

At the time of consent, the participants must confirm they will be willing to complete an eDiary using either an application downloaded to their smartphone or using a device that will be provided at the time of enrollment. Before enrollment on Day 1, the participant will be instructed to download the eDiary application or will be provided an eDiary device to record solicited ARs (Section 7.4.3) on Day 1.

On Day 1 (dosing day), participants will be instructed on thermometer usage to measure body temperature, ruler usage to measure injection site erythema and swelling/induration (hardness), and self-assessment for localized axillary swelling or tenderness on the same side as the injection arm.

On Day 1 (dosing day), participants will record data into the eDiary starting approximately 30 minutes after the injection under supervision of the study site staff to ensure successful entry of assessments. The study site staff will perform any retraining as necessary. Participants will continue to record data in the eDiary after they leave the study site, preferably in the evening and at the same time each day, on the day of injection and for 6 days following injection.

Participants will record the following data in the eDiary:

- Solicited local and systemic reactogenicity ARs, as defined in Section 7.4.3, that occur on the day of vaccination and during the 7 days after vaccination (ie, the day of injection and 6 subsequent days). Any solicited AR that is ongoing beyond Day 7 will be reported in the eDiary until it has resolved, and not to exceed 28 days after vaccination. ARs recorded in the eDiary beyond Day 7 should be reviewed by the study site staff either during the next scheduled telephone call or at the next study site visit.

- Daily oral body temperature measurement should be performed at approximately the same time each day using the thermometer provided by the study site. If body temperature is taken more than once in a given day, only the highest temperature reading should be recorded.
- Other measurements, as applicable, for solicited local ARs (injection site erythema and swelling/induration) will be performed using the ruler provided by the study site.
- Any medications taken to treat or prevent pain or fever on Day 1 or for the next 6 days.

The eDiary will be the only source document allowed for solicited systemic or local ARs (including body temperature measurements). Participants will be instructed to complete eDiary entries daily. The participant will have a limited window on the following day to complete assessments for the previous day; quantitative temperature recordings and measurement of any injection site erythema or swelling/induration reported on the following day may be excluded from the analyses of solicited ARs.

Any new safety information reported during safety telephone calls or at site visits (including a solicited AR) that is not already captured in the eDiary will be described in the source documents as a verbally reported event. Any AR reported in this manner must be described as an unsolicited event and therefore entered on the AE eCRF.

Study site staff will review eDiary data with participants during the safety call 7 days after vaccination.

The eDiary will also be used every 2 weeks from Day 36 to Day 162, and from Day 202 to Day 342 (Parts A.1, B, C, D, F, and G only) or every 2 weeks from Day 36 to Day 162 (Parts A.2, H, and J only), to capture the occurrence of MAAEs, AESI, SAEs, and AEs leading to withdrawal. The eDiary will prompt the participant to complete an eDiary questionnaire that collects the following data:

- Changes in health since last completing the questionnaire or since in contact with the study site
- Known exposure to someone with known COVID-19 or SARS-CoV-2 infection
- Any experience of symptoms of COVID-19
- Any MAAEs, AESI, or SAEs

If an eDiary record results in identification of relevant safety events or of symptoms of COVID-19, a follow-up safety call will be triggered.

Apart from the safety telephone calls described in Section 7.1.2, each participant will complete a questionnaire in an eDiary as shown in the SoE (Table 17, Table 18, Table 19, and Table 20). The eDiary responses will be reviewed by study site personnel and may result in a follow-up safety call by the site to the participant.

###### 7.1.1.1. Ancillary Supplies for Participant Use

Study sites will distribute Sponsor-provided oral thermometers and rulers for use by participants to assess body temperature and injection site reactions, respectively, for recording solicited ARs in the eDiaries. Based on availability, smartphone devices may be provided to those participants who do not have their own device to use for eDiary activities.

##### 7.1.2. Safety Telephone Call

A safety telephone call is a telephone call made to the participant by a trained site personnel. This call will follow an approved script, which will facilitate the collection of relevant safety information. There will be a safety telephone call on Day 8 for each participant to discuss their health and review their eDiary. Subsequent safety calls by the site to each participant will occur every 2 weeks from Day 43 to Day 169, and from Day 209 to Day 349 (Parts A.1, B, C, D, F, and G only) (Table 17) and every 2 weeks from Day 43 to Day 169 for Parts A.2 (Table 18), H (Table 19), and J (Table 20). The participant will be interviewed according to the script about the occurrence of AEs, MAAEs, AESI, SAEs, AEs leading to withdrawal, concomitant medications associated with those events, and any non-study vaccinations (Section 7.4.7). In addition, study personnel will collect information on known participant exposure to someone with COVID-19 or SARS-CoV-2 infection and on the participant's experience of COVID-19 symptoms. All safety information collected from the telephone call must be documented in the source documents as described by the participant and not documented on the script used for the safety telephone contact. As noted in Section 7.1.1, an unscheduled follow-up safety call may be triggered if an eDiary record results in identification of a relevant safety event.

##### 7.1.3. Laboratory Assessments

No routine safety laboratory assessments are planned for this study.

A point-of-care urine pregnancy test will be performed at Day 1 before vaccination. At any time, a pregnancy test either via blood or point-of-care urine can be performed, at the discretion of the investigator. If not documented in a female participant's medical records, an FSH test may be performed at the Screening Visit, as necessary and at the discretion of the investigator, to confirm postmenopausal status.

##### 7.1.4. Vital Sign Measurements

Vital sign measurements will include systolic and diastolic blood pressure, heart rate, respiratory rate, and body temperature (preferred route is oral). The participant will be seated for at least 5 minutes before all measurements are taken. Vital signs will be measured at the time points indicated in the SoE (Table 17, Table 18, Table 19, and Table 20). Vital signs are to be collected pre- and post-dosing on the day of injection (Day 1) only. When applicable, vital sign measurements should be performed before blood collection.

Participants who are febrile (body temperature ≥ 38.0°C [100.4°F]) before injection on Day 1 must be rescheduled within the relevant window period to receive the injection. Afebrile participants with minor illnesses may be vaccinated at the discretion of the investigator.

##### 7.1.5. Physical Examinations

A full physical examination, including height and weight, will be performed at Day 1 as indicated in the SoE (Table 17, Table 18, Table 19, and Table 20). The full examination will include assessment of skin, head, ears, eyes, nose, throat, neck, thyroid, lungs, heart, cardiovascular system, abdomen, lymph nodes, and musculoskeletal system and extremities. Any clinically significant finding identified during a study visit should be reported as an MAAE.

Symptom-directed physical examinations may be performed at other time points at the discretion of the investigator. On the day of vaccination, before injection, the arm receiving the injection should be examined and the associated lymph nodes should be evaluated.

##### 7.1.6. Assessment for SARS-CoV-2 Infection

Participants will have NP samples collected for SARS-CoV-2 testing at time points specified in the SoE (Table 17, Table 18, Table 19, and Table 20).

A study illness visit or a consultation will be arranged within 24 hours or as soon as possible to collect an NP swab (Table 12 and Table 13) to ascertain the presence of SARS-CoV-2 via RT-PCR if a participant experiences any of the following (the presence of any one of these symptoms lasting at least 48 hours [except for fever and/or respiratory symptoms]):

- Signs or symptoms of SARS-CoV-2 infection as defined by the CDC (CDC 2020), including:
  - Fever (temperature ≥ 38.0°C [100.4°F]) or chills (of any duration, including ≤ 48 hours)
  - Cough (of any duration, including ≤ 48 hours)
  - Shortness of breath and/or difficulty breathing (of any duration, including ≤ 48 hours)
  - Fatigue
  - Muscle or body aches
  - Headache
  - New loss of taste and/or smell
  - Sore throat, congestion, or runny nose
  - Nausea or vomiting
  - Diarrhea
- MAAE suggesting a SARS-CoV-2 infection
- Clinical or radiographical evidence of pneumonia

Additionally, clinical information will be carefully collected to evaluate the severity of the clinical case. All findings will be recorded in the eCRF.

It is important to note that some of the symptoms of COVID-19 overlap with solicited systemic ARs that are expected after vaccination with mRNA-1273 (eg, myalgia, headache, fever, and chills). During the first 7 days after vaccination, when these solicited ARs are common, investigators should use their clinical judgment to decide whether an NP swab should be collected. The collection of an NP swab prior to the Day 1 and Day 29 vaccination can help ensure that cases of COVID-19 are not overlooked. Any study participant reporting respiratory symptoms during the 7-day period after vaccination should be evaluated for COVID-19.

If scheduled, a study site illness visit may include additional assessments such as medical history, physical examination, and blood sampling for clinical laboratory testing. The NP swab sample may be tested by multiplex RT-PCR for respiratory viruses besides SARS-CoV-2 to evaluate the severity of the clinical case. Radiologic imaging studies may be conducted. Blood samples will be collected at all illness visits for potential future immunologic assessment of SARS-CoV-2 infection.

Cases are defined as participants meeting clinical criteria based both on symptoms for COVID-19 and on RT-PCR detection of SARS-CoV-2 from samples collected within 72 hours of the study participant reporting symptoms meeting the definition of COVID-19. Participants who are hospitalized for COVID-19 without the opportunity for a clinic visit will also be considered cases, assuming that the symptomology criteria for COVID-19 are met and a respiratory sample is positive for SARS-CoV-2 by PCR at a Clinical Laboratory Improvement Amendments (CLIA)-certified or CLIA-certified waiver laboratory. Investigators are encouraged to try to obtain a respiratory sample during the course of hospitalization for submission to the study central laboratory, if feasible.

Symptomatic COVID-19 is defined by the presence of one of the CDC-listed symptoms (CDC 2020) and a positive RT-PCR test on a respiratory sample. Asymptomatic SARS-CoV-2 infection is defined as a positive RT-PCR test on a respiratory sample in the absence of symptoms or a positive serologic test for antinucleocapsid antibody after a negative test result at the time of enrollment, with the serologic assay detecting previously resolved SARS-CoV-2 infections that may have occurred between visits, and the RT-PCR to detect active viral infection at the time of a visit. If participants are confirmed to have SARS-CoV-2 infection and are symptomatic or asymptomatic, the investigator will notify the participants' primary care physicians of the diagnosis and the local public health authorities as required per local regulations.

If the participant had known exposure to COVID-19 (eg, exposure to someone with a confirmed case of COVID-19), it will be captured in the COVID-19 exposure form, and the participant will continue to follow all remaining study assessments as scheduled. Likewise, participants with a confirmed case of COVID-19 will continue to follow all remaining study assessments as scheduled.

Any confirmed symptomatic COVID-19 infection occurring in participants will be captured as an MAAE along with relevant concomitant medications and details about severity, seriousness, and outcome.

#### 7.2. Immunogenicity Assessments

Blood samples for immunogenicity assessments will be collected at the time points indicated in the SoE (Table 17, Table 18, Table 19, and Table 20). The following immunogenicity assessments will be measured contemporaneously with Day 57 samples from participants in the mRNA-1273-P301 (COVE) study (historical control samples; further details will be provided in the SAP):

- Serum binding antibody (bAb) level against SARS-CoV-2 as measured by ligand-binding assay specific to the SARS-CoV-2 S protein and the S protein RBD.
- Serum nAb level against SARS-CoV-2 as measured by pseudovirus neutralization assays.
- Testing for serologic markers for SARS-CoV-2 infection as measured by anti-nucleocapsid antibodies detected by immunoassay (Day 1, Day 29, Day 91 [Parts A.2, F, G, H, and J only], Day 181, and Day 366 [Parts A.1, B, C, D, F, and G only]).
- Sequencing of SARS-CoV-2 genome in samples positive for viral infection by PCR testing.
- PBMCs may be collected for a subset of participants at selected sites to characterize T-cell and B-cell response against SARS-CoV-2 and variants.

Sample aliquots will be designed to ensure that backup samples are available and that vial volumes are likely to be adequate for future testing needs. The actual time and date of each sample collected will be recorded in the eCRF. Handling and preparation of the samples for analysis, as well as shipping and storage requirements, will be provided in a separate study manual.

Measurement of bAb and nAb levels will be performed in a laboratory designated by the Sponsor.

According to the ICF (Section 10.2.6), excess serum from immunogenicity testing may be used for future research, which may be performed at the discretion of the Sponsor to further characterize the immune response to SARS-CoV-2, additional assay development, and the immune response across CoV.

Nasopharyngeal swab samples to be collected are also shown (Table 12 and Table 13).

**Table 12: Blood and Nasopharyngeal Swab Sampling for Parts A.1, B, C, D, F, and G**

| Sample Name | D1 (Baseline) | D15 | D29 | D91 (Part F and Part G only) | D181 | D366 | UNS / Illness |
|---|---|---|---|---|---|---|---|
| Anti-SARS-CoV-2 | 1 | | 1 | 1 | 1 | 1 | 1 |
| Immunogenicity | 1 | 1 | 1 | 1 | 1 | 1 | 1 |
| RT-PCR/Sequencing | 1 | | 1 | 1 | 1 | | 1 |
| BioFire 2.1 | | | | | | | 1 |
| PBMC may be collected for a subset of participants at selected sites | 1 | 1 | 1 | 1 | | | |

Abbreviations: D =day; N/A = not applicable; NP = nasopharyngeal; PBMC = Peripheral Blood Mononuclear Cells; RT-PCR = reverse transcriptase polymerase chain reaction; SST = serum separator tube; UNS = unscheduled visit.

**Table 13: Blood and Nasopharyngeal Swab Sampling for Parts A.2, H, and J**

| Sample Name | D1 (Baseline) | D15 | D29 | D91 | D181 | UNS / Illness |
|---|---|---|---|---|---|---|
| Anti-SARS-CoV-2 | 1 | | 1 | 1 | 1 | 1 |
| Immunogenicity | 1 | 1 | 1 | 1 | 1 | 1 |
| RT-PCR/Sequencing | 1 | | 1 | 1 | 1 | 1 |
| BioFire 2.1 | | | | | | 1 |

Abbreviations: D =day; N/A = not applicable; NP = nasopharyngeal; PBMC = Peripheral Blood Mononuclear Cells; RT-PCR = reverse transcriptase polymerase chain reaction; SST = serum separator tube; UNS = unscheduled visit.

#### 7.3. Efficacy Assessments

Vaccine efficacy will not be formally assessed in this study, but active surveillance for COVID-19 and SARS-CoV-2 infection through weekly contact and blood draws (see Table 17, Table 18, Table 19, and Table 20), will be performed.

#### 7.4. Safety Definitions and Related Procedures

##### 7.4.1. Adverse Event

An AE is defined as any untoward medical occurrence associated with the use of a drug in humans, whether or not considered drug related.

**Events Meeting the Adverse Event Definition**

- Exacerbation of a chronic or intermittent pre-existing condition including either an increase in frequency and/or intensity of the condition
- New conditions detected or diagnosed after vaccination even though they may have been present before the start of the study

**Events NOT Meeting the Adverse Event Definition**

- Procedures planned before study entry (eg, hospitalization for preplanned surgical procedure).
- Medical or surgical procedure (eg, endoscopy, appendectomy): the condition that leads to the procedure should be the AE.
- Situations in which an untoward medical occurrence did not occur (social and/or convenience admission to a hospital).

An AR is any AE for which there is a reasonable possibility that the vaccine caused the AE (Section 7.4.3). For the purposes of investigational new drug safety reporting, "reasonable possibility" means that there is evidence to suggest a causal relationship between the vaccine and the AE.

An unsolicited AE is any AE reported by the participant that is not specified as a solicited AR in the protocol or is specified as a solicited AR but starts outside the protocol-defined period for reporting solicited ARs (ie, 7 days after vaccination). Any unsolicited AE which began during mRNA-1273-P301 (COVE) but is ongoing at the time of enrollment in this study, should be documented as medical history.

##### 7.4.2. Serious Adverse Events

An AE (including an AR) is considered an SAE if, in the view of either the investigator or Sponsor, it results in any of the following outcomes:

- **Death**
  A death that occurs during the study or that comes to the attention of the investigator during the protocol-defined follow-up period must be reported to the Sponsor, whether or not it is considered related to the IP.
- **Is life-threatening**
  An AE is considered life-threatening if, in the view of either the investigator or the Sponsor, its occurrence places the participant at immediate risk of death. It does not include an AE that, had it occurred in a more severe form, might have caused death.
- **Inpatient hospitalization or prolongation of existing hospitalization**
  In general, inpatient hospitalization indicates the participant was admitted to the hospital or emergency ward for at least one overnight stay for observation and/or treatment that would not have been appropriate in the physician's office or outpatient setting. The hospital or emergency ward admission should be considered an SAE regardless of whether opinions differ as to the necessity of the admission. Complications that occur during inpatient hospitalization will be recorded as an AE; however, if a complication/AE prolongs hospitalization or otherwise fulfills SAE criteria, the complication/AE will be recorded as a separate SAE.
- **Persistent or significant incapacity or substantial disruption of the ability to conduct normal life functions**
  This definition is not intended to include experiences of relatively minor medical significance such as uncomplicated headache, nausea/vomiting, diarrhea, influenza, and accidental trauma (eg, sprained ankle) which may interfere with or prevent everyday life functions but do not constitute a substantial disruption.
- **Congenital anomaly or birth defect**
- **Medically important event**
  Medical judgment should be exercised in deciding whether SAE reporting is appropriate in other situations, such as important medical events that may not be immediately life-threatening or result in death or hospitalization but may jeopardize the participant or require medical or surgical intervention to prevent one of the other outcomes listed in the above definition. These events should usually be considered serious. Examples of such medical events include allergic bronchospasm requiring intensive treatment in an emergency room or at home, blood dyscrasias or convulsions that do not result in inpatient hospitalization, or the development of drug dependency or drug abuse.

##### 7.4.3. Solicited Adverse Reactions

The term "reactogenicity" refers to the occurrence and intensity of selected signs and symptoms (ARs) occurring after IP injection. The eDiary will solicit daily participant reporting of ARs using a structured checklist (Section 7.1.1). Participants will record such occurrences in an eDiary during the 7 days after vaccination (ie, the day of injection and 6 subsequent days).

Severity grading of reactogenicity will occur automatically based on participant entry into the eDiary according to the grading scales presented in Table 14 modified from the Toxicity Grading Scale for Healthy Adult and Adolescent Volunteers Enrolled in Preventive Vaccine Clinical Trials (DHHS 2007).

If a solicited local or systemic AR continues beyond 7 days after dosing, the participant will be prompted daily to capture solicited local or systemic AR in the eDiary until resolution. ARs recorded in eDiaries beyond Day 7 should be reviewed by the study staff either via phone call or at the next study visit. All solicited ARs (local and systemic) will be considered causally related to dosing.

**Table 14: Solicited Adverse Reactions and Grades**

| Reaction | Grade 0 | Grade 1 | Grade 2 | Grade 3 | Grade 4 |
|---|---|---|---|---|---|
| Injection site pain | None | Does not interfere with activity | Repeated use of over-the-counter pain reliever > 24 hours or interferes with activity | Any use of prescription pain reliever or prevents daily activity | Requires emergency room visit or hospitalization |
| Injection site erythema (redness) | < 25 mm/ < 2.5 cm | 25 – 50 mm/ 2.5 – 5 cm | 51 – 100 mm/ 5.1 – 10 cm | > 100 mm/ > 10 cm | Necrosis or exfoliative dermatitis |
| Injection site swelling/induration (hardness) | < 25 mm/ < 2.5 cm | 25 – 50 mm/ 2.5 – 5 cm | 51 – 100 mm/ 5.1 – 10 cm | > 100 mm/ > 10 cm | Necrosis |
| Axillary (underarm) swelling or tenderness ipsilateral to the side of injection | None | No interference with activity | Repeated use of over-the-counter (non-narcotic) pain reliever > 24 hours or some interference with activity | Any use of prescription (narcotic) pain reliever or prevents daily activity | Emergency room visit or hospitalization |
| Headache | None | No interference with activity | Repeated use of over-the-counter pain reliever > 24 hours or some interference with activity | Significant; any use of prescription pain reliever or prevents daily activity | Requires emergency room visit or hospitalization |

| **Reaction** | **Grade 0** | **Grade 1** | **Grade 2** | **Grade 3** | **Grade 4** |
|---|---|---|---|---|---|
| Fatigue | None | No interference with activity | Some interference with activity | Significant; prevents daily activity | Requires emergency room visit or hospitalization |
| Myalgia (muscle aches all over body) | None | No interference with activity | Some interference with activity | Significant; prevents daily activity | Requires emergency room visit or hospitalization |
| Arthralgia (joint aches in several joints) | None | No interference with activity | Some interference with activity | Significant; prevents daily activity | Requires emergency room visit or hospitalization |
| Nausea/vomiting | None | No interference with activity or 1-2 episodes/ 24 hours | Some interference with activity or > 2 episodes/24 hours | Prevents daily activity, requires outpatient intravenous hydration | Requires emergency room visit or hospitalization for hypotensive shock |
| Chills | None | No interference with activity | Some interference with activity not requiring medical intervention | Prevents daily activity and requires medical intervention | Requires emergency room visit or hospitalization |
| Fever (oral) | < 38.0°C<br>< 100.4°F | 38.0 – 38.4°C<br>100.4 – 101.1°F | 38.5 – 38.9°C<br>101.2 – 102.0°F | 39.0 – 40.0°C<br>102.1 – 104.0°F | > 40.0°C<br>> 104.0°F |

Any solicited AR that meets any of the following criteria must be entered into the participant's source document and must also be recorded by the study site staff on the solicited AR page of the participant's eCRF:

- Solicited local or systemic AR that results in a visit to a healthcare provider (HCP), to be recorded as an MAAE (Section 7.4.4)
- Solicited local or systemic AR leading to the participant withdrawing from the study or the participant being withdrawn from the study by the investigator (AE leading to withdrawal)
- Solicited local or systemic AR lasting beyond 7 days post-injection
- Solicited local or systemic AR that otherwise meets the definition of an SAE

##### 7.4.4. Medically Attended Adverse Events

An MAAE is an AE that leads to an unscheduled visit to an HCP. This would include visits to a study site for unscheduled assessments (eg, abnormal laboratory follow-up, COVID-19 [Section 7.1.6]) and visits to HCPs external to the study site (eg, urgent care, primary care physician). Investigators will review unsolicited AEs for the occurrence of any MAAEs. Unsolicited AEs will be captured on the AE page of the eCRF.

All confirmed COVID-19 cases will be recorded as MAAEs.

All suspected cases of anaphylaxis should be recorded as MAAEs and reported as an SAE, based on the criteria for a medically important event, unless the event meets other serious criteria. As an SAE, the event should be reported to the Sponsor or designee immediately and in all circumstances within 24 hours per Section 7.4.11. The investigator will submit any updated anaphylaxis case data to the Sponsor within 24 hours of it being available. For reporting purposes, a participant who displays signs or symptoms consistent with anaphylaxis (as follows) should be reported as a potential case of anaphylaxis. This is provided as general guidance for investigators and is based on the Brighton Collaboration case definition (Rüggeberg et al 2007).

Anaphylaxis is an acute hypersensitivity reaction with multi-organ system involvement that can present as, or rapidly progress to, a severe life-threatening reaction. It may occur following exposure to allergens from a variety of sources.

Anaphylaxis is a clinical syndrome characterized by the following:

- Sudden onset AND
- Rapid progression of signs and symptoms AND
- Involves 2 or more organ systems, as follows:
  - **Skin/mucosal:** urticaria (hives), generalized erythema, angioedema, generalized pruritus with skin rash, generalized prickle sensation, and red and itchy eyes.
  - **Cardiovascular:** measured hypotension, clinical diagnosis of uncompensated shock, loss of consciousness or decreased level of consciousness, and evidence of reduced peripheral circulation.
  - **Respiratory:** bilateral wheeze (bronchospasm), difficulty breathing, stridor, upper airway swelling (lip, tongue, throat, uvula, or larynx), respiratory distress, persistent dry cough, hoarse voice, sensation of throat closure, sneezing, and rhinorrhea.
  - **Gastrointestinal:** diarrhea, abdominal pain, nausea, and vomiting.

##### 7.4.5. Adverse Event of Special Interest

An AESI is an AE (serious or nonserious) of scientific and medical concern specific to the Sponsor's product or program for which ongoing monitoring and immediate notification by the investigator to the Sponsor is required and documentation is in the form of a case narrative. Such events may require further investigation to characterize and understand them. Section 10.4 (Appendix 4) provides a list of AESI pertinent to this study. All AESI will be collected through the entire study period and must be reported to the Sponsor or designee immediately and in all circumstances within 24 hours of becoming aware of the event via the electronic data capture (EDC) system. If a site receives a report of a new AESI from a study participant or receives updated data on a previously reported AESI and the eCRF has been taken offline, then the site can report this information on a paper AESI form using the SAE Mailbox, or the SAE Fax line (Section 7.4.11).

**Myocarditis and/or Pericarditis**

Very rare events of myocarditis and/or pericarditis have been reported after vaccination with the mRNA-1273 vaccine. All suspected cases of probable and confirmed myocarditis, pericarditis, or myopericarditis should be reported as an AESI even if it does not meet criteria per the CDC case definition. The event should also be reported as an SAE, if it meets seriousness criteria. As an SAE, the event should be reported to the Sponsor or designee immediately and in all circumstances within 24 hours as per Section 7.4.2. The investigator will submit any updated myocarditis, pericarditis, or myopericarditis case data to the Sponsor within 24 hours of it being available. For reporting purposes, any events suspicious for myocarditis, pericarditis, or myopericarditis should be reported as an AESI. The CDC case definition is displayed below as guidance (Gargano et al, 2021). These definitions are intended to serve as a guide to help reporting of suspected cases of myocarditis, pericarditis, or myopericarditis, but the diagnosis of suspected cases are left to the investigator’s clinical judgment.

**Acute Myocarditis Case Definition**

Presence of ≥ 1 new or worsening of the following clinical symptoms (persons who lack the listed symptoms but who meet other criteria may be classified as subclinical myocarditis [probable or confirmed]):

- Chest pain/pressure/discomfort
- Dyspnea/shortness of breath/pain with breathing
- Palpitations
- Syncope

AND

For probable case:

≥ 1 new finding of:

- Troponin level above upper limit of normal (any type of troponin)
- Abnormal electrocardiogram (ECG or EKG) or rhythm monitoring findings consistent with myocarditis
  - To meet the ECG or rhythm monitoring criterion, a probable case must include at least one of:
    - ST segment or T-wave abnormalities
    - Paroxysmal or sustained atrial, supraventricular, or ventricular arrhythmias

  - AV nodal conduction delays or intraventricular conduction defects

- Abnormal cardiac function or wall motion abnormalities on echocardiogram
- cMRI finding consistent with myocarditis (Ferreira et al, 2018)

AND

- No other identifiable cause of the symptoms and findings

<u>For confirmed case</u>:

- Histopathologic confirmation of myocarditis (using Dallas criteria [Aretz et al, 1987])

OR

- cMRI findings consistent with myocarditis in the presence of troponin level above upper limit of normal (any type of troponin)

AND

- No other identifiable cause of the symptoms and findings

**Acute Pericarditis Case Definition**

Presence of ≥ 2 new or worsening of the following clinical features (Adler et al, 2015):

- Acute chest pain (Typically described as pain made worse by lying down, deep inspiration, or cough; and relieved by sitting up or leaning forward, although other types of chest pain may occur)
- Pericardial rub on exam
- New ST-elevation or PR-depression on EKG
- New or worsening pericardial effusion on echocardiogram or MRI

**Case Definition of Myopericarditis**

Participants who meet criteria for both myocarditis and pericarditis may be described under myopericarditis.

An independent Cardiac Event Adjudication Committee (CEAC) that includes pediatric and adult cardiologists will review suspected cases of myocarditis and pericarditis to determine if they meet CDC criteria of “probable” or “confirmed” events, and to assess severity (Gargano et al 2021). Any cases that the CEAC assesses as representing probably or confirmed cases of myocarditis or pericarditis will be referred to the Sponsor, who will then make a final decision on whether to suspend further enrollment and/or study dosing based on assessment of the overall potential risk to study participants.

The CEAC will operate under the rules of an approved charter that will be written and reviewed at the organizational meeting of the CEAC. Details regarding the CEAC composition, responsibilities, procedures, and frequency of data review will be defined in its charter.

##### 7.4.6. Recording and Follow-up of Pregnancy

Female participants who have a positive pregnancy test at screening should not be enrolled; participants who have a positive pregnancy test at any time during the study should receive no further dosing with IP but should be asked to remain in the study and be monitored for safety.

Details of all pregnancies in female participants will be collected after the start of study treatment and until the end of their participation in the study.

- If a pregnancy is reported, the investigator should inform the Sponsor within 24 hours of learning of the pregnancy and should follow the procedures outlined in this section.
- Abnormal pregnancy outcomes (eg, spontaneous abortion, fetal death, stillbirth, congenital anomalies, ectopic pregnancy) will be considered as SAEs.

Pregnancies occurring in participants after enrollment must be reported to Sponsor or designee within 24 hours of the site learning of its occurrence, using the SAE Mailbox, or the SAE Fax line (Section 7.4.11). If the participant agrees to submit this information, the pregnancy must be followed to determine the outcome, including spontaneous or voluntary termination, details of the birth, and the presence or absence of any birth defects, congenital abnormalities, or maternal and/or newborn complications. This follow-up should occur even if intended duration of the safety follow-up for the study has ended. Pregnancy report forms will be distributed to the study site to be used for this purpose. The investigator must immediately (within 24 hours of awareness) report to the Sponsor any pregnancy resulting in an abnormal outcome according to the procedures described for SAEs.

##### 7.4.7. Eliciting and Documenting Adverse Events

The investigator is responsible for ensuring that all AEs and SAEs are recorded in the eCRF and reported to the Sponsor.

Solicited ARs will be collected from Day 1 through 7 days after vaccination. Other (unsolicited) AEs will be collected from Day 1 through 28 days after vaccination.

The MAAEs, AESI, AE leading to withdrawal, AESI, and SAEs will be collected from participants as specified in the SoE (Table 17, Table 18, Table 19, and Table 20) until the end of their participation in the study. Any AEs occurring before receipt of IP will be analyzed separately from AEs occurring after receipt of the study vaccine.

At every study site visit or telephone contact, participants will be asked a standard question to elicit any medically related changes in their well-being (including COVID-19 symptoms) according to the scripts provided. Participants will also be asked if they have been hospitalized, had any accidents, used any new medications, changed concomitant medication regimens (both prescription and over-the-counter medications), or had any non-study vaccinations.

In addition to participant observations, physical examination findings, or other documents relevant to participant safety classified as an AE will be documented on the AE page of the eCRF.

After the initial AE/SAE report, the investigator is required to proactively follow each participant at subsequent visits/contacts. All AEs and SAEs will be treated as medically appropriate and followed until resolution, stabilization, the event is otherwise explained, or the participant is LTFU (as defined in Section 6.3).

##### 7.4.8. Assessment of Intensity

An event is defined as "serious" when it meets at least one of the predefined outcomes as described in the definition of an SAE (Section 7.4.2), NOT when it is rated as severe.

The severity (or intensity) of an AR or AE refers to the extent to which it affects the participant's daily activities. The Toxicity Grading Scale for Healthy Adult and Adolescent Volunteers Enrolled in Preventive Vaccine Clinical Trials (DHHS 2007) will be used to categorize local and systemic reactogenicity events (solicited ARs), clinical laboratory test results, and vital sign measurements observed during this study. Specific criteria for local and systemic reactogenicity events are presented in Section 7.4.3.

The determination of severity for all unsolicited AEs should be made by the investigator based upon medical judgment and the definitions of severity as follows:

- Mild: These events do not interfere with the participant's daily activities.
- Moderate: These events cause some interference with the participant's daily activities and require limited or no medical intervention.
- Severe: These events prevent the participant's daily activity and require intensive therapeutic intervention.

Study staff should elicit from the participant the impact of AEs on the participant's activities of daily living to assess severity and document appropriately in the participant's source documentation. Changes in the severity of an AE should be documented in the participant's source documentation to allow an assessment of the duration of the event at each level of intensity to be performed. An AE characterized as intermittent requires documentation of onset and duration of each episode. An AE that fluctuates in severity during the course of the event is reported once in the eCRF at the highest severity observed.

##### 7.4.9. Assessment of Causality

The investigator's assessment of an AE's relationship to IP is part of the documentation process but is not a factor in determining what is or is not reported in the study.

The investigator will assess causality (ie, whether there is a reasonable possibility that the IP caused the event) for all AEs and SAEs. The relationship will be characterized using the following classification:

**Not related:** There is not a reasonable possibility of a relationship to the IP. Participant did not receive the IP OR temporal sequence of the AE onset relative to administration of the IP is not reasonable OR the AE is more likely explained by another cause than the IP.

**Related:** There is a reasonable possibility of a relationship to the IP. There is evidence of exposure to the IP. The temporal sequence of the AE onset relative to the administration of the IP is reasonable. The AE is more likely explained by the IP than by another cause.

##### 7.4.10. Reporting Adverse Events

The investigator is responsible for reporting all AEs that are observed or reported during the study, regardless of their relationship to IP or their clinical significance. If there is any doubt as to whether a clinical observation is an AE, the event should be reported.

All unsolicited AEs reported or observed during the study will be recorded on the AE page of the eCRF. Information to be collected includes type of event, time of onset, investigator-specified assessment of severity (impact on activities of daily living) and relationship to IP, time of resolution of the event, seriousness, as well as any required treatment or evaluations, and outcome. The unsolicited AEs resulting from concurrent illnesses, reactions to concurrent illnesses, reactions to concurrent medications, or progression of disease states must also be reported. All AEs will be followed until they are resolved or stable or judged by the investigator to be not clinically significant. The Medical Dictionary for Regulatory Activities (MedDRA) will be used to code all unsolicited AEs.

Any medical condition that is present at the time of screening but does not deteriorate should not be reported as an unsolicited AE. However, if it deteriorates at any time during the study, it should be recorded as an unsolicited AE.

##### 7.4.11. Reporting SAEs

Prompt notification by the investigator to the Sponsor of an SAE is essential so that legal obligations and ethical responsibilities toward the safety of participants and the safety of a study intervention under clinical investigation are met.

Any AE considered serious by the investigator or that meets SAE criteria (Section 7.4.2) must be reported to the Sponsor immediately (within 24 hours of becoming aware of the SAE) via the EDC system. The investigator will assess whether there is a reasonable possibility that the IP caused the SAE. The Sponsor will be responsible for notifying the relevant regulatory authorities of any SAE as outlined in 21 US CFR Parts 312 and 320. The investigator is responsible for notifying the IRB directly.

If the eCRF is unavailable at the time of the SAE, the following contact information is to be used for SAE reporting:

- SAE Mailbox:
- SAE Fax Line (USA and Canada): +1-617-649-3910

Regulatory reporting requirements for SAEs are described in Section 7.4.15.

The investigator and any qualified designees are responsible for detecting, documenting, and recording events that meet the definition of an AE, including SAEs, and remain responsible for following up AEs that are serious, considered related to IP or study procedures, or that caused the participant to discontinue the study.

##### 7.4.12. Time Period and Frequency for Collecting AE, AESI, and SAE Information

Medical occurrences that begin before IP dosing but after obtaining informed consent will be recorded in the Medical History/Current Medical Conditions section of the eCRF and not in the

AE section; however, if the condition worsens at any time during the study, it will be recorded and reported as an AE.

AEs may be collected as follows:

- Observing the participant.
- Receiving an unsolicited complaint from the participant.
- Questioning the participant in an unbiased and nonleading manner.

Solicited AEs will be collected from the day of injection through 6 days after vaccination. Other (unsolicited) AEs will be collected from the day of injection through 28 days after vaccination.

Serious AEs (including AESI) will be collected from the start of IP dosing until the last day of study participation.

All SAEs and AESI will be recorded and reported to the Sponsor or designee immediately and under no circumstance should this exceed 24 hours of becoming aware of the event via the EDC system. If a site receives a report of a new SAE or AESI from a study participant or receives updated data on a previously reported SAE or AESI and the eCRF has been taken offline, then the site can report this information on a paper SAE or AESI form using the SAE Mailbox or the SAE Fax line (Section 7.4.11).

An abnormal value or result from a clinical or laboratory evaluation can also indicate an AE if it is determined by the investigator to be clinically significant (eg, leads to study drug discontinuation, or meets any serious criteria). If this is the case, it must be recorded in the source document and as an AE on the appropriate AE form(s). The evaluation that produced the value or result should be repeated until that value or result returns to normal or is stabilized and the participant's safety is not at risk.

Investigators are not obligated to actively seek AEs or SAEs after EoS participation. However, if the investigator learns of any SAE (including a death) at any time after a participant has withdrawn from or completed the study, and the investigator considers the event to be reasonably related to the IP or study participation, the investigator must promptly notify the Sponsor.

##### 7.4.13. Method of Detecting AEs and SAEs

Electronic diaries have specifically been designed for this study by the Sponsor. The diaries will include prelisted AEs (solicited ARs) and intensity scales; they will also include blank space for the recording of information on other AEs (unsolicited AEs) and concomitant medications/vaccinations.

The investigator is responsible for the documentation of AEs regardless of study arm or suspected causal relationship to IP. For all AEs, the investigator must pursue and obtain information adequate to determine the outcome of the AE and to assess whether the AE meets the criteria for classification as an SAE requiring immediate notification to the Sponsor or its designated representative.

Care will be taken not to introduce bias when detecting AEs and/or SAEs. Open-ended and nonleading verbal questioning of the participant is the preferred method to inquire about AE occurrences.

###### 7.4.14. Follow-up of AEs and SAEs

After the initial AE/SAE report, the investigator is required to proactively follow each participant at subsequent visits and contacts.

All AEs and SAEs will be treated as medically appropriate and followed until resolution, stabilization, the event is otherwise explained, or the participant is LTFU, as defined in Section 6.3.

###### 7.4.15. Regulatory Reporting Requirements for SAEs

- Prompt notification by the investigator to the Sponsor of an SAE is essential so that legal obligations and ethical responsibilities toward the safety of participants and the safety of a study intervention under clinical investigation are met.
- The Sponsor has a legal responsibility to notify both the local regulatory authority and other regulatory agencies about the safety of a study intervention under clinical investigation. The Sponsor will comply with country-specific regulatory requirements relating to safety reporting to the regulatory authority, IRBs, and investigators.
- Investigator safety reports must be prepared for suspected unexpected serious ARs according to local regulatory requirements and Sponsor policy and forwarded to investigators as necessary.
- An investigator who receives an investigator safety report describing an SAE or other specific safety information (eg, summary or listing of SAEs) from the Sponsor will review and then file it along with the IB and will notify the IRB, if appropriate according to local requirements.

#### 7.5. Safety Monitoring

No safety monitoring committee or data safety monitoring board is planned for this study.

Safety monitoring for this study will include study team members, inclusive of, at a minimum, the Sponsor medical monitor, Sponsor safety physician (from Pharmacovigilance), and CRO medical monitor. The study team will conduct ongoing safety reviews during the study and will be responsible to monitor for safety concerns during the study as described in the Safety Management Plan.

An Independent CEAC that includes pediatric and adult cardiologists will review suspected cases of myocarditis and pericarditis to determine if they meet CDC criteria of "probable" or "confirmed" events, and to assess severity.

#### 7.6. Treatment of Overdose

As the study treatment is to be administered by a healthcare professional, it is unlikely that an overdose will occur. Dose deviations will be tracked as protocol deviations (Section 10.2.8).

#### 7.7. Pharmacokinetics

Pharmacokinetic parameters are not evaluated in this study.

##### 7.8. Pharmacodynamics

Pharmacodynamic parameters are not evaluated in this study.

#### 7.9. Biomarkers

Immunogenicity assessments are described in Section 7.2. Biomarkers are not evaluated in this study.

#### 7.10. Health Economics

Health economics are not evaluated in this study.

### 8. STATISTICAL ANALYSIS PLAN

This section summarizes the planned statistical analysis strategy and procedures for the study. The details of statistical analysis will be provided in the SAP, which will be finalized before the clinical database lock for the study. If changes are made to primary and/or secondary objectives or the related statistical methods after the study has begun, then the protocol will be amended (consistent with ICH Guideline E9). Changes to other secondary or exploratory analyses made after the protocol has been finalized, along with an explanation as to when and why they occurred, will be listed in the SAP or CSR for the study. Ad hoc exploratory analyses, if any, will be clearly identified in the CSR.

#### 8.1. Blinding and Responsibility for Analyses

This is an open-label study. For Part J, participants will be randomized in a 1:1 ratio to receive one of 2 treatments. Randomization will not be blinded.

#### 8.2. Statistical Hypotheses

**Part A.1:**

Each dose level of mRNA-1273.211 (50 µg, 100 µg) will be assessed with respect to mRNA-1273 primary series historical control.

For the primary objective on immune response, there are 4 null hypotheses to be tested for each arm:

A. $H_0^1$: 50 or 100 µg mRNA-1273.211, as a single booster dose, is inferior to the primary series of (100 µg) mRNA-1273 based on GMT ratio against ancestral SARS-CoV-2 with a non-inferiority margin of 1.5.

B. $H_0^2$: 50 or 100 µg mRNA-1273.211, as a single booster dose, is inferior to the primary series of (100 µg) mRNA-1273 based on difference in SRR against ancestral SARS-CoV-2 with a non-inferiority margin of 10%.

C. $H_0^3$: 50 or 100 µg mRNA-1273.211 against the variant B.1.351 is inferior to the primary series of (100 µg) mRNA-1273 against ancestral SARS-CoV-2 based on the GMT ratio of mRNA-1273.211 against the variant B.1.351 at Day 29 compared to mRNA-1273 against ancestral SARS-CoV-2 at Day 57 with a non-inferiority margin of 1.5.

D. $H_0^4$: 50 or 100 µg mRNA-1273.211 against the variant B.1.351 is inferior to the primary series of (100 µg) mRNA-1273 against ancestral SARS-CoV-2 based on the difference in SRR (SRR of mRNA-1273.211 against the variant B.1.351 at Day 29 – SRR of mRNA-1273 against ancestral SARS-CoV-2 at Day 57) with a non-inferiority margin of 10%.

**Part A.2:**

No statistical hypothesis testing will be performed for Part A.2.

**Part B:**

For the primary objective on immune response for a single booster dose of 100 µg mRNA-1273, there are 2 null hypotheses to be tested, and Part B would be considered to meet its primary objective if both null hypotheses are rejected:

A. $H_0^1$: 100 µg mRNA-1273, as a single booster dose, is inferior to the primary series of (100 µg) mRNA-1273 based on GMT ratio against ancestral SARS-CoV-2 with a non-inferiority margin of 1.5.

B. $H_0^2$: 100 µg mRNA-1273, as a single booster dose, is inferior to the primary series of mRNA-1273 based on difference in SRR against ancestral SARS-CoV-2 with a non-inferiority margin of 10%.

**Part C:**

Each dose level of mRNA-1273.617.2 booster dose (50 µg, 100 µg) will be assessed with respect to mRNA-1273 primary series historical control.

For the primary objective on immune response, there are 2 null hypotheses to be tested for each arm:

A. $H_0^1$: 50 µg or 100 µg mRNA-1273.617.2, as a single booster dose, against the variant B.1.617.2 is inferior to the primary series of (100 µg) mRNA-1273 against ancestral SARS-CoV-2 based on the GMT ratio of mRNA-1273.617.2 against the variant B.1.617.2 at Day 29 compared to mRNA-1273 against ancestral SARS-CoV-2 at Day 57 with a non-inferiority margin of 1.5.

B. $H_0^2$: 50 µg or 100 µg mRNA-1273.617.2, as a single booster dose, against the variant B.1.617.2 is inferior to the primary series of (100 µg) mRNA-1273 against ancestral SARS-CoV-2 based on the difference in SRR (SRR of mRNA-1273.617.2 against the variant B.1.617.2 at Day 29 – SRR of mRNA-1273 against ancestral SARS-CoV-2 at Day 57) with a non-inferiority margin of 10%.

**Part D:**

Each dose level of mRNA-1273.213 booster dose (50 µg, 100 µg) will be assessed with respect to mRNA-1273 primary series historical control.

For the primary objective on immune response, there are 4 null hypotheses to be tested for each arm:

A. $H_0^1$: 50 µg or 100 µg mRNA-1273.213, as a single booster dose, against the variant B.1.617.2 is inferior to the primary series of (100 µg) mRNA-1273 against ancestral SARS-CoV-2 based on the GMT ratio of mRNA-1273.213 against the variant B.1.617.2 at Day 29 compared to mRNA-1273 against ancestral SARS-CoV-2 at Day 57 with a non-inferiority margin of 1.5.

B. $H_0^2$: 50 µg or 100 µg mRNA-1273.213, as a single booster dose, against the variant B.1.617.2 is inferior to the primary series of (100 µg) mRNA-1273 against ancestral SARS-CoV-2 based on the difference in SRR (SRR of mRNA-1273.213 against the variant B.1.617.2 at Day 29 – SRR of mRNA-1273 against ancestral SARS-CoV-2 at Day 57) with a non-inferiority margin of 10%.

C. $H_0^3$: 50 µg or 100 µg mRNA-1273.213, as a single booster dose, against the variant B.1.351 is inferior to the primary series of (100 µg) mRNA-1273 against ancestral SARS-CoV-2 based on the GMT ratio of mRNA-1273.213 against the variant B.1.351 at Day 29 compared to mRNA-1273 against ancestral SARS-CoV-2 at Day 57 with a non-inferiority margin of 1.5.

D. $H_0^4$: 50 µg 100 µg mRNA-1273.213, as a single booster dose, against the variant B.1.351 is inferior to the primary series of (100 µg) mRNA-1273 against ancestral SARS-CoV-2 based on the difference in SRR (SRR of mRNA-1273.213 against the variant B.1.351 at Day 29 – SRR of mRNA-1273 against ancestral SARS-CoV-2 at Day 57) with a non-inferiority margin of 10%.

**Part F (Cohort 1):**

50 µg mRNA-1273.529 booster dose (as a first booster dose) will be assessed with respect to mRNA-1273 booster dose (as a first booster dose) using an external comparator (details will be provided in SAP).

For the primary immunogenicity objectives, there are 3 hypotheses to be tested:

A. $H_1^1$: 50 µg mRNA-1273.529, as a single booster dose, against the variant B.1.1.529 is non-inferior to the booster dose of (50 µg) mRNA-1273 against B.1.1.529 based on the GMT ratio of mRNA-1273.529 against the variant B.1.1.529 at Day 29 compared to mRNA-1273 against the variant B.1.1.529 at Day 29 with a non-inferiority margin of 1.5.

B. $H_1^2$: 50 µg mRNA-1273.529, as a single booster dose, against the variant B.1.1.529 is non-inferior to the booster dose of (50 µg) mRNA-1273 against B.1.1.529 based on the difference in SRR at Day 29 with a non-inferiority margin of 10%.

C. $H_1^3$: 50 µg mRNA-1273.529, as a single booster dose, against the variant B.1.1.529 is superior to the booster dose of (50 µg) mRNA-1273 against B.1.1.529 based on the GMT ratio of mRNA-1273.529 against the variant B.1.1.529 at Day 29 compared to mRNA-1273 against the variant B.1.1.529 at Day 29.

The primary immunogenicity objective is considered met if non-inferiority against B.1.1.529 based on GMR and SRR difference is demonstrated.

**Part F (Cohort 2):**

50 µg mRNA-1273.529 as the second booster dose will be compared to 50 µg mRNA-1273 as the second booster dose

For the primary immunogenicity objective, there are 3 hypotheses to be tested:

A. $H_1^1$: 50 µg mRNA-1273.529, as a second booster dose, against the variant B.1.1.529 is non-inferior to the second booster dose of (50 µg) mRNA-1273 against B.1.1.529 based on the GMT ratio of mRNA-1273.529 against the variant B.1.1.529 at Day 29 compared to mRNA-1273 against the variant B.1.1.529 at Day 29 with a non-inferiority margin of 1.5.

B. $H_1^2$: 50 µg mRNA-1273.529, as a second booster dose, against the variant B.1.1.529 is non-inferior to the second booster dose of (50 µg) mRNA-1273 against B.1.1.529 based on the difference in SRR at Day 29 with a non-inferiority margin of 10%.

C. $H_1^3$: 50 µg mRNA-1273.529, as a second booster dose, against the variant B.1.1.529 is superior to the second booster dose of (50 µg) mRNA-1273 against B.1.1.529 based on the GMT ratio of mRNA-1273.529 against the variant B.1.1.529 at Day 29 compared to mRNA-1273 against the variant B.1.1.529 at Day 29.

The primary immunogenicity objective is considered met if non-inferiority against B.1.1.529 based on GMR and SRR difference is demonstrated.

**Part G:**

50 µg mRNA-1273.214 as the second booster dose will be compared to 50 µg mRNA-1273 as the second booster dose (active control arm in Part F, Cohort 2)

For the primary objective on immune response, there are 8 hypotheses (4 hypotheses at Day 29 and 4 hypotheses at Day 91).

A. $H_1^1$: 50 µg mRNA-1273.214, as a second booster dose, against the variant B.1.1.529 is non-inferior to the second booster dose of (50 µg) mRNA-1273 against B.1.1.529 based on the GMT ratio of mRNA-1273.214 against the variant B.1.1.529 at Day 29 compared to mRNA-1273 against the variant B.1.1.529 at Day 29 with a non-inferiority margin of 1.5.

B. $H_1^2$: 50 µg mRNA-1273.214, as a second booster dose, against the variant B.1.1.529 is non-inferior to the second booster dose of (50 µg) mRNA-1273 against B.1.1.529 based on the difference in SRR at Day 29 with a non-inferiority margin of 10%.

C. $H_1^3$: 50 µg mRNA-1273.214, as a second booster dose, against ancestral SARS-CoV-2 is non-inferior to the second booster dose of (50 µg) mRNA-1273 against ancestral SARS-CoV-2 based on the GMT ratio of mRNA-1273.214 against the ancestral SARS-CoV-2 at Day 29 compared to mRNA-1273 against the ancestral SARS-CoV-2 at Day 29 with a non-inferiority margin of 1.5.

D. $H_1^4$: 50 µg mRNA-1273.214, as a second booster dose, against the variant B.1.1.529 is superior to the second booster dose of (50 µg) mRNA-1273 against B.1.1.529 based on the GMT ratio of mRNA-1273.214 against the variant B.1.1.529 at Day 29 compared to mRNA-1273 against the variant B.1.1.529 at Day 29.

E. $H_1^5$: 50 µg mRNA-1273.214, as a second booster dose, against the variant B.1.1.529 is non-inferior to the second booster dose of (50 µg) mRNA-1273 against B.1.1.529 based on the GMT ratio of mRNA-1273.214 against the variant B.1.1.529 at Day 91 compared to mRNA-1273 against the variant B.1.1.529 at Day 91 with a non-inferiority margin of 1.5.

F. $H_1^6$: 50 µg mRNA-1273.214, as a second booster dose, against the variant B.1.1.529 is non-inferior to the second booster dose of (50 µg) mRNA-1273 against B.1.1.529 based on the difference in SRR at Day 91 with a non-inferiority margin of 10%.

G. $H_1^7$: 50 µg mRNA-1273.214, as a second booster dose, against ancestral SARS-CoV-2 is non-inferior to the second booster dose of (50 µg) mRNA-1273 against ancestral SARS-CoV-2 based on the GMT ratio of mRNA-1273.214 against ancestral SARS-CoV-2 at Day 91 compared to mRNA-1273 against ancestral SARS-CoV-2 at Day 91 with a non-inferiority margin of 1.5

H. $H_1^8$: 50 µg mRNA-1273.214, as a second booster dose, against the variant B.1.1.529 is superior to the second booster dose of (50 µg) mRNA-1273 against B.1.1.529 based on the GMT ratio of mRNA-1273.214 against the variant B.1.1.529 at Day 91 compared to mRNA-1273 against the variant B.1.1.529 at Day 91.

For the primary immunogenicity objective, an alpha of 0.05 (two-sided) will be allocated to the 2 time points (Day 29 and Day 91). Day 29 and Day 91 will each have an alpha of 0.025 (two-sided) for hypotheses testing.

The primary immunogenicity objective is considered met if non-inferiority against B.1.1.529 based on GMR, SRR difference. and non-inferiority against ancestral SARS-CoV-2 based on GMR are demonstrated at Day 29 or Day 91.

For the key secondary immunogenicity objective, there are 2 hypotheses to be tested (Day 29 and Day 91 will each have alpha of 0.025 [two-sided] for hypotheses testing):

I. $H_1^9$: 50 µg mRNA-1273.214, as a second booster dose, against ancestral SARS-CoV-2 is non-inferior to the booster dose of (50 µg) mRNA-1273 against ancestral SARS-CoV-2 based on the difference in SRR at Day 29 with a non-inferiority margin of 10%.

J. $H_1^{10}$: 50 µg mRNA-1273.214, as a second booster dose, against ancestral SARS-CoV-2 is non-inferior to the booster dose of (50 µg) mRNA-1273 against ancestral SARS-CoV-2 based on the difference in SRR at Day 91 with a non-inferiority margin of 10%.

**Part H:**

50 µg mRNA-1273.222 given as a second booster dose will be compared to 50 µg mRNA-1273 given as a second booster dose (in Part F, Cohort 2)

For the primary immunogenicity objective, there are 5 hypotheses to be tested as shown in Figure 2:

A. $H_1^1$: 50 µg mRNA-1273.222, as a second booster dose, against Omicron BA.4/5 is non-inferior to the second booster dose of (50 µg) mRNA-1273 against Omicron BA.4/5 based on the GMT ratio of mRNA-1273.222 against Omicron BA.4/5 at Day 29 compared to mRNA-1273 against Omicron BA.4/5 at Day 29 with a non-inferiority margin of 1.5.

B. $H_1^2$: 50 µg mRNA-1273.222, as a second booster dose, against Omicron BA.4/5 is non-inferior to the second booster dose of (50 µg) mRNA-1273 against Omicron BA.4/5 based on the difference in SRR at Day 29 with a non-inferiority margin of 5%.

C. $H_1^3$: 50 µg mRNA-1273.222, as a second booster dose, against ancestral SARS-CoV-2 D614G is non-inferior to the second booster dose of (50 µg) mRNA-1273 against ancestral SARS-CoV-2 D614G based on the GMT ratio of mRNA-1273.222 against ancestral SARS-CoV-2 D614G at Day 29 compared to mRNA-1273 against ancestral SARS-CoV-2 D614G at Day 29 with a non-inferiority margin of 1.5.

D. $H_1^4$: 50 µg mRNA-1273.222, as a second booster dose, against ancestral SARS-CoV-2 D614G is non-inferior to the second booster dose of (50 µg) mRNA-1273 against ancestral SARS-CoV-2 D614G based on the difference in SRR at Day 29 with a non-inferiority margin of 10%.

E. $H_1^5$: 50 µg mRNA-1273.222, as a second booster dose, against Omicron BA.4/5 is superior to the second booster dose of (50 µg) mRNA-1273 against Omicron BA.4/5 based on the GMT ratio of mRNA-1273.222 against Omicron BA.4/5 at Day 29 compared to mRNA-1273 against Omicron BA.4/5 at Day 29.

**Part J:**

No formal hypothesis testing will be performed for Part J and all safety and immunogenicity analyses will be descriptive.

The primary immunogenicity objective will be assessed using the Per-Protocol Set for Immunogenicity. Comparisons on immunogenicity response between the mRNA-1273.815 and mRNA-1273.231 treatment arms may be performed.

An ANCOVA model will be used to compare antibody response between treatment arms. Day 15 and Day 29 antibody titers (against SARS-CoV-2 variants) will be included in the model as a dependent variable and treatment group variables (mRNA-1273.815 and mRNA-1273.231) will be included as fixed effect. The model may adjust for other covariates as appropriate. Geometric mean ratio and the corresponding 95% CI will be estimated from the model. Seroresponse rate difference at Day 15 and Day 29 will be compared and 95% CI will be provided using Miettinen-Nurminen method.

#### 8.3. Sample Size Determination

**Part A.1:**

Each dose level of mRNA-1273.211 will be assessed at a 2-sided type I error rate of 5%.

The target enrollment of the 50-µg mRNA-1273.211 arm is approximately 300 participants. Assuming 10% of participants will be excluded from the PP Set for Immunogenicity, with approximately 270 participants in the mRNA-1273.211 50 µg study arm and from the mRNA-1273 primary series historical control arm in the PP Set for Immunogenicity, there is approximately 75% power to reject all null hypotheses for the primary objectives. With approximately 270 participants in the mRNA-1273.211 50 µg study arm and 526 participants from the mRNA-1273 primary series historical control arm, there is approximately 90% power to reject all null hypotheses for the primary objectives based on GMR and the difference in SRR against ancestral SARS-CoV-2 and variant (B.1.351) virus strain at 2-sided alpha of 5.0%. The assumptions are: the true GMR (50 µg mRNA-1273.211 vs. 100 µg mRNA-1273) against ancestral SARS-CoV-2 and variant virus strain is 1, the standard deviation of the log-transformed titer is 1.5, with a non-inferiority margin of 1.5; the true SRR against ancestral SARS-CoV-2 and variant virus strain after a single booster dose of mRNA-1273.211 is 90%, SRR against ancestral SARS-CoV-2 after mRNA-1273 primary series is 90%, and non-inferiority margin for SRR difference is 10%.

The target enrollment of 100 µg mRNA-1273.211 arm is approximately 584 participants. Assuming 10% of participants will be excluded from the PP Set for Immunogenicity, with approximately 526 participants in mRNA-1273.211 100 µg and mRNA-1273 primary series historical control arm respectively in the PP Set for Immunogenicity, there is approximately 80%

power to reject all null hypotheses for the primary objectives based on GMR and difference in SRR against ancestral SARS-CoV-2 at 2-sided alpha of 5.0%. The assumptions are: the true GMR against ancestral SARS-CoV-2 and the variant (B.1.351) is 0.9, the standard deviation of the log-transformed titer is 1.5, non-inferiority margin for GMR is 1.5; the true SRR against ancestral SARS-CoV-2 and variant after a single booster dose of mRNA-1273.211 is 90%, and SRR against ancestral SARS-CoV-2 and variant after mRNA-1273 primary series is 90%, and non-inferiority margin for SRR difference is 10%.

Approximately 584 participants in the mRNA-1273-P301 (COVE) study will be selected as the mRNA-1273 primary series historical control. Summary of the sampling plan for the selection of mRNA-1273-P301 (COVE) participant is in Section 8.5.3.2 and in the SAP. The same historical comparator arm will be used for the subsequent Parts B, C, and D.

With approximately 300 and 584 participants exposed to 50 and 100 µg of mRNA-1273.211, respectively, there is at least 90% probability to observe one participant at each dose level reporting an AE if the true rate of AEs is 1%.

**Part A.2**

We anticipate approximately 300 participants will be enrolled in Part A.2, there is no statistical hypothesis testing in Part A.2.

**Part B:**

The target enrollment of the 100 µg mRNA-1273 arm is approximately 300 participants. Assuming 10% of participants will be excluded from the PP Set for Immunogenicity, with approximately 270 participants in the 100 µg mRNA-1273 study arm and approximately 526 from the mRNA-1273 primary series historical control arm in the PP Set for Immunogenicity, there is >90% power to reject both null hypotheses for the primary objective based on GMR and the difference in SRR against ancestral SARS-CoV-2 (mRNA-1273) at 2-sided alpha of 5.0%. The assumptions are: the true GMR (100 µg mRNA-1273 booster vs. 100 µg mRNA-1273 primary series historical control) against ancestral SARS-CoV-2 is 1.5, the standard deviation of the log-transformed titer is 1.5, with a non-inferiority margin of 1.5; the true SRR against ancestral SARS-CoV-2 after a single booster dose of mRNA-1273 is 90%, SRR against ancestral SARS-CoV-2 after mRNA-1273 primary series is 90%, and non-inferiority margin for SRR difference is 10%.

With approximately 300 participants exposed to 100 µg of mRNA-1273, there is at least 90% probability to observe one participant reporting an AE if the true rate of AEs is 1%.

**Part C**

Each dose level of mRNA-1273.617.2 will be assessed at a 2-sided type I error rate of 5%.

The target enrollment is approximately 584 participants for each dose level of mRNA-1273.617.2. Assuming 10% of participants will be excluded from the PP Set for Immunogenicity, with approximately 526 participants in each dose level of mRNA-1273.617.2 and mRNA-1273 primary series historical control arm respectively in the PP Set for Immunogenicity, there is approximately 90% power to reject both null hypotheses for the primary objectives based on GMR and difference in SRR. The assumptions are: the true GMR (mRNA-1273.617.2 booster vs. 100 mRNA-1273 primary series historical control) against the variant (B.1.617.2) is 0.9 compared to 100 µg mRNA-1273 against ancestral SARS-CoV-2, the standard deviation of the log-transformed titer is 1.5, non-inferiority margin for GMR is 1.5; the true SRR against variant after a single booster dose of mRNA-1273.617.2 is 90%, and SRR against ancestral SARS-CoV-2 and variant after mRNA-1273 primary series is 90%, and non-inferiority margin for SRR difference is 10%. The immune response assumptions are the same for 50 µg and 100 µg mRNA-1273.617.2.

With approximately 584 participants exposed to each dose of mRNA-1273.617.2, there is at least 90% probability in each group to observe one participant reporting an AE if the true rate of AEs is 1%.

**Part D**

Each dose level of mRNA-1273.213 will be assessed at a 2-sided type I error rate of 5%.

The target enrollment is approximately 584 participants for each dose level of mRNA-1273.213. Assuming 10% of participants will be excluded from the PP Set for Immunogenicity, with approximately 526 participants in each dose level of mRNA-1273.213 and mRNA-1273 primary series historical control arm respectively in the PP Set for Immunogenicity, there is approximately 80% power to reject all null hypotheses for the primary objectives based on GMR and difference in SRR. The assumptions are: the true GMR (mRNA-1273.213 vs. 100 µg mRNA-1273 primary series historical control) against the 2 variants (B.1.617.2, B.1.351) is 0.9 compared to 100 ug mRNA-1273 against ancestral SARS-CoV-2, the standard deviation of the log-transformed titer is 1.5, non-inferiority margin for GMR is 1.5; the true SRR against variants (B.1.617.2, B.1.351) after a single booster dose of mRNA-1273.213 is 90%, and SRR against ancestral SARS-CoV-2 and variant after mRNA-1273 primary series is 90%, and non-inferiority margin for SRR difference is 10%. The immune response assumptions are the same for 50 µg and 100 µg mRNA-1273.617.2.

With approximately 584 participants exposed to each dose of mRNA-1273.213, there is at least 90% probability in each group to observe one participant reporting an AE if the true rate of AEs is 1%.

**Part F**

mRNA-1273.529 in each cohort will be assessed at a 2-sided type I error rate of 5%.

**Cohort 1:**

The target enrollment is approximately 375 participants for 50 µg mRNA-1273.529. Assuming 20% of participants will be excluded from the PP Set for Immunogenicity-SARS-CoV2 negative, with approximately 300 participants in 50 µg mRNA-1273.529 and 300 participants in 50 µg mRNA-1273 (external comparator) in the PP Set for Immunogenicity-SARS-CoV-2 negative, there is approximately 89% global power for the primary immunogenicity objectives with alpha level of 0.05 (2-sided). The assumptions are: the true GMR (mRNA-1273.529 booster vs. 50 µg mRNA-1273 booster) against the variant (B.1.1.529) is 1.5, the standard deviation of the log-transformed titer is 1.5, and the non-inferiority margin for GMR is 1.5; the true SRR against B.1.1.529 after a single booster dose of 50 µg mRNA-1273.529 is 90% (same assumption for 50 µg mRNA-1273), and non-inferiority margin for SRR difference is 10%.

With approximately 375 participants exposed to 50 µg mRNA-1273.529, there is at least 90% probability in this group to observe 1 participant reporting an AE if the true rate of AEs is 1%.

There may be an urgency to perform the Day 29 analysis as early as possible and depending on the testing capability of assays of antibodies against B.1.1.529, the Sponsor may decide using an external arm with less than 300 participants. Such decision will be documented in SAP prior to the planned Day 29 analysis.

**Cohort 2:**

The target enrollment is approximately 750 participants for 50 µg mRNA-1273.529 and 50 µg mRNA-1273 (1:1 ratio). Assuming 20% of participants will be excluded from the PP Set for Immunogenicity – SARS-CoV-2 negative, with approximately 300 participants each in 50 µg mRNA-1273.529 and 50 µg mRNA-1273 in the PP Set for Immunogenicity and SARS-CoV-2 negative, there is approximately 89% global power to demonstrate the primary immunogenicity objectives of alpha level of 0.05 (2-sided). The assumptions are: the true GMR (mRNA-1273.529 as the second booster vs 50 µg mRNA-1273 as the second booster) against the variant (B.1.1.529) is 1.5, the standard deviation of the log-transformed titer is 1.5, and the non-inferiority margin for GMR is 1.5; the true SRR against B.1.1.529 after 50 µg mRNA-1273.529 as a second booster dose is 90% (same assumption for 50 µg 1273), and non-inferiority margin for SRR difference is 10%.

With approximately 375 participants exposed to each group, there is at least 90% probability in each group to observe 1 participant reporting an AE if the true rate of AEs is 1%.

**Part G**

The target enrollment is approximately 375 participants for 50 µg mRNA-1273.214. Hypotheses testing will be performed at Day 29 and Day 91, alpha of 0.025 (2-sided) will be allocated equally to each one of the 2 time points. Assuming 20% of participants will be excluded from the PP Set for Immunogenicity – SARS-CoV-2 negative, with approximately 300 participants in 50 µg mRNA-1273.214 and 300 participants in 50 µg mRNA-1273 (Part F, Cohort 2, 50 µg mRNA-1273) in the PP Set for Immunogenicity and SARS-CoV-2 negative, there is approximately 71% global power to demonstrate the primary immunogenicity objectives with alpha of 0.025 (2-sided) at each time point. The assumptions are: the true GMR (mRNA-1273.214 second booster vs. 50 µg mRNA-1273 second booster) against the variant (B.1.1.529) is 1.5, the true GMR against ancestral SARS-CoV-2 is 1, the standard deviation of the log-transformed titer is 1.5, and the non-inferiority margin for GMR is 1.5, the true SRR against B.1.1.529 after mRNA-1273.214 as a second booster dose is 90% (same assumption for both 50 µg mRNA-1273.214 and 50 µg mRNA-1273), and non-inferiority margin for SRR difference is 10%.

With approximately 375 participants exposed to 50 µg mRNA-1273.214, there is at least 90% probability in this group to observe 1 participant reporting an AE if the true rate of AEs is 1%.

**Part H**

The target enrollment is approximately 500 participants for 50 µg mRNA-1273.222. Assuming 40% of participants will be excluded from the PP Set for Immunogenicity – SARS-CoV-2 negative (due to a SARS-CoV-2 infection prebooster), with approximately 300 participants in 50 µg mRNA-1273.222 and 260 participants in 50 µg mRNA-1273 (Part F, Cohort 2-50 µg mRNA-1273) in the PP Set for Immunogenicity and SARS-CoV-2 negative, there is approximately 60% power to demonstrate the primary immunogenicity objectives with an alpha of 0.05 (2-sided) at

Day 29. The assumptions are: the true GMR (mRNA-1273.222 second booster vs. mRNA-1273 second booster) against Omicron BA.4/5 is 1.5, GMR (mRNA-1273.222 second booster vs. mRNA-1273 second booster) against ancestral SARS-CoV-2 D614G is 1, the standard deviation of the log-transformed titer is 1.5, and the non-inferiority margin for GMR is 1.5. The true SRR against Omicron BA.4/5 after mRNA-1273.222 as a second booster dose is 95% (same assumption for 50 µg mRNA-1273), and non-inferiority margin for SRR difference against Omicron BA.4/5 is 5%. The true SRR against ancestral SARS-CoV-2 D614G after mRNA-1273.222 as a second booster dose is 95% (same assumption for 50 µg mRNA-1273), and the non-inferiority margin for SRR difference against ancestral SARS-CoV-2 D614G is 10%.

With approximately 500 participants exposed to 50 µg mRNA-1273.222, there is at least 90% probability in this group to observe 1 participant reporting an AE if the true rate of AEs is 1%.

**Part J**

The sample size for Part J is not driven by statistical assumptions for formal hypothesis testing. The number of proposed participants is considered sufficient to provide a descriptive summary of the safety and immunogenicity of each treatment arm.

The target enrollment is approximately 100 participants who will be randomized in a 1:1 ratio to receive either mRNA-1273.815 (50 µg) or mRNA-1273.231 (50 µg).

#### 8.4. Analysis Sets

The analysis sets are described in Table 15, same definitions across Parts A (1, 2), B, C, D, F, G, H, and J when applicable.

**Table 15: Analysis Sets**

| Set | Description |
|---|---|
| Full Analysis Set (FAS) | The FAS consists of all participants who receive IP. |
| Modified Intent-to-Treat (mITT) Set | The mITT Set consists of all participants in the FAS who have no serologic or virologic evidence of prior SARS-CoV-2 infection (both negative RT-PCR test for SARS-CoV-2 and negative serology test based on bAb specific to SARS-CoV-2 nucleocapsid) prebooster, ie, all FAS participants with baseline SARS-CoV-2 negative status at prebooster. |
| Per-Protocol (PP) Set for Immunogenicity | The PP Set for Immunogenicity consists of all participants in the FAS who received the planned dose of study vaccination and no major protocol deviations that impact key or critical data.<br>The PP Set will be used as the primary analysis set for analyses of immunogenicity for immunobridging. |
| PP Set for Immunogenicity - SARS-CoV-2 negative (PPSI-Neg) | Participants in the PPSI who have no serologic or virologic evidence of SARS-CoV-2 infection at baseline, ie, who are SARS-CoV-2 negative, defined by both negative RT-PCR test for SARS-CoV-2 and negative serology test based on bAb specific to SARS-CoV-2 nucleocapsid |

| Set | Description |
|---|---|
| | PPSI-Neg will be the primary analysis set for analyses of immunogenicity for between booster comparisons. |
| Solicited Safety Set | The Solicited Safety Set consists of all participants who receive IP and contribute any solicited AR data.<br>The Solicited Safety Set will be used for the analyses of solicited ARs. Participants will be included in the study arm corresponding to the dose of IP that they actually received. |
| Safety Set | The Safety Set consists of all participants who receive IP. The Safety Set will be used for all analyses of safety except for the solicited ARs. Participants will be included in the study arm corresponding to the dose of IP that they actually received. |
| Per-Protocol Set for Efficacy | The PP Set for Efficacy consists of all participants in the FAS who receive the planned dose of study vaccination, who are SARS-CoV-2 negative at baseline (ie, have a negative RT-PCR test for SARS-CoV-2 and a negative serology test based on bAb specific to SARS-CoV-2 nucleocapsid at baseline), and have no major protocol deviations that impact key or critical data. |

#### 8.5. Statistical Methods

##### 8.5.1. Baseline Characteristics and Demographics

Demographic variables (eg, age, gender, race, ethnicity, height, weight, and body mass index) and baseline characteristics will be summarized by study arm. For study parts with more than one dose, data will be summarized for each arm and for dose levels of the same vaccine type combined. Summary statistics (mean, standard deviation for continuous variable, and number and percentage for categorical variables) will be provided.

##### 8.5.2. Efficacy Analysis

Descriptive summaries of symptomatic COVID-19 disease, asymptomatic SARS-CoV-2 infection, as well as COVID-19 regardless of symptoms will be provided for each study arm. Vaccine efficacy may be estimated if the number of cases accrued is deemed to be sufficient. Efficacy analyses will be performed using the mITT and PP Set for Immunogenicity.

##### 8.5.3. Immunogenicity Analysis

The primary analysis population for immunogenicity will be the PP Set for Immunogenicity for immunobridging analysis, PPSI-Neg will be the primary analysis for immunogenicity analyses for between booster group comparisons. Each arm will be evaluated separately.

###### 8.5.3.1. Sampling of Historical Control: Participants from Study mRNA-1273-P301 Immunogenicity Analysis

Approximately 584 Study mRNA-1273-P301 participants on primary series of mRNA-1273 will be selected to serve as the mRNA-1273 comparator (historical control arm), and immunogenicity data at Day 57 after the primary series of mRNA-1273 from these participants will be used to be compared with those at Day 29 after a booster dose this study.

In Study mRNA-1273-P301, a random subcohort of study participants have already been selected for the secondary immunogenicity objective, including approximately 1,000 participants who were baseline SARS-CoV-2 negative and randomized to mRNA-1273. The PP Random Subcohort for Immunogenicity is the primary analysis population for the secondary immunogenicity objective in mRNA-1273-P301 (mRNA-1273-P301 SAP v2.0).

In Study mRNA-1273-P301, approximately 75% participants were ≥18 and <65 years old and 25% participants were ≥ 65 years old. In Study P205, for each study arm, the plan is to enroll participants with similar distribution of the 2 age groups (approximately 75% ≥ 18 and <65 years old and approximately 25% participants were ≥ 65 years old).

Therefore, a total of approximately 584 mRNA-1273-P301 participants with baseline SARS-CoV-2 negative status and randomized to mRNA-1273 with similar distribution of the 2 age groups will be selected from the mRNA-1273-P301 PP Random Subcohort as historical control for Study P205. Within each age group, mRNA-1273-P301 participants with baseline SARS-CoV-2 negative status and randomized to mRNA-1273 in the mRNA-1273-P301 PP Random Subcohort will be randomly selected.

If there is not enough number of participants in the mRNA-1273-P301 PP Random Subcohort either for an age group or overall, additional participants will be randomly selected from mRNA-1273-P301 who were not selected for the PP Random Subcohort. If the planned number of target sample size in P205 is changed, the number of participants from mRNA-1273-P301 to serve as historical control will be updated accordingly.

##### 8.5.3.2. Analysis for the Primary Immunogenicity Objective

###### 8.5.3.2.1. Analysis for the Primary Immunogenicity Objective for Part A.1

Each dose level of mRNA-1273.211 (50 and 100 µg) will be assessed with respect to mRNA-1273 primary series historical control.

For the primary objective on immune response, there are 4 null hypotheses to be tested for each arm.

A. $H_0^1$: 50 or 100 µg mRNA-1273.211, as a single booster dose, is inferior to the primary series of (100 µg) mRNA-1273 based on GMT ratio against ancestral SARS-CoV-2 with a non-inferiority margin of 1.5.

B. $H_0^2$: 50 or 100 µg mRNA-1273.211, as a single booster dose, is inferior to the primary series of (100 µg) mRNA-1273 based on difference in SRR against ancestral SARS-CoV-2 with a non-inferiority margin of 10%.

C. $H_0^3$: 50 or 100 µg mRNA-1273.211 against the variant B.1.351 is inferior to the primary series of (100 µg) mRNA-1273 against ancestral SARS-CoV-2 based on the GMT ratio of mRNA-1273.211 against the variant B.1.351 at Day 29 compared to mRNA-1273 against ancestral SARS-CoV-2 at Day 57 with a non-inferiority margin of 1.5.

D. $H_0^4$: 50 or 100 µg mRNA-1273.211 against the variant B.1.351 is inferior to the primary series of (100 µg) mRNA-1273 against ancestral SARS-CoV-2 based on the difference in SRR (SRR of mRNA-1273.211 against the variant B.1.351 at Day 29 – SRR of mRNA-1273 against ancestral SARS-CoV-2 at Day 57) with a non-inferiority margin of 10%.

Pseudotyped virus neutralizing antibody will be used as the basis to assess non-inferiority in immune response. The assays that will be used to assess the immune response to vaccination will be described in the SAP.

Each dose level of mRNA-1273.211 will be assessed with respect to mRNA-1273 primary series historical control separately. An ANCOVA model will be carried out with antibody titers (Day 29 on 50 or 100 µg mRNA-1273.211 and Day 57 on mRNA-1273 primary series historical control) against ancestral SARS-CoV-2 as a dependent variable and a group variable (50 or 100 µg mRNA-1273.211 and mRNA-1273 primary series historical control) as the fixed effect, adjusting for age groups (<65, ≥ 65). The GMT will be estimated by the GLSM from the model for each group and corresponding 95% CI will be provided for each group. The GMR (ratio of GMTs) for each dose level of mRNA-1273.211 with respect to mRNA-1273 will be estimated by the ratio of GLSM from the model, the estimated between group differences along with the corresponding 95% CI. The 95% CI for the ratio of GLSM will be provided to assess the between group difference in immune response against ancestral SARS-CoV-2 for each dose level of mRNA-1273.211 at Day 29 compared to mRNA-1273 primary series historical control at Day 57.

For each dose level of mRNA-1273.211, the non-inferiority of immune response to mRNA-1273 will be considered demonstrated if the lower bound of the corresponding CI of the GMR against ancestral SARS-CoV-2 is > 0.667 based on the non-inferiority margin of 1.5.

The primary definition of seroresponse is defined as ≥ 4 × LLOQ for those with baseline < LLOQ; ≥ 4-foldrise for those with baseline ≥ LLOQ. This definition will be used as the basis for non-inferiority hypothesis testing. The number and percentage (rate) of participants achieving seroresponse at Day 29 will be summarized with 95% CI calculated using the Clopper-Pearson method for each group. The difference of SRRs between each dose level of mRNA-1273.211 at Day 29 and mRNA-1273 primary series historical control at Day 57 in mRNA-1273-P301 against ancestral SARS-CoV-2 will be calculated with 95% CI. The non-inferiority in SRR of each dose level of mRNA-1273.211 compared to mRNA-1273 primary series historical control will be considered demonstrated if the lower bound of the 95% of the SRR difference is > -10% based on the non-inferiority margin of 10%.

The same analysis methods for immune response against variant B.1.351 will be used for $H_0^3$ and $H_0^4$ hypotheses testing.

For each dose level of mRNA-1273.211, the primary immunogenicity objective (against ancestral SARS-CoV-2 mRNA-1273 and variant [B.1.351]) is considered met if non-inferiority is demonstrated based on both GMR and SRR difference, specifically:

- If the lower bound of the 95% CI of the GMT ratio against ancestral SARS-CoV-2 between mRNA-1273.211 and mRNA-1273 is > 0.667 based on the non-inferiority margin of 1.5, and
- The lower bound of the 95% CI of the SRR difference (mRNA-1273.211 – mRNA-1273) against ancestral SARS-CoV-2 strain is > -10%, and

- The lower bound of the 95% CI of the GMT ratio between mRNA-1273.211 against the variant (B.1.351) at Day 29 as compared to 100 µg mRNA-1273 against ancestral SARS-CoV-2 is at Day 57 > 0.667 based on the non-inferiority margin of 1.5, and
- The lower bound of the 95% CI of the SRR difference (mRNA-1273.211 against the variant [B.1.351] at Day 29 – 100 µg mRNA-1273 against ancestral SARS-CoV-2 strain at Day 57) is > -10%.

###### 8.5.3.2.2. Analysis for the Primary Immunogenicity Objective for Part A.2

There is no hypothesis testing for Part A.2.

For Part A.2 participants, Day 29 and Day 181 immune response after mRNA-1273.214 (50 µg) as a second booster dose will be compared with their own Day 29 and Day 181 immune response of mRNA-1273.211 (50 µg) received as the first booster dose. GMT ratios will be calculated by back transforming the mean of paired differences of antibody titer data on the logarithmic scale between Day 29 and Day 181 post mRNA-1273.214 and Day 29 and Day 181 of antibody titer data post mRNA-1273.211. CIs for the GMT ratio will be based on t-distribution of the log-transformed values then back-transformed to the original scale for presentation. Seroresponse rates at Day 29 and Day 181 post mRNA-1273.214 will be compared with their seroresponse rates at Day 29 and Day 181 post mRNA-1273.211. The difference in seroresponse rates and its corresponding 95% CI based on adjusted Wald method will be provided.

###### 8.5.3.2.3. Analysis for the Primary Immunogenicity Objective for Part B

For the primary objective on immune response for a single booster dose of 100 µg mRNA-1273, there are 2 null hypotheses to be tested, and Part B would be considered to meet its primary objective if both null hypotheses are rejected:

A. $H_0^1$: 100 µg mRNA-1273, as a single booster dose, is inferior to the primary series of (100 µg) mRNA-1273 based on GMT ratio against ancestral SARS-CoV-2 with a non-inferiority margin of 1.5.

B. $H_0^2$: 100 µg mRNA-1273, as a single booster dose, is inferior to the primary series of (100 µg) mRNA-1273 based on difference in SRR against ancestral SARS-CoV-2 with a non-inferiority margin of 10%.

The same analysis methods described in Section 8.5.3.2.1 will be used for the primary immunogenicity objective for Part B.

The primary immunogenicity objective (against ancestral SARS-CoV-2 mRNA-1273) is considered met if non-inferiority is demonstrated based on both GMR and SRR difference, specifically:

- The lower bound of the 95% CI of the GMT ratio against ancestral SARS-CoV-2 between 100 µg mRNA-1273 booster dose and 100 µg mRNA-1273 primary series is > 0.667 based on the non-inferiority margin of 1.5, and
- The lower bound of the 95% CI of the SRR difference (100 µg mRNA-1273 booster dose – 100 µg mRNA-1273 primary series) against ancestral SARS-CoV-2 strain is > -10%

###### 8.5.3.2.4. Analysis for the Primary Immunogenicity Objective for Part C

Each dose level of mRNA-1273.617.2 (50 and 100 µg) will be assessed with respect to mRNA-1273 primary series historical control separately. For the primary objective on immune response, there are 2 null hypotheses to be tested for each arm.

A. $H_0^1$: 50 µg or 100 µg mRNA-1273.617.2, as a single booster dose, against the variant B.1.617.2 is inferior to the primary series of (100 µg) mRNA-1273 against ancestral SARS-CoV-2 based on GMT ratio of mRNA-1273.617.2 against the variant B.1.617.2 at Day 29 compared to mRNA-1273 against ancestral SARS-CoV-2 at Day 57 with a non-inferiority margin of 1.5.

B. $H_0^2$: 50 µg or 100 µg mRNA-1273.617.2, as a single booster dose, against the variant B.1.617.2 is inferior to the primary series of (100 µg) mRNA-1273 against ancestral SARS-CoV-2 based on difference in SRR (SRR of mRNA-1273.617.2 against the variant B.1.617.2 at Day 29 – SRR of mRNA-1273 against ancestral SARS-CoV-2 at Day 57) with a non-inferiority margin of 10%.

The same analysis methods described in Section 8.5.3.2.1 will be used for the primary immunogenicity objective for Part C.

The primary immunogenicity objective (against the variant) is considered met if non-inferiority is demonstrated based on both GMR and SRR difference, specifically:

- The lower bound of the 95% CI of the GMT ratio between mRNA-1273.617.2 against the variant (B.1.617.2) at Day 29 as compared to 100 µg mRNA-1273 against ancestral SARS-CoV-2 at Day 57 is > 0.667 based on the non-inferiority margin of 1.5, and
- The lower bound of the 95% CI of the SRR difference (mRNA-1273.617.2 against the variant [B.1.617.2] at Day 29 – 100 µg mRNA-1273 against ancestral SARS-CoV-2 at Day 57 is > -10%)

###### 8.5.3.2.5. Analysis for the Primary Immunogenicity Objective for Part D

Each dose level of mRNA-1273.213 (50 and 100 µg) will be assessed with respect to mRNA-1273 primary series historical control separately. For the primary objective on immune response, there are 4 null hypotheses to be tested for each arm.

A. $H_0^1$: 50 µg or 100 µg mRNA-1273.213, as a single booster dose, against the variant B.1.617.2 is inferior to the primary series of (100 µg) mRNA-1273 against ancestral SARS-CoV-2 based on the GMT ratio of mRNA-1273.617.2 against the variant B.1.617.2 at Day 29 compared to mRNA-1273 against ancestral SARS-CoV-2 at Day 57 with a non-inferiority margin of 1.5

B. $H_0^2$: 50 µg or 100 µg mRNA-1273.213, as a single booster dose, against the variant B.1.617.2 is inferior to the primary series of (100 µg) mRNA-1273 against ancestral SARS-CoV-2 based on the difference in SRR (SRR of mRNA-1273.617.2 against the variant B.1.617.2 at Day 29 – SRR of mRNA-1273 against ancestral SARS-CoV-2 at Day 57) with a non-inferiority margin of 10%

C. $H_0^3$: 50 µg or 100 µg mRNA-1273.213, as a single booster dose, against the variant B.1.351 is inferior to the primary series of (100 µg) mRNA-1273 against ancestral SARS-CoV-2 based on the GMT ratio of mRNA-1273.213 against the variant B.1.351 at Day 29 compared to mRNA-1273 against ancestral SARS-CoV-2 at Day 57 with a non-inferiority margin of 1.5

D. $H_0^4$: 50 µg or 100 µg mRNA-1273.213, as a single booster dose, against the variant B.1.351 is inferior to the primary series of (100 µg) mRNA-1273 against ancestral SARS-CoV-2 based on the difference in SRR (SRR of mRNA-1273.213 against the variant B.1.351 at Day 29 – SRR of mRNA-1273 against ancestral SARS-CoV-2 at Day 57) with a non-inferiority margin of 10%

The same analysis methods described in Section 8.5.3.2.1 will be used for the primary immunogenicity objective for Part D.

The primary immunogenicity objective (against the variant) is considered met if non-inferiority is demonstrated based on both GMR and SRR difference, specifically:

- The lower bound of the 95% CI of the GMT ratio between mRNA-1273.213 against the variant (B.1.617.2) at Day 29 as compared to 100 µg mRNA-1273 against ancestral SARS-CoV-2 at Day 57 is > 0.667 based on the non-inferiority margin of 1.5, and
- The lower bound of the 95% CI of the SRR difference (mRNA-1273.213 against the variant [B.1.617.2] at Day 29 – 100 µg mRNA-1273 against ancestral SARS-CoV-2 at Day 57 is > -10%), and
- The lower bound of the 95% CI of the GMT ratio between mRNA-1273.213 against the variant (B.1.351) at Day 29 as compared to 100 µg mRNA-1273 against ancestral SARS-CoV-2 at Day 57 is > 0.667 based on the non-inferiority margin of 1.5, and
- The lower bound of the 95% CI of the SRR difference (mRNA-1273.213 against the variant [B.1.351] at Day 29 – 100 µg mRNA-1273 against ancestral SARS-CoV-2 at Day 57 is > -10%).

###### 8.5.3.2.6. Analysis for the Primary Immunogenicity Objective for Part F

Cohort 1:

50 µg mRNA-1273.529 booster dose (first booster dose) will be assessed with respect to mRNA-1273 booster dose (first booster dose) using an external comparator (details regarding the external historical control will be included in the SAP). For the primary immunogenicity objective, there are 3 hypotheses to be tested.

A. $H_1^1$: 50 µg mRNA-1273.529, as a single booster dose, against the variant B.1.1.529 is non-inferior to the booster dose of (50 µg) mRNA-1273 against B.1.1.529 at Day 29 compared to mRNA-1273 against the variant B.1.1.529 at Day 29 with a non-inferiority margin of 1.5.

B. $H_1^2$: 50 µg mRNA-1273.529, as a single booster dose, against the variant B.1.1.529 is non-inferior to the booster dose of (50 µg) mRNA-1273 against B.1.1.529 based on the difference in SRR at Day 29 with a non-inferiority margin of 10%.

C. $H_1^3$: 50 µg mRNA-1273.529, as a single booster dose, against the variant B.1.1.529 is superior to the booster dose of (50 µg) mRNA-1273 against B.1.1.529 based on the GMT ratio of mRNA-1273.529 against the variant B.1.1.529 at Day 29 compared to mRNA-1273 against the variant B.1.1.529 at Day 29.

An ANCOVA model will be performed to assess the difference in immune response between mRNA-1273.529 and mRNA-1273 (using external comparator) as the first booster dose. For immune response against the B.1.1.529 strain, in the ANCOVA model, antibody titers at Day 29 postbooster against the B.1.1.529 strain will be a dependent variable, and a group variable (mRNA-1273.529 and mRNA-1273) will be the fixed effect, adjusting for age groups (< 65, ≥ 65 years) and prebooster antibody titer level, if applicable.

The GMT will be estimated by the GLSM from the model and its corresponding 95% will be provided for each group. The GMR (ratio of GMTs) for mRNA-1273.529 with respect to mRNA-1273 will be estimated by the ratio of GLSM from the model and the corresponding 95% CIs will be provided. The 95% CI for GMR will be used to assess the between group difference in immune response against the B.1.1.529 strain for mRNA-1273.529 at Day 29 compared to the mRNA-1273.

The number and percentage (rate) of participants achieving seroresponse at Day 29 will be summarized with 95% CI calculated using the Clopper-Pearson method for each group. The difference of SRR between mRNA-1273.529 and mRNA-1273 will be calculated with 95% CI based on Miettinen-Nurminen method. The non-inferiority in SRR of mRNA-1273.529 compared to mRNA-1273 will be considered demonstrated if the lower bound of the 95% of the SRR difference is > -10% based on the non-inferiority margin of 10%.

The primary immunogenicity objective (against the B.1.1.529) is considered met if non-inferiority is demonstrated based on GMR and SRR difference, specifically:

- The lower bound of the 95% CI of the GMT ratio between mRNA1273.529 against the variant (B.1.1.529) at Day 29 as compared to 50 µg mRNA1273 against B.1.1.529 at Day 29 is > 0.667 based on the non-inferiority margin of 1.5
- The lower bound of the 95% CI of the SRR difference (50 µg mRNA-1273.529 against the variant B.1.1.529 at Day 29–- 50 µg mRNA-1273 against B.1.1.529 at Day 29) is >-10%
- If non-inferiority is demonstrated (based on GMT ratio and SRR difference), the lower bound of 95% CI of the GMT ratio will be compared to 1, if it's greater than 1, then superiority is demonstrated.

Cohort 2

A. $H_1^1$: 50 µg mRNA-1273.529, as a second booster dose, against the variant B.1.1.529 is non-inferior to the second booster dose of (50 µg) mRNA-1273 against B.1.1.529 based on the GMT ratio of mRNA-1273.529 against the variant B.1.1.529 at Day 29 compared to mRNA-1273 against the variant B.1.1.529 at Day 29 with a non-inferiority margin of 1.5.

B. $H_1^2$: 50 µg mRNA-1273.529, as a second booster dose, against the variant B.1.1.529 is non-inferior to the second booster dose of (50 µg) mRNA-1273 against B.1.1.529 based on the difference in SRR at Day 29 with a non-inferiority margin of 10%.

C. $H_1^3$: 50 µg mRNA-1273.529, as a second booster dose, against the variant B.1.1.529 is superior to the second booster dose of (50 µg) mRNA-1273 against B.1.1.529 based on the GMT ratio of mRNA-1273.529 against the variant B.1.1.529 at Day 29 compared to mRNA-1273 against the variant B.1.1.529 at Day 29.

The primary immunogenicity objective (against the B.1.1.529) is considered met if non-inferiority is demonstrated based on GMR and SRR difference, specifically:

- The lower bound of the 95% CI of the GMT ratio between mRNA1273.529 against the variant (B.1.1.529) at Day 29 as compared to 50 µg mRNA1273 against B.1.1.529 at Day 29 is > 0.667 based on the non-inferiority margin of 1.5
- The lower bound of the 95% CI of the SRR difference (50 µg mRNA-1273.529 against the variant B.1.1.529 at Day 29–- 50 µg mRNA-1273 against B.1.1.529 at Day 29) is >-10%
- If non-inferiority is demonstrated (based on GMT ratio and SRR difference), the lower bound of 95% CI of GMT ratio will be compared to 1, if it's greater than 1, then superiority is demonstrated.

###### 8.5.3.2.7. Analysis for the Primary Immunogenicity Objective for Part G

50 µg mRNA-1273.214 as the second booster dose will be compared to 50 µg mRNA-1273 as the second booster dose (active control arm in Part F, Cohort 2)

For the primary immunogenicity objective, there are 8 hypotheses (4 hypotheses at Day 29 and 4 hypotheses at Day 91). Figure 1 depicts the hypotheses testing strategy.

A. $H_1^1$: 50 µg mRNA-1273.214, as a second booster dose, against the variant B.1.1.529 is non-inferior to the second booster dose of (50 µg) mRNA-1273 against B.1.1.529 based on the GMT ratio of mRNA-1273.529 against the variant B.1.1.529 at Day 29 compared to mRNA-1273 against the variant B.1.1.529 at Day 29 with a non-inferiority margin of 1.5.

B. H12: 50 µg mRNA-1273.214, as a second booster dose, against the variant B.1.1.529 is non-inferior to the second booster dose of (50 µg) mRNA-1273 against B.1.1.529 based on the difference in SRR at Day 29 with a non-inferiority margin of 10%.

C. $H_1^3$: 50 µg mRNA-1273.214, as a second booster dose, against the ancestral SARS-CoV-2 is non-inferior to the second booster dose of (50 µg) mRNA-1273 against ancestral SARS-CoV-2 based on the GMT ratio of mRNA-1273.214 against ancestral SARS-CoV-2 at Day 29 compared to mRNA-1273 against ancestral SARS-CoV-2 at Day 29 with a non-inferiority margin of 1.5.

D. $H_1^4$: 50 µg mRNA-1273.214, as a second booster dose, against the variant B.1.1.529 is superior to the second booster dose of (50 µg) mRNA-1273 against B.1.1.529 based on the GMT ratio of mRNA-1273.214 against the variant B.1.1.529 at Day 29 compared to mRNA-1273 against the variant B.1.1.529 at Day 29.

E. $H_1^5$: 50 µg mRNA-1273.214, as a second booster dose, against the variant B.1.1.529 is non-inferior to the second booster dose of (50 µg) mRNA-1273 against B.1.1.529 based on the GMT ratio of mRNA-1273.214 against the variant B.1.1.529 at Day 91 compared to mRNA-1273 against the variant B.1.1.529 at Day 91 with a non-inferiority margin of 1.5.

F. $H_1^6$: 50 µg mRNA-1273.214, as a second booster dose, against the variant B.1.1.529 is non-inferior to the second booster dose of (50 µg) mRNA-1273 against B.1.1.529 based on the difference in SRR at Day 91 with a non-inferiority margin of 10%.

G. $H_1^7$: 50 µg mRNA-1273.214, as a second booster dose, against ancestral SARS-CoV-2 is non-inferior to the second booster dose of (50 µg) mRNA-1273 against ancestral SARS-CoV-2 based on the GMT ratio of mRNA-1273.214 against ancestral SARS-CoV-2 at Day 91 compared to mRNA-1273 against ancestral SARS-CoV-2 at Day 91 with a non-inferiority margin of 1.5.

H. $H_1^8$: 50 µg mRNA-1273.214, as a second booster dose, against the variant B.1.1.529 is superior to the second booster dose of (50 µg) mRNA-1273 against B.1.1.529 based on the GMT ratio of mRNA-1273.214 against the variant B.1.1.529 at Day 91 compared to mRNA-1273 against the variant B.1.1.529 at Day 91.

**Figure 1: Statistical Hypotheses Testing Strategy for Part G**

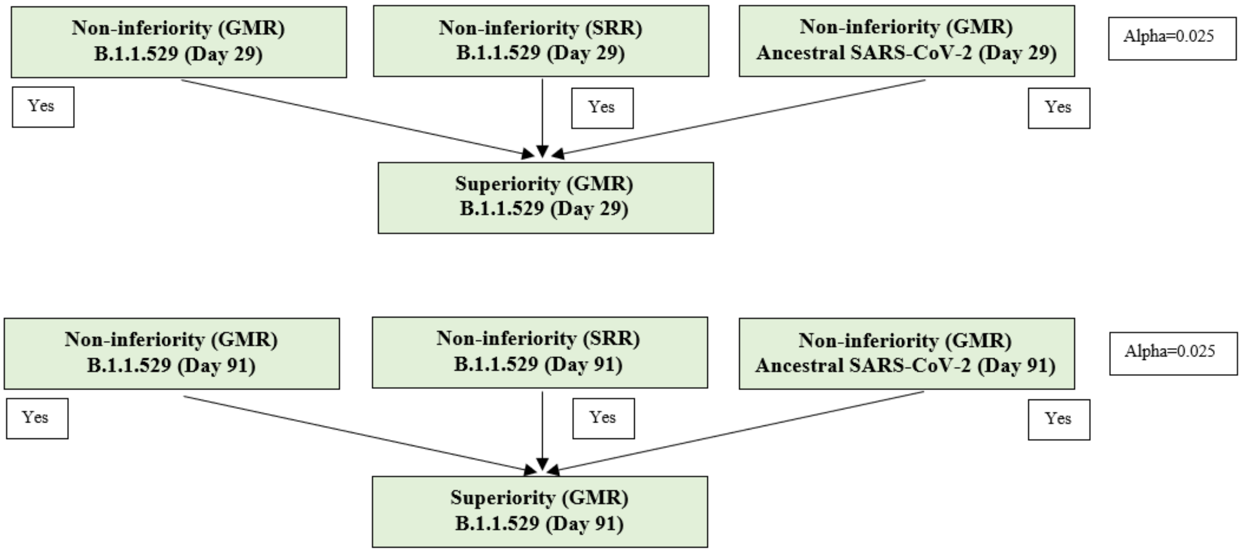

For the primary immunogenicity objective, an alpha of 0.05 (two-sided) will be allocated to the 2 time points. Day 29 and Day 91 will each have an alpha of 0.025 (two-sided) for hypotheses testing.

The analyses method described in Part F Cohort 1 will be used for Part G.

The primary immunogenicity objective is considered met if non-inferiority against B.1.1.529 based on GMR, SRR difference, and non-inferiority against ancestral SARS-CoV-2 based on GMR are demonstrated at Day 29 or Day 91.

Day 29: alpha = 0.025 (2-sided)

- The lower bound of the 97.5% CI of the GMT ratio between mRNA-1273.214 against the variant (B.1.1.529) at Day 29 as compared to 50 µg mRNA-1273 against B.1.1.529 at Day 29 is > 0.667 based on the non-inferiority margin of 1.5
- The lower bound of the 97.5% CI of the SRR difference (50 µg mRNA-1273.214 against the variant B.1.1.529 at Day 29–- 50 µg mRNA-1273 against B.1.1.529 at Day 29) is >-10%
- The lower bound of the 97.5% CI of the GMT ratio between mRNA-1273.214 against ancestral SARS-CoV-2 at Day 29 as compared to 50 µg mRNA-1273 against ancestral SARS-CoV-2 at Day 29 is > 0.667 based on the non-inferiority margin of 1.5
- If non-inferiority is demonstrated for both B.1.1.529 (based on GMR and SRR) and ancestral SARS-CoV-2 (based on GMR), the lower bound of 97.5% CI of GMR will be compared to 1, if it's greater than 1, then superiority against B.1.1.529 is demonstrated

Day 91: alpha=0.025 (2-sided)

Hypotheses testing at Day 91 will be performed in the same manner, first test 2 non-inferiority hypotheses (2 against the B.1.1.529 strain and one against ancestral SARS-CoV-2) at alpha of 0.025 level (two-sided). Once non-inferiority is demonstrated for both B.1.1529 and ancestral SARS-CoV-2, then superiority testing against the B.1.1.529 at alpha of 0.025 level (two-sided) will be performed.

For the key secondary objective, there are 2 hypotheses to be tested (Day 29 and Day 91 each with alpha level of 0.025, 2-sided):

I. $H_1^9$: 50 µg mRNA-1273.214, as a second booster dose, against ancestral SARS-CoV-2 is non-inferior to the booster dose of (50 µg) mRNA-1273 against ancestral SARS-CoV-2 based on the difference in SRR at Day 29 with a non-inferiority margin of 10%.

J. $H_1^{10}$: 50 µg mRNA-1273.214, as a second booster dose, against ancestral SARS-CoV-2 is non-inferior to the booster dose of (50 µg) mRNA-1273 against ancestral SARS-CoV-2 based on the difference in SRR at Day 91 with a non-inferiority margin of 10%.

If the lower bound of the 97.5% CI of the SRR difference (50 µg mRNA-1273.214 against ancestral SARS-CoV-2–- 50 µg mRNA-1273 against ancestral SARS-CoV-2) is >-10% at Day 29 or Day 91, then the key secondary objective will be considered met.

In the event that an early assessment of the 1273.214 data is needed due to public health concerns, a two-staged approach will be used. Specifically, a subset of participants' (ie, 50 first enrolled participants) serum samples will first be tested against ancestral SARS-CoV-2 and various VOCs. For the Day 29 and Day 91 immunogenicity analyses, all participants' immune data will be used in the formal analysis to evaluate the primary immunogenicity objective.

###### 8.5.3.2.8. Analysis for the Primary Immunogenicity Objective for Part H

The primary immunogenicity objective will be assessed in the Per-Protocol Set for Immunogenicity, SARS-CoV-2 negative, descriptive summaries of antibody GMTs, GMFR, and SRR will be provided (refer to Section 8.5.3.4 for analysis methods).

For the primary immunogenicity objective, there are 5 hypotheses to be tested as shown in Figure 2:

A. $H_1^1$: 50 µg mRNA-1273.222, as a second booster dose, against Omicron BA.4/5 is non-inferior to the second booster dose of (50 µg) mRNA-1273 against Omicron BA.4/5 based on the GMT ratio of mRNA-1273.222 against Omicron BA.4/5 at Day 29 compared to mRNA-1273 against Omicron BA.4/5 at Day 29 with a non-inferiority margin of 1.5.

B. $H_1^2$: 50 µg mRNA-1273.222, as a second booster dose, against Omicron BA.4/5 is non-inferior to the second booster dose of (50 µg) mRNA-1273 against Omicron BA.4/5 based on the difference in SRR at Day 29 with a non-inferiority margin of 5%.

C. $H_1^3$: 50 µg mRNA-1273.222, as a second booster dose, against ancestral SARS-CoV-2 D614G is non-inferior to the second booster dose of (50 µg) mRNA-1273 against ancestral SARS-CoV-2 D614G based on the GMT ratio of mRNA-1273.222 against ancestral SARS-CoV-2 D614G at Day 29 compared to mRNA-1273 against ancestral SARS-CoV-2 D614G at Day 29 with a non-inferiority margin of 1.5.

D. $H_1^4$: 50 µg mRNA-1273.222, as a second booster dose, against ancestral SARS-CoV-2 D614G is non-inferior to the second booster dose of (50 µg) mRNA-1273 against ancestral SARS-CoV-2 D614G based on the difference in SRR at Day 29 with a non-inferiority margin of 10%.

E. $H_1^5$: 50 µg mRNA-1273.222, as a second booster dose, against Omicron BA.4/5 is superior to the second booster dose of (50 µg) mRNA-1273 against Omicron BA.4/5 based on the GMT ratio of mRNA-1273.222 against Omicron BA.4/5 at Day 29 compared to mRNA-1273 against Omicron BA.4/5 at Day 29.

**Figure 2: Statistical Hypotheses Testing for Part H**

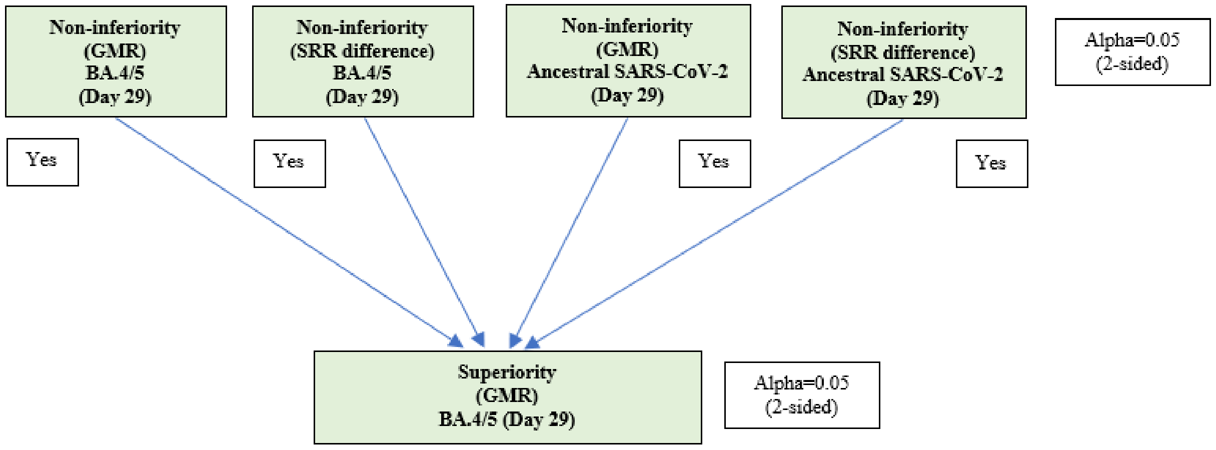

The analyses methods described in Part F Cohort 1 will be used for Part H.

The primary immunogenicity objective is considered met if non-inferiority against BA.4/5 and ancestral SARS-CoV-2 D614G based on GMR, SRR difference at Day 29 are demonstrated.

###### 8.5.3.2.1. Analysis for the Primary Immunogenicity Objective for Part J

No formal hypothesis testing will be performed for Part J and all safety and immunogenicity analyses will be descriptive.

The primary immunogenicity objective will be assessed using the Per-Protocol Set for Immunogenicity. Comparisons on immunogenicity response between the 2 arms may be performed.

An ANCOVA model will be used to compare antibody response between treatment arms. Day 15 and Day 29 antibody titers (against SARS-CoV-2 variants) will be included in the model as a dependent variable and treatment group variable (mRNA-1273.815, mRNA-1273.231) will be included as fixed effect. The model may adjust for other covariates as appropriate. Geometric mean ratio and the corresponding 95% CI will be estimated from the model. Seroresponse rate difference at Day 15 and Day 29 will be compared and 95% CI will be provided using Miettinen-Nurminen method.

##### 8.5.3.3. Analysis for Secondary Immunogenicity Objective

###### 8.5.3.3.1. Parts A.1, B, C, and D

The same analysis methods Section 8.5.3.2.1 will be used to evaluate each booster arm against variant as compared with 100 µg mRNA-1273 primary series against ancestral SARS-CoV-2, specifically:

- To compare immune response to circulating VOCs after a single booster dose of 50 µg or 100 µg mRNA-1273.211 (Part A.1) with 100 µg mRNA-1273 primary series against ancestral SARS-CoV-2
- To compare immune response to circulating VOCs after a single booster dose of 100 µg mRNA-1273 (Part B) with 100 µg mRNA-1273 primary series against ancestral SARS-CoV-2
- To compare immune response to circulating VOCs after a single booster dose of 50 µg or 100 µg mRNA-1273.617.2 (Part C) with 100 µg mRNA-1273 primary series against ancestral SARS-CoV-2
- To compare immune response to circulating VOCs after a single booster dose of 50 µg or 100 µg mRNA-1273.213 (Part D) with 100 µg mRNA-1273 primary series against ancestral SARS-CoV-2

##### 8.5.3.4. Other Analysis of Immunogenicity

SARS-CoV-2-specific bAb and nAb are assessed at multiple timepoints in each part of this study.

For each of the antibodies of interest, eg, levels of SARS-CoV-2–specific bAb and SARS-CoV-2-specific nAb, the GMT or level with corresponding 95% CI at each time point, and GMFR of post-baseline/baseline titers or levels with corresponding 95% CI at each post-baseline time point will be provided for each arm. The 95% CIs will be calculated based on the t-distribution of the log-transformed values then back-transformed to the original scale for presentation. The following descriptive statistics will also be provided at each time point: number of participants (n), median, minimum, and maximum.

The MMRM will be used to analyze all postbooster measures for between booster comparison when applicable, the model will include treatment group, analysis visit, treatment by visit interaction, and adjusting for age groups and prebooster titer levels. An unstructured covariance structure will be used to model the within-participant errors. The GMT will be estimated from the model and its corresponding 95% CI will be provided for each group at each postboost timepoint. The GMR (ratio of GMTs) will be estimated from the model and the corresponding 95% CI will be provided at each postboost timepoint.

The SRR of each arm against ancestral SARS-CoV-2 and variants, defined as the percentage of participants achieving seroresponse against ancestral SARS-CoV-2 and variants respectively, will be provided for each arm with the 95% CI calculated using the Clopper-Pearson method.

The primary definition of seroresponse is defined as ≥ 4 × LLOQ for those with pre-dose 1 of primary series baseline < LLOQ; ≥ 4-foldrise for those with pre-dose 1 of primary series baseline ≥ LLOQ. The secondary definition of seroresponse is defined as ≥ 4 × LLOQ for those with prebooster baseline < LLOQ; ≥ 4-foldrise for those with prebooster ≥ LLOQ. SRR will be summarized using both definitions for all the study parts.

##### 8.5.4. Safety Analyses

All safety analyses are descriptive in nature and will be based on the Safety Set, except summaries of solicited ARs, which will be based on the Solicited Safety Set. All safety analyses will be provided by study arm.

Safety and reactogenicity will be assessed by clinical review of all relevant parameters including solicited ARs (local and systemic ARs), unsolicited AEs, treatment-related AEs, severe AEs, SAEs, MAAEs, AESI, AEs leading to withdrawal, vital sign measurements, and physical examination findings.

The number and percentage of participants with any solicited local AR, with any solicited systemic AR, with any solicited AR during the 7-day follow-up period after each vaccination, and with Grade 3 or higher solicited AR will be provided. A 2-sided 95% exact CI using the Clopper-Pearson method will be provided for the percentage of participants with any solicited AR.

The number and percentage of participants with unsolicited AEs, treatment-related AEs, severe AEs, SAEs, MAAEs, AESI, and AEs leading to withdrawal will be summarized. Unsolicited AEs will be presented by MedDRA system organ class and preferred term. Unsolicited AEs will be coded according to the MedDRA Dictionary for Adverse Reaction Terminology.

The number of events of solicited ARs, unsolicited AEs/SAEs, MAAEs, AEs leading to withdrawal, and AESI will be reported in summary tables accordingly. Pregnancy outcomes will also be summarized.

Table 16 summarizes the analysis strategy for safety parameters. For all other safety parameters, descriptive summary statistics will be provided. Further details will be described in the SAP.

**Table 16: Analysis Strategy for Safety Parameters**

| Safety Endpoint | Number and Percentage of Participants, Number of Events | 95% CI for Each Study Arm |
|---|---|---|
| Any Solicited AR (overall and by local, systemic) | X | X |
| Any Unsolicited AE | X | — |
| Any SAE | X | — |
| Any Unsolicited MAAE | X | — |
| Any Unsolicited AESI | X | — |
| Any Unsolicited Treatment-Related AE | X | — |
| Any Treatment-Related SAE | X | — |
| Any Unsolicited AE Leading to Withdrawal from Study Participation | X | — |
| Any Severe Unsolicited AE | X | — |
| Any Treatment-Related Severe Unsolicited AE | X | — |

Abbreviations: AE = adverse event; AESI = adverse event of special interest; AR = adverse reaction; CI = confidence interval; MAAE = medically attended adverse event; SAE = serious adverse event.
Notes: 95% CI using the Clopper-Pearson method, X = results will be provided.

##### 8.5.5. Exploratory Analyses

Exploratory analyses on immune response to selected virus strains may be performed to compare booster regimens including:

- Compare immune response to ancestral SARS-CoV-2 and variants after a single booster dose of 50 µg mRNA-1273.211 (Part A.1) vs. a single booster dose of 100 µg mRNA-1273.211 (Part A.1)
- Compare immune response to ancestral SARS-CoV-2 and variants after a single booster dose of 50 µg mRNA-1273.211 (Part A.1) vs. a single booster dose of 100 µg mRNA-1273 (Part B)
- Compare immune response to ancestral SARS-CoV-2 and variants after a single booster dose of 100 µg mRNA-1273.211 (Part A.1) vs. a single booster dose of 100 µg mRNA-1273 (Part B)
- Compare immune response to ancestral SARS-CoV-2 and variants after a single booster dose of 50 µg or 100 µg mRNA-1273.617.2 (Part C) vs. a single booster dose of 100 µg mRNA-1273 (Part B)
- Compare immune response to ancestral SARS-CoV-2 and variants after a single booster dose of 50 µg mRNA-1273.617.2 (Part C) vs. a single booster dose of 100 µg mRNA-1273.617.2 (Part C)
- Compare immune response to ancestral SARS-CoV-2 and variants after a single booster dose of 50 µg or 100 µg mRNA-1273.211 (Part A.1) vs. a single booster dose of 50 µg or 100 µg mRNA-1273.617.2 (Part C)

- Compare immune response to ancestral SARS-CoV-2 and variants after a single booster dose of 50 µg mRNA-1273.213 (Part D) vs. a single booster dose of 100 µg mRNA-1273.213 (Part D)
- To compare immune response to ancestral SARS-CoV-2 and variants after a single booster dose of 50 µg or 100 µg mRNA-1273.213 (Part D) vs. a single booster dose of 50 µg or 100 µg mRNA-1273.211 (Part A.1)
- To compare immune response to ancestral SARS-CoV-2 and variants after a single booster dose of 50 µg or 100 µg mRNA-1273.213 (Part D) vs. a single booster dose of 100 µg mRNA-1273 (Part B)
- To compare immune response to ancestral SARS-CoV-2 and variants after a single booster dose of 50 µg or 100 µg mRNA-1273.213 (Part D) vs. a single booster dose of 50 µg or 100 µg mRNA-1273.617.2 (Part C)

Other exploratory analyses to compare immune response of each booster against variants compared to the priming series of mRNA-1273 against the variant may be performed.

Exploratory analyses not addressed in Section 8.5.3 will be described in the SAP before database lock.

##### 8.5.6. Subgroup Analyses

Immunogenicity will be assessed in the following subgroups; however, subgroup analyses may not be performed for Part J due to small sample size:

- Age (18 to < 65, and ≥ 65 years)
- Sex (female, male)
- Baseline/prebooster SARS-CoV-2 status (negative, positive) if there is enough number of prebooster positives
- Race and ethnicity group (non-Hispanic White, communities of color)

Safety may be assessed for the same subgroups.

#### 8.6. Planned Analyses

##### 8.6.1. Interim Analysis

The interim analysis will be conducted based on safety and immunogenicity data collected through Day 15 or Day 29. The interim analysis may be conducted either after all participants in Parts A.1, A.2, B, C, D, F, G, H, or J have completed their Day 15 or Day 29 visit assessments and/or subsequent timepoint visits (eg, Day 91 for Parts F, G, H, and J) or combined after the last participant of each study part (Parts A.1, A.2, B, C, D, F, G, H, or J) dose arm, or pre-specified subset of the dose arm has completed their Day 15 or Day 29 visit assessments.

##### 8.6.2. Final Analyses

The final analysis of all endpoints will be performed after all participants have completed all planned study procedures. Results of this analysis will be presented in a final CSR, including individual listings. The final CSR will include full analyses of all safety and immunogenicity through Day 366 (Month 12) for Parts A.1, B, C, D, F, and G and through D181 (Month 6) for Parts A.2, H, and J.

### 10. SUPPORTING DOCUMENTATION AND OPERATIONAL CONSIDERATIONS

#### 10.1. APPENDIX 1: Schedule of Events

##### Table 17: Schedule of Events for Parts A.1, B, C, D, F, and G

| **Visit Number** | Screening[1] | V1 | | V2 | V3 | V3a | | | V4 | | | V5 | UNS |
|---|---|---|---|---|---|---|---|---|---|---|---|---|---|
| **Type of Visit** | C | C | SC | C | C | C | SFU (eDiary) | SFU (SC) | C | SFU (eDiary) | SFU (SC) | C | C |
| **Month Timepoint** | M0 | M0 | | | M1 | M3 | | | M6 | | | M12 | -- |
| **Study Visit Day** | D0[1] | D1[1] | D8 | D15 | D29 | D91 (Parts F and G only) | Every 2 weeks D36–D162[2] | Every 2 weeks D43-D169 | D181 | Every 2 weeks D202–D342[2] | Every 2 weeks D209-D349 | D366 | |
| **Window Allowance (Days)** | -7 | 0 | +3 | ±3 | -7 | ±7 | ±2 | ±3 | ±14 | ±2 | ±3 | ±14 | -- |
| **Days Since Booster Injection** | | 0 | 7 | 14 | 28 | 90 | -- | | 180 | -- | | 365 | -- |
| Informed consent form | X | | | | | | | | | | | | |
| Study injection (including 30-minute post-dosing observation period) | | X | | | | | | | | | | | |
| Confirm participant meets inclusion and exclusion criteria | X | | | | | | | | | | | | |
| Physical examination including vital signs[3] | X | X | | | X | X | | | X | | | X | X |
| Pregnancy testing | | X | | | | | | | | | | | |
| Blood for SARS-CoV-2 serology (antinucleocapsid antibody) | | X | | | X | X | | | X | | | X | X |
| Blood for immune response to vaccination (binding and neutralizing antibody)[4] | | X | | X | X | X | | | X | | | X | X |

| **Visit Number** | Screening[1] | V1 | | V2 | V3 | V3a | | | V4 | | | V5 | UNS |
|---|---|---|---|---|---|---|---|---|---|---|---|---|---|
| **Type of Visit** | C | C | SC | C | C | C | SFU (eDiary) | SFU (SC) | C | SFU (eDiary) | SFU (SC) | C | C |
| **Month Timepoint** | M0 | M0 | | | M1 | M3 | | | M6 | | | M12 | -- |
| **Study Visit Day** | D0[1] | D1[1] | D8 | D15 | D29 | D91 (Parts F and G only) | Every 2 weeks D36–D162[2] | Every 2 weeks D43-D169 | D181 | Every 2 weeks D202–D342[2] | Every 2 weeks D209-D349 | D366 | |
| **Window Allowance (Days)** | -7 | 0 | +3 | ±3 | -7 | ±7 | ±2 | ±3 | ±14 | ±2 | ±3 | ±14 | -- |
| **Days Since Booster Injection** | | 0 | 7 | 14 | 28 | 90 | -- | | 180 | -- | | 365 | -- |
| Nasopharyngeal swab sample for SARS-CoV-2[4] | | X | | | X | X | | | X | | | X | X[5] |
| eDiary activation for recording solicited local and systemic reactogenicity adverse reactions (7 days) | | X | | | | | | | | | | | |
| Review of eDiary | | | X | | | | | | | | | | |
| Follow-up safety calls[6] | | | X | | | | | X | | | X | | |
| Recording of unsolicited AEs | | X | X | X | X | | | | | | | | |
| Recording of MAAEs and concomitant medications relevant to or for the treatment of the MAAE[7] | | X | X | X | X | X | X | X | X | X | X | X | |
| Recording of SAEs and concomitant medications relevant to or for the treatment of the SAE[7] | | X | X | X | X | X | X | X | X | X | X | X | |
| Recording of AESI | | X | X | X | X | X | X | X | X | X | X | X | |

| **Visit Number** | Screening[1] | V1 | | V2 | V3 | V3a | | | V4 | | | V5 | UNS |
|---|---|---|---|---|---|---|---|---|---|---|---|---|---|
| **Type of Visit** | C | C | SC | C | C | C | SFU (eDiary) | SFU (SC) | C | SFU (eDiary) | SFU (SC) | C | C |
| **Month Timepoint** | M0 | M0 | | | M1 | M3 | | | M6 | | | M12 | -- |
| **Study Visit Day** | D0[1] | D1[1] | D8 | D15 | D29 | D91 (Parts F and G only) | Every 2 weeks D36–D162[2] | Every 2 weeks D43-D169 | D181 | Every 2 weeks D202–D342[2] | Every 2 weeks D209-D349 | D366 | |
| **Window Allowance (Days)** | -7 | 0 | +3 | ±3 | -7 | ±7 | ±2 | ±3 | ±14 | ±2 | ±3 | ±14 | -- |
| **Days Since Booster Injection** | | 0 | 7 | 14 | 28 | 90 | -- | | 180 | -- | | 365 | -- |
| Recording of concomitant medications and non-study vaccinations[8] | | X | X | X | X | | | | | | | X | |
| Study completion | | | | | | | | | | | | X | |
| PBMCs<br>May be collected for a subset of participants at selected sites. | X | | | X | X | X | | | | | | | |

Abbreviations: AE = adverse event; AESI = adverse event of special interest; C = clinic visit; COVID-19 = coronavirus disease 2019; D = day; eDiary = electronic diary; M = month; MAAE = medically attended adverse event; NP = nasopharyngeal; RT-PCR = reverse transcriptase polymerase chain reaction; PBMC = peripheral blood mononuclear cells; SAE = serious adverse event; SARS-CoV-2 = severe acute respiratory syndrome coronavirus 2; SC = safety telephone call; SFU = safety follow-up; UNS = unscheduled visit; V = visit.

Note: In accordance with “FDA Guidance on Conduct of Clinical Trials of Medical Products during COVID-19 Public Health Emergency” (DHHS 2020), investigators may convert study site visits to telemedicine visits with the approval of the Sponsor.

1. The Screening Visit and Day 1 (vaccination) visit can be combined and occur on the same day.
2. Safety follow-up via eDiary questionnaire will be performed every 2 weeks from at Day 36 to Day 162, and from Day 202 to Day 342. These study days are relative to Day 1 vaccination. Adverse reactions recorded in the eDiary beyond Day 7 should be reviewed by the study site staff either during the next scheduled telephone call or at the next study site visit.
3. Physical examination: A full physical examination, including height and weight, will be performed on Day 1. Symptom-directed physical examinations may be performed at other time points at the discretion of the investigator. Any clinically significant finding identified during a study visit should be reported as an MAAE. Vital signs are to be collected pre- and post-dosing on the day of injection (Day 1) only. When applicable, vital sign measurements should be performed before blood collection. For participants who are febrile (body temperature ≥ 38.0°C [100.4°F]) before injection on Day 1, the visit must be

rescheduled within the relevant window period to receive the injection. Afebrile participants with minor illnesses can be administered investigational product at the discretion of the investigator.

4. The NP swab sample, collected prior to vaccination on Day 1, will be used to ascertain the presence of SARS-CoV-2 via RT-PCR. The NP swab sample will also be collected within 24 hours if participant experience signs and symptoms of SARS-CoV-2 infection. It is important to note that some of the symptoms of COVID-19 overlap with solicited local and systemic reactogenicity ARs, that are expected after vaccination with mRNA-1273.211 (eg, myalgia, headache, fever, and chills). During the first 7 days after vaccination, when these solicited ARs are common, investigators should use their clinical judgment to decide if an NP swab should be collected. The collection of an NP swab prior to the Day 1 vaccination can help ensure that cases of COVID-19 are not overlooked. Any study participant reporting respiratory symptoms during the 7-day period after vaccination should be evaluated for COVID-19.
5. Nasopharyngeal swabs will be collected during the unscheduled/illness visit for testing for the presence of SARS-CoV-2 via RT-PCR and for assessing non-SARS-CoV-2 causes of upper and lower respiratory tract infection.
6. Trained site personnel will call all participants to collect information relating to any AEs, MAAEs, SAEs, AEs leading to withdrawal, information on concomitant medications associated with those events, AESI, and any non-study vaccinations. In addition, study personnel will collect information on known participant exposure to someone with known COVID-19 or SARS-CoV-2 infection and on participant experience of COVID-19 symptoms. Sites will collect this information for eDiary days only if eDiary responses indicate the need for follow-up via telephone.
7. All concomitant medications relevant to or for the treatment of an SAE or MAAE will be recorded from Day 1 through the end of study visit (Day 366).
8. All concomitant medications and non-study vaccinations will be recorded through 28 days following injection.

**Table 18: Schedule of Events for Part A.2**

| **Visit Number** | A.2 Screening[1] | V6 | | V7 | V8 | V9 | | | V10 | A.2 UNS |
|---|---|---|---|---|---|---|---|---|---|---|
| **Type of Visit** | C | C | SC | C | C | C | SFU (eDiary) | SFU (SC) | C | C |
| **Month Timepoint** | M0 | M0 | | | M1 | M3 | | | M6 | -- |
| **Study Visit Day** | D0[1] | D1[1,9] | D8 | D15 | D29 | D91 | Every 2 weeks D36–D162[2] | Every 2 weeks D43-D169 | D181 | |
| **Window Allowance (Days)** | -7 | 0 | +3 | ±3 | -7 | ±7 | ±2 | ±3 | ±14 | -- |
| **Days Since Booster Injection** | | 0 | 7 | 14 | 28 | 90 | -- | | 180 | -- |
| Informed consent form | X | | | | | | | | | |
| Study injection (including 30-minute post-dosing observation period) | | X | | | | | | | | |
| Confirm participant meets inclusion and exclusion criteria | X | | | | | | | | | |
| Physical examination including vital signs[3] | X | X | | | X | X | | | X | X |
| Pregnancy testing | | X | | | | | | | | |
| Blood for SARS-CoV-2 serology (antinucleocapsid antibody) | | X | | | X | X | | | X | X |
| Blood for immune response to vaccination (binding and neutralizing antibody)[4] | | X | | X | X | X | | | X | X |
| Nasopharyngeal swab sample for SARS-CoV-2[4] | | X | | | X | X | | | X | X[5] |

| Visit Number | A.2 Screening[1] | V6 | | V7 | V8 | V9 | | | V10 | A.2 UNS |
|---|---|---|---|---|---|---|---|---|---|---|
| **Type of Visit** | C | C | SC | C | C | C | SFU (eDiary) | SFU (SC) | C | C |
| **Month Timepoint** | M0 | M0 | | | M1 | M3 | | | M6 | -- |
| **Study Visit Day** | D0[1] | D1[1,9] | D8 | D15 | D29 | D91 | Every 2 weeks D36–D162[2] | Every 2 weeks D43-D169 | D181 | |
| **Window Allowance (Days)** | -7 | 0 | +3 | ±3 | -7 | ±7 | ±2 | ±3 | ±14 | -- |
| **Days Since Booster Injection** | | 0 | 7 | 14 | 28 | 90 | -- | | 180 | -- |
| eDiary activation for recording solicited local and systemic reactogenicity adverse reactions (7 days) | | X | | | | | | | | |
| Review of eDiary | | | X | | | | | | | |
| Follow-up safety calls[6] | | | X | | | | | X | | |
| Recording of unsolicited AEs | | X | X | X | X | | | | | |
| Recording of MAAEs and concomitant medications relevant to or for the treatment of the MAAE[7] | | X | X | X | X | X | X | X | X | |
| Recording of SAEs and concomitant medications relevant to or for the treatment of the SAE[7] | | X | X | X | X | X | X | X | X | |
| Recording of AESI | | X | X | X | X | X | X | X | X | |
| Recording of concomitant medications and non-study vaccinations[8] | | X | X | X | X | | | | X | |
| Study completion | | | | | | | | | X | |

Abbreviations: AE = adverse event; AESI = adverse event of special interest; C = clinic visit; COVID-19 = coronavirus disease 2019; D = day; eDiary = electronic diary; M = month; MAAE = medically attended adverse event; NP = nasopharyngeal; RT-PCR = reverse transcriptase polymerase chain reaction; SAE = serious adverse event; SARS-CoV-2 = severe acute respiratory syndrome coronavirus 2; SC = safety telephone call; SFU = safety follow-up; UNS = unscheduled visit; V = visit.

Note: In accordance with "FDA Guidance on Conduct of Clinical Trials of Medical Products during COVID-19 Public Health Emergency" (DHHS 2020), investigators may convert study site visits to telemedicine visits with the approval of the Sponsor.

1. The Screening Visit and Day 1 (vaccination) visit can be combined and occur on the same day.
2. Safety follow-up via eDiary questionnaire will be performed every 2 weeks from Day 36 to Day 162. These study days are relative to Day 1 vaccination. Adverse reactions recorded in the eDiary beyond Day 7 should be reviewed by the study site staff either during the next scheduled telephone call or at the next study site visit.
3. Physical examination: A full physical examination, including height and weight, will be performed on Day 1. Symptom-directed physical examinations may be performed at other timepoints at the discretion of the investigator. Any clinically significant finding identified during a study visit should be reported as an MAAE. Vital signs are to be collected pre- and post-dosing on the day of injection (Day 1) only. When applicable, vital sign measurements should be performed before blood collection. For participants who are febrile (body temperature ≥ 38.0°C [100.4°F]) before injection on Day 1, the visit must be rescheduled within the relevant window period to receive the injection. Afebrile participants with minor illnesses can be administered investigational product at the discretion of the investigator.
4. The NP swab sample, collected prior to injection on Day 1, will be used to ascertain the presence of SARS-CoV-2 via RT-PCR. The NP swab sample will also be collected within 24 hours if participant experience signs and symptoms of SARS-CoV-2 infection. It is important to note that some of the symptoms of COVID-19 overlap with solicited local and systemic reactogenicity ARs, that are expected after vaccination with mRNA-1273.211 (eg, myalgia, headache, fever, chills). During the first 7 days after vaccination, when these solicited ARs are common, investigators should use their clinical judgment to decide if an NP swab should be collected. The collection of a NP swab prior to the Day 1 vaccination can help ensure that cases of COVID-19 are not overlooked. Any study participant reporting respiratory symptoms during the 7-day period after injection should be evaluated for COVID-19.
5. Nasopharyngeal swabs will be collected during the unscheduled/illness visit for testing for the presence of SARS-CoV-2 via RT-PCR and for assessing non-SARS-CoV-2 causes of upper and lower respiratory tract infection.
6. Trained site personnel will call all participants to collect information relating to any AEs, MAAEs, SAEs, AEs leading to withdrawal, information on concomitant medications associated with those events, AESI, and any non-study vaccinations. In addition, study personnel will collect information on known participant exposure to someone with known COVID-19 or SARS-CoV-2 infection and on participant experience of COVID-19 symptoms. Sites will collect this information for eDiary days only if eDiary responses indicate the need for follow-up via telephone.
7. All concomitant medications relevant to or for the treatment of an SAE or MAAE will be recorded from Day 1 through the end of study visit (Day 181).
8. All concomitant medications and non-study vaccinations will be recorded through 28 days following injection.
9. If Part A.2 Day 1 falls within the visit window for Part A.1 D366, the study procedures listed for Part A.1 Day 366, Part A.2 Screening, and Part A.2 Day 1 can be completed on the same day. All Part A.1 Day 366 samples will be collected prior to administration of mRNA-1273.214. Additional samples would not be collected for Part A.2 Day 1 if it occurs on the same day as Part A.1 Day 366. The samples collected pre-dose will be used for both Part A.1 Day 366 and Part A.2 Day 1.

**Table 19: Schedule of Events for Part H**

| **Visit Number** | H Screening[1] | V1 | | V2 | V3 | V3a | | | V4 | H UNS |
|---|---|---|---|---|---|---|---|---|---|---|
| **Type of Visit** | C | C | SC | C | C | C | SFU (eDiary) | SFU (SC) | C | C |
| **Month Timepoint** | M0 | M0 | | | M1 | M3 | | | M6 | -- |
| **Study Visit Day** | D0[1] | D1[1] | D8 | D15 | D29 | D91 | Every 2 weeks D36-D162[2] | Every 2 weeks D43-D169 | D181 | |
| **Window Allowance (Days)** | -7 | 0 | +3 | ±3 | -7 | ±7 | ±2 | ±3 | ±14 | -- |
| **Days Since Booster Injection** | | 0 | 7 | 14 | 28 | 90 | -- | | 180 | -- |
| Informed consent form | X | | | | | | | | | |
| Study injection (including 30-minute post-dosing observation period) | | X | | | | | | | | |
| Confirm participant meets inclusion and exclusion criteria | X | | | | | | | | | |
| Physical examination including vital signs[3] | X | X | | | X | X | | | X | X |
| Pregnancy testing | | X | | | | | | | | |
| Blood for SARS-CoV-2 serology (antinucleocapsid antibody) | | X | | | X | X | | | X | X |
| Blood for immune response to vaccination (binding and neutralizing antibody)[4] | | X | | X | X | X | | | X | X |
| Nasopharyngeal swab sample for SARS-CoV-2[4] | | X | | | X | X | | | X | X[5] |
| eDiary activation for recording solicited local and systemic reactogenicity adverse reactions (7 days) | | X | | | | | | | | |

| **Visit Number** | H Screening[1] | V1 | | V2 | V3 | V3a | | | V4 | H UNS |
|---|---|---|---|---|---|---|---|---|---|---|
| **Type of Visit** | C | C | SC | C | C | C | SFU (eDiary) | SFU (SC) | C | C |
| **Month Timepoint** | M0 | M0 | | | M1 | M3 | | | M6 | -- |
| **Study Visit Day** | D0[1] | D1[1] | D8 | D15 | D29 | D91 | Every 2 weeks D36-D162[2] | Every 2 weeks D43-D169 | D181 | |
| **Window Allowance (Days)** | -7 | 0 | +3 | ±3 | -7 | ±7 | ±2 | ±3 | ±14 | -- |
| **Days Since Booster Injection** | | 0 | 7 | 14 | 28 | 90 | -- | | 180 | -- |
| Review of eDiary | | | X | | | | | | | |
| Follow-up safety calls[6] | | | X | | | | | X | | |
| Recording of unsolicited AEs | | X | X | X | X | | | | | |
| Recording of MAAEs and concomitant medications relevant to or for the treatment of the MAAE[7] | | X | X | X | X | X | X | X | X | |
| Recording of SAEs and concomitant medications relevant to or for the treatment of the SAE[7] | | X | X | X | X | X | X | X | X | |
| Recording of AESI | | X | X | X | X | X | X | X | X | |
| Recording of concomitant medications and non-study vaccinations[8] | | X | X | X | X | | | | X | |
| Study completion | | | | | | | | | X | |

Abbreviations: AE = adverse event; AESI = adverse event of special interest; C = clinic visit; COVID-19 = coronavirus disease 2019; D = day; eDiary = electronic diary; M = month; MAAE = medically attended adverse event; NP = nasopharyngeal; RT-PCR = reverse transcriptase polymerase chain reaction; SAE = serious adverse event; SARS-CoV-2 = severe acute respiratory syndrome coronavirus 2; SC = safety telephone call; SFU = safety follow-up; UNS = unscheduled visit; V = visit.

Note: In accordance with “FDA Guidance on Conduct of Clinical Trials of Medical Products during COVID-19 Public Health Emergency” (DHHS 2020), investigators may convert study site visits to telemedicine visits with the approval of the Sponsor.

1. The Screening Visit and Day 1 (vaccination) visit can be combined and occur on the same day.
2. Safety follow-up via eDiary questionnaire will be performed every 2 weeks from at Day 36 to Day 162. These study days are relative to Day 1 vaccination. Adverse reactions recorded in the eDiary beyond Day 7 should be reviewed by the study site staff either during the next scheduled telephone call or at the next study site visit.
3. Physical examination: A full physical examination, including height and weight, will be performed on Day 1. Symptom-directed physical examinations may be performed at other timepoints at the discretion of the investigator. Any clinically significant finding identified during a study visit should be reported as an MAAE. Vital signs are to be collected pre- and post-dosing on the day of injection (Day 1) only. When applicable, vital sign measurements should be performed before blood collection. For participants who are febrile (body temperature ≥ 38.0°C [100.4°F]) before injection on Day 1, the visit must be rescheduled within the relevant window period to receive the injection. Afebrile participants with minor illnesses can be administered investigational product at the discretion of the investigator.
4. The NP swab sample, collected prior to injection on Day 1, will be used to ascertain the presence of SARS-CoV-2 via RT-PCR. The NP swab sample will also be collected within 24 hours if a participant experiences signs and symptoms of SARS-CoV-2 infection. It is important to note that some of the symptoms of COVID-19 overlap with solicited local and systemic reactogenicity ARs, which are expected after vaccination with mRNA-1273.222 (eg, myalgia, headache, fever, chills). During the first 7 days after vaccination, when these solicited ARs are common, investigators should use their clinical judgment to decide if an NP swab should be collected. The collection of a NP swab prior to the Day 1 vaccination can help ensure that cases of COVID-19 are not overlooked. Any study participant reporting respiratory symptoms during the 7-day period after injection should be evaluated for COVID-19.
5. Nasopharyngeal swabs will be collected during the unscheduled/illness visit for testing for the presence of SARS-CoV-2 via RT-PCR and for assessing non-SARS-CoV-2 causes of upper and lower respiratory tract infection.
6. Trained site personnel will call all participants to collect information relating to any AEs, MAAEs, SAEs, AEs leading to withdrawal, information on concomitant medications associated with those events, AESI, and any non-study vaccinations. In addition, study personnel will collect information on known participant exposure to someone with known COVID-19 or SARS-CoV-2 infection and on participant experience of COVID-19 symptoms. Sites will collect this information for eDiary days only if eDiary responses indicate the need for follow up via telephone.
7. All concomitant medications relevant to or for the treatment of an SAE or MAAE will be recorded from Day 1 through the end of study visit (Day 181).
8. All concomitant medications and non-study vaccinations will be recorded through 28 days following injection.

**Table 20: Schedule of Events for Part J**

| **Visit Number** | J Screening[1] | V1 | | V2 | V3 | V3a | | | V4 | J UNS |
|---|---|---|---|---|---|---|---|---|---|---|
| **Type of Visit** | C | C | SC | C | C | C | SFU (eDiary) | SFU (SC) | C | C |
| **Month Timepoint** | M0 | M0 | | | M1 | M3 | | | M6 | -- |
| **Study Visit Day** | D0[1] | D1[1] | D8 | D15 | D29 | D91 | Every 2 weeks D36-D162[2] | Every 2 weeks D43-D169 | D181 | |
| **Window Allowance (Days)** | -7 | 0 | +3 | ±3 | -7 | ±7 | ±2 | ±3 | ±14 | -- |
| **Days Since Booster Injection** | | 0 | 7 | 14 | 28 | 90 | -- | | 180 | -- |
| Informed consent form | X | | | | | | | | | |
| Randomization for treatment assignment and study injection (including 30-minute post-dosing observation period) | | X | | | | | | | | |
| Confirm participant meets inclusion and exclusion criteria | X | | | | | | | | | |
| Physical examination including vital signs[3] | X | X | | | X | X | | | X | X |
| Pregnancy testing | | X | | | | | | | | |
| Blood for SARS-CoV-2 serology (antinucleocapsid antibody) | | X | | | X | X | | | X | X |
| Blood for immune response to vaccination (binding and neutralizing antibody)[4] | | X | | X | X | X | | | X | X |
| Nasopharyngeal swab sample for SARS-CoV-2[4] | | X | | | X | X | | | X | X[5] |
| eDiary activation for recording solicited local and systemic reactogenicity adverse reactions (7 days) | | X | | | | | | | | |

| **Visit Number** | J Screening[1] | V1 | | V2 | V3 | V3a | | | V4 | J UNS |
|---|---|---|---|---|---|---|---|---|---|---|
| **Type of Visit** | C | C | SC | C | C | C | SFU (eDiary) | SFU (SC) | C | C |
| **Month Timepoint** | M0 | M0 | | | M1 | M3 | | | M6 | -- |
| **Study Visit Day** | D0[1] | D1[1] | D8 | D15 | D29 | D91 | Every 2 weeks D36-D162[2] | Every 2 weeks D43-D169 | D181 | |
| **Window Allowance (Days)** | -7 | 0 | +3 | ±3 | -7 | ±7 | ±2 | ±3 | ±14 | -- |
| **Days Since Booster Injection** | | 0 | 7 | 14 | 28 | 90 | -- | | 180 | -- |
| Review of eDiary | | | X | | | | | | | |
| Follow-up safety calls[6] | | | X | | | | | X | | |
| Recording of unsolicited AEs | | X | X | X | X | | | | | |
| Recording of MAAEs and concomitant medications relevant to or for the treatment of the MAAE[7] | | X | X | X | X | X | X | X | X | |
| Recording of SAEs and concomitant medications relevant to or for the treatment of the SAE[7] | | X | X | X | X | X | X | X | X | |
| Recording of AESI and AEs leading to withdrawal from study | | X | X | X | X | X | X | X | X | |
| Recording of concomitant medications and non-study vaccinations[8] | | X | X | X | X | | | | X | |
| Study completion | | | | | | | | | X | |

Abbreviations: AE = adverse event; AESI = adverse event of special interest; C = clinic visit; COVID-19 = coronavirus disease 2019; D = day; eDiary = electronic diary; M = month; MAAE = medically attended adverse event; NP = nasopharyngeal; RT-PCR = reverse transcriptase polymerase chain reaction; SAE = serious adverse event; SARS-CoV-2 = severe acute respiratory syndrome coronavirus 2; SC = safety telephone call; SFU = safety follow-up; UNS = unscheduled visit; V = visit.

Note: In accordance with "FDA Guidance on Conduct of Clinical Trials of Medical Products during COVID-19 Public Health Emergency" (DHHS 2020), investigators may convert study site visits to telemedicine visits with the approval of the Sponsor.

1. The Screening Visit and Day 1 (vaccination) visit can be combined and occur on the same day.

2. Safety follow-up via eDiary questionnaire will be performed every 2 weeks from at Day 36 to Day 162. These study days are relative to Day 1 vaccination. Adverse reactions recorded in the eDiary beyond Day 7 should be reviewed by the study site staff either during the next scheduled safety telephone call or at the next study site visit.
3. Physical examination: A full physical examination, including height and weight, will be performed on Day 1. Symptom-directed physical examinations may be performed at other timepoints at the discretion of the investigator. Any clinically significant finding identified during a study visit should be reported as an MAAE. Vital signs are to be collected pre- and post-dosing on the day of injection (Day 1) only. When applicable, vital sign measurements should be performed before blood collection. For participants who are febrile (body temperature ≥ 38.0°C [100.4°F]) before injection on Day 1, the visit must be rescheduled within the relevant window period to receive the injection. Afebrile participants with minor illnesses can be administered investigational product at the discretion of the investigator.
4. The NP swab sample, collected prior to injection on Day 1, will be used to ascertain the presence of SARS-CoV-2 via RT-PCR. The NP swab sample will also be collected within 24 hours if a participant experiences signs and symptoms of SARS-CoV-2 infection. It is important to note that some of the symptoms of COVID-19 overlap with solicited local and systemic reactogenicity ARs, which are expected after vaccination with mRNA-1273.222 (eg, myalgia, headache, fever, chills). During the first 7 days after vaccination, when these solicited ARs are common, investigators should use their clinical judgment to decide if an NP swab should be collected. The collection of a NP swab prior to the Day 1 vaccination can help ensure that cases of COVID-19 are not overlooked. Any study participant reporting respiratory symptoms during the 7-day period after injection should be evaluated for COVID-19.
5. Nasopharyngeal swabs will be collected during the unscheduled/illness visit for testing for the presence of SARS-CoV-2 via RT-PCR and for assessing non-SARS-CoV-2 causes of upper and lower respiratory tract infection.
6. Trained site personnel will call all participants to collect information relating to any AEs, MAAEs, SAEs, AESI, AEs leading to withdrawal from study participation, information on concomitant medications associated with those events, AESI, and any non-study vaccinations. In addition, study personnel will collect information on known participant exposure to someone with known COVID-19 or SARS-CoV-2 infection and on participant experience of COVID-19 symptoms. Sites will collect this information for eDiary days only if eDiary responses indicate the need for follow up via telephone.
7. All concomitant medications relevant to or for the treatment of an SAE or MAAE will be recorded from Day 1 through the end of study visit (Day 181).
8. All concomitant medications and non-study vaccinations will be recorded through 28 days following injection.

#### 10.2. APPENDIX 2: Study Governance Considerations

##### 10.2.1. Regulatory and Ethical Considerations

This study will be conducted in accordance with the protocol and with the following:

- Consensus ethical principles derived from international guidelines including the Declaration of Helsinki and Council for International Organizations of Medical Sciences International Ethical Guidelines.
- Applicable ICH GCP Guidelines.
- Applicable laws and regulatory requirements.
- The protocol, protocol amendments, ICF, IB, and other relevant documents (eg, advertisements) must be submitted to an IRB by the investigator and reviewed and approved by the IRB before the study is initiated.
- Any amendments to the protocol will require IRB approval before implementation of changes made to the study design, except for changes necessary to eliminate an immediate hazard to study participants.
- The investigator will be responsible for the following:
  - Providing written summaries of the status of the study to the IRB annually or more frequently in accordance with the requirements, policies, and procedures established by the IRB
  - Notifying the IRB of SAEs or other significant safety findings as required by IRB procedures
  - Providing oversight of the conduct of the study at the site and adherence to requirements of 21 CFR, ICH guidelines, the IRB, European regulation 536/2014 for clinical studies (if applicable), and all other applicable local regulations

##### 10.2.2. Study Monitoring

Before an investigational site can enter a participant into the study, a representative of the Sponsor or its representatives will visit the investigational study site to:

- Determine the adequacy of the facilities.
- Discuss with the investigator(s) and other personnel their responsibilities with regard to protocol adherence, and the responsibilities of the Sponsor or its representatives. This will be documented in a Clinical Study Agreement between the Sponsor, the designated CRO, and the investigator.

According to ICH GCP guideline, the Sponsor of the study is responsible for ensuring the proper conduct of the study with regard to protocol adherence and validity of data recorded on the eCRFs. The study monitor's duties are to aid the investigator and the Sponsor in the maintenance of complete, accurate, legible, well-organized, and easily retrievable data. The study monitor will advise the investigator of the regulatory necessity for study-related monitoring, audits, IRB review, and inspection by providing direct access to the source data and/or documents. In addition, the study monitor will explain to and interpret for the investigator all regulations applicable to the clinical evaluation of an IP as documented in ICH guidelines.

It is the study monitor's responsibility to inspect the eCRFs and source documentation throughout the study to protect the rights of the participants; to verify adherence to the protocol; to verify completeness, accuracy, and consistency of the data; and to confirm adherence of study conduct to any local regulations. Details will be outlined in the clinical monitoring plan. During the study, a monitor from the Sponsor or a representative will have regular contacts with the investigational site, for the following:

- Provide information and support to the investigator(s).
- Confirm that facilities remain acceptable.
- Confirm that the investigational team is adhering to the protocol, that the data are being accurately recorded in the eCRFs, and that IP accountability checks are being performed.
- Perform source data verification. This includes a comparison of the data in the eCRFs with the participant's medical records at the hospital or practice, and other records relevant to the study. This will require direct access to all original records for each participant (eg, clinical charts or electronic medical record system).
- Record and report any protocol deviations not previously sent.
- Confirm AEs and SAEs have been properly documented on eCRFs and confirm any SAEs have been forwarded to the SAE Mailbox, and those SAEs that met criteria for reporting have been forwarded to the IRB.

The monitor will be available between visits if the investigator(s) or other staff needs information or advice.

###### 10.2.3. Audits and Inspections

The Sponsor, their designee(s), the IRB, or regulatory authorities will be allowed to conduct site visits to the investigational facilities for the purpose of monitoring or inspecting any aspect of the study. The investigator agrees to allow the Sponsor, their designee(s), the IRB, or regulatory authorities to inspect the IP storage area, IP stocks, IP records, participant charts and study source documents, and other records relative to study conduct.

Authorized representatives of the Sponsor, a regulatory authority, and the IRB may visit the site to perform audits or inspections, including source data verification. The purpose of a Sponsor audit or inspection is to systematically and independently examine all study-related activities and documents to determine whether these activities were conducted, and data were recorded, analyzed, and accurately reported according to the protocol, ICH GCP (R2), and any applicable regulatory requirements. The investigator should contact the Sponsor immediately if contacted by a regulatory agency about an inspection.

The principal investigator must obtain IRB approval for the investigation. Initial IRB approval and all materials approved by the IRB for this study including the participant consent form and recruitment materials must be maintained by the investigator and made available for inspection.

##### 10.2.4. Financial Disclosure

The investigator is required to provide financial disclosure information to allow the Sponsor to submit the complete and accurate certification or disclosure statements required under 21 CFR 54. In addition, the investigator must provide the Sponsor with a commitment to promptly update this information if any relevant changes occur during the course of the investigation and for 1 year following the completion of the study.

The Sponsor, the CRO, and the study site are not financially responsible for further testing or treatment of any medical condition that may be detected during the screening process. In addition, in the absence of specific arrangements, the Sponsor, the CRO, and the study site are not financially responsible for further treatment of the disease under study.

##### 10.2.5. Recruitment Procedures

Advertisements to be used for the recruitment of study participants and any other written information regarding this study to be provided to the participant should be submitted to the Sponsor for approval. All documents must be approved by the IRB.

##### 10.2.6. Informed Consent/Assent Process

The informed consent document(s) must meet the requirements of 21 CFR 50, local regulations, ICH guidelines, Health Insurance Portability and Accountability Act (HIPAA) requirements, where applicable, and the IRB or study center. All consent documents will be approved by the appropriate IRB. The actual ICF used at each center may differ, depending on local regulations and IRB requirements. However, all versions must contain the standard information found in the sample ICF provided by the Sponsor. Any change to the content of the ICF must be approved by the Sponsor and the IRB prior to the form being used.

If new information becomes available that may be relevant to the participant’s willingness to continue participation in the study, this will be communicated to them in a timely manner. Such information will be provided via a revised ICF or an addendum to the original ICF.

The investigator or his/her representative will explain the nature of the study to the participant and answer all questions regarding the study.

The investigator is responsible for ensuring that the participant fully understands the nature and purpose of the study. Information should be given in both oral and written form whenever possible.

No participant should be obliged to participate in the study. The participant must be informed that participation is voluntary. Participants, their relatives, guardians, or (if applicable) legal representatives must be given ample opportunity to inquire about details of the study. The information must make clear that refusal to participate in the study or withdrawal from the study at any stage is without any prejudice to the participant’s subsequent care.

The participant must be allowed sufficient time to decide whether they wish to participate.

The participant must be made aware of and give consent to direct access to his/her source medical records by study monitors, auditors, the IRB, and regulatory authorities. The participant should be informed that such access will not violate participant confidentiality or any applicable regulations. The participant should also be informed that he/she is authorizing such access by signing the ICF.

A copy of the ICF(s) must be provided to the participant.

A participant who is rescreened is not required to sign another ICF if the rescreening occurs within 28 days from the previous ICF signature date (within the initial screening period).

The ICF will also explain that excess serum from immunogenicity testing may be used for future research, which may be performed at the discretion of the Sponsor to further characterize the immune response to SARS-CoV-2, additional assay development, and the immune response across CoV.

##### 10.2.7. Protocol Amendments

No change or amendment to this protocol may be made by the investigator or the Sponsor after the protocol has been agreed to and signed by all parties unless such change(s) or amendment(s) has (have) been agreed upon by the investigator or the Sponsor. Any change agreed upon will be recorded in writing, and the written amendment will be signed by the investigator and the Sponsor. Institutional review board approval is required prior to the implementation of an amendment, unless overriding safety reasons warrant immediate action, in which case the IRB(s) will be promptly notified.

Any modifications to the protocol or the ICF, which may impact the conduct of the study, potential benefit of the study, or may affect participant safety, including changes of study objectives, study design, participant population, sample sizes, study procedures, or significant administrative aspects will require a formal amendment to the protocol. Such amendment will be released by the Sponsor, agreed by the investigator(s), and approved by the relevant IRB(s) prior to implementation. A signed and dated statement that the protocol, any subsequent relevant amended documents and the ICF have been approved by relevant IRB(s) must be provided to the Sponsor before the study is initiated.

Administrative changes to the protocol are minor corrections and/or clarifications that have no effect on the way the study is to be conducted. These administrative changes will be released by the Sponsor, agreed by the investigators, and notified to the IRB(s).

##### 10.2.8. Protocol Deviations

Noncompliance may be either on the part of the participant, the investigator, or the study site staff. As a result of deviations, corrective actions are to be developed by the site and implemented promptly.

It is the responsibility of the site investigator to use continuous vigilance to identify and report deviations to the Sponsor or its designee. All deviations must be addressed in study source documents and reported to the study monitor. Protocol deviations must be sent to the reviewing IRB per their policies. The site investigator is responsible for knowing and adhering to the reviewing IRB requirements.

##### 10.2.9. Data Protection

Participants will be assigned a unique identifier by the Sponsor. Any participant records or datasets that are transferred to the Sponsor will contain the identifier only; participant names or any information which would make the participant identifiable will not be transferred.

The participant must be informed that his/her personal study-related data will be used by the Sponsor in accordance with local data protection law. The level of disclosure must also be explained to the participant.

The participant must be informed that his/her medical records may be examined by Clinical Quality Assurance auditors or other authorized personnel appointed by the Sponsor, by appropriate IRB members, and by inspectors from regulatory authorities.

Individual participant medical information obtained as a result of this study is considered confidential, and disclosure to third parties is prohibited. Information will be accessible to authorized parties or personnel only. Medical information may be given to the participant's physician or to other appropriate medical personnel responsible for the participant's well-being. Each participant will be asked to complete a form allowing the investigator to notify the participant's primary health care provider of his/her participation in this study.

All laboratory specimens, evaluation forms, reports, and other records will be identified in a manner designed to maintain participant confidentiality. All records will be kept in a secure storage area with limited access. Clinical information will not be released without the written permission of the participant, except as necessary for monitoring and auditing by the Sponsor, its designee, the relevant regulatory authority, or the IRB.

The investigator and all employees and coworkers involved with this study may not disclose or use for any purpose other than performance of the study, any data, record, or other unpublished, confidential information disclosed to those individuals for the purpose of the study. Prior written agreement from the Sponsor or its designee must be obtained for the disclosure of any confidential information to other parties.

##### 10.2.10. Sample Retention and Future Biomedical Research

The retention period of laboratory samples will be 20 years, or as permitted by local regulations, to address further scientific questions related to mRNA-1273.211 or anti-respiratory virus immune response. In addition, identifiable samples can be destroyed at any time at the request of the participant. During the study, or during the retention period, in addition to the analysis outlined in the study endpoints, exploratory analysis may be conducted using other measures of adaptive immunity to SARS-CoV-2 to include humoral and cellular immune assay methodologies on any remaining blood or serum samples, including samples from participants who are screened but are not subsequently enrolled. These analyses will extend the search for other potentially relevant biomarkers to investigate the effect of mRNA-1273.211 as well as to determine how changes in biomarkers may relate to exposure and clinical outcomes. A decision to perform such exploratory research may arise from new scientific findings related to the drug class or disease, as well as reagent and assay availability.

##### 10.2.11. Safety Oversight

Safety monitoring for the study is described in Section 7.5.

##### 10.2.12. Dissemination of Clinical Study Data

The Sponsor shares information about clinical trials and results on publicly accessible websites, based on international and local legal and regulatory requirements, and other clinical trial disclosure commitments established by pharmaceutical industry associations. These websites include clinicaltrials.gov, EU clinical trial register (eu.ctr), as well as some national registries.

##### 10.2.13. Data Quality Assurance and Quality Control

Data collection is the responsibility of the clinical study staff at the site under the supervision of the site investigator. The investigator is responsible for ensuring the accuracy, completeness, legibility, and timeliness of the data reported.

- All participant data relating to the study will be recorded in the eCRF unless transmitted to the Sponsor or designee electronically (eg, laboratory data). The investigator is responsible for verifying that data entries are accurate and correct by physically or electronically signing the eCRF.
- The investigator must maintain accurate documentation (source data) that supports the information entered in the eCRF.
- The investigator must permit study-related monitoring, audits, IRB review, and regulatory agency inspections and provide direct access to source data documents.
- Monitoring details describing strategy (eg, risk-based initiatives in operations and quality such as Risk Management and Mitigation Strategies and Analytical Risk-Based Monitoring), methods, responsibilities and requirements, including handling of noncompliance issues and monitoring techniques (central, remote, or onsite monitoring) are provided in the clinical monitoring plan.
- The Sponsor or designee is responsible for the data management of this study including quality checking of the data.
- The Sponsor assumes accountability for actions delegated to other individuals (eg, CROs).
- Study monitors will perform ongoing source data verification to confirm that data entered into the eCRF by authorized site personnel are accurate, complete, and verifiable from source documents; that the safety and rights of participants are being protected; and that the study is being conducted in accordance with the currently approved protocol and any other study agreements, ICH GCP, and all applicable regulatory requirements.
- Records and documents, including signed ICFs, pertaining to the conduct of this study must be retained by the investigator for a period of at least 2 years after the last marketing application approval or, if not approved, 2 years following the discontinuance of the test article for investigation. If this requirement differs from any local regulations, the local regulations will take precedence unless the local retention policy is less than 2 years. No records may be destroyed during the retention period without the written approval of the Sponsor. No records may be transferred to another location or party without written notification to the Sponsor.

Quality assurance includes all the planned and systematic actions that are established to ensure that the clinical study is performed and the data are generated, documented (recorded), and reported according to ICH GCP and local/regional regulatory standards.

A quality assurance representative from Sponsor or qualified designee, who is independent of and separated from routine monitoring, may periodically arrange inspections/audits of the clinical study by reviewing the data obtained and procedural aspects. These inspections may include onsite inspections/audits and source data checks. Direct access to source documents is required for the purpose of these periodic inspections/audits.

##### 10.2.14. Data Collection and Management

This study will be conducted in compliance with ICH CGP guidelines. This study will also be conducted in accordance with the most recent version of the Declaration of Helsinki.

This study will use electronic data collection to collect data directly from the study site using eCRFs. The investigator is responsible for ensuring that all sections of each eCRF are completed promptly and correctly and that entries can be verified against any source data.

Study monitors will perform source document verification to identify inconsistencies between the eCRFs and source documents. Discrepancies will be resolved in accordance with the principles of GCP. Detailed study monitoring procedures are provided in the clinical monitoring plan.

AEs will be coded with MedDRA. Concomitant medications will be coded using WHO – Drug Reference List.

##### 10.2.15. Source Documents

Source documents are original documents or certified copies, and include, but are not limited to, eDiaries, medical and hospital records, screening logs, ICFs, telephone contact logs, and worksheets. Source documents provide evidence for the existence of the participant and substantiate the integrity of the data collected. Source documents are filed at the investigator’s site.

Data reported on the case report form or entered in the eCRF that are transcribed from source documents must be consistent with the source documents or the discrepancies must be explained. The investigator may need to request previous medical records or transfer records, depending on the study. Also, current medical records must be available.

The Sponsor or its designee requires that the investigator prepare and maintain adequate and accurate records for each participant treated with the IP. Source documents such as any hospital, clinic, or office charts and the signed ICFs are to be included in the investigator’s files with the participant’s study records.

##### 10.2.16. Retention of Records

The principal investigator must maintain all documentation relating to the study for a period of at least 2 years after the last marketing application approval or, if not approved, 2 years following the discontinuance of the test article for investigation. If this requirement differs from any local regulations, the local regulations will take precedence unless the local retention policy is less than 2 years.

If it becomes necessary for the Sponsor or the regulatory authority to review any documentation relating to the study, the investigator must permit access to such records. No records will be destroyed without the written consent of the Sponsor, if applicable. It is the responsibility of the Sponsor to inform the investigator when these documents no longer need to be retained.

##### 10.2.17. Study and Site Closure

If the study is prematurely terminated or suspended, the Sponsor shall promptly inform the investigators, the IRBs, the regulatory authorities, and any CRO(s) used in the study of the reason for termination or suspension, as specified by the applicable regulatory requirements. The investigator shall promptly inform the participant and should assure appropriate participant therapy and/or follow-up.

The Sponsor or designee reserves the right to close the study site or terminate the study at any time for any reason at the sole discretion of the Sponsor.

The investigator may initiate study site closure at any time, provided there is reasonable cause and sufficient notice is given in advance of the intended termination.

Reasons for the early closure of a study site by the Sponsor or investigator may include but are not limited to:

- Continuation of the study represents a significant medical risk to participants
- Failure of the investigator to comply with the protocol, the requirements of the IRB or local health authorities, the Sponsor's procedures, or GCP guidelines
- Inadequate recruitment of participants by the investigator
- Discontinuation of further mRNA-1273.211 development

Study sites will be closed upon study completion. A study site is considered closed when all required documents and study supplies have been collected and a study site closure visit has been performed.

##### 10.2.18. Publication Policy

The results of this study may be published or presented at scientific meetings. If this is foreseen, the investigator agrees to submit all manuscripts or abstracts to the Sponsor before submission. This allows the Sponsor to protect proprietary information and to provide comments.

The Sponsor will comply with the requirements for publication of study results. In accordance with standard editorial and ethical practice, the Sponsor will generally support publication of multicenter studies only in their entirety and not as individual site data. In this case, a coordinating investigator will be designated by mutual agreement.

Authorship will be determined by mutual agreement and in line with International Committee of Medical Journal Editors authorship requirements.

The clinical study plan and the results of the study will be published on www.ClinicalTrials.gov in accordance with 21 CFR 50.25(c). The results of and data from this study belong to the Sponsor.

#### 10.3. APPENDIX 3: Contraceptive Guidance

**Definitions: Woman of Childbearing Potential**

Women of childbearing potential are those who are considered fertile following menarche and until becoming postmenopausal unless permanently sterile (see below). If fertility is unclear (eg, amenorrhea in adolescents or athletes) and a menstrual cycle cannot be confirmed before vaccination at Day 1, additional evaluation should be considered.

Women in the following categories are not considered women of childbearing potential:

1. Premenarchal
2. Premenopausal, surgically sterile female with 1 of the following:
    a. Documented complete hysterectomy
    b. Documented surgical sterilization

    For individuals with permanent infertility due to an alternate medical cause other than the above (eg, Müllerian agenesis, androgen insensitivity), investigator discretion should be applied to determining study entry.

    Note: Documentation can come from the site personnel's review of the participant's medical records, medical examination, or medical history interview.
3. Postmenopausal female
    - A postmenopausal state is defined as no menses for 12 months without an alternative medical cause. The following age-specific requirements apply:
        – Women < 50 years of age would be considered postmenopausal if they have been amenorrheic for 12 months or more following cessation of exogenous hormonal treatments and if they have luteinizing hormone and FSH levels in the postmenopausal range for the institution.
        – Women ≥ 50 years of age would be considered postmenopausal if they have been amenorrheic for 12 months or more, had radiation-induced menopause with last menses > 1 year ago, had chemotherapy-induced menopause with last menses > 1 year ago.
    - A high FSH level in the postmenopausal range may be used to confirm a postmenopausal state in women not using hormonal replacement therapy (HRT).
    - Females on HRT and whose menopausal status is in doubt will be required to use one of the nonestrogen hormonal highly effective contraception methods if they wish to continue their HRT during the study. Otherwise, they must discontinue HRT to allow confirmation of postmenopausal status before study enrollment.

**Contraception Guidance:**

Adequate female contraception is defined as consistent and correct use of an FDA-approved contraceptive method in accordance with the product label. For example:

- Barrier method (such as condoms, diaphragm, or cervical cap) used in conjunction with spermicide

- Intrauterine device
- Prescription hormonal contraceptive taken or administered via oral (pill), transdermal (patch), subdermal, or IM route
- Sterilization of a female participant's monogamous male partner prior to entry into the study

Note that periodic abstinence (eg, calendar, ovulation, symptothermal, postovulation methods) and withdrawal are not acceptable methods of contraception.

#### 10.4. APPENDIX 4: Adverse Events of Special Interest Terms

**The Investigator's medical judgment must be applied to assess an event as an AESI, as most AESI are based on medical concepts. The table below does not provide a comprehensive list of terms.**

The following table describes events/medical concepts that are of interest in COVID-19 vaccine safety surveillance. Some are specific to vaccines; however, some are of interest due to their occurrence in the context of concurrent or recent COVID-19. Events falling into the descriptions below should be reported as AESI, per protocol, even when they occur during/following COVID infection.

**Please note: COVID-19 itself is not an AESI.**

| Medical Concept | Medical Concept Descriptions/Guidance |
|---|---|
| **Anosmia, Ageusia** | New onset of anosmia or ageusia associated with COVID-19 or idiopathic etiology.<br>DOES NOT INCLUDE anosmia or ageusia associated with sinus/nasal congestion, congenital, or traumatic etiologies |
| **Subacute thyroiditis** | Acute inflammatory disease of the thyroid (immune-mediated or idiopathic).<br>DOES NOT INCLUDE new onset of chronic thyroiditis |
| **Acute pancreatitis** | New onset of pancreatitis in the absence of a clear, alternate etiology, such as alcohol, gallstones, trauma, recent invasive procedure, etc. |
| **Appendicitis** | Any event of appendicitis |
| **Rhabdomyolysis** | New onset of rhabdomyolysis in the absence of a clear, alternate etiology, such as drug/alcohol abuse, excessive exercise, trauma, etc. |
| **Acute respiratory distress syndrome (ARDS)** | New onset of ARDS/respiratory failure due to acute inflammatory lung injury.<br>DOES NOT INCLUDE non-specific symptoms of shortness of breath or dyspnea, nor events with underlying etiologies of heart failure or fluid overload |
| **Coagulation disorders** | New onset of thrombosis, thromboembolic event, or non-traumatic hemorrhage/bleeding disorder (eg, stroke, DVT, pulmonary embolism, disseminated intravascular coagulation (DIC), etc.) |
| **Acute cardiovascular injury** | New onset of clinically confirmed, acute cardiovascular injury, such as myocarditis, pericarditis, arrhythmia confirmed by ECG (eg, atrial fibrillation, atrial flutter, supraventricular tachycardia), stress cardiomyopathy, heart failure, acute coronary syndrome, myocardial infarction, etc.<br>DOES NOT INCLUDE transient sinus tachycardia/bradycardia, non-specific symptoms such as palpitations, racing heart, heart fluttering or pounding, irregular heartbeats, shortness of breath, chest pain/discomfort, etc. |
| **Acute kidney injury** | New onset of acute kidney injury or acute renal failure in the absence of a clear, alternate etiology, such as urinary tract infection/urosepsis, trauma, tumor, nephrotoxic medications/substances, etc.;<br>Increase in serum creatinine by ≥ 0.3 mg/dl (or ≥26.5 μmol/l) within 48 hours; OR<br>Increase in serum creatinine to ≥ 1.5 times baseline, known or presumed to have occurred within prior 7 days |

| Medical Concept | Medical Concept Descriptions/Guidance |
|---|---|
| **Acute liver injury** | New onset in the absence of a clear, alternate etiology, such as trauma, tumor, hepatotoxic medications/substances, etc.:<br>>3-fold elevation above the upper normal limit for ALT or AST; OR<br>>2-fold elevation above the upper normal limit for total serum bilirubin or GGT or ALP |
| **Dermatologic findings** | Chilblain-like lesions<br>Single organ cutaneous vasculitis<br>Erythema multiforme<br>Bullous rash<br>Severe cutaneous adverse reactions, such as Stevens-Johnson syndrome, Toxic epidermal necrolysis, Drug reaction with eosinophilia and systemic symptoms (DRESS), fixed drug eruptions, and necrotic or exfoliative reactions |
| **Systemic inflammatory syndromes** | Multisystem inflammatory syndrome in adults (MIS-A) or children (MIS-C)<br>Kawasaki's disease<br>Hemophagocytic lymphohistiocytosis (HLH) |
| **Thrombocytopenia** | Platelet count < 150 x $10^9$/L (thrombocytopenia)<br>New clinical diagnosis, or worsening, of thrombocytopenic condition, such as immune thrombocytopenia, thrombocytopenic purpura, or HELLP syndrome |
| **Acute aseptic arthritis** | Clinical syndrome characterized by acute onset of signs and symptoms of joint inflammation without recent trauma for a period of no longer than 6 weeks, synovial increased leukocyte count and the absence of microorganisms on Gram stain, routine culture and/or PCR.<br>DOES NOT INCLUDE new onset of chronic arthritic conditions |
| **New onset, or worsening, of neurological disease** | Immune-mediated neurological disorders<br>Guillain-Barre Syndrome<br>Acute disseminated encephalomyelitis (ADEM)<br>Peripheral facial nerve palsy (Bell's palsy)<br>Transverse myelitis<br>Encephalitis/Encephalomyelitis<br>Aseptic meningitis<br>Seizures/convulsions/epilepsy<br>Narcolepsy/hypersomnia |
| **Anaphylaxis** | Anaphylaxis associated with study drug administration |
| **Other syndromes** | Fibromyalgia<br>Postural Orthostatic Tachycardia Syndrome<br>Chronic Fatigue Syndrome<br>Myalgic encephalomyelitis<br>Post viral fatigue syndrome<br>Myasthenia gravis |

Abbreviations: ALP = alkaline phosphatase; ALT = alanine aminotransferase; AST = aspartate aminotransferase; COVID = coronavirus disease; DVT = deep vein thrombosis; ECG = electrocardiogram; GGT = gamma-glutamyl transferase; HELLP = hemolysis, elevated liver enzymes, and low platelets

#### 10.5. APPENDIX 5: Protocol Amendment History

##### 10.5.1. Amendment 9, 30 Sep 2022

**Main Rationale for the Amendment**

The main purpose of this amendment is to update the statistical analyses of Part H (vaccine mRNA-1273.222) to include a primary immunogenicity objective of a comparison between the mRNA-1273.222 and mRNA-1273 groups of the antibody response against the ancestral SARS-CoV-2 D614G.

The summary of changes table provided below describes the major changes made to Amendment 9 relative to Amendment 8, including the sections modified and corresponding rationales. The synopsis of Amendment 9 has been modified to correspond to changes in the body of the protocol. Minor copy edits and administrative updates were made throughout the protocol to align with new content and/or for accuracy.

| Section # and Name | Description of Change | Brief Rationale |
|---|---|---|
| Title Page, Protocol Approval Page, and Protocol Amendment Summary of Changes | Updated protocol version and date. Added Summary of Changes. | Updated to reflect the new protocol version. Summary of Changes added to be in line with Moderna guidelines for protocol amendments. |
| Synopsis; Section 2.0: Objectives and Endpoints Part H; Section 8.2: Statistical Hypotheses; Section 8.5.3.2.8: Analysis for the Primary Immunogenicity Objective for Part H | Added evaluation of the neutralizing antibody response elicited by mRNA-1273.222 vs. mRNA-1273 against the ancestral SARS-CoV-2 D614G strain as a co-primary immunogenicity endpoint | Because mRNA-1273.222 is a bivalent vaccine with an antigenic component that also targets the ancestral SARS-CoV-2 D614G strain, a comparison of the antibody responses against the ancestral SARS-CoV-2 between the vaccine groups is now included in the primary immunogenicity objectives as a comparison. |
| Synopsis; Section 8.0: Statistical Analysis Plan | Revised the seroresponse rate success criteria against BA.4/5 for Part H using a 5% non-inferiority margin. | This was updated based on the SRR data from another Omicron-containing bivalent vaccine, mRNA-1273.214. |

| Section # and Name | Description of Change | Brief Rationale |
|---|---|---|
| Synopsis;<br>Section 8.5.3.2: Analysis for Primary Immunogenicity | Revised the GMR non-inferiority boundary for all study parts, from ≥ 0.67 to > 0.667. | This is a technical update of the GMR non-inferiority success criterion. |
| Synopsis; Section 8.0: Statistical Analysis Plan | Added evaluation of seroresponse based on secondary definition to Parts F (Cohort 2), G, and H. | This update will enable use of both seroresponse rate definitions in the data analysis. |
| Synopsis, Section 3.1: General Design;<br>Section 4.0: Study Population; Section 8.3: Sample Size Determination | In Part H, approximately 500 (updated from 400) participants will receive a second booster dose of mRNA-1273.222 50 µg to achieve 300 (updated from 240) evaluable subjects. | The sample size increase will further accommodate statistical comparisons. |

##### 10.5.2. Amendment 8, 01 Aug 2022

**Amendment 8, 01 Aug 2022**

This amendment is considered to be substantial based on the criteria set forth in Article 10(a) of Directive 2001/20/EC of the European Parliament and the Council of the European Union.

**Main Rationale for the Amendment**

The main purpose of this amendment is to introduce a new study part (part H) for the Omicron BA.4/5-containing bivalent vaccine candidate mRNA-1273.222.

The summary of changes table provided below describes the major changes made to Amendment 8 relative to Amendment 7, including the sections modified and corresponding rationales. The synopsis of Amendment 8 has been modified to correspond to changes in the body of the protocol. Minor copy edits and administrative updates were made throughout the protocol to align with new content and/or for accuracy.

| Section # and Name | Description of Change | Brief Rationale |
|---|---|---|
| Title Page, Protocol Approval Page, and Protocol Amendment Summary of Changes | Updated protocol version and date. Added Summary of Changes. | Updated to reflect the new protocol version. Summary of Changes added to be in line with Moderna guidelines for protocol amendments. |
| Global | New study part (H) added to the study protocol to evaluate the immunogenicity, safety, and reactogenicity of the mRNA-1273.222 vaccine candidate. | To evaluate the safety, reactogenicity, and immunogenicity of the mRNA-1273.222 candidate. |

| Section # and Name | Description of Change | Brief Rationale |
|---|---|---|
| Global | Participant safety follow up shortened from 1 year to 6 months post study injection in Part A.2. | It was determined that 6 months post study injection provides adequate long-term safety follow up in participants who have already been followed for 1 year prior to the booster dose in Part A.2. |

##### 10.5.3. Amendment 7, 26 Apr 2022

This amendment is considered to be substantial based on the criteria set forth in Article 10(a) of Directive 2001/20/EC of the European Parliament and the Council of the European Union.

**Main Rationale for the Amendment**

The main purpose of this amendment is to introduce a sub-part in Part A of the study to offer the mRNA-1273.214 (50 µg) booster vaccine candidate (prototype/Omicron multivalent vaccine) as a second booster dose to participants who have previously received a first booster dose of mRNA-1273.211 (50 µg).

The summary of changes table provided below describes the major changes made to Amendment 7 relative to Amendment 6, including the sections modified and corresponding rationales. The synopsis of Amendment 7 has been modified to correspond to changes in the body of the protocol. Minor copy edits and administrative updates were made throughout the protocol to align with new content and/or for accuracy.

| Section # and Name | Description of Change | Brief Rationale |
|---|---|---|
| Title Page, Protocol Approval Page, and Protocol Amendment Summary of Changes | Updated protocol version and date. Added Summary of Changes. | Updated to reflect the new protocol version. Summary of Changes added to be in line with Moderna guidelines for protocol amendments. |
| Global | New sub-part to Part A (Part A.2) being added to the study.<br>Part A.2 evaluates the immunogenicity, safety, and reactogenicity of the mRNA-1273.214 vaccine candidate given as a second booster dose (50 µg) in Part A participants who have previously received 2 doses of mRNA-1273 as a primary series and a single dose of mRNA-1273.211 (50 µg) as a booster dose. | The Part A.2 primary objective is to evaluate the safety, reactogenicity and immunogenicity of the mRNA-1273.214 candidate as a second booster dose in participants who have previously received the mRNA-1273.211 50 µg booster candidate as a first booster dose. |
| Synopsis, Section 4.1 Inclusion Criteria | Updated inclusion criterion #6 for Part A.2-specific requirements. | Updated as part of the Part A.2 additions described above. |

| Section # and Name | Description of Change | Brief Rationale |
|---|---|---|
| Synopsis, Section 7.2 Immunogenicity Assessments | Added Part A.2 to the testing for serologic markers for SARS-CoV-2 infection as measured by anti-nucleocapsid antibodies detected by immunoassay. | Updated as part of the Part A.2 additions described above. |
| Appendix 1: Schedule of Events, Table 15 | Updated the visit labels. | This update to the table aligns with protocol amendment 6 administrative memo #2, dated 01 Apr 2022. |
| Appendix 4: Adverse Events of Special Interest Terms | Updated the appendix language. | Updated to reflect current language regarding adverse events of special interest. |

##### 10.5.4. Amendment 6, 17 Mar 2022

This amendment is considered to be substantial based on the criteria set forth in Article 10(a) of Directive 2001/20/EC of the European Parliament and the Council of the European Union.

**Main Rationale for the Amendment**

The main purpose of this amendment is to modify the statistical hypothesis testing by: 1) adding non-inferiority testing against the Omicron variant (B.1.1.529) and the ancestral SARS-CoV-2 for multiple timepoints (Day 29, Day 91) based on geometric mean titer ratio and seroresponse rate (SRR) difference to the co-primary endpoints in Part G (booster vaccine candidate mRNA-1273.214); 2) adding superiority testing against the Omicron variant for multiple timepoints (Day 29, Day 91) in Part G; 3) include the SRR difference in the primary endpoints for study parts F and G.

The summary of changes table provided below describes the major changes made to Amendment 6 relative to Amendment 5, including the sections modified and corresponding rationales. The synopsis of Amendment 6 has been modified to correspond to changes in the body of the protocol. Minor copy edits and administrative updates were made throughout the protocol to align with new content and/or for accuracy.

| Section # and Name | Description of Change | Brief Rationale |
|---|---|---|
| Title Page, Protocol Approval Page, and Protocol Amendment Summary of Changes | Updated protocol version and date. Added Summary of Changes. | Updated to reflect the new protocol version. Summary of Changes added to be in line with Moderna guidelines for protocol amendments. |
| Protocol Synopsis | Added sequential enrollment of dose levels to Part F Cohort 2. | Updated to clarify the timing of enrollment of the 2 dose levels in Part F Cohort 2 relative to one another. |
| Synopsis, Section 2 (Objectives and Endpoints), Section 8.2 (Statistical | Changed the primary objectives and endpoints of Part G to include 2 | Added hypothesis testing at Day 91 to evaluate the |

| Section # and Name | Description of Change | Brief Rationale |
|---|---|---|
| Hypotheses), Section 8.3 (Sample Size Determination), Section 8.5.3.2.5 (Analysis for the Primary Immunogenicity Objective for Part F), Section 8.5.3.2.6 (Analysis for the Primary Immunogenicity Objective for Part G), Section 8.6.1 (Interim Analysis) | timepoints (Day 29, Day 91) for hypothesis testing.<br>Changed the SRR endpoint against Omicron from key secondary to a co-primary endpoint for Parts F and G. | durability of the antibody response.<br>SRR against Omicron will be evaluated as a co-primary endpoint; hypothesis testing for non-inferiority against Omicron will be based on GMT ratio and SRR difference and superiority will be based on GMT ratio. |
| Section 3.1 (General Design) | Deleted the final paragraph of this section. | Removed text for clarity and to match updates to the protocol. |
| Section 5.3.1 (Preparation of Study Vaccine) | Part F and G corrected to reflect the correct volume of injections for mRNA-1273 and mRNA-1273.214 | Updated to correct a typographical error. |
| Section 5.3.4 (Study Vaccine Packaging and Labeling) | Part F corrected to provide the correct volume of mRNA-1273 in the glass vials. | Updated to correct a typographical error. |
| Section 8.5.3.2.6 (Analysis for Primary Immunogenicity Objective for Part G) | Added a new heading for Part G. | The heading was added for clarity to differentiate the analyses in Part F and Part G. |
| Section 8.5.3.4 (Other Analysis of Immunogenicity) | Removed reference to Day 29 immunogenicity as a the timepoint of primary interest in the study. | Updated to reflect the new Objectives and Endpoints. |

##### 10.5.5. Amendment 5, 10 Feb 2022

This amendment is considered to be substantial based on the criteria set forth in Article 10(a) of Directive 2001/20/EC of the European Parliament and the Council of the European Union.

**Main Rationale for the Amendment**

The main purposes of this amendment are:

(i) to introduce mRNA-1273 (50 µg) arm, administered as a second booster dose, and to introduce non-inferiority and superiority hypothesis testing for the mRNA-1273.529 (50 µg) Omicron variant booster vaccine candidate in Part F of the study

(ii) to introduce Part G of the study to evaluate the mRNA-1273.214 (50 µg) multivalent booster vaccine candidate.

The summary of changes table provided here describes the changes made in Amendment 5 relative to Amendment 4, including the sections modified and the corresponding rationale. The synopsis of Amendment 5 has been modified to correspond to changes in the body of the protocol. Minor editorial and grammatical corrections were also made.

| Section # and Name | Description of Change | Brief Rationale |
|---|---|---|
| Title Page, Protocol Approval Page, and Protocol Amendment Summary of Changes | Updated protocol version and date. Added Summary of Changes. | Updated to reflect the new protocol version. Summary of Changes added to be in line with Moderna guidelines for protocol amendments. |
| Global | Part G is being added to the study.<br>Part G evaluates the immunogenicity, safety, and reactogenicity of mRNA-1273.214 vaccine candidate as a single booster dose (50 µg) in adults who have previously received 2 doses of mRNA-1273 as a primary series and a single dose of mRNA-1273 (50 µg) as a booster dose. | The Part G primary objectives support non-inferiority and superiority hypothesis testing for the mRNA-1273.214 booster candidate compared to the mRNA-1273 booster. |
| Global | Part F was updated:<br>The 100 µg dose level of mRNA-1273.529 was removed from Cohort 1 and Cohort 2 of Part F. A 50 µg dose level of mRNA-1273 was added to Cohort 2. The part F objectives and endpoints were also revised. | The Part F primary objectives support non-inferiority and superiority hypothesis testing for the mRNA-1273.529 booster candidate compared to the mRNA-1273 booster. |
| Protocol Synopsis, Section 2 (Objectives and Endpoints), Section 8.5 (Statistical Methods) | Updated the Objectives and Endpoints for Part F of the study. | Updated to reflect the changes in the primary and secondary endpoints for Part F. |
| Protocol Synopsis, Section 3.1 (General Design), Section 3.3 (Justification for the Dose, Control Product, and Choice of Study Population), Section 4.1 (Inclusion Criteria) | Added language allowing subjects to have been given their mRNA-1273 booster as part of the mRNA-1273-P301 (COVE) study or under the EUA. | Added for clarification that individuals who received their booster through Study mRNA-1273-P301 or through EUA are eligible to participate in the study. |
| Protocol Synopsis, Section 3.1 (General Design), Section 3.2 (Scientific Rationale for Study Design), Section 7.2 (Immunogenicity Assessments), Section 10.1 (Appendix 1: Schedule of Events) | Addition of a Day 91 Visit for Part F and Part G. | Added to allow serology draws, immunogenicity draws, and nasopharyngeal swab collection, at Day 91 in Part F and Part G. |

| Section # and Name | Description of Change | Brief Rationale |
|---|---|---|
| Section 1.1 (Study Rationale), Section 1.1.1 (mRNA-1273), Section 1.2.3 (Overall Benefit/Risk Conclusion), | Language added to reference the approval of the mRNA-1273 primary series BLA. | Updated to reflect licensure of the mRNA-1273 primary series. |

##### 10.5.6. Amendment 4, 04 Jan 2022

This amendment is considered to be substantial based on the criteria set forth in Article 10(a) of Directive 2001/20/EC of the European Parliament and the Council of the European Union.

**Main Rationale for the Amendment**

The main purposes of this amendment are:

- To add the Omicron variant-specific vaccine candidate, mRNA-1273.529, (50 µg and 100 µg) as first and second booster doses to the protocol in a new study part (Part F)
- To update protocol based on updates (12 November 2021) made in the statistical analysis plan.

**Summary of Major Changes in Protocol Amendment 4:**

| Section # and Name | Description of Change | Brief Rationale |
|---|---|---|
| Title Page, Protocol Approval Page, and Protocol Amendment Summary of Changes | Updated protocol version and date. Added Protocol Amendment Summary of Changes. | Updated to reflect new version, and date of protocol. Protocol Summary of Changes added to be in line with Moderna guidelines for protocol amendments. |
| Global | Minor copy edits were made. | Made for alignment with new content, readability and/or accuracy. |
| Global | Part F is being added to the study.<br>Part F evaluates the immunogenicity, safety, and reactogenicity of mRNA-1273.529 vaccine candidate as a single booster dose (50 µg, 100 µg) in adults who have previously received 2 doses of mRNA-1273 as a primary series (Cohort 1) or have received 2 doses of mRNA-1273 as a primary series and a single dose of mRNA-1273 (50 µg) as a booster dose (Cohort 2). | The Part F primary objectives will enable a comparison between the Omicron-specific immune response elicited from a mRNA-1273.529 booster dose with the ancestral SARS-CoV-2-specific immune response elicited from the mRNA-1273 primary series immunization (historical cohort). These immunogenicity objectives adhere to the regulatory guidance for variant vaccines. |

| Section # and Name | Description of Change | Brief Rationale |
|---|---|---|
| Protocol Synopsis, Section 1.2.5 (mRNA-1273.529), Section 5.1 (Investigational Products Administered), Section 5.3.1 (Preparation of Study Vaccine), Section 5.3.2 (Study Vaccine Administration), Section 5.3.4 (Study Vaccine Packaging and Labeling), and Section 5.3.5 (Study Vaccine Storage) | Part F language was added to discuss mRNA-1273.529 preparation and packaging/labeling. | Part F language was added to include language related to the mRNA-1273.529 IP. |
| Protocol Synopsis, Section 2 (Objectives and Endpoints), Section 8.2 (Statistical Hypothesis), Section 8.5.3.2 (Analysis for the Primary Immunogenicity Objective) | Removed GMT ratio ≥1 point estimator requirement for 50 µg dose arms in multiple study parts. | The point estimator of GMT ratio ≥1 for the 50-µg booster dose of the P205 vaccine candidates is being removed to align with the upversioned statistical analysis plan (SAP version 2). The SAP was upversioned after the interim analysis of Study P201B demonstrated non-inferior immune responses, after the 50-µg booster dose of mRNA-1273, compared to the mRNA-1273 primary series. |
| Protocol Synopsis, Section 2 (Objectives and Endpoints) | Reference to viral sequencing changed from spike protein sequencing to genomic sequencing. | Updated to reflect current practice of sequencing the entire viral genome rather than just the sequence of the spike protein. |
| Protocol Synopsis, Section 3.1 (General Design) | Added discussion of Part E, a site-specific study part that is not described in the global amendment. | Added explanation to clarify the reason Part F follows Part D in this protocol amendment. |
| Protocol Synopsis, Section 3.1 (General Design), Section 10.4: Appendix 1 (Schedule of Events) | Added optional collection of PBMC samples. | To allow for the collection, by some sites, of PBMC samples at specific visits. |
| Protocol Synopsis, Section 4.1 (Inclusion Criteria) | Inclusion criterion 6 was changed to also allow for participants who have received a primary vaccine series of mRNA-1273 and a 50 µg mRNA-1273 booster dose at least 3 months before enrollment to be included in the study. | Addition to the inclusion criterion to allow for enrollment of participants in Cohort 2 of Part F of the study. |

| Section # and Name | Description of Change | Brief Rationale |
| --- | --- | --- |
| Protocol Synopsis, Section 4.2 (Exclusion Criteria) | Exclusion criterion 2 was updated to reflect the new time frame for exclusion due to history of prior SARS-CoV-2 infection. | Updated to clarify the change to the exclusion criterion. |
| Protocol Synopsis, Section 4.2 (Exclusion Criteria) | Exclusion criterion 9 updated to only exclude due to myocarditis or pericarditis within the last 2 months. | Updated to reflect removal AESI as exclusionary except for myocarditis or pericarditis within the last 2 months. |
| Protocol Synopsis, Section 7.2 (Immunogenicity Assays) | Added reference to viral genome sequencing of PCR positive samples. | To clarify that viral genome sequencing is being carried out in alignment with exploratory endpoints. |
| Protocol Synopsis, Section 8.2 (Statistical Hypotheses), Section 8.5.3.2 (Analysis for the Primary Immunogenicity Objective) | For study parts with both, 50 µg and 100 µg doses, removed testing sequence for any interim and for the final analyses. | Testing sequence was removed to align with the upversioned statistical analysis plan (SAP version 2). The SAP was upversioned after the interim analysis of study P201 demonstrated non-inferior immune responses, after the 50-µg booster dose of mRNA-1273, compared to the mRNA-1273 primary series. |
| Section 1.1 (Study Rationale) | Background information regarding the Omicron (B.1.1.529) variant emergence and genotype added to the study rationale. | To provide background and context for the addition of the mRNA-1273.529 variant vaccine to the study. |
| Section 1.2.7 (Clinical Studies) | Updated language to indicate that the Phase 2a study of mRNA-1273 (NCT04405076) has been completed. | To reflect completion of the study. |
| Section 7.2 (Immunogenicity Assessments) | Deleted the “Draw Tube” and “Site Instructions” columns from Table 8. | This information is already contained in the Laboratory Manual and Collections Flow Chart. |
| Section 8.5.3.2 (Analysis for the Primary Immunogenicity Objective) | Updated language to reflect the seroresponse definitions provided in the statistical analysis plan (SAP). | To clarify the seroresponse definition based on pre-dose 1 of primary series as the primary definition for seroresponse. |

##### 10.5.7. Amendment 3, 15 Sep 2021

This amendment is considered to be substantial based on the criteria set forth in Article 10(a) of Directive 2001/20/EC of the European Parliament and the Council of the European Union.

**Main Rationale for the Amendment:**

The main purpose of this amendment is to add mRNA-1273.617.2 50 µg and mRNA-1273.213 50 µg and 100 µg.

**Summary of Major Changes in Protocol Amendment 3:**

| Section # and Name | Description of Change | Brief Rationale |
|---|---|---|
| Title Page, Protocol Approval Page, and Protocol Amendment Summary of Changes | Updated protocol version and date. Added Protocol Amendment Summary of Changes. | Updated to reflect new version, and date of protocol. Protocol Summary of Changes added to be in line with Moderna guidelines for protocol amendments. |
| Global | Minor copy edits were made. | Made for alignment with new content, readability and/or accuracy. |
| Synopsis | Updated the estimated date of last participant and total number of sites. | Changes made to reflect the addition of new study sites and additional parts to the study. |
| Synopsis, Section 2 (Objectives and Endpoints), Section 3.1 (General Design), Section 3.3 (Justification for Dose, Control Product, and Choice of Study Population), Section 4 (Study Population), Section 5.1 (Investigational Products Administered) Section 5.3.1 (Preparation of Study Vaccine), Section 5.3.2 (Study Vaccine Administration), Section 5.3.5 (Study Vaccine Storage), Section 8.6.1 (Interim Analyses), and Appendix 1, Table 10 | 50 µg mRNA-1273.617.2 is being added to Part C which evaluates the immunogenicity, safety, and reactogenicity of mRNA-1273.617.2 vaccine as a single booster dose (50 µg, 100 µg) in adults who have previously received 2 doses of mRNA-1273 as a primary series. | The Part C primary endpoints will enable a comparison between the immune response elicited from mRNA-1273.617.2 as a booster dose, against the B.1.617.2 variant, with the immune response elicited from mRNA-1273 after primary series immunization (historical cohort) against the parental viral strain. |
| | Part D is being added to Study P205. Part D evaluates the immunogenicity, safety, and reactogenicity of mRNA-1273.213 vaccine as a single booster dose (50 µg, 100 µg) in adults who have previously received 2 doses of mRNA-1273 as a primary series. | The Part D primary endpoints will enable a comparison between the immune response elicited from mRNA-1273.213 as a booster dose, against the B.1.617.2 and B.1.351 variants, with the immune response elicited from mRNA-1273 after primary series immunization (historical cohort) against the parental viral strain. |
| Synopsis and Section 4.1 (Inclusion Criteria) | Criterion 6 was updated to include participants that received 2 doses of mRNA-1273 vaccine under the Emergency Use Authorization (EUA) | Study participants might have received the primary series vaccination with an mRNA vaccine for the prevention of |

| Section # and Name | Description of Change | Brief Rationale |
|---|---|---|
| | with their second dose at least 6 months prior to enrollment in mRNA-1273-P205, and able to provide proof of vaccination status at the time of screening (Day 1). | coronavirus disease 19 (COVID-19) outside of mRNA-1273 studies. |
| Synopsis and Section 4.2 (Exclusion Criteria) | Criterion 2 was updated to exclude participants with a known history of SARS-CoV-2 infection within the last 18 months. | Criterion was updated to allow enrollment of participants outside of those that participated in the mRNA-1273-P301 (COVE) study. |
| Synopsis, Section 1.2.4 (mRNA-1273.213), Section 5.1 (Investigational Products Administered), Section 5.3.1 (Preparation of Study Vaccine), and Section 5.3.4 (Study Vaccine Packaging and Labeling) | Part D (mRNA-1273.213) language was added to discuss mRNA-1273.213, preparation, and packaging/labeling. | The Part D language was added to include the information related to the mRNA-1273.213 IP. |
| Synopsis, Section 8.2 (Statistical Hypotheses), Section 8.3 (Sample Size Determination), Section 8.5.3.2 (Analysis for the Primary Immunogenicity Objective), Section 8.5.3.3 (Analysis for Secondary Immunogenicity Objective), and Section 8.5.5 (Exploratory Analyses) | Part C 50 µg mRNA-1273.617.2 hypotheses were added to reflect the addition of Part C Objectives.<br><br>Part D hypotheses were added to reflect the addition of Part D Objectives. | Study hypotheses were updated to reflect the addition of 50 µg mRNA-1273.617.2 to Part C Objectives.<br><br>Study hypotheses were updated to reflect addition of Part D. |
| Section 1.3.1 (Known Potential Benefits) | Known potential benefits was updated to include that mRNA-1273.213 vaccine may be an effective vaccine against COIVD-19 variants of concern (VOC). | Vaccine mRNA-1273.213 includes variant-matched spike protein sequences, from circulating variants of concern |
| Section 7.4.5 (Adverse Event of Special Interest) | Added documentation for AESI and the definition of subclinical myocarditis. | Added for alignment with approved verbiage across other clinical studies |

##### 10.5.8. Amendment 2, 26 Jul 2021

This amendment is considered to be substantial based on the criteria set forth in Article 10(a) of Directive 2001/20/EC of the European Parliament and the Council of the European Union.

**Main Rationale for the Amendment:**

The main purpose of this amendment is to add mRNA-1273 and mRNA-1273.617.2 study arms of 100 µg and update the primary and secondary objectives and endpoints to reflect regulatory guidance.

**Summary of Major Changes in Protocol Amendment 2:**

| Section # and Name | Description of Change | Brief Rationale |
|---|---|---|
| Title Page, Protocol Approval Page, Declaration of Investigator, and Protocol Amendment Summary of Changes | Updated protocol version and date. Added Protocol Amendment Summary of Changes. | Updated to reflect new version, and date of protocol. Protocol Summary of Changes added to be in line with Moderna guidelines for protocol amendments. |
| Global | Minor copy edits were made. | Made for alignment with new content, readability and/or accuracy. |
| Synopsis | Updated the estimated date of last participant and total number of sites. | Changes made to reflect the addition of new study sites and additional parts to the study. |
| Synopsis, Section 2 (Objectives and Endpoints), Section 3.1 (General Design), Section 3.3 (Justification for Dose, Control Product, and Choice of Study Population), and Section 4 (Study Population) | The study is now 3 open-label parts: A, B, and C. The objectives and endpoints were also broken up to address each of the below.<br><br>Part A evaluates the immunogenicity, safety, and reactogenicity of 2 dose levels (50 µg and 100 µg) of mRNA-1273.211 vaccine administered as a single booster dose to adults who have previously received 2 doses of mRNA-1273 as a primary series.<br><br>Part B evaluates the immunogenicity, safety, and reactogenicity of mRNA-1273 vaccine administered as a single booster dose (100 µg) to adults who have previously received 2 doses of mRNA-1273 as a primary series.<br><br>Part C evaluates the immunogenicity, safety, and reactogenicity of mRNA-1273.617.2 vaccine as a single booster dose (100 µg) to adults who have previously received 2 doses of mRNA-1273 as a primary series. | The study is now a 3-part study. Part A is the original study, including 2 dose levels of mRNA-1273.211 (50 and 100 µg). Part B will include a new study arm of mRNA-1273 100 µg. Part C will include a mRNA-1273.617.2 100 µg study arm.<br><br>The part B primary endpoints will enable a comparison between the immune response elicited from mRNA-1273 as a booster dose with the immune response elicited from mRNA-1273 after primary series immunization (historical cohort) using the prototype strain as basis for comparison.<br><br>The part C primary endpoints will enable a comparison between the immune response elicited from mRNA-1273.617.2 as a booster dose, |

| Section # and Name | Description of Change | Brief Rationale |
|---|---|---|
| | | against the B.1.617.2 variant, with the immune response elicited from mRNA-1273 after primary series immunization (historical cohort) against the prototype viral strain. |
| Synopsis and Section 2 (Objectives and Endpoints) | The Key Secondary objectives and endpoints were upgraded to co-primary endpoints for Part A of Amendment 2. | Key secondary objectives and endpoints were updated to co-primary to reflect considerations discussed with regulatory guidance.<br>The updated co-primary objectives and endpoints will enable a comparison of the immune response to booster dose of mRNA-1273.211 against the B.1.351 variant with the immune response to priming doses of the prototype vaccine (historical control) against prototype strain. |
| Synopsis and Section 2 (Objectives and Endpoints) | Symptomatic and asymptomatic SARS-CoV-2 infection terminology language was updated in the Exploratory Objectives:<br>"An alternative definition of symptomatic COVID-19 that is identical to the one used in mRNA-1273-P301" was updated to Primary case definition per the mRNA-1273-P301 COVE study"<br>The CDC definition was updated to Secondary case definition based on the CDC criteria. | To align the naming convention across studies. |
| Synopsis, Section 5.1 (Investigational Products Administered), and Section 5.3.1 (Preparation of Study Vaccine) | Part A (mRNA-1273.211) language was updated to correct concentration of mRNA-1273.211 from 0.5 mg/mL to 0.2 mg/mL in a 20 mM tris buffer and from 10.7 mM to 4.3 mM sodium acetate at pH 7.5.<br><br>Part B (mRNA-1273) language was added to discuss mRNA-1273, preparation, and packaging/labeling. | Part A language was a correction. The Part B and C language was added to include the information related to the mRNA-1273 and mRNA-1273.617.2 IP. |

| Section # and Name | Description of Change | Brief Rationale |
|---|---|---|
| | Part C (mRNA-1273.617.2) language was added to discuss mRNA-1273.617.2, preparation, and packaging/labeling. | |
| Section 7.1.6 (Assessment of SARS-CoV-2 Infection) | "Part A The blinded phase only" language was removed. | This language applies to all parts and the study is open-label. |
| Synopsis, Section 7.4.5 (Adverse Events of Special Interest), and Section 7.5 (Safety Monitoring) | An adjudication committee will be utilized to review any suspected cases of myocarditis, pericarditis, and myopericarditis. | An independent cardiac event adjudication committee will review any suspect cases of myocarditis, pericarditis, and myopericarditis. |
| Synopsis, Section 8.2 (Statistical Hypotheses), Section 8.3 (Sample Size Determination), Section 8.5.4.2 (Analysis for the Primary Immunogenicity Objective) | Part A hypotheses were updated to from 2 Primary and 2 Key Secondary to 4 Primary hypotheses.<br><br>Part B hypotheses were added to reflect the Part B Objectives.<br><br>Part C hypotheses were added to reflect the addition of Part C Objectives. | Key secondary objectives and endpoints (part A) were updated to co-primary to reflect considerations discussed with regulatory guidance. Study hypotheses were updated accordingly. |
| Synopsis and Section 8.3 (Sample Size Determination) | Part A additional power estimation was performed for 50 ug mRNA-1273.211 (270 vs. 526 in PP Set for Immunogenicity). | Updated plan to include all historical control patients in comparison of 50 ug mRNA-1273.211 vs. historical control. |
| Synopsis and Section 8.6.1 (Interim Analysis) | Section 8.6.1 was renamed from "Primary Analysis" to "Interim Analysis."<br><br>Language was also included to describe when the analysis may occur with the addition of Part B and C. | Section heading and content was updated for clarity. |
| Section 1.3.2 (Risks from Study Participation and Their Mitigation) | Safety language related to myocarditis and pericarditis was added. | Language was updated to reflect very rare reports of myocarditis/pericarditis after vaccination. |
| Section 5.3.5 (Study Vaccine Storage) | Language was added to indicate that mRNA-1273.617.2 must be stored at -60°C to -90°C (-76°F to -130°F), and mRNA-1273 must be stored at -25°C to 15°C. | Storage directions were added for mRNA-1273.617.2 and mRNA-1273 for clarity. |

| Section # and Name | Description of Change | Brief Rationale |
|---|---|---|
| Section 7.4.4 (Medically Attended Adverse Events) | The following language was updated from “All MAAEs must be fully reported on the MAAE page of the eCRF” to “Unsolicited AEs will be captured on the AE page of the eCRF.” | Language was updated to reflect actual process. |
| Section 7.4.5 (Adverse Events of Special Interest) | Myocarditis, pericarditis, and myopericarditis case definitions were added for investigator reference. | The CDC definition of myocarditis and pericarditis (both are Adverse Events of Special Interest) was added to assist investigators with the evaluation of any suspect cases. |
| Section 8.5.5 (Exploratory Analysis) | Added 2 exploratory analyses.<br>Exploratory analyses on immune response to selected virus strains may be performed to compare booster regimens (study arms).<br>Other exploratory analyses to compare immune response of boosters against variants compared to the priming series of mRNA-1273 against the variant may be performed. | To assess product cross-protection against variant. |

##### 10.5.9. Amendment 1, 23 Jun 2021

**Main Rationale for the Amendment:**

The main purpose of this amendment is to add an mRNA-1273.211 study arm of 100 µg and update the primary and secondary objectives and endpoints to reflect regulatory guidance.

**Summary of Major Changes in Protocol Amendment 1:**

| Section # and Name | Description of Change | Brief Rationale |
|---|---|---|
| Title Page, Protocol Approval Page, and Protocol Amendment Summary of Changes | Updated protocol version and date. Added Protocol Amendment Summary of Changes. | Updated to reflect new version, and date of protocol. Protocol Summary of Changes added to be in line with Moderna guidelines for protocol amendments. |
| Global | Minor grammar and formatting corrections were made throughout the document. | Updates were made for clarity and readability. |

| Section # and Name | Description of Change | Brief Rationale |
|---|---|---|
| Synopsis | Estimated date last participant completed was updated to July 2022 from May 2022. | New study completion date is based on last participant in mRNA-1273.211 100 µg arm enrolled in July 2021. |
| Synopsis, Section 1.1 (Study Rationale), Section 2 (Objectives and Endpoints), Section 3.1 (General Design), Section 3.3 (Justification for Dose, Control Product, and Choice of Study Population), Section 4 (Study Population), Section 5.1 (Investigational Products Administered), Section 5.3.1 (Preparation of Study Vaccine), Section 8.2 (Statistical Hypotheses), Section 8.3 (Sample Size Determination), and Section 8.6.1 (Primary Analysis) | An mRNA-1273.211 study arm of 100 µg was added to the study. | An mRNA-1273.211 100 µg arm was added to evaluate the safety and immunogenicity of a higher booster dose. The 100 µg arm will be included in the study scientific hypothesis and objectives to demonstrate non-inferiority of the immune responses elicited by mRNA-1273.211 compared with the immune response elicited by mRNA-1273. |
| Synopsis and Section 2 (Primary Objectives and Endpoints) | The primary and secondary objectives and endpoints were updated. Key secondary objectives and endpoints were added before the secondary endpoint. | Primary and secondary objectives and endpoints were updated to reflect considerations discussed with regulatory guidance.<br>The updated primary endpoints will enable a comparison between the immune responses elicited from mRNA-1273.211 and mRNA-1273 (historical cohort) using the prototype strain as basis for comparison.<br>The key secondary objective and endpoint will enable a comparison of immune response to booster dose of mRNA-1273.211 against the B1.1.351 variant with the immune response to priming doses of the prototype vaccine (historical control) against prototype strain. |

| Section # and Name | Description of Change | Brief Rationale |
|---|---|---|
| Synopsis and Section 2 (Objectives and Endpoints) | Secondary objectives and endpoints were modified to include comparisons of the immune response to a booster dose of mRNA-1273.211 with the response to priming doses of mRNA-1273 (historical control) against other variants of interest and concern (including B.1.1617.2). These objectives and endpoints were moved from exploratory and modified.<br><br>Exploratory Endpoint language was also updated for clarity. | Secondary endpoints were updated to perform a comparison of the immune response to the booster dose of mRNA-1273.211 against other viral variants of interest or concern, compared to the immune response to priming doses of the prototype vaccine (historical control) against the prototype strains. |
| Synopsis and Section 3.1 (Scientific Rationale for Study Design) | Additional language was added to specify when the primary analysis will be completed. | This language was added to clarify timing for primary analysis. |
| Synopsis, Section 3.1 (General Design), Section 4 (Study Population), and Section 8.3 (Sample Size Determination) | Language was added to indicate that the 100 µg study arm is expected to have approximately 584 participants, to achieve 526 evaluable participants. | An mRNA-1273.211 100 µg arm was added to evaluate the safety and immunogenicity of a higher booster dose. The 100 µg arm will be included in the study scientific hypothesis, and the study arm size was determined based on statistical calculations, in order to meet the study objectives. |
| Synopsis, Section 1.1 (Study Rationale), Section 8.2 (Statistical Hypotheses), and Section 8.5.4.2 (Analysis for the Primary Immunogenicity Objective) | The primary hypotheses language was updated. | The primary hypotheses language was updated to reflect the changes in the primary objectives as previously specified. |
| Synopsis, Section 8.2 (Statistical Hypotheses), and Section 8.5.4.3 (Analysis for the Key Secondary Immunogenicity Objective) | The hypotheses and supporting language for the key secondary objective were added.<br>The Section 8.5.4.3 (Analysis for the Key Secondary and Secondary Immunogenicity Objective) was added. | The hypotheses language for the key secondary was updated to reflect the changes in the key secondary objectives as previously specified. |
| Synopsis and Section 8.5.4.4 (Other Analysis of Immunogenicity) | More information about other immunogenicity analyses was added. | Information was added to align with the updated primary, key secondary and secondary endpoints. |

| Section # and Name | Description of Change | Brief Rationale |
|---|---|---|
| Synopsis and Section 8.3 (Sample Size Determination) | Sample size language and rationale was updated for clarity and language pertaining to the 100 µg study arm was added. | mRNA-1273.211 100 µg arm was added to evaluate the safety and immunogenicity of a higher booster dose. The 100 µg arm will be included in the study scientific hypothesis, and the study arm size was determined based on statistical calculations, in order to meet the study objectives. |
| Synopsis, Section 8.4 (Analysis Sets), and Section 8.5.2 (Efficacy Analysis) | The Per-Protocol Set title was updated to include 'for Immunogenicity' and the description was updated.<br><br>The Modified Intent-to-Treat Set, and the Per-Protocol Set for Efficacy were also added.<br><br>Language was added to clarify that Efficacy analyses will be performed using Modified Intent-to-Treat Set and Per-Protocol Set. | The modified intent-to-treat and per-protocol analysis sets were added to align with protocols across the SARS-CoV-2 program. |
| Section 7.1.6 (Assessment for SARS-CoV-2 Infection) and Section 10.1 (Appendix 1: Schedule of Events) - Footnote 4 | Language was added to discuss the use of clinical judgment to determine whether a nasopharyngeal swab collection is warranted due to symptoms of COVID-19 overlap with solicited systemic adverse reactions. | This information was added to further support clinical evaluation of suspected COVID-19 events given some overlap between adverse reaction and symptoms of COVID-19. |
| Section 7.2 (Immunogenicity Assessments) - Table 3 Blood and Nasopharyngeal Swab Sampling | A 1 was added to the row PCR/Sequencing under the UNS/Illness column. | This addition was to clarify that polymerase chain reaction testing would take place at acute illness/unscheduled visits to evaluate for SARS-CoV-2 infection. |
| Section 8.5.4.1 (Sampling of Historical Control: Participants from Study mRNA-1273-P301 Immunogenicity Analysis) | Section 8.5.4.1 (Sampling of Historical Control: Participants from Study mRNA-1273-P301 Immunogenicity Analysis) was added to provide further description of the historical control arm. | Information regarding process for selecting samples from the historical controls of participants who received priming doses of mRNA-1273 in Study mRNA-1273-P301 is included. |

| Section # and Name | Description of Change | Brief Rationale |
|---|---|---|
| Section 8.5.4.2 (Analysis for the Primary Immunogenicity Objective) | Section 8.5.4.2 (Analysis for the Primary Immunogenicity Objective) was added to provide further description of the primary immunogenicity objective and its parameters. | Information was added to align with the updated primary objectives. |
| Section 8.5.4.3 (Analysis for the Key Secondary and Secondary Immunogenicity Objective) | Section 8.5.4.3 (Analysis for the Key Secondary and Secondary Immunogenicity Objective) was added to provide further description of the key secondary and secondary immunogenicity objective and its parameters. | Information was added to align with the updated key secondary and secondary objectives. |
| Section 8.5.4.4 (Other Analysis of Immunogenicity) | Section 8.5.4.4 (Other Analysis of Immunogenicity) was added to provide clarification on other immunogenicity analyses language. | Clarifications were made within the immunogenicity analyses language to align with the modified objectives. |

Signature Page for VV-CLIN-009865 v1.0

| | |
|---|---|
| 2nd Approval | Rituparna Das<br>Clinical<br>27-Mar-2023 14:33:48 GMT+0000 |

Signature Page for VV-CLIN-009865 v1.0

**ModernaTX, Inc.**

**Protocol mRNA-1273-P205**

### A Phase 2/3 Study to Evaluate the Immunogenicity and Safety of mRNA Vaccine Boosters for SARS-CoV-2 Variants

#### Statistical Analysis Plan

**SAP Version 6.0**
**Version Date of SAP: 02 May 2023**

Prepared by:

PPD, part of Thermo Fisher Scientific
929 North Front Street
Wilmington, NC 28401-3331
United States

Confidential

6.3.1.1. Incidence of Adverse Events ... 54
6.3.1.2. TEAEs by System Organ Class and Preferred Term ... 55
*6.3.2. Solicited Adverse Reactions ... 55*
6.3.2.1. Analysis of Solicited Adverse Reactions ... 55
*6.3.3. Pregnancy Tests ... 57*
*6.3.4. Vital Sign Measurements ... 57*
6.4. IMMUNOGENICITY ANALYSIS ... 58
*6.4.1. Immunogenicity Assessments ... 58*
*6.4.2. Selecting Study P301 (COVE) Subjects as Historical Control and P201B Subjects as External Comparator ... 58*
*6.4.3. Primary Analysis of Antibody-Mediated Immunogenicity Endpoints ... 59*
*6.4.4. Secondary Analysis of Antibody-Mediated Immunogenicity Endpoints ... 68*
*6.4.5. Exploratory Analysis of Antibody-Mediated Immunogenicity Endpoints ... 70*
*6.4.6. Sensitivity Analysis ... 72*
*6.4.7. Seroresponse ... 73*
6.5. EFFICACY ANALYSIS ... 73
*6.5.1. Endpoint Definition/Derivation ... 74*
6.5.1.1. Derivation of SARS-CoV-2 Infection ... 74
6.5.1.2. Derivation of Asymptomatic SARS-CoV-2 Infection ... 74
6.5.1.3. Derivation of Symptomatic SARS-CoV-2 Infection (COVID-19) ... 75
*6.5.2. Analysis Method ... 77*
*6.5.3. Sensitivity Analysis ... 77*
*6.5.4. SARS-CoV-2 Exposure and Symptoms ... 77*
6.6. INTERIM ANALYSIS ... 78
6.7. DATA SAFETY MONITORING BOARD ... 78
6.8. FINAL ANALYSIS ... 78

**7. REFERENCES ... 78**

**8. LIST OF APPENDICES ... 80**

8.1. APPENDIX A. STANDARDS FOR SAFETY AND IMMUNOGENICITY VARIABLE DISPLAY IN TFLS 80
8.2. APPENDIX B. ANALYSIS VISIT WINDOWS FOR SAFETY AND IMMUNOGENICITY ANALYSIS 80
8.3. APPENDIX C. IMPUTATION RULES FOR MISSING PRIOR/CONCOMITANT MEDICATIONS AND NON-STUDY VACCINATIONS ... 82
8.4. APPENDIX D. IMPUTATION RULES FOR MISSING AE DATES ... 83
8.5. APPENDIX E. IMMUNOGENICITY ENDPOINTS BY STUDY PART ... 84
8.6. APPENDIX F. SUMMARY OF MAJOR CHANGES IN PREVIOUS VERSIONS OF SAP ... 89

#### List of Abbreviations

| Abbreviation | Definition |
|---|---|
| AE | adverse event |
| AESI | adverse event of special interest |
| ANCOVA | analysis of covariance |
| AR | adverse reaction |
| BMI | body mass index |
| bAb | binding antibody |
| CI | confidence interval |
| CSP | clinical study protocol |
| CSR | clinical study report |
| DHHS | Department of Health and Human Services |
| eCRF | electronic case report form |
| eDiary | electronic diary |
| EUA | Emergency Use Authorization |
| FAS | full analysis set |
| GLSM | geometric least square mean |
| GM | geometric mean |
| GMFR | geometric mean fold rise |
| GMT | geometric mean titer |
| GMR | geometric mean ratio |
| IP | investigational product |
| LLOQ | lower limit of quantification |
| MAAEs | medically-attended adverse events |
| MedDRA | Medical Dictionary for Regulatory Activities |
| MMRM | mixed effect model repeated measure |
| mRNA | messenger ribonucleic acid |
| nAb | neutralizing antibody |
| PP | per-protocol |
| PT | preferred term |
| RT-PCR | reverse transcriptase polymerase chain reaction |
| SAE | serious adverse event |
| SAP | statistical analysis plan |
| SARS-CoV-2 | severe acute respiratory syndrome coronavirus 2 |
| SD | standard deviation |
| SOC | system organ class |
| SoE | schedule of events |
| SRR | seroresponse rate |
| TEAE | treatment-emergent adverse event |
| ULOQ | upper limit of quantification |
| VOC | variants of concern |
| WHO | World Health Organization |
| WHODD | World Health Organization drug dictionary |

#### 1. Introduction

This statistical analysis plan (SAP), version 6.0, is based on the clinical study protocol (CSP), Amendment 10, dated 23-Mar-2023 and the most recently approved electronic case report form (eCRF) Release 23, dated 18-Apr-2023. The main purpose of this SAP amendment is to align the analysis plan for the newly added study part J.

**Table 1: Summary of major changes in SAP Version 6.0**

| Section | Brief Description of Changes | Rationale |
| --- | --- | --- |
| 2.1 (Primary Objectives), 2.3 (Exploratory Objectives) | Added objectives for Part J. | Updated according to protocol amendment 10. |
| 3.1 (Primary Endpoints), 3.3 (Exploratory Endpoints) | Added endpoints for Part J. | Updated according to protocol amendment 10. |
| 4.1 (Overall Study Design), 4.2 (Statistical Hypotheses), 4.3 (Sample Size and Power), 4.4 (Randomization) | Added details for Part J. | Updated according to protocol amendment 10. |
| 6.4.3 (Primary Analysis of Antibody-Mediated Immunogenicity Endpoints) | Added details for Part J. | Updated according to protocol amendment 10. |
| 6.5.1.1 (Derivation of SARS-CoV-2 Infection), 6.5.1.2 (Derivation of Asymptomatic SARS-CoV-2 Infection) | Added clarification on derivation. SARS-CoV-2 negative status is required when positive test result is based on serology test post baseline. | Added clarification. |
| 6.6 Interim Analysis | Added information for Part J. | Updated according to protocol amendment 10. |
| 6.8 Final Analysis | Added information for Part J. | Updated according to protocol amendment 10. |

This SAP provides statistical analysis details/data derivations. It also documents modifications or additions to the analysis plan that are not “principal” in nature and result from information that was not available at the time of protocol finalization. If the methods in this SAP differ from the methods described in the protocol, the SAP will prevail.

Study mRNA-1273-P205 is a phase 2/3, open-label study to evaluate the immunogenicity, safety, and reactogenicity of mRNA-1273.211, mRNA-1273, mRNA-1273.617.2, mRNA-1273.213, mRNA-1273.529, mRNA-1273.214, mRNA-1273.222, mRNA-1273.815, and mRNA-1273.231 vaccines. The study consists of 9 parts: Part A.1 will evaluate 2 dose levels (50 or 100 µg) of mRNA-1273.211, Part B will evaluate a single dose of 100 µg mRNA-1273, Part C will evaluate 2 dose levels (50 or 100 µg) of mRNA-1273.617.2, Part D will evaluate 2 dose levels (50 or 100 µg) of mRNA-1273.213, Part E will evaluate a single dose of 100 µg mRNA-1273.213 at a single clinical study site, Part F Cohort 1 will evaluate 50 µg of mRNA-1273.529 as a single booster, Part F Cohort 2 will evaluate 50 µg of the mRNA-1273.529 vaccine and 50 µg of the mRNA-1273 as a second booster in individuals who have previously received a single booster dose of 50 µg mRNA-1273, Part G will evaluate 50 µg of mRNA-1273.214 as a second booster in individuals who have previously received a single booster dose of 50 µg mRNA-1273, Part A.2 will evaluate 50 µg of mRNA-1273.214 as a second booster in individuals who have previously received the mRNA-1273.211 booster, Part H will evaluate 50 µg of mRNA-1273.222 as a second booster in individuals who have previously received a single booster dose of 50 µg mRNA-1273, and Part J will evaluate 50 µg of mRNA-1273.815 and 50 µg of mRNA-1273.231 as a booster in individuals who previously received a primary series of mRNA vaccine, a first booster dose of a monovalent mRNA vaccine, and a second booster dose of a bivalent mRNA vaccine against SARS-CoV-2.

The PPD Biostatistics and programming team, designee of Moderna Biostatistics and Programming department, will perform the statistical analysis of the immunogenicity, safety, reactogenicity and efficacy data. SAS version 9.4 or higher will be used to generate all statistical outputs (tables, figures, listings, and datasets).

In this document, subject and participant are used interchangeably; injection of IP, injection, and dose are used interchangeably. For historical control arm, Day 57 refers Day 57 post dose 1 of primary series.

### 2. Study Objectives

#### 2.1. Primary Objectives

This study consists of 9 parts: A (1, 2), B, C, D, E, F, G, H and J. The primary objectives for each study part are the following:

**Part A.1**

To be assessed for each booster dose level of mRNA-1273.211 (50 µg mRNA-1273.211 and 100 µg mRNA-1273.211):

- To demonstrate non-inferior immune response of a single booster dose of mRNA-1273.211 compared with 2 priming doses of mRNA-1273 in the pivotal Phase 3 efficacy trial (study mRNA-1273-P301 [COVE]):
    - To demonstrate non-inferiority based on geometric mean titer (GMT) ratio (mRNA-1273.211 vs. mRNA-1273) against the ancestral SARS-CoV-2 with a non-inferiority margin of 1.5.
    - To demonstrate non-inferiority based on the seroresponse rate (SRR) (mRNA-1273.211 - mRNA-1273) against the ancestral SARS-CoV-2 with a non-inferiority margin of 10%.
    - To demonstrate non-inferior immune response based on GMT ratio of mRNA-1273.211 as a single booster dose against the variant B.1.351, compared to mRNA-1273 after 2 priming doses against the ancestral SARS-CoV-2 strain with a non-inferiority margin of 1.5.
    - To demonstrate non-inferior immune response based on the SRR of a single booster dose of mRNA-1273.211 against the variant B.1.351 as compared to 2 priming doses of mRNA-1273 against the ancestral SARS-CoV-2 strain with a non-inferiority margin of 10%.
- To evaluate the safety and reactogenicity of mRNA-1273.211.

**Part A.2**

- To evaluate the immunogenicity of mRNA-1273.214 (50 µg) as a second booster dose in participants who previously received a first booster dose of mRNA-1273.211 (50 µg).
- To assess the safety and reactogenicity of the mRNA-1273.214 (50 µg) given as a second booster dose in participants who previously received a first booster dose of mRNA-1273.211 (50 µg).

**Part B**

To be assessed for a single booster dose of 100 µg mRNA-1273:

- To demonstrate non-inferior immune response of a single booster dose of 100 µg mRNA-1273 compared with 2 priming doses of mRNA-1273 in the pivotal Phase 3 efficacy trial (study mRNA-1273-P301 [COVE]):
    - To demonstrate non-inferiority based on GMT ratio (100 µg mRNA-1273 single booster dose vs. 100 µg primary series mRNA-1273) against the ancestral SARS-CoV-2 with a non-inferiority margin of 1.5.
    - To demonstrate non-inferiority based on the SRR (100 µg mRNA-1273 single booster dose vs. 100 µg primary series mRNA-1273) against the ancestral SARS-CoV-2 with a non-inferiority margin of 10%.
- To evaluate the safety and reactogenicity of mRNA-1273.

**Part C**

To be assessed for each dose level of mRNA-1273.617.2 (50 µg mRNA-1273.617.2 and 100 µg mRNA-1273.617.2):

- To demonstrate non-inferior immune response of a single booster dose of mRNA-1273.617.2 compared with 2 priming doses of mRNA-1273 in the pivotal Phase 3 efficacy trial (study mRNA-1273-P301 [COVE]):
    - To demonstrate non-inferiority based on GMT ratio (mRNA-1273.617.2 as a single booster dose against the variant B.1.617.2 vs. 100 µg primary series mRNA-1273) against the ancestral SARS-CoV-2 with a non-inferiority margin of 1.5.
    - To demonstrate non-inferiority based on the SRR of a single booster dose of mRNA-1273.617.2 against the variant B.1.617.2 vs. 100 µg primary series mRNA-1273 against the ancestral SARS-CoV-2 with a non-inferiority margin of 10%.
- To evaluate the safety and reactogenicity of mRNA-1273.617.2.

**Part D**

To be assessed for each dose level of mRNA-1273.213 (50 µg mRNA-1273.213 and 100 µg mRNA-1273.213):

- To demonstrate non-inferior immune response of a single booster dose of mRNA-1273.213 compared with 2 priming doses of mRNA-1273 in the pivotal Phase 3 efficacy trial (study mRNA-1273-P301 [COVE]):
    - To demonstrate non-inferiority based on GMT ratio of mRNA-1273.213 as a single booster dose against the variant B.1.617.2 vs. 100 µg primary series mRNA-1273 against the ancestral SARS-CoV-2 with a non-inferiority margin of 1.5.
    - To demonstrate non-inferiority based on the SRR of a single booster dose of mRNA-1273.213 against the variant B.1.617.2 vs. 100 µg primary series mRNA-1273 against the ancestral SARS-CoV-2 with a non-inferiority margin of 10%.
    - To demonstrate non-inferiority based on GMT ratio of mRNA-1273.213 as a single booster dose against the variant B.1.351 vs. 100 µg primary series mRNA-1273 against the ancestral SARS-CoV-2 with a non-inferiority margin of 1.5.
    - To demonstrate non-inferiority based on the SRR of a single booster dose of mRNA-1273.213 against the variant B.1.351 vs. 100 µg primary series mRNA-1273 against the ancestral SARS-CoV-2 with a non-inferiority margin of 10%.
- To evaluate the safety and reactogenicity of mRNA-1273.213.

**<u>Part E</u>**

To be assessed for a single booster dose of 100 µg mRNA-1273.213 at a single clinical study site:

- To evaluate the safety and reactogenicity of mRNA-1273.213.

**<u>Part F (Cohort 1)</u>**

50 µg mRNA-1273.529 for participants who previously received 100 µg primary series and have not received a mRNA-1273 booster dose previously:

- To demonstrate non-inferiority of the antibody response against the Omicron variant (B.1.1.529) of a first booster dose of mRNA-1273.529 compared to a first booster dose of mRNA-1273 (50 µg) based on GMT ratio and SRR difference.

- To demonstrate superiority of the antibody response against the Omicron variant (B.1.1.529) of a first booster dose of mRNA-1273.529 compared to a first booster dose of mRNA-1273 (50 µg) based on GMT ratio.
- To evaluate the safety and reactogenicity of mRNA-1273.529.

**Part F (Cohort 2)**

Second booster dose of 50 µg mRNA-1273.529 or 50 µg mRNA-1273 for participants who previously received 100 µg primary series and a booster dose of 50 µg mRNA-1273:

- To demonstrate non-inferiority of the antibody response against the Omicron variant (B.1.1.529) of a second booster dose of mRNA-1273.529 compared to a second booster dose of mRNA-1273 (50 µg) based on GMT ratio and SRR difference.
- To demonstrate superiority of the antibody response against the Omicron variant (B.1.1.529) of a second booster dose of mRNA-1273.529 compared to a second booster dose of mRNA-1273 (50 µg) based on GMT ratio.
- To evaluate the safety and reactogenicity of mRNA-1273.529.

**Part G**

Second booster dose of 50 µg mRNA-1273.214 for participants who previously received 100 µg mRNA-1273 primary series and a booster dose of 50 µg mRNA-1273:

- To demonstrate non-inferiority of the antibody response of a second booster dose of mRNA-1273.214 compared to mRNA-1273 (50 µg) when administered as a second booster dose against the Omicron variant (B.1.1.529) based on GMT ratio and SRR difference at Day 29 and Day 91.
- To demonstrate superiority of the antibody response of a second booster dose of mRNA-1273.214 compared to mRNA-1273 (50 µg) when administered as a second booster dose against the Omicron variant (B.1.1.529) based on GMT ratio at Day 29 and Day 91.
- To demonstrate non-inferiority of the antibody response of a second booster dose of mRNA-1273.214 compared to mRNA-1273 (50 µg) when administered as a second booster dose against the ancestral SARS-CoV-2 based on GMT ratio at Day 29 and Day 91.

- To evaluate the safety and reactogenicity of mRNA-1273.214.

**Part H**

Second booster dose of 50 µg mRNA-1273.222 for participants who previously received 100 µg mRNA-1273 primary series and a booster dose of 50 µg mRNA-1273:

- To demonstrate non-inferiority of the antibody response of a second booster dose of mRNA-1273.222 50 µg compared to mRNA-1273 50 µg when administered as a second booster dose against the Omicron BA.4/5 based on GMT ratio and SRR difference at Day 29.
- To demonstrate superiority of the antibody response of a second booster dose of mRNA-1273.222 compared to mRNA-1273 (50 µg) administered as a second booster dose against the Omicron BA.4/5 based on GMT ratio at Day 29.
- To demonstrate non-inferiority of the antibody response of mRNA-1273.222 50 µg compared to mRNA-1273 50 µg when administered as a second booster dose against the ancestral SARS-CoV-2 D614G based on GMT ratio and SRR difference at Day 29.
- To evaluate the safety and reactogenicity of mRNA-1273.222 50 µg.

**Part J**

Booster dose of mRNA-1273.815 (50 µg) or mRNA-1273.231 (50 µg) for participants who previously received a primary series of mRNA vaccine, a first booster dose of a monovalent mRNA vaccine, and a second booster dose of a bivalent mRNA vaccine against SARS-CoV-2:

- To evaluate the immunogenicity of mRNA-1273.815 (50 µg) and mRNA-1273.231 (50 µg) against SARS-CoV-2 Omicron BA.4/BA.5, BQ.1.1, and XBB.1.5 subvariants at Day 15 and Day 29.
- To evaluate the safety and reactogenicity of mRNA-1273.815 (50 µg) and mRNA-1273.231 (50 µg).

#### 2.2. Secondary Objective

The secondary objective will be assessed for each dose level in each study part and is described below:

**Part A.1 and B**

- To compare immune response of mRNA vaccines in each study part as booster against variants compared to the priming series of mRNA-1273 against the ancestral SARS-CoV-2.

**Part A.2**

- To evaluate the immunogenicity of mRNA-1273.214 (50 µg) as a second booster dose in participants who previously received a first booster dose of mRNA-1273.211 (50 µg).

**Part C and D**

- To compare immune response of mRNA vaccines in each study part as booster against the ancestral SARS-CoV-2 and variants compared to the priming series of mRNA-1273 against the ancestral SARS-CoV-2.

**Part E**

- To evaluate the immune response of 100 µg mRNA-1273.213 as booster against the ancestral SARS-CoV-2 and variants at a single clinical study site.

**Part F (Cohort 1)**

- To evaluate the immunogenicity of a mRNA-1273.529 dose compared to a mRNA-1273 administered as a first booster dose at all timepoints post-boost.
- To compare the immune response of mRNA-1273.529 as a first booster dose against the Omicron variant compared to the priming series of mRNA-1273.

**Part F (Cohort 2)**

- To evaluate the immunogenicity of a mRNA-1273.529 booster dose compared to mRNA-1273 booster administered as a second booster dose at all timepoints post-boost.
- To compare the immune response of mRNA-1273.529 as a second booster dose against the Omicron variant compared to the priming series of mRNA-1273.

**Part G**

Key secondary objective:

- To demonstrate non-inferiority based on the SRR against the ancestral SARS-CoV-2 of a second booster dose of mRNA-1273.214 compared to a second booster dose of mRNA-1273 (50 µg) at Day 29 and Day 91.

Secondary objectives:

- To evaluate the immunogenicity of a mRNA-1273.214 booster compared to mRNA-1273 booster administered as a second booster dose at all timepoints post-boost.
- To compare immune response of mRNA-1273.214 as a second dose against the Omicron variant compared to the priming series of mRNA-1273.

**Part H**

- To evaluate the immunogenicity of mRNA-1273.222 50 µg as a second booster dose against the ancestral SARS-CoV-2 (and other variants) compared to a second booster dose of mRNA-1273 50 µg at all timepoints post-booster.

#### 2.3. Exploratory Objectives

The common exploratory objectives shared by all study parts (A.1, A.2, B, C, D, E, F, G, H and J) is the following:

- To assess for symptomatic and asymptomatic SARS-CoV-2 infection.

The common exploratory objectives shared by study parts A.1, A.2, B, C, D, E, F, G, and H is the following:

- To evaluate the genetic and/or phenotypic relationships of isolated SARS-CoV-2 strains to the vaccine sequence.

The exploratory objectives that are specific to each study part are as follows:

**Part A.1**

- To compare the immune response of 50 µg of mRNA-1273.211 with 100 µg of mRNA-1273.211 against the same viral strains.
- To compare immune response of mNRA-1273.211 as booster against variants compared to the priming series of mRNA-1273 against the variants.

**Part B**

- To compare the immune response to ancestral SARS-CoV-2  and variants after a single booster dose of 50 or 100 µg mRNA-1273.211 (Part A) vs. a single booster dose of 100 µg mRNA-1273.
- To compare immune response of mRNA 1273 as booster against variants compared to the priming series of mRNA 1273 against the variants.

**<u>Part C</u>**

- To compare the immune response of 50 µg mRNA-1273.617.2 with 100 µg of mRNA-1273.617.2 against the same viral strains.
- To compare immune response to ancestral SARS-CoV-2 and variants after a single booster dose of mRNA-1273.617.2 vs. a single booster dose of 100 µg mRNA-1273 (Part B).
- To compare immune response to ancestral SARS-CoV-2 and variants after a single booster dose of mRNA-1273.617.2 vs. a single booster dose of 50 or 100 µg mRNA-1273.211 (Part A).
- To compare immune response of mRNA-1273.617.2 as a booster against variants compared to the priming series of mRNA-1273 against the variants.

**<u>Part D</u>**

- To compare the immune response of 50 µg mRNA-1273.213 with 100 µg of mRNA-1273.213 against the same viral strains.
- To compare immune response to ancestral SARS-CoV-2 and variants after a single booster dose of mRNA-1273.213 vs. a single booster dose of 50 or 100 µg mRNA-1273.211 (Part A).
- To compare immune response to ancestral SARS-CoV-2 and variants after a single booster dose of mRNA-1273.213 vs. a single booster dose of 100 µg mRNA-1273 (Part B).
- To compare immune response to ancestral SARS-CoV-2 and variants after a single booster dose of mRNA-1273.213 vs. a single booster dose of 50 or 100 µg mRNA-1273.617.2 (Part C).
- To compare immune response of mRNA-1273.213 as a booster against variants compared to the priming series of mRNA-1273 against the variants.

**Part F (Cohort 1 and 2)**

- To characterize the cellular immune response of mRNA-1273.529 as a booster against the ancestral SARS-CoV-2 and other variants.

**Part G**

- To characterize the cellular immune response of mRNA-1273.214 as a booster against the ancestral SARS-CoV-2 and other variants.

**Part J**

- To evaluate the immunogenicity of mRNA-1273.815 (50 µg) and mRNA-1273.231 (50 µg) against SARS-CoV-2 variants at any study timepoint.

### 3. Study Endpoints

#### 3.1. Primary Endpoints

The primary immunogenicity objectives will be evaluated by the following endpoints for each study part:

**Part A.1**

To be assessed for each booster dose level of mRNA-1273.211 (50 µg mRNA-1273.211 and 100 µg mRNA-1273.211):

- GMT ratio of GMT of mRNA-1273.211 against the ancestral SARS-CoV-2 at Day 29 after the booster dose over GMT of 100 µg mRNA-1273 against the ancestral SARS-CoV-2 at Day 57 (historical control)
- SRR difference between mRNA-1273.211 against the ancestral SARS-CoV-2 at Day 29 after the booster dose and 100 µg mRNA-1273 against the ancestral SARS-CoV-2 at Day 57 (historical control)
- GMT ratio of GMT of mRNA-1273.211 against the variant B.1.351 at Day 29 after the booster dose over GMT of 100 µg mRNA-1273 against the ancestral SARS-CoV-2 strain at Day 57 (historical control)
- SRR difference between mRNA-1273.211 against the variant B.1.351 at Day 29 after the booster dose and 100 µg mRNA-1273 against the ancestral SARS-CoV-2 strain at Day 57 (historical control)

**Part A.2**

- GMT ratio and SRR difference of mRNA-1273.214 (50 µg) as a second booster dose against the ancestral SARS-CoV-2 and against SARS-CoV-2 variants (including Omicron) compared to mRNA-1273.211 (50 µg) dose against the ancestral SARS-CoV-2 and against SARS-CoV-2 variants as the first booster dose at Day 29 and Day 181

**Part B**

- GMT ratio of 100 µg mRNA-1273 against the ancestral SARS-CoV-2 at Day 29 after the booster dose over GMT of 100 µg mRNA-1273 against the ancestral SARS-CoV-2 at Day 57 (historical control)
- SRR difference between 100 µg mRNA-1273 against the ancestral SARS-CoV-2 at Day 29 after the booster dose and 100 µg mRNA-1273 against the ancestral SARS-CoV-2 at Day 57 (historical control)

**Part C**

To be assessed for each dose level of mRNA-1273.617.2 (50 µg mRNA-1273.617.2 and 100 µg mRNA-1273.617.2):

- GMT ratio of mRNA-1273.617.2 against the variant B.1.617.2 at Day 29 after the booster dose over GMT of 100 µg mRNA-1273 against the ancestral SARS-CoV-2 at Day 57 (historical control)
- SRR difference between mRNA-1273.617.2 against the variant B.1.617.2 at Day 29 after the booster dose and 100 µg mRNA-1273 against the ancestral SARS-CoV-2 at Day 57 (historical control)

**Part D**

To be assessed for each booster dose level of mRNA-1273.213 (50 µg mRNA-1273.213 and 100 µg mRNA-1273.213):

- GMT ratio of mRNA-1273.213 against the variant B.1.617.2 at Day 29 after the booster dose over GMT of 100 µg mRNA-1273 against the ancestral SARS-CoV-2 at Day 57 (historical control)

- SRR difference between mRNA-1273.213 against the variant B.1.617.2 at Day 29 after the booster dose and 100 µg mRNA-1273 against the ancestral SARS-CoV-2 at Day 57 (historical control)
- GMT ratio of mRNA-1273.213 against the variant B.1.351 at Day 29 after the booster dose over GMT of 100 µg mRNA-1273 against the ancestral SARS-CoV-2 at Day 57 (historical control)
- SRR difference between mRNA-1273.213 against the variant B.1.351 at Day 29 after the booster dose and 100 µg mRNA-1273 against the ancestral SARS-CoV-2 at Day 57 (historical control)

**Part F (Cohort 1)**

50 µg mRNA-1273.529 for participants who previously received 100 µg primary series and have not received a mRNA-1273 booster dose previously:

- Day 29 post-boost GMT ratio of Omicron-specific GMT of mRNA-1273.529 over the Omicron-specific GMT of mRNA-1273 (historical mRNA-1273 booster dose control)
- Day 29 SRR difference between mRNA-1273.529 against Omicron and mRNA-1273 against Omicron

**Part F (Cohort 2)**

Second booster dose of 50 µg mRNA-1273.529 or 50 µg mRNA-1273 for participants who previously received 100 µg primary series and one booster dose of 50 µg mRNA-1273:

- Day 29 post-boost GMT ratio of Omicron-specific GMT of mRNA-1273.529 over the Omicron-specific GMT of mRNA-1273
- Day 29 SRR difference between mRNA-1273.529 against Omicron and mRNA-1273 against Omicron

**Part G**

Second booster dose of 50 µg mRNA-1273.214 for participants who previously received 100 µg primary series and one booster dose of 50 µg mRNA-1273:

- GMT ratio of Omicron-specific GMT of mRNA-1273.214 over the Omicron-specific GMT of mRNA-1273 (Part F, Cohort 2, 50 µg mRNA-1273) at Day 29 and Day 91
- SRR difference between mRNA-1273.214 against Omicron variant and mRNA-1273 against Omicron variant at Day 29 and Day 91
- GMT ratio of ancestral SARS-CoV-2 GMT of mRNA-1273.214 over the ancestral SARS-CoV-2 GMT of mRNA-1273 (Part F, Cohort 2, 50 µg mRNA-1273) at Day 29 and Day 91

**Part H**

Second booster dose of 50 µg mRNA-1273.222 for participants who previously received 100 µg primary series and a booster dose of 50 µg mRNA-1273:

- GMT ratio of Omicron BA.4/5 GMT of mRNA-1273.222 over the Omicron BA.4/5 GMT of mRNA-1273 (Part F, Cohort 2, 50 µg mRNA-1273) at Day 29
- SRR difference between mRNA-1273.222 against Omicron BA.4/5 and mRNA-1273 against Omicron BA.4/5 at Day 29
- GMT ratio of the ancestral SARS-CoV-2 D614G GMT of mRNA-1273.222 over the ancestral SARS-CoV-2 D614G GMT of mRNA-1273 (Part F, Cohort 2, 50 µg mRNA-1273) at Day 29
- SRR difference between mRNA-1273.222 and mRNA-1273 against the ancestral SARS-CoV-2 D614G at Day 29

**Part J**

Booster dose of mRNA-1273.815 (50 µg) or mRNA-1273.231 (50 µg) for participants who previously received a primary series of mRNA vaccine, a first booster dose of a monovalent mRNA vaccine, and a second booster dose of a bivalent mRNA vaccine against SARS-CoV-2:

- Antibody response of mRNA-1273.815 (50 µg) and mRNA-1273.231(50 µg) against SARS-CoV-2 Omicron BA.4/BA.5, BQ.1.1, and XBB.1.5 subvariants by GMT, geometric mean fold rise (GMFR), and SRR at Day 15 and Day 29

The primary safety objective will be evaluated by the following endpoints for all study parts (A.1, A.2, B, C, D, E, F, G, H and J):

- Solicited local and systemic reactogenicity adverse reactions (ARs) during a 7-day follow-up period after vaccination
- Unsolicited adverse events (AEs) during the 28-day follow-up period after vaccination
- Serious AEs (SAEs), medically attended AEs (MAAEs), AEs leading to withdrawal and AEs of special interest (AESIs) from Day 1 to end of study

#### 3.2. Secondary Endpoints

The secondary objective for each dose level in each study part will be evaluated by the following endpoints:

**Part A.1 and B**

- GMT ratio and SRR difference of each booster arm as a booster dose against variants (as appropriate to each study part) compared to the priming series of mRNA-1273 against the ancestral SARS-CoV-2
    - Variant strains that may be considered: B.1.351, P1, B.1.1.7, B.1.617.2, B.1.1.529 etc.

**Part A.2**

- Antibody response of the mRNA-1273.214 (50 µg) against the ancestral SARS-CoV-2 and against SARS-CoV-2 variants (including Omicron) by GMT and SRR at multiple time points after the mRNA-1273.214 booster dose

**Part C and D**

- GMT ratio and SRR difference of each booster arm as a booster dose against the ancestral SARS-CoV-2 and variants (as appropriate to each study part) compared to the priming series of mRNA-1273 against the ancestral SARS-CoV-2
    - Variant strains that may be considered: B.1.351, P1, B.1.1.7, B.1.617.2, B.1.1.529 etc.

**Part E**

- Immune response of 100 µg mRNA-1273.213 against the ancestral SARS-CoV-2 and variants at all timepoints by GMT, geometric mean fold rise (GMFR), and SRR

    - Variant strains that may be considered: B.1.351, P1, B.1.1.7, B.1.617.2, B.1.1.529 etc.

**Part F (Cohort 1 and 2)**

- GMT ratio of mRNA-1273.529 and mRNA-1273 against the Omicron variant at all timepoints post-boost
- SRR difference between mRNA-1273.529 against the Omicron variant and mRNA-1273 against the Omicron variant
- GMT ratio of mRNA-1273.529 and mRNA-1273 against the ancestral SARS-CoV-2 and other variants at all timepoints post-boost
- SRR difference between mRNA-1273.529 against the ancestral SARS-CoV-2 and other variants and mRNA-1273 against the ancestral SARS-CoV-2 and other variants
- GMT ratio and SRR difference of mRNA-1273.529 as a booster dose (first or second) against the Omicron variant compared to the priming series of mRNA-1273 against the ancestral SARS-CoV-2 (historical control)

**Part G**

Key secondary endpoint:

- SRR difference between mRNA-1273.214 against the Omicron variant and mRNA-1273 against the Omicron variant at all timepoints post-boost

Secondary endpoints:

- GMT ratio of mRNA-1273.214 and mRNA-1273 against the Omicron variant at all timepoints post-boost
- SRR difference between mRNA-1273.214 against the Omicron variant and mRNA-1273 against the Omicron variant at all timepoints post-boost
- GMT ratio of mRNA-1273.214 and mRNA-1273 against the ancestral SARS-CoV-2 and other variants at all timepoints post-boost
- SRR difference between mRNA-1273.214 against the ancestral SARS-CoV-2 and other variants and mRNA-1273 against the ancestral SARS-CoV-2 and other variants at all timepoints post-boost

- GMT ratio and SRR difference of mRNA-1273.214 as a second booster dose against the Omicron variant compared to the priming series of mRNA-1273 against the ancestral SARS-CoV-2 (historical control group)

**Part H**

- GMT ratio of mRNA-1273.222 50 µg and mRNA-1273 50 µg against ancestral SARS-CoV-2 (and other variants) at all timepoints post-booster
- SRR difference between mRNA-1273.222 50 µg and mRNA-1273 50 µg against ancestral SARS-CoV-2 (and other variants) at all timepoints post-booster

##### 3.3. Exploratory Endpoints

The common exploratory endpoints shared by all study parts (A.1, A.2, B, C, D, E, F, G, H and J) are the following:

- Laboratory-confirmed symptomatic or asymptomatic SARS-CoV-2 infection will be defined in participants:
    - Primary case definition per the P301 (COVE) study
    - Secondary case definition based on the CDC criteria: the presence of one of the CDC-listed symptoms (https://www.cdc.gov/coronavirus/2019-ncov/symptoms-testing/symptoms.html) and a positive reverse transcriptase polymerase chain reaction (RT-PCR) test on a respiratory sample
    - Asymptomatic SARS-CoV-2 infection is defined as a positive RT-PCR test on a respiratory sample in the absence of symptoms or a positive serologic test for antinucleocapsid antibody after a negative test at time of enrollment

The common exploratory endpoints shared by study parts A.1, A.2, B, C, D, E, F, G and H) are the following:

- Characterization of the SARS-CoV-2 spike genetic sequence of viral isolates and comparison with the vaccine sequence
- Characterization of immune responses to vaccine breakthrough isolates

Exploratory endpoints for each dose level unique to each study part are the following:

**Part A.1**

- GMT ratio and SRR difference of 50 µg mRNA-1273.211 compared to 100 µg against the same viral strains
- GMT ratio and SRR difference of mRNA-1273.211 as a booster dose against variants compared to the priming series of mRNA-1273 against the variants

**Part B**

- GMT ratio and SRR difference of 50 or 100 µg mRNA-1273.211 (Part A) as a booster dose against ancestral SARS-CoV-2 and variants compared to a booster dose of 100 µg mRNA-1273 against the ancestral SARS-CoV-2 and variants
- GMT ratio and SRR difference of mRNA-1273 as a booster dose against variants compared to the priming series of mRNA-1273 against the variants

**Part C**

- GMT ratio and SRR difference of 50 µg mRNA-1273.617.2 compared to 100 µg of mRNA-1273.617.2 against the same viral strains
- GMT ratio and SRR difference of mRNA-1273.617.2 as a booster dose against ancestral SARS-CoV-2 and variants compared to a booster dose of 100 µg mRNA-1273 (Part B) against the ancestral SARS-CoV-2 and variants
- GMT ratio and SRR difference of mRNA-1273.617.2 as a booster dose against ancestral SARS-CoV-2 and variants compared to a booster dose of 50 or 100 µg mRNA-1273.211 (Part A) against the ancestral SARS-CoV-2 and variants
- GMT ratio and SRR difference of mRNA-1273.617.2 as a booster dose against variants compared to the priming series of mRNA-1273 against the variants

**Part D**

- GMT ratio and SRR difference of 50 µg mRNA-1273.213 compared to 100 µg of mRNA-1273.213 against the same viral strains
- GMT ratio and SRR difference of mRNA-1273.213 as a booster dose against ancestral SARS-CoV-2 and variants compared to a booster dose of 50 or 100 µg mRNA-1273.211 (Part A) against the ancestral SARS-CoV-2 and variants
- GMT ratio and SRR difference of mRNA-1273.213 as a booster dose against ancestral SARS-CoV-2 and variants compared to a booster dose of 100 µg mRNA-1273 (Part B) against the ancestral SARS-CoV-2 and variants

- GMT ratio and SRR difference of mRNA-1273.213 as a booster dose against ancestral SARS-CoV-2 and variants compared to a booster dose of 50 or 100 µg mRNA-1273.617.2 (Part C) against the ancestral SARS-CoV-2 and variants
- GMT ratio and SRR difference of mRNA-1273.213 as a booster dose against variants compared to the priming series of mRNA-1273 against the variants

**Part F (Cohort 1 and 2)**

- T-cell and B-cell response after the mRNA-1273.529 booster

**Part G**

- T-cell and B-cell response after the mRNA-1273.214 booster

**Part J**

- Antibody response of mRNA-1273.815 (50 µg) and mRNA-1273.231(50 µg) against SARS-CoV-2 variants by GMT, GMFR, and SRR at any study timepoint

### 4. Study Design

#### 4.1. Overall Study Design

This is an open-label, Phase 2/3 study to evaluate the immunogenicity, safety, and reactogenicity of mRNA-1273.211 (Part A.1), mRNA-1273 (Part B), mRNA-1273.617.2 (Part C), mRNA-1273.213 (Part D and E), mRNA-1273.529 (Part F), mRNA-123.214 (Part G and Part A.2), mRNA-1273.222 (Part H), mRNA-1273.815 (Part J) and mRNA-1273.231 (Part J) vaccines.

**Part A.1**

Part A.1 will evaluate two dose levels (50 or 100 µg total mRNA content) of the mRNA-1273.211 vaccine administered as a single booster dose to adult participants of the mRNA-1273-P301 (COVE) study who have previously received 2 doses of mRNA-1273 as a primary series. Enrollment will begin with the 50 µg dose arm, followed by the 100 µg dose arm.

**Part A.2**

Part A.2 will evaluate the immunogenicity, safety, and reactogenicity of the mRNA-1273.214 vaccine administered as a second booster dose to adult participants of the mRNA-1273-P205 study who have previously received 2 doses of mRNA-1273 as a primary series and a first booster of (50 µg total mRNA content) of the mRNA-1273.211 in Part A.1 of this study.

**Part B**

Part B will evaluate a single dose of mRNA-1273 100 µg vaccine administered as a single booster dose to adult participants of the mRNA-1273-P301 (COVE) study who have previously received 2 doses of mRNA-1273 as a primary series. Enrollment will begin upon the completion of enrollment of Part A of the study.

**Part C**

Part C will evaluate two dose levels (50 or 100 µg) of the mRNA-1273.617.2 vaccine administered as a single booster dose to adult participants of the mRNA-1273-P301 (COVE) study who have previously received 2 doses of mRNA-1273 as a primary series in Study mRNA-1273-P301 (COVE) or under the EUA. Enrollment of the 100 µg dose arm will begin upon completion of enrollment of Part B of the study and the 50 µg dose arm enrollment will begin after completion of the 100 µg dose level arm in both Part C and D.

**Part D**

Part D will evaluate two dose levels (50 or 100 µg) of the mRNA-1273.213 vaccine administered as a single booster dose to adult participants of the mRNA-1273-P301 (COVE) study who have previously received 2 doses of mRNA-1273 as a primary series in Study mRNA-1273-P301 (COVE) or under the EUA. Enrollment of the 100 µg dose level arm will begin upon completion of enrollment of Part C 100 µg dose level arm of the study followed by the 50 µg dose arm, which may run in parallel with Part C 50 µg dose arm enrollment.

**Part E**

Part E will be enrolled at a single clinical study site and will evaluate a single dose of mRNA-1273.213 100 µg vaccine administered as a single booster dose to adult participants who have previously received 2 doses of any SARS-CoV-2 mRNA authorized vaccine as a primary series. Enrollment of Part E will begin upon completion of enrollment of Part C of the study and will run concurrently with Part D.

**Part F**

Part F will consist of two cohorts.

Cohort 1 will evaluate 50 µg of the mRNA-1273.529 vaccine administered as a single booster dose to adult participants who have previously received 2 doses of mRNA-1273 as a primary series.

Cohort 2 will evaluate 50 µg of the mRNA-1273.529 vaccine  and 50 µg of the mRNA-1273 when administered as a second booster dose to adult participants who have previously received 2 doses of 100 µg mRNA-1273 as a primary series and 1 booster dose of 50 µg mRNA-1273.

Enrollment of the mRNA-1273.529 Cohort 1 will run in parallel with the mRNA-1273.529 in Cohort 2. Enrollment of the 50 µg mRNA-1273 arm in Cohort 2 will begin upon completion of enrollment of the mRNA-1273.529 Cohort 2 arm and may run in parallel with the enrollment of the mRNA-1273.529 Cohort 1 arm.

**Part G**

Part G will evaluate 50 µg of the mRNA-1273.214 vaccine administered as a second booster dose to adults who have previously received 2 doses of 100 µg mRNA-1273 as a primary series and a single booster dose of 50 µg mRNA-1273. Enrollment of the mRNA-1273.214 50 µg second boost arm will begin upon completion of enrollment of the mRNA-1273 50 ug arm in Cohort 2 of Part F. Enrollment of the mRNA-1273.214 50µg second boost arm may run in parallel with the enrollment of the mRNA-1273.529 Cohort 1 arm of Part F.

**Part H**

Part H will evaluate 50 µg of the mRNA-1273.222 vaccine administered as a second booster dose to adults who have previously received 2 doses of 100 µg mRNA-1273 as a primary series and a single booster dose of 50 µg mRNA-1273.

**Part J**

Part J will evaluate mRNA-1273.815 (50 µg) and mRNA-1273.231 (50 µg) administered as a booster to adults who previously received a primary series of mRNA vaccine, a first booster dose of a monovalent mRNA vaccine, and a second booster dose of a bivalent mRNA vaccine against SARS-CoV-2. Participants will be randomized in a 1:1 ratio to receive either mRNA-1273.815 (50 µg) or mRNA-1273.231 (50 µg).

Overall, study parts A.1, B, C, and D will assess whether a single booster dose of mRNA vaccines in each study part boosts antibody responses to the ancestral SARS-CoV-2 and variants similar to antibody responses to the ancestral SARS-CoV-2 elicited by 2 doses of mRNA-1273 (100 µg) against the ancestral SARS-CoV-2, using a historical control arm from the mRNA-1273-P301 (COVE). Part E will assess a single booster dose in participants who previously received any SARS-CoV-2 mRNA authorized vaccine. Study Part F Cohort 1 will assess whether a single booster dose of the mRNA-1273.529 as the first booster dose elicits a similar or superior antibody response to the B.1.1.529 compared to a single booster dose of mRNA-1273, using an external historical comparator. Study Part F Cohort 2 will assess whether a single booster dose of the mRNA-1273.529 as a second booster dose elicits a similar or superior antibody response to the B.1.1.529 compared to a single booster dose of mRNA-1273 as a second booster dose. Study part G will assess whether a single booster dose of the mRNA-1273.214 as a second booster dose elicits a similar or superior antibody response to the B.1.1.529 compared to a single booster dose of mRNA-1273 as a second booster dose (Part F, Cohort 2, 50 µg mRNA-1273). Study Part A.2 will evaluate mRNA-1273.214 administered as a second booster dose to participants who rolled over from Part A.1. Study Part H will assess whether a single booster dose of the mRNA-1273.222 50 µg as a second booster dose elicits a superior antibody response against Omicron BA.4/5 compared to a single booster dose of mRNA-1273 as a second booster dose (Part F, Cohort 2, 50 µg mRNA-1273). Part J will evaluate whether mRNA-1273.815 (50 µg) or mRNA-1273.231 (50 µg) administered as a booster will elicit antibody response against Omicron subvariants BA.4/BA.5, BQ.1.1, and XBB.1.5 in adults who previously received a primary series of mRNA vaccine, a first booster dose of a monovalent mRNA vaccine, and a second booster dose of a bivalent mRNA vaccine against SARS-CoV-2.

**Table 2: Study Arm**

| Study Part | Study Arm | Dose[1] | N |
|---|---|---|---|
| Part A.1 | mRNA-1273.211 | 50 µg | ~300 |
| | mRNA-1273.211 | 100 µg | ~584 |
| Part A.2[2] | mRNA-1273.214 | 50 µg | ~300 |
| Part B | mRNA-1273 | 100 µg | ~300 |
| Part C | mRNA-1273.617.2 | 50 µg | ~584 |
| | mRNA-1273.617.2 | 100 µg | ~584 |
| Part D | mRNA-1273.213 | 50 µg | ~584 |
| | mRNA-1273.213 | 100 µg | ~584 |
| Part E | mRNA-1273.213 | 100 µg | ~50-100 |
| Part F (Cohort 1) | mRNA-1273.529 | 50 µg | ~375 |
| Part F (Cohort 2) | mRNA-1273.529 | 50 µg | ~375 |
| | mRNA-1273 | 50 µg | ~375 |
| Part G | mRNA-1273.214 | 50 µg | ~375 |
| Part H | mRNA-1273.222 | 50 µg | ~500 |
| Part J (3rd booster)[3, 4] | mRNA-1273.815 | 50 µg | ~50 |
| | mRNA-1273.231 | 50 µg | ~50 |

[1] Dose is total mRNA.
[2] Participants rolled over from Part A.1 to Part A.2.
[3] Participants may be rolled over from Part H.
[4] Participants will be randomized in a 1:1 ratio to receive either mRNA-1273.815 or mRNA-1273.231.

#### 4.2. Statistical Hypotheses

##### <u>Part A.1</u>

Each dose of mRNA-1273.211 (50 µg, 100 µg) will be assessed with respect to mRNA-1273 primary series historical control.

For the primary objective on immune response, there are 4 null hypotheses to be tested for each arm.

- $H_0^1$: 50 or 100 µg mRNA-1273.211, as a single booster dose, is inferior to the primary series (100 µg) of mRNA-1273 based on GMT ratio of immune response against the ancestral SARS-CoV-2 with a non-inferiority margin of 1.5.

- $H_0^2$: 50 or 100 µg mRNA-1273.211, as a single booster dose, is inferior to the primary series of mRNA-1273 based on difference in SRR against the ancestral SARS-CoV-2 with a non-inferiority margin of 10%.
- $H_0^3$: 50 or 100 µg mRNA-1273.211 against the variant B.1.351 is inferior to the primary series of (100 µg) mRNA-1273 against the ancestral SARS-CoV-2 based on the GMT ratio of mRNA-1273.211 against the variant B.1.351 at Day 29 compared to mRNA-1273 against the ancestral SARS-CoV-2 at Day 57 with a non-inferiority margin of 1.5.
- $H_0^4$: 50 or 100 µg mRNA-1273.211 against the variant B.1.351 is inferior to the primary series of (100 µg) mRNA-1273 against the ancestral SARS-CoV-2 based on the difference in SRR (SRR of mRNA-1273.211 against the variant B.1.351 at Day 29 – SRR of mRNA-1273 against the ancestral SARS-CoV-2 at Day 57) with a non-inferiority margin of 10%.

**Part A.2**

No statistical hypothesis testing will be performed for Part A.2.

**Part B**

For the primary objective on immune response for a single booster dose of 100 µg mRNA-1273, there are 2 null hypotheses to be tested, and Part B would be considered to meet its primary objective if both null hypotheses are rejected:

- $H_0^1$: 100 µg mRNA-1273, as a single booster dose, is inferior to the primary series of (100 µg) mRNA-1273 based on GMT ratio against the ancestral SARS-CoV-2 with a non-inferiority margin of 1.5.
- $H_0^2$: 100 µg mRNA-1273, as a single booster dose, is inferior to the primary series of mRNA-1273 based on difference in SRR against the ancestral SARS-CoV-2 with a non-inferiority margin of 10%.

**Part C**

Each dose of mRNA-1273.617.2 (50 µg, 100 µg) will be assessed with respect to mRNA-1273 primary series historical control.

For the primary objective on immune response, there are 2 null hypotheses to be tested for each arm:

- $H_0^1$: 50 µg or 100 µg mRNA-1273.617.2, as a single booster dose, against the variant strain B.1.617.2 is inferior to the primary series of (100 µg) mRNA-1273 against the ancestral SARS-CoV-2 based on the GMT ratio of mRNA-1273.617.2 against the variant B.1.617.2 at Day 29 compared to mRNA-1273 against the ancestral SARS-CoV-2 at Day 57 with a non-inferiority margin of 1.5.
- $H_0^2$: 50 µg or 100 µg mRNA-1273.617.2, as a single booster dose, against the variant B.1.617.2 is inferior to the primary series of (100 µg) mRNA-1273 against the ancestral SARS-CoV-2 based on the difference in SRR (SRR of mRNA-1273.617.2 against the variant B.1.617.2 at Day 29 – SRR of mRNA-1273 against the ancestral SARS-CoV-2 at Day 57) with a non-inferiority margin of 10%.

**Part D**

Each dose of mRNA-1273.213 (50 µg, 100 µg) will be assessed with respect to mRNA-1273 primary series historical control.

For the primary objective on immune response, there are 4 null hypotheses to be tested for each arm:

- $H_0^1$: 50 µg or 100 µg mRNA-1273.213, as a single booster dose, against the variant B.1.617.2 is inferior to the primary series of (100 µg) mRNA-1273 against the ancestral SARS-CoV-2 based on the GMT ratio of mRNA-1273.213 against the variant B.1.617.2 at Day 29 compared to mRNA-1273 against the ancestral SARS-CoV-2 at Day 57 with a non-inferiority margin of 1.5.
- $H_0^2$: 50 µg or 100 µg mRNA-1273.213, as a single booster dose, against the variant B.1.617.2 is inferior to the primary series of (100 µg) mRNA-1273 against the ancestral SARS-CoV-2 based on the difference in SRR (SRR of mRNA-1273.213 against the variant B.1.617.2 at Day 29 – SRR of mRNA-1273 against the ancestral SARS-CoV-2 at Day 57) with a non-inferiority margin of 10%.
- $H_0^3$: 50 µg or 100 µg mRNA-1273.213, as a single booster dose, against the variant B.1.351 is inferior to the primary series of (100 µg) mRNA-1273 against the ancestral SARS-CoV-2 based on the GMT ratio of mRNA-1273.213 against the variant B.1.351 at Day 29 compared to mRNA-1273 against the ancestral SARS-CoV-2 at Day 57 with a non-inferiority margin of 1.5.

- $H_0^4$: 50 µg or 100 µg mRNA-1273.213, as a single booster dose, against the variant B.1.351 is inferior to the primary series of (100 µg) mRNA-1273 against the ancestral SARS-CoV-2 based on the difference in SRR (SRR of mRNA-1273.213 against the variant B.1.351 at Day 29 – SRR of mRNA-1273 against the ancestral SARS-CoV-2 at Day 57) with a non-inferiority margin of 10%.

**Part E**

There will be no formal hypothesis testing for Part E. All analyses will be descriptive.

**Part F (Cohort 1)**

50 µg mRNA-1273.529 as a first booster dose will be assessed with respect to mRNA-1273 as a first booster dose using mRNA-1273-P201 part B 50 µg booster after 100 µg primary series as an external comparator.

For the primary objective on immune response, hypotheses are:

- 50 µg mRNA-1273.529, as a single booster dose, against the variant B.1.1.529 is either non-inferior or superior to the booster dose of (50 µg) mRNA-1273 against B.1.1.529 based on the GMT ratio of mRNA-1273.529 against the variant B.1.1.529 at Day 29 compared to mRNA-1273 against the variant B.1.1.529 at Day 29 with a non-inferiority margin of 1.5.
- 50 µg mRNA-1273.529, as a single booster dose, against the variant B.1.1.529 is non-inferior to the booster dose of (50 µg) mRNA-1273 against B.1.1.529 based on the difference in SRR at Day 29 with a non-inferiority margin of 10%.

The primary immunogenicity objective is considered met if non-inferiority against B.1.1.529 (based on GMR and SRR difference) is demonstrated.

**Part F (Cohort 2)**

50 µg mRNA-1273.529 as a second booster dose will be compared to 50 µg mRNA-1273 as a second booster dose.

For the primary objective on immune response, hypotheses are:

- 50 µg mRNA-1273.529, as a second booster dose, against the variant B.1.1.529 is either non-inferior or superior to the second booster dose of (50 µg) mRNA-1273 against B.1.1.529 based on the GMT ratio of mRNA-1273.529 against the variant B.1.1.529 at Day 29 compared to mRNA-1273 against the variant B.1.1.529 at Day 29 with a non-inferiority margin of 1.5.

- 50 µg mRNA-1273.529, as a second booster dose, against the variant B.1.1.529 is non-inferior to the booster dose of (50 µg) mRNA-1273 against B.1.1.529 based on the difference in SRR with a non-inferiority margin of 10%.

The primary immunogenicity objective is considered met if non-inferiority against B.1.1.529 (based on GMR and SRR difference) is demonstrated.

**Part G**

50 µg mRNA-1273.214 as a second booster dose will be compared to 50 µg mRNA-1273 as the second booster dose (active control arm in Part F, Cohort 2, booster-to-booster comparison for the 2$^{nd}$ booster).

The primary objective on immune response will be tested with 0.025 two-sided alpha each initially allocated to Day 29 and Day 91 respectively. For the primary objective of immune response, an alpha of 0.05 (two-sided) will be allocated to the two time points. Day 29 and Day 91 will each have an alpha of 0.025 (two-sided) for hypotheses testing. If all objectives are met at Day 29, hypotheses at Day 91 can be tested at an alpha of 0.05 (two-sided).

Figure 1 below demonstrates the testing strategy at Day 29 and Day 91.

**Figure 1: Statistical Hypotheses Testing Strategy for Part G**

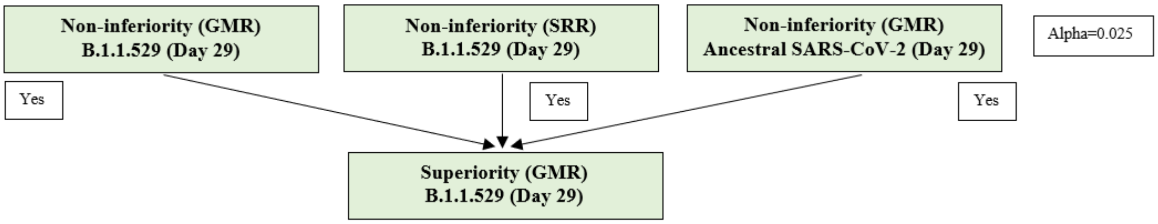

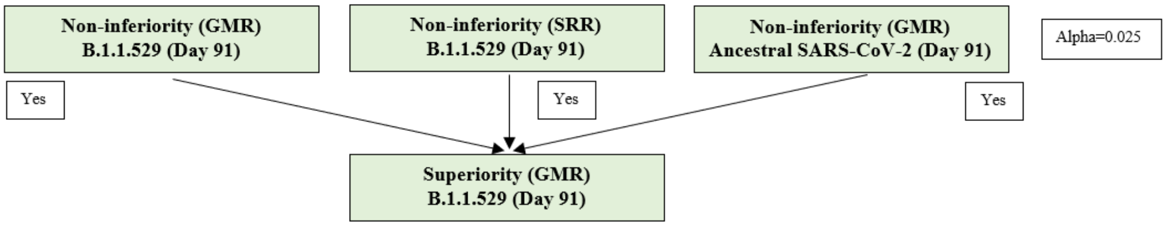

Note: If all objectives are met at Day 29, hypotheses at Day 91 can be tested at an alpha of 0.05 (two-sided).

Hypotheses at Day 29:

- 50 µg mRNA-1273.214, as a second booster dose, against the variant B.1.1.529 is non-inferior to the second booster dose of (50 µg) mRNA-1273 against B.1.1.529 based on the GMT ratio of mRNA-1273.214 against the variant B.1.1.529 at Day 29 compared to mRNA-1273 against the variant B.1.1.529 at Day 29 with a non-inferiority margin of 1.5.
- 50 µg mRNA-1273.214, as a second booster dose, against the variant B.1.1.529 is non-inferior to the second booster dose of (50 µg) mRNA-1273 against B.1.1.529 based on the difference in SRR at Day 29 with a non-inferiority margin of 10%.
- 50 µg mRNA-1273.214, as a second booster dose, against the ancestral SARS-CoV-2 is non-inferior to the second booster dose of (50 µg) mRNA-1273 against the ancestral SARS-CoV-2 based on the GMT ratio of mRNA-1273.214 against the ancestral SARS-CoV-2 at Day 29 compared to mRNA-1273 against the ancestral SARS-CoV-2 at Day 29 with a non-inferiority margin of 1.5.
- 50 µg mRNA-1273.214, as a second booster dose, against the variant B.1.1.529 is superior to the second booster dose of (50 µg) mRNA-1273 against B.1.1.529 based on the GMT ratio of mRNA-1273.214 against the variant B.1.1.529 at Day 29 compared to mRNA-1273 against the variant B.1.1.529 at Day 29. Please note

this hypothesis will only be tested when the above 3 hypotheses have been demonstrated.

Hypotheses at Day 91:

- 50 µg mRNA-1273.214, as a second booster dose, against the variant B.1.1.529 is non-inferior to the second booster dose of (50 µg) mRNA-1273 against B.1.1.529 based on the GMT ratio of mRNA-1273.214 against the variant B.1.1.214 at Day 91 compared to mRNA-1273 against the variant B.1.1.529 at Day 91 with a non-inferiority margin of 1.5.
- 50 µg mRNA-1273.214, as a second booster dose, against the variant B.1.1.529 is non-inferior to the second booster dose of (50 µg) mRNA-1273 against B.1.1.529 based on the difference in SRR at Day 91 with a non-inferiority margin of 10%.
- 50 µg mRNA-1273.214, as a second booster dose, against the ancestral SARS-CoV-2 is non-inferior to the second booster dose of (50 µg) mRNA-1273 against the ancestral SARS-CoV-2 based on the GMT ratio of mRNA-1273.214 against the ancestral SARS-CoV-2 at Day 91 compared to mRNA-1273 against the ancestral SARS-CoV-2 at Day 91 with a non-inferiority margin of 1.5.
- 50 µg mRNA-1273.214, as a second booster dose, against the variant B.1.1.529 is superior to the second booster dose of (50 µg) mRNA-1273 against B.1.1.529 based on the GMT ratio of mRNA-1273.214 against the variant B.1.1.529 at Day 91 compared to mRNA-1273 against the variant B.1.1.529 at Day 91. Please note this hypothesis will only be tested when the above 3 hypotheses have been demonstrated.

The primary immunogenicity objective is considered met if non-inferiority against B.1.1.529 based on GMR SRR difference and non-inferiority against the ancestral SARS-CoV-2 based on GMR are demonstrated either at Day 29 or Day 91.

For the key secondary objective, hypotheses are (Day 29 and 91 will each have an alpha of 0.025 (two-sided) for hypotheses testing):

- 50 µg mRNA-1273.214, as a second booster dose, against the ancestral SARS-CoV-2 is non-inferior to the booster dose of (50 µg) mRNA-1273 against the ancestral SARS-CoV-2 based on the difference in SRR at Day 29 with a non-inferiority margin of 10%.

- 50 µg mRNA-1273.214, as a second booster dose, against the ancestral SARS-CoV-2 is non-inferior to the booster dose of (50 µg) mRNA-1273 against the ancestral SARS-CoV-2 based on the difference in SRR at Day 91 with a non-inferiority margin of 10%.

**Part H**

50 µg mRNA-1273.222 given as a second booster dose will be compared to 50 µg mRNA-1273 given as a second booster dose (in Part F, Cohort 2).

For the primary immunogenicity objective, there are 5 hypotheses to be tested. Figure 2 below demonstrates the testing strategy.

**Figure 2: Statistical Hypotheses Testing Strategy for Part H**

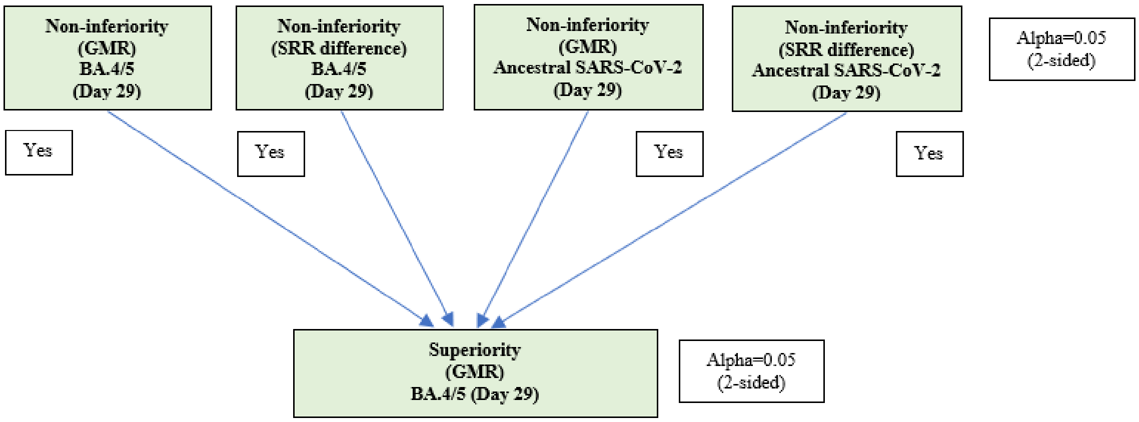

For the primary immunogenicity objective, the 5 hypotheses to be tested are the following:

- 50 µg mRNA-1273.222, as a second booster dose, against Omicron BA.4/5 is non-inferior to the second booster dose of (50 µg) mRNA-1273 against Omicron BA.4/5 based on the GMT ratio of mRNA-1273.222 against Omicron BA.4/5 at Day 29 compared to mRNA-1273 against Omicron BA.4/5 at Day 29 with a non-inferiority margin of 1.5.
- 50 µg mRNA-1273.222, as a second booster dose, against Omicron BA.4/5 is non-inferior to the second booster dose of (50 µg) mRNA-1273 against Omicron BA.4/5 based on the difference in SRR at Day 29 with a non-inferiority margin, if

- the lower bound of the 95% CI of the SRR difference (50 µg mRNA-1273.222 against Omicron BA.4/5 at Day 29 - 50 µg mRNA-1273 against Omicron BA.4/5 at Day 29) is >-5%, then the non-inferiority of mRNA-1273.222 against Omicron BA. 4/5 compared to that of mRNA-1273 is demonstrated based on a non-inferiority margin of 5%.
- the lower bound of the 95% CI of the SRR difference (50 µg mRNA-1273.222 against Omicron BA.4/5 at Day 29 - 50 µg mRNA-1273 against Omicron BA.4/5 at Day 29) is >-10% but ≤ -5%, then non-inferiority of mRNA-1273.222 against Omicron BA. 4/5 compared to that of mRNA-1273 is demonstrated based on a non-inferiority margin of 10%.

- 50 µg mRNA-1273.222, as a second booster dose, against the ancestral SARS-CoV-2 D614G is non-inferior to the second booster dose of (50 µg) mRNA-1273 against the ancestral SARS-CoV-2 D614G based on the GMT ratio of mRNA-1273.222 against the ancestral SARS-CoV-2 D614G at Day 29 compared to mRNA-1273 against the ancestral SARS-CoV-2 D614G at Day 29 with a non-inferiority margin of 1.5.
- 50 µg mRNA-1273.222, as a second booster dose, against the ancestral SARS-CoV-2 D614G is non-inferior to the second booster dose of (50 µg) mRNA-1273 against ancestral SARS-CoV-2 D614G based on the difference in SRR at Day 29 with a non-inferiority margin 10%.
- 50 µg mRNA-1273.222, as a second booster dose, against Omicron BA.4/5 is superior to the second booster dose of (50 µg) mRNA-1273 against Omicron BA.4/5 based on the GMT ratio of mRNA-1273.222 against Omicron BA.4/5 at Day 29 compared to mRNA-1273 against Omicron BA.4/5 at Day 29.

**Part J**

No formal hypothesis testing will be performed for Part J and all safety and immunogenicity analyses will be descriptive.

#### 4.3. Sample Size and Power

**Part A.1**

Each dose level of mRNA-1273.211 will be assessed at a 2-sided type I error rate of 5%.

The target enrollment of the 50 µg mRNA-1273.211 arm is approximately 300 participants. Assuming 10% of participants will be excluded from the PP Set for Immunogenicity, with approximately 270 participants in the mRNA-1273.211 50 µg study arm and from the mRNA-1273 (primary series) historical control arm in the PP Set for Immunogenicity, there is approximately 75% power to reject all null hypotheses for the primary objectives. With approximately 270 participants in the mRNA-1273.211 50 µg study arm and 526 participants from the mRNA-1273 primary series historical control arm, there is approximately 90% power to reject all null hypotheses for the primary objectives based on GMR and the difference in SRR against the ancestral SARS-CoV-2 and variant (B.1.351) at 2-sided alpha of 5.0%. The assumptions are: the true GMR (50 µg mRNA-1273.211 vs. 100 µg mRNA-1273) against the ancestral SARS-CoV-2 and the variant (B.1.351) is 1, the standard deviation of the log-transformed titer is 1.5, with a non-inferiority margin of 1.5; the true SRR against ancestral SARS-CoV-2 and the variant (B.1.351) after a single booster dose of mRNA-1273.211 is 90%, SRR against ancestral SARS-CoV-2 after mRNA-1273 primary series is 90%, and non-inferiority margin for SRR difference is 10%.

The target enrollment of 100 µg mRNA-1273.211 arm is approximately 584 participants. Assuming 10% of participants will be excluded from the PP Set for Immunogenicity, with approximately 526 participants in mRNA-1273.211 100 µg and mRNA-1273 primary series historical control arm respectively in the PP Set for Immunogenicity, there is approximately 80% power to reject all null hypotheses for the primary objectives based on GMR and difference in SRR against the ancestral SARS-CoV-2 at 2-sided alpha of 5.0%. The assumptions are: the true GMR against the ancestral SARS-CoV-2 and the variant (B.1.351) is 0.9, the standard deviation of the log-transformed titer is 1.5, non-inferiority margin for GMR is 1.5; the true SRR against ancestral SARS-CoV-2 and the variant (B.1.351) after a single booster dose of mRNA-1273.211 is 90%, and SRR against ancestral SARS-CoV-2 and the variant (B.1.351) after mRNA-1273 primary series is 90%, and non-inferiority margin for SRR difference is 10%.

Approximately 584 participants in the mRNA-1273-P301 (COVE) study will be selected as the mRNA-1273 priming series historical control arm. The sampling plan for the selection of mRNA-1273-P301 (COVE) participants is included in Section 4.6. The same historical comparator arm will be used for the subsequent Part B, C, and D.

With approximately 300 and 584 participants exposed to 50 and 100 µg of mRNA-1273.211, respectively, there is at least 90% probability to observe one participant at each dose level reporting an AE if the true rate of AEs is 1%.

**Part A.2**

We anticipate approximately 300 participants will be enrolled in Part A.2, there is no statistical hypothesis testing in Part A.2.

**Part B**

The target enrollment of the 100 µg mRNA-1273 arm is approximately 300 participants. Assuming 10% of participants will be excluded from the PP Set for Immunogenicity, with approximately 270 participants in the 100 µg mRNA-1273 study arm and approximately 526 from the mRNA-1273 primary series historical control arm in the PP Set for Immunogenicity, there is >90% power to reject both null hypotheses for the primary objective based on GMR and the difference in SRR against the ancestral SARS-CoV-2 (mRNA-1273) at 2-sided alpha of 5.0%. The assumptions are: the true GMR (100 µg mRNA-1273 booster vs. mRNA-1273 primary series historical control) against the ancestral SARS-CoV-2 is 1.5, the standard deviation of the log-transformed titer is 1.5, with a non-inferiority margin of 1.5; the true SRR against ancestral SARS-CoV-2 after a single booster dose of mRNA-1273 is 90%, SRR against ancestral SARS-CoV-2 after mRNA-1273 primary series is 90%, and non-inferiority margin for SRR difference is 10%.

With approximately 300 participants exposed to 100 µg of mRNA-1273, there is at least 90% probability to observe one participant reporting an AE if the true rate of AEs is 1%.

**Part C**

Each dose level of mRNA-1273.617.2 will be assessed at a 2-sided type I error rate of 5%.

The target enrollment is approximately 584 participants for each dose level of mRNA-1273.617.2. Assuming 10% of participants will be excluded from the PP Set for Immunogenicity, with approximately 526 participants in each dose level of mRNA-1273.617.2 and mRNA-1273 primary series historical control arm respectively in the PP Set for Immunogenicity, there is approximately 90% power to reject both null hypotheses for the primary objectives based on GMR and difference in SRR. The assumptions are: the true GMR (mRNA-1273.617.2 booster vs. 100 µg mRNA-1273 primary series historical control) against the variant (B.1.617.2) is 0.9 compared to 100 µg mRNA-1273 against ancestral SARS-CoV-2, the standard deviation of the log-transformed titer is 1.5, non-inferiority margin for GMR is 1.5; the true SRR against variant strain after a single booster dose of mRNA-1273.617.2 is 90%, and SRR against ancestral SARS-CoV-2 and variant (B.1.617.2) after mRNA-1273 primary series is 90%, and non-inferiority margin for SRR difference is 10%. The immune response assumptions are the same for 50 µg and 100 µg mRNA-1273.617.2.

With approximately 584 participants exposed to each dose of mRNA-1273.617.2, there is at least 90% probability in each group to observe one participant reporting an AE if the true rate of AEs is 1%.

**Part D**

Each dose level of mRNA-1273.213 will be assessed at a 2-sided type I error rate of 5%.

The target enrollment is approximately 584 participants for each dose level of mRNA-1273.213. Assuming 10% of participants will be excluded from the PP Set for Immunogenicity, with approximately 526 participants in each dose level of mRNA-1273.213 and mRNA-1273 primary series historical control arm respectively in the PP Set for Immunogenicity, there is approximately 80% power to reject all null hypotheses for the primary objectives based on GMR and difference in SRR. The assumptions are: the true GMR (mRNA-1273.213 vs. 100 µg mRNA-1273 primary series historical control) against the two variant strains (B.1.617.2, B.1.351) is 0.9 compared to 100 µg mRNA-1273 against ancestral SARS-CoV-2, the standard deviation of the log-transformed titer is 1.5, non-inferiority margin for GMR is 1.5; the true SRR against variants (B.1.617.2, B.1.351) after a single booster dose of mRNA-1273.213 is 90%, SRR against ancestral SARS-CoV-2 and variant after mRNA-1273 primary series is 90%, and non-inferiority margin for SRR difference is 10%. The immune response assumptions are the same for 50 µg and 100 µg mRNA-1273.213.

With approximately 584 participants exposed to each dose of mRNA-1273.213, there is at least 90% probability to observe one participant reporting an AE if the true rate of AEs is 1%.

**Part E**

The target enrollment of the mRNA-1273.213 in this part of the study is between 50 and 100 participants, with a subset of enrolled participants who previously received Moderna's mRNA-1273 authorized vaccine as a primary series. The sample size for Part E is not driven by statistical assumptions for hypothesis testing.

**Part F**

mRNA-1273.529 in each cohort will be assessed at a 2-sided type I error rate of 5%.

Cohort 1:

The target enrollment is approximately 375 participants for 50 µg mRNA-1273.529. Assuming 20% of participants will be excluded from the PP Set for Immunogenicity-SARS-CoV2 negative, with approximately 300 participants in 50 µg mRNA-1273.529 and 300 participants in 50 µg mRNA-1273 (external comparator, see Section 4.7) in the PP Set for Immunogenicity-SARS-CoV-2 negative, there is approximately 89% global power for the primary immunogenicity objectives with alpha level of 0.05 (2-sided). The assumptions are: the true GMR (mRNA-1273.529 booster vs. 50 µg mRNA-1273 booster) against the variant (B.1.1.529) is 1.5, the standard deviation of the log-transformed titer is 1.5, and the non-inferiority margin for GMR is 1.5; the true SRR against B.1.1.529 after a single booster dose of 50 µg mRNA-1273.529 is 90% (same assumption for 50 µg mRNA-1273), and non-inferiority margin for SRR difference is 10%.

With approximately 375 participants exposed to 50 µg mRNA-1273.529, there is at least 90% probability in this group to observe 1 participant reporting an AE if the true rate of AEs is 1%.

Cohort 2:

The target enrollment is approximately 750 participants for 50 µg mRNA-1273.529 and 50 µg mRNA-1273 (1:1 ratio). Assuming 20% of participants will be excluded from the PP Set for Immunogenicity – SARS-CoV-2 negative, with approximately 300 participants each in 50 µg mRNA-1273.529 and 50 µg mRNA-1273 in the PP Set for Immunogenicity and SARS-CoV-2 negative, there is approximately 89% global power to demonstrate the primary immunogenicity objectives of alpha level of 0.05 (2-sided). The assumptions are: the true GMR (mRNA-1273.529 as the second booster vs 50 µg mRNA-1273 as the second booster) against the variant (B.1.1.529) is 1.5, the standard deviation of the log-transformed titer is 1.5, and the non-inferiority margin for GMR is 1.5; the true SRR against B.1.1.529 after 50 µg mRNA-1273.529 as a second booster dose is 90% (same assumption for 50 µg 1273), and non-inferiority margin for SRR difference is 10%.

With approximately 375 participants exposed to each group, there is at least 90% probability in each group to observe 1 participant reporting an AE if the true rate of AEs is 1%.

**Part G**

The target enrollment is approximately 375 participants for 50 µg mRNA1273.214. Hypotheses testing will be performed at Day 29 and Day 91, alpha of 0.025 (2-sided) will be allocated equally to each one of the two time points. Assuming 20% of participants will be excluded from the PP Set for Immunogenicity – SARS-CoV-2 negative, with approximately 300 participants in 50 µg mRNA1273.214 and 300 participants in 50 µg mRNA-1273 (Part F, Cohort 2-50 µg mRNA1273) in the PP Set for Immunogenicity and SARS-CoV-2 negative, there is approximately 71% global power to demonstrate the primary immunogenicity objectives with alpha of 0.025 (2-sided) at each time point. The assumptions are: the true GMR (mRNA-1273.214 second booster vs. 50 µg mRNA-1273 second booster) against the variant (B.1.1.529) is 1.5, the true GMR against ancestral SARS-CoV-2 is 1, the standard deviation of the log-transformed titer is 1.5, and the non-inferiority margin for GMR is 1.5, the true SRR against B.1.1.529 after mRNA-1273.214 as a second booster dose is 90% (same assumption for both 50 µg mRNA-1273.214 and 50 µg mRNA-1273), and non-inferiority margin for SRR difference is 10%.

With approximately 375 participants exposed to 50 µg mRNA-1273.214, there is at least 90% probability in this group to observe 1 participant reporting an AE if the true rate of AEs is 1%.

**Part H**

The target enrollment is approximately 500 participants for 50 µg mRNA1273.222. Assuming 40% of participants will be excluded from the PP Set for Immunogenicity – SARS-CoV-2 negative (due to a SARS-CoV-2 infection pre-booster), with approximately 300 participants in 50 µg mRNA1273.222 and 260 participants in 50 µg mRNA-1273 (Part F, Cohort 2-50 µg mRNA1273) in the PP Set for Immunogenicity and SARS-CoV-2 negative, there is approximately 60% power to demonstrate the primary immunogenicity objectives with alpha of 0.05 (2-sided) at Day 29. The assumptions are: the true GMR (mRNA-1273.222 second booster vs. 50 µg mRNA-1273 second booster) against Omicron BA.4/5 is 1.5, GMR (mRNA-1273.222 second booster vs. mRNA-1273 second booster) against ancestral SARS-CoV-2 D614G is 1, the standard deviation of the log-transformed titer is 1.5, and the non-inferiority margin for GMR is 1.5. The true SRR against Omicron BA.4/5 after mRNA-1273.222 as a second booster dose is 95% (same assumption for 50 µg mRNA-1273), and non-inferiority margin for SRR difference against Omicron BA.4/5 is 5%. The true SRR against ancestral SARS-CoV-2 D614G after mRNA-1273.222 as a second booster dose is 95% (same assumption for 50 µg mRNA-1273), and the non-inferiority margin for SRR difference against ancestral SARS-CoV-2 D614G is 10%.

With approximately 500 participants exposed to 50 µg mRNA-1273.222, there is at least 90% probability in this group to observe 1 participant reporting an AE if the true rate of AEs is 1%.

**Part J**

The sample size for Part J is not driven by statistical assumptions for formal hypothesis testing. The number of proposed participants is considered sufficient to provide a descriptive summary of the safety and immunogenicity of each treatment arm.

The target enrollment is approximately 100 participants who will be randomized in a 1:1 ratio to receive either mRNA-1273.815 (50 µg) or mRNA-1273.231 (50 µg).

#### 4.4. Randomization

Randomization will be performed for participants in Part J. For Part J, participants will be randomized in a 1:1 ratio to receive one of 2 treatments. Randomization will not be blinded.

#### 4.5. Blinding and Unblinding

Not applicable.

#### 4.6. Sampling Plan for Selecting 301 (COVE) Subjects

Approximately 584 Study P301 participants on primary series of mRNA-1273 will be selected to serve as the mRNA-1273 comparator (historical control arm) for Part A, B, C, and D, and immunogenicity data at Day 57 after the primary series of mRNA-1273 from these participants will be used to be compared with those at Day 29 after a booster dose in this study.

In Study P301, a random subcohort of study participants have already been selected for the secondary immunogenicity objective, including approximately 1,000 participants who were baseline SARS-CoV-2 negative and randomized to mRNA-1273. The PP Random Subcohort for Immunogenicity is the primary analysis population for the secondary immunogenicity objective in P301 (P301 SAP v2.0).

In Study P301, approximately 75% participants were ≥18 and <65 years old and 25% participants were ≥ 65 years old. In Study P205, for each study arm, the plan is to enroll participants with similar distribution of the 2 age groups (approximately 75% ≥ 18 and <65 years old and approximately 25% participants were ≥ 65 years old).

Therefore, a total of approximately 584 P301 participants with baseline SARS-CoV-2 negative status and randomized to mRNA-1273 with similar distribution of the 2 age groups will be selected from the P301 PP Random Subcohort as historical control for Study P205. Within each age group, P301 participants with baseline SARS-CoV-2 negative status and randomized to mRNA-1273 in the P301 PP Random Subcohort will be randomly selected.

If there is not enough number of participants in the P301 PP Random Subcohort either for an age group or overall, additional participants will be randomly selected from P301 who were not selected for the PP Random Subcohort. If the planned number of target sample size in P205 is changed, the number of participants from P301 to serve as historical control will be updated accordingly.

#### 4.7. External Comparator for Part F Cohort 1 - Study mRNA-1273-P201 Part B Subjects

Study mRNA-1273-P201 (P201) Part B, is the Open-Label Interventional Phase of the study, and was prompted by the authorization of a COVID-19 vaccine under an EUA. In Part B, all participants who previously received 1 or 2 injections of mRNA-1273 (50 µg or 100 µg) vaccine were able to receive a single booster dose of mRNA-1273 (50 µg). A total of approximately 150 participants in the P201 50 µg mRNA-1273 booster arm who were primed with 2 doses of 100 µg mRNA-1273 will serve as an external comparator for the P205 Part F 50 µg mRNA-1273.529 arm (Cohort 1).

### 5. Analysis Sets

The following analysis sets are defined: Full Analysis Set, Modified Intent-to-Treat Set, Per-Protocol Set for Immunogenicity, Per-Protocol Sensitivity Set for Immunogenicity, Per-Protocol Set for Immunogenicity – SARS-CoV-2 negative, Solicited Safety Set, Safety Set, and Per-Protocol Set for Efficacy. Definitions are the same across study Parts A (1, 2), B, C, D, E, F, G, H and J when applicable.

#### 5.1. Full Analysis Set

The Full Analysis Set (FAS) consists of all participants who receive IP.

#### 5.2. Modified Intent-to-Treat (mITT) Set

The mITT Set consists of all participants in the FAS who have no serologic or virologic evidence of prior SARS-CoV-2 infection (both negative RT-PCR test for SARS-CoV-2 and negative serology test based on bAb specific to SARS-CoV-2 nucleocapsid) pre-booster, ie, all FAS participants with baseline SARS-CoV-2 negative status pre-booster.

#### 5.3. Per-Protocol (PP) Set for Immunogenicity

The Per-Protocol (PP) Set for immunogenicity consists of all P205 participants in the FAS who meet all the criteria listed below, P301 historical control who were in the Per-protocol random subcohort for immunogenicity in P301, and P201 external comparator who were in the Per-Protocol Set for Part B.

**mRNA-1273-P205 participants**

a) Received the planned dose of study vaccination per schedule
b) For Parts A.1, A.2, B, C, D, F, G, H: had pre-booster and Day 29 (occurring between 21 and 42 days after vaccination) neutralizing antibody data; for Part J: had pre-booster and Day 15 neutralizing antibody data

- Part A.1: against B.1.351 variant
- Part A.2: against B.1.1.529 variant
- Part B: against ancestral SARS-CoV-2
- Part C: against B.1.617.2 variant
- Part D: against B.1.351 variant and B.1.617.2 variant
- Part F (Cohort 1 and Cohort 2): against B.1.1.529 variant
- Part G: against B.1.1.529 variant

- Part H: against BA.4/5 variant
- Part J: against XBB.1.5 variant

c) Had no major protocol deviations that impact key or critical data
d) Had no previous HIV infection

**mRNA-1273-P301 historical control**

Consists of mRNA-1273-P301 participants who were selected for historical control arm and who were in Per-Protocol set for immunogenicity in P301, i.e. Per-protocol Random Subcohort for immunogenicity in P301 (please refer to P301 SAP version 2.0 Section 6.7.2 for details).

**mRNA-1273-P201 external comparator**

Consists of P201B participants after 100 µg primary series who were in Per-Protocol Set for Part B (please refer to P201 SAP version 5.0 Section 5.6 for details).

The PP Set for immunogenicity will be used to summarize immunogenicity data and used as the primary analysis set for analyses of immunogenicity for immunobridging unless otherwise specified.

#### 5.4. Per-Protocol (PP) Sensitivity Set for Immunogenicity

For Parts A.1, A.2, B, C, D, F, G, H, the PP Sensitivity Set for immunogenicity will be used for sensitivity analysis and will consist of participants who meet all requirements for the PP Immunogenicity Set with the exception of the time window for Day 29 neutralizing antibody data.

**mRNA-1273-P205 participants**

a) Received the planned dose of study vaccination per schedule
b) Had pre-booster and Day 29

- Part A.1: against B.1.351 variant
- Part A.2: against B.1.1.529 variant
- Part B: against ancestral SARS-CoV-2
- Part C: against B.1.617.2 variant
- Part D: against B.1.351 variant and B.1.617.2 variant
- Part F (Cohort 1 and Cohort 2): against B.1.1.529 variant
- Part G: against B.1.1.529 variant

- Part H: against BA.4/5 variant

c) Had no major protocol deviations that impact key or critical data

d) Had no previous HIV infection

**mRNA-1273-P301 historical control**

Consists of mRNA-1273-P301 participants who were selected for historical control arm and who were in Per-Protocol set for immunogenicity in P301, i.e. Per-protocol Random Subcohort for immunogenicity in P301 (please refer to Section 6.4.2 for details).

##### 5.5. Per-Protocol Set for Immunogenicity – SARS-CoV-2 Negative (PPSI-Neg)

The PPSI-Neg Set consists of P205 participants who meet all requirements for the PP Immunogenicity Set and have no serologic or virologic evidence of SARS-CoV-2 infection at baseline and P201B participants who were in the Per-Protocol Set for Part B.

**mRNA-1273-P205 participants**

a) Received the planned dose of study vaccination per schedule

b) For Parts A.1, A.2, B, C, D, F, G, H: had pre-booster and Day 29 (occurring between 21 and 42 days after vaccination) neutralizing antibody data; for Part J: had pre-booster and Day 15 neutralizing antibody data

- Part A.1: against B.1.351 variant
- Part A.2: against B.1.1.529 variant
- Part B: against ancestral SARS-CoV-2
- Part C: against B.1.617.2 variant
- Part D: against B.1.351 variant and B.1.617.2 variant
- Part F (Cohort 1 and Cohort 2): against B.1.1.529 variant
- Part G: against B.1.1.529 variant
- Part H: against BA.4/5 variant
- Part J: against XBB.1.5 variant

c) Had no major protocol deviations that impact key or critical data

d) Had no previous HIV infection

e) Have no serologic or virologic evidence of SARS-CoV-2 infection at baseline (ie, who are SARS-CoV-2 negative, defined by both negative RT-PCR test for SARS-CoV-2 and negative serology test based on bAb specific to SARS-CoV-2 nucleocapsid)

**mRNA-1273-P201B external comparator**

Consists of P201B participants after 100 µg primary series who were in Per-Protocol Set for Part B (please refer to P201 SAP version 5.0 Section 5.6 for details).

The PPSI-Neg Set will used as the primary analysis set for analyses of immunogenicity for between booster comparisons for Part F, G, and H, and PPSI Set will be used as primary analysis set for part J.

#### 5.6. Solicited Safety Set

The Solicited Safety Set consists of all participants who receive IP and contribute any solicited AR data.

The Solicited Safety Set will be used for the analyses of solicited ARs. Participants will be included in the study arm corresponding to the dose of IP that they actually received.

#### 5.7. Safety Set

The Safety Set consists of all participants who receive IP.

The Safety Set will be used for all analyses of safety except for the solicited ARs.

#### 5.8. Per-Protocol (PP) Set for Efficacy

The PP Set for efficacy consists of all participants in the FAS who received the planned dose of study vaccination, who are SARS-CoV-2 negative pre-booster (ie, have a negative RT-PCR test for SARS-CoV-2 and a negative serology test based on bAb specific to SARS-CoV-2 nucleocapsid pre-booster), and have no major protocol deviations that impact key or critical data.

The PP Set for efficacy will be used as the primary analysis set for analyses of efficacy unless otherwise specified.

### 6. Statistical Analysis

#### 6.1. General Considerations

Please refer to the protocol for Schedule of Events (SoE).

Continuous variables will be summarized using the following descriptive summary statistics: the number of subjects (n), mean, standard deviation (SD), median, minimum (min), and maximum (max).

Categorical variables will be summarized using counts and percentages.

Baseline value, unless specified otherwise, is defined as the most recent non-missing measurement (scheduled or unscheduled) collected before the dose of IP in this study. Pre-booster and baseline are used interchangeably for the study arms in P205. For immunogenicity tests, the baseline is defined as the most recent non-missing measurement (scheduled or unscheduled) collected before the dose of IP.

For the summary statistics of all numerical variables, unless otherwise specified, the display precision will follow programming standards. Refer to Appendix A for variable display standards.

When count data are presented, the percentage will be suppressed when the count is zero in order to draw attention to the non-zero counts. “Missing” will be included in count tabulations where specified on the shells to account for dropouts and missing values. The denominator for all percentages will be the number of subjects in the corresponding group, unless otherwise specified.

Pre-booster SARS-CoV-2 status is determined by using virologic and serologic evidence of SARS-CoV-2 infection on or before Day 1 (pre-booster).

Positive SARS-CoV-2 status at pre-booster is defined as a positive RT-PCR test for SARS-CoV-2, and/or a positive serology test based on bAb specific to SARS-CoV-2 nucleocapsid (as measured by Roche Elecsys Anti-SARS-CoV-2 assay) on or before Day 1.

Negative status at pre-booster is defined as a negative RT-PCR test for SARS-CoV-2 and a negative serology test based on bAb specific to SARS-CoV-2 nucleocapsid (as measured by Roche Elecsys Anti-SARS-CoV-2 assay) on or before Day 1.

**Study day relative to the** injection will be calculated as follows:

a) study day prior to the injection will be calculated as: date of assessment/event – date of the injection;

b) study day on or after the date of the injection will be calculated as: date of assessment/event – date of the injection + 1;

For calculation of antibody levels/titers, antibody values reported as below LLOQ will be replaced by 0.5 × LLOQ. Values that are greater than the upper limit of quantification (ULOQ) will be converted to the ULOQ if actual values are not available. Missing results will not be imputed unless specified otherwise.

The following **analysis periods for safety analyses** will be used:

- Up to 28 days after vaccination: from the day of vaccination (Day 1) and continues through the earliest date of (the day of vaccination and 27 subsequent days, the day of study discontinuation). This analysis period will be used as the primary analysis period for safety analyses including unsolicited AEs, except for solicited ARs, unless specified otherwise.
- Throughout the study: from the day of vaccination (Day 1) and continues through the earliest date of (study completion, discontinuation from the study, or death).

**Unscheduled visits:** Unscheduled visit measurements will be included in the analysis as follows:

- In scheduled visit windows per specified visit windowing rules.
- In the derivation of baseline/last on-treatment measurements.
- In the derivation of maximum/minimum on-treatment values and maximum/minimum change from baseline values for safety analyses.
- In individual subject data listings as appropriate.

**Visit windowing rules:** The analysis visit windows for protocol-defined visits are provided in Appendix B.

**Incomplete/missing data:**

- Imputation rules for missing or incomplete days of medications, non-study vaccinations and procedures are provided in Appendix C.
- Imputation rules for missing or incomplete AE dates are provided in Appendix D.

- If the laboratory results are reported as below the LLOQ (e.g., <0.1), the numeric values will be imputed by 0.5 × LLOQ for the calculation of summary values. If the laboratory results are reported as greater than the ULOQ (e.g., >3000), the numeric values will be imputed by ULOQ in the summary if actual values are not available.
- Other incomplete/missing data will not be imputed, unless specified otherwise.

**Treatment groups**

- Part A.1: 50 µg of mRNA-1273.211 and 100 µg of mRNA-1273.211
- Part A.2: 50 µg of mRNA-1273.214
- Part B: 100 µg of mRNA-1273
- Part C: 50 µg of mRNA-1273.617.2 and 100 µg of mRNA-1273.617.2
- Part D: 50 µg of mRNA-1273.213 and 100 µg of mRNA-1273.213
- Part E: 100 µg of mRNA-1273.213
- Part F Cohort 1: 50 µg of mRNA-1273.529
- Part F Cohort 2: 50 µg of mRNA-1273.529 and 50 µg of mRNA-1273
- Part G: 50 µg of mRNA-1273.214
- Part H: 50 µg of mRNA-1273.222
- Part J: 50 µg of mRNA-1273.815, 50 µg of mRNA-1273.231

**External comparator**

- 100 µg of mRNA-1273 (historical control arm from Study mRNA-1273-P301 used for immunogenicity comparison only)
- 50 µg of mRNA-1273 (booster arm primed with 2 doses of 100 µg mRNA-1273 from Study mRNA-1273-P201 used as an external comparator for P205 Part F Cohort 1).

**Subgroup Analysis**

Immunogenicity will be assessed in the following subgroups:

- Age (18 to <65, and ≥65 years)
- Sex (female, male)
- Pre-booster SARS-CoV-2 status (negative, positive) if there is enough of pre-booster positives
- Race (White, Black or African American, Other)
- Ethnicity (Hispanic or Latino, Not Hispanic or Latino)

Safety may be assessed for the same subgroups.

Subgroup analyses may not be performed for Part J due to small sample size.

**Analyses Approach**

There are multiple parts in P205, all analyses and data summaries/displays will be provided by study arm for each study part using appropriate analysis population. For study parts with more than one dose level, data summaries/displays will be provided for each arm and for dose levels of the same vaccine type combined, unless otherwise specified.

**Subjects Received Off-study COVID-19 Vaccine**

If a participant received off-study COVID-19 vaccine, the participant's immune response and effectiveness data will be censored at the earlier date of date of off-study COVID-19 vaccine, database lock/data snapshot or data cutoff date, or date of last observed in study.

All analyses will be conducted using SAS Version 9.4 or higher.

#### 6.2. Background Characteristics

##### 6.2.1. Subject Disposition

The number and percentage of subjects in the following categories will be summarized as defined in Section 6.1 based on:

- Full Analysis Set
- Modified Intent-to-Treat Set
- Per-Protocol Set for Immunogenicity
- Per-Protocol Sensitivity Set for Immunogenicity
- Per Protocol Set for Immunogenicity - SARS-CoV-2 Negative
- Solicited Safety Set
- Safety Set
- Per-Protocol Set for efficacy

The percentage will be based on the number of subjects in all enrolled subjects.

The number and percentage of subjects in each of the following disposition categories will be summarized based on the Full Analysis Set:

- Received the dose of IP

- Completed study
- Prematurely discontinued the study and the reason for discontinuation

This study treatment only consists of a 1-dose booster, thus discontinuation from study treatment is not applicable to this study. A subject who completed 12 months of follow up after the injection is considered to have completed the study for study parts A.1, B, C, D, E, F, and G. A subject who completed 6 months of follow up after the injection is considered to have completed the study for study parts A.2, H and J.

A subject disposition listing for participants who discontinued the study early will be provided, including informed consent, subjects who were vaccinated, subjects who completed the study, subjects discontinued from the study, with reasons for discontinuation.

##### 6.2.2. Demographics

Descriptive statistics will be calculated for the following continuous demographic and baseline characteristics: age (years), weight (kg), height (cm), and body mass index (BMI) (kg/m2). Number and percentage of subjects will be provided for categorical variables such as age group (18 to <65, and ≥65 years), gender, race, ethnicity, pre-booster SARS-CoV-2 status, and time duration from completion of primary series to booster dose (205 first booster dose date minus P301 second vaccine dose date). The summaries will be provided based on the Safety Set and Per-Protocol immunogenicity Set. If the subjects in two or more analysis sets are identical, only one table will be provided for such analysis sets.

##### 6.2.3. Medical History

Medical history data will be coded by system organ class (SOC) and preferred term (PT) using the Medical Dictionary for Regulatory Activities (MedDRA).

The number and percentage of participants with any medical history will be summarized by SOC and PT based on the Safety Set. A participant will be counted only once for multiple events within each SOC and PT. SOCs will be displayed in internationally agreed order and, within each SOC, PTs will be displayed in descending order of frequency based on total (combined arm) and then alphabetically.

###### 6.2.4. Prior and Concomitant Medications

Prior and concomitant medications and non-study vaccination will be coded using the World Health Organization (WHO) drug dictionary (WHODD). The summary of concomitant medications will be based on the Safety Set.

The number and percentage of subjects using concomitant medications and non-study vaccinations during the 7-day follow-up period (i.e., on the day of injection and the 6 subsequent days) and during the 28-day follow-up period after the injection (i.e., on the day of injection and the 28 subsequent days) will be summarized as defined in Section 6.1 as follows:

- Any concomitant medications and non-study vaccination within 7 days Post Injection
- Any concomitant medications and non-study vaccination within 28 days Post Injection
- Seasonal influenza vaccine within 28 days Post Injection
- Antipyretic or analgesic medication within 28 days Post Injection

A summary table of concomitant medications and non-study vaccination that continued or newly received at or after the injection through 28 days will be provided by PT in descending frequency based on total (combined arms).

Medications taken to prevent pain or fever will be collected on eDiary and summaries will be provided based on the Solicited Safety Set as defined in Section 6.1, including within 7 days after injection, beyond 7 days after injection and after injection.

###### 6.2.5. Study Exposure

Summary of study exposure will be summarized.

Study duration, defined as time on study from the injection/booster to study discontinuation, study completion, last contact date, or data cutoff date, whichever occurs earlier, will be summarized.

###### 6.2.6. Major Protocol Deviations

Major protocol deviations are a subset of protocol deviations that may significantly impact the completeness, accuracy, or reliability of the study data or that may significantly affect a subject's rights, safety, or well-being. Major protocol deviations rules will be developed and finalized before database lock.

The number and percentage of the subjects with each major protocol deviation type will be provided as defined in Section 6.1 based on the FAS.

Major protocol deviations will be presented in a listing.

Select major protocol deviations are deemed to impact critical data and lead to exclusion from the Per-Protocol Set for Immunogenicity or Per-Protocol Set for Efficacy. Number of subjects with such major protocol deviations leading to exclusion from the PP Sets will be summarized.

###### 6.2.7. COVID-19 Impact

A listing will be provided for COVID-19 impact.

##### 6.3. Safety Analysis

Safety and reactogenicity will be assessed by clinical review of all relevant parameters including solicited ARs (local and systemic), unsolicited treatment-emergent AEs, treatment-related AEs, severe AEs, SAEs, MAAEs, AESIs, AEs leading to withdrawal from study participation, vital signs, and physical examinations-findings. Unsolicited treatment-emergent AEs will be coded by SOC and PT according to the MedDRA. The Toxicity Grading Scale for Healthy Adult and Adolescent Volunteers Enrolled in Preventative Vaccine Clinical Trials (DHHS 2007) is used in this study for solicited ARs.

Safety analyses will be based on the Safety Set, except that the Solicited Safety Set will be used for analyses of solicited AR.

###### 6.3.1. Adverse Events

A treatment-emergent AE (TEAE) is defined as any event occurring during the study not before exposure to study vaccine or any event already present that worsens after exposure to study vaccine. [Note: worsening of a pre-existing condition after vaccination will be reported as a new AE.]

Adverse events will also be evaluated by the investigator for the coexistence of MAAE which is defined as an AE that leads to an unscheduled visit to a healthcare practitioner.

Overview of unsolicited AEs will be summarized by stage, up to 28 days after vaccination and throughout the study (see Section 6.1).

All summary tables (except for the overall summary of AEs) for unsolicited AEs will be presented by SOC and PT for TEAEs with counts of subjects included. SOC will be displayed in internationally agreed order. PT will be displayed in descending order of frequency and then alphabetically within SOC. When summarizing the number and percentage of subjects with an event, subjects with multiple occurrences of the same AE or a continuing AE will be counted once. Subjects will be presented according to the highest severity in the summaries by severity, if subjects reported multiple events under the same SOC and/or PT. If severity (causality) is missing, the AE will be imputed as 'Severe' ('Related').

Percentages will be based upon the number of subjects in the Safety Set.

###### 6.3.1.1. Incidence of Adverse Events

An overall summary of unsolicited TEAEs including the number and percentage of subjects who experience the following will be presented:

- Any unsolicited TEAEs
- Any serious TEAEs
- Any fatal TEAEs
- Any unsolicited medically-attended TEAEs
- Any unsolicited TEAEs leading to discontinuation from participation in the study
- Any unsolicited severe TEAEs
- Any unsolicited AESIs
- Any unsolicited non-serious TEAEs
- Any unsolicited severe non-serious TEAEs

The table will also include number and percentage of subjects with unsolicited TEAEs that are treatment-related per PI's assessment in each of the above categories.

In addition, separate listings containing individual subject adverse event data for unsolicited AEs, unsolicited TEAEs leading to discontinuation from participation in the study, serious AEs and unsolicited medically-attended AEs will be provided separately.

###### 6.3.1.2. TEAEs by System Organ Class and Preferred Term

The following summary tables of TEAEs will be provided by SOC and PT using frequency counts and percentages (i.e., number and percentage of subjects with an event), up to 28 days after vaccination. Select TEAEs that will be collected throughout the study will also be summarized throughout the study when applicable:

- All unsolicited TEAEs
- All unsolicited TEAEs that are treatment-related
- All serious TEAEs
- All serious TEAEs that are treatment-related
- All unsolicited TEAEs leading to discontinuation from participation in the study
- All unsolicited Severe TEAEs
- All unsolicited Severe TEAEs that are treatment-related
- All unsolicited medically-attended TEAEs
- All unsolicited medically-attended TEAEs that are treatment-related
- All unsolicited AESIs
- All unsolicited AESIs that are treatment-related

##### 6.3.2. Solicited Adverse Reactions

###### 6.3.2.1. Analysis of Solicited Adverse Reactions

The solicited ARs are recorded by the subject in eDiary. If a solicited local or systemic AR continues beyond 7 days post injection, the participant will be prompted to capture solicited local or systemic AR in the eDiary until resolution.

Analyses of solicited ARs will be provided based on the Solicited Safety Set. The following summaries will be provided.

- Summary of SAR Within 7 Days (SAR eDiary and SAR eCRF)

  i. The number and percentage of subjects who reported each individual solicited local AR and solicited systemic AR during the 7-day follow-up period after the injection will be tabulated by severity grade, and by severity grade and day of reporting.

     A two-sided 95% exact confidence interval (CI) using the Clopper-Pearson method will be provided for the percentage of subjects who reported any solicited local AR, solicited systemic AR, or any solicited AR.

  ii. The number and percentage of subjects who reported each individual solicited local AR and solicited systemic AR during the 7-day follow-up period after the injection will be summarized by onset day (Day 1 through Day 7). The onset of individual solicited AR is defined as the time point after the injection at which the respective solicited AR first occurred.

- Summary of SAR Duration (SAR eDiary and SAR eCRF)

  i. Duration is calculated as the last day – the first day + 1 when the solicited adverse reaction was reported starting within the 7 days of injection.

- Summary of SAR Persisting Beyond 7 Days (SAR eDiary and SAR eCRF)

  i. The number and percentage of subjects who reported each individual solicited local AR and solicited systemic AR that persist beyond 7 days after the injection (i.e., occurred before day 7, but persisting after day 7 regardless of duration) will be tabulated by severity grade.

     A two-sided 95% exact confidence interval (CI) using the Clopper-Pearson method will be provided for the percentage of subjects who reported any solicited local AR, solicited systemic AR, or any solicited AR persisting beyond 7 days after injection.

- Summary of SAR with Onset after Day 7 (SAR eCRF only)

i. The incidence for each individual solicited local AR and solicited systemic AR with onset day after the 7-day follow-up period after the injection (i.e., after Day 7) will be tabulated.

ii. The onset day of each individual solicited local AR and solicited systemic AR with onset day after the 7-day follow-up period after the injection (i.e., after Day 7) will be summarized descriptively.

iii. The number of days reporting each individual solicited local AR and solicited systemic AR with onset after the 7-day follow-up period after the injection (i.e., after Day 7) will be summarized descriptively, similar to SAR duration summary.

- Summary of Onset Day for Local Reactions (SAR eDiary and SAR eCRF)

i. The number and percentage of subjects who reported local reactions will be tabulated by onset day (within 7 days and beyond). The onset day is defined similarly in 'Summary of SAR Within 7 Days' section.

##### 6.3.3. Pregnancy Tests

A point-of-care urine pregnancy test will be performed on Day 1. At any time, a pregnancy test either via blood or point-of-care urine can be performed, at the discretion of the investigator. A by-subject listing will be provided for pregnancy tests with positive results.

##### 6.3.4. Vital Sign Measurements

Vital sign measurements, including systolic and diastolic blood pressures, heart rate, respiratory rate, and body temperature, will be collected at the time points indicated in the SoE table in the protocol with pre- and post-dosing on the day of injection (Day 1) only and they will be presented in a listing. The abnormalities meeting the toxicity grading criteria (Grade 2 or higher) in any vital sign measurement will be provided in the listing.

Shift from baseline in the toxicity grades at each time point and shift from baseline in the toxicity grades to the worst post-baseline result will also be summarized.

#### 6.4. Immunogenicity Analysis

The primary analysis population for immunogenicity will be the PP Set for Immunogenicity for immunobridging to primary series analysis and the PPSI-Neg will be the primary analysis population for immunogenicity analyses for between booster group comparisons and are summarized in Appendix E. Each arm in each study part will be evaluated separately and compared to the historical control arm of mRNA 1273 primary series. Exploratory analyses may be performed to compare between booster dose, refer to Appendix E for a summary of immunogenicity endpoints.

The GMT and geometric mean (GM) level will be calculated using the following formula:

$$10^{\left\{\frac{\sum_{i=1}^{n}\log_{10}(t_i)}{n}\right\}}$$

where $t_1, t_2, \ldots, t_n$ are *n* observed immunogenicity titers or levels.

The geometric mean fold-rise (GMFR) measures the changes in immunogenicity titers or levels within subjects. The GMFR will be calculated using the following formula:

$$10^{\left\{\frac{\sum_{i=1}^{n}\log_{10}\left(v_{ij}/v_{ik}\right)}{n}\right\}} = 10^{\left\{\frac{\sum_{i=1}^{n}\log_{10}(v_{ij})-\log_{10}(v_{ik})}{n}\right\}}$$

where, for *n* subjects, $v_{ij}$ and $v_{ik}$ are observed immunogenicity titers or levels for subject *i* at time points *j* and *k*, $j \neq k$

##### 6.4.1. Immunogenicity Assessments

Blood samples for immunogenicity assessments will be collected at the time points indicated in the SoE table in the protocol.

##### 6.4.2. Selecting Study P301 (COVE) Subjects as Historical Control and P201B Subjects as External Comparator

Please refer to Section 4.6 regarding the selection/sampling plan of COVE participants as historical control whose immunogenicity 28 days after completion of the primary series will be compared with that for each of the P205 study arm after a single booster dose.

For each age group (<65, ≥ 65), simple random sampling was used with a random number seed of 231.

Please refer to Section 4.7 regarding the selection of P201B participants as an external comparator whose immunogenicity will be compared with that for 50 µg mRNA-1273.529 arm (Part F, Cohort 1).

##### 6.4.3. Primary Analysis of Antibody-Mediated Immunogenicity Endpoints

**Part A.1**

Each dose level of mRNA-1273.211 (50 and 100 µg) will be assessed with respect to mRNA-1273 primary series historical control for the primary objective on immune response. Pseudotyped virus neutralizing antibody will be used as the basis to assess non-inferiority in immune response.

For the primary objective on immune response, there are 4 null hypotheses to be tested for each arm (see Section 4.2).

Test of $H_0^1$

In order to test the first hypothesis of whether or not a single booster dose of mRNA-1273.211 is inferior to the two priming doses of 100 µg of mRNA-1273, an analysis of variance (ANCOVA) will be carried out with antibody titers (Day 29 on 50 or 100 µg mRNA-1273.211 and Day 57 on mRNA-1273 primary series historical control) against the ancestral SARS-CoV-2 as a dependent variable and a group variable (50 or 100 µg mRNA-1273.211 and mRNA-1273 primary series historical control) as the fixed effect, adjusting for age group (<65, ≥ 65). The GMT values will be estimated by the geometric least square mean (GLSM) from the model for each group and corresponding 95% confidence interval (CI) will be provided for each group. The GMR (ratio of GMTs) for each dose level of mRNA-1273.211 with respect to mRNA-1273 will be estimated by the ratio of GLSM from the model. The 95% CI for the ratio of GLSM will be provided to assess the between group difference in immune response against the ancestral SARS-CoV-2 for each dose level of mRNA-1273.211 at Day 29 compared to mRNA-1273 primary series historical control at Day 57.

For each dose level, the hypothesis will be rejected (ie, non-inferiority of immune response to mRNA-1273 based on GMT ratio will be considered demonstrated) if the lower bound of the corresponding 95% CI of the GMR against the ancestral SARS-CoV-2 is > 0.667 based on the non-inferiority margin of 1.5.

Test of $H_0^2$

In order to test the second hypothesis of whether or not a single booster dose of mRNA-1273.211 is inferior to two priming doses of 100 µg of mRNA-1273, the number and percentage (rate) of participants achieving seroresponse at Day 29 will be summarized with 95% CI calculated using the Clopper-Pearson method for each group. The difference of SRRs between each dose level of mRNA-1273.211 at Day 29 and mRNA-1273 primary series historical control at Day 57 in P301 against ancestral SARS-CoV-2 will be calculated with 95% CI using Miettinen-Nurminen (score) method.

The non-inferiority in SRR of each dose level of MRNA-1279.211 compared to mRNA-1273 primary series historical control will be considered demonstrated if the lower bound of the 95% CI of the SRR difference is > -10% based on the non-inferiority margin of 10%.

The same analysis methods for immune response against variant B.1.351 will be used for $H_0^3$ and $H_0^4$ hypotheses testing.

For each dose level of mRNA-1273.211, the primary immunogenicity objective (against ancestral SARS-CoV-2 and variant B.1.351) is considered met if non-inferiority is demonstrated based on both GMR and SRR difference, specifically:

- If the lower bound of the 95% CI of the GMT ratio against the ancestral SARS-CoV-2 between mRNA-1273.211 and mRNA-1273 is > 0.667 based on the non-inferiority margin of 1.5, and
- The lower bound of the 95% CI of the SRR difference (mRNA-1273.211 - mRNA-1273) against ancestral SARS-CoV-2 is > -10%, and

- The lower bound of the 95% CI of the GMT ratio between mRNA-1273.211 against the variant (B.1.351) at Day 29 as compared to 100 µg mRNA-1273 against the ancestral SARS-CoV-2 is at Day 57 > 0.667 based on the non-inferiority margin of 1.5, and
- The lower bound of the 95% CI of the SRR difference (mRNA-1273.211 against the variant [B.1.351] at Day 29 – 100 µg mRNA-1273 against ancestral SARS-CoV-2 at Day 57) is > -10%.

**Part A.2**

For Part A.2 participants, Day 29 and Day 181 immune response after mRNA-1273.214 (50 µg) as a second booster dose will be compared with their own Day 29 and Day 181 immune response of mRNA-1273.211 (50 µg) received as the first booster dose. GMT ratios will be calculated by back transforming the mean of paired differences of antibody titer data on the logarithmic scale between Day 29 and Day 181 post mRNA-1273.214 and Day 29 and Day 181 of antibody titer data post mRNA-1273.211. CIs for the GMT ratio will be based on t-distribution of the log-transformed values then back transformed to the original scale for presentation. Seroresponse rates at Day 29 and Day 181 post mRNA-1273.214 will be compared with their seroresponse rates at Day 29 and Day 181 post mRNA-1273.211. The difference in seroresponse rates and its corresponding 95% CI based on adjusted Wald method will be provided.

**Part B**

For the primary objective on immune response for a single booster dose of 100 µg mRNA-1273, there are 2 null hypotheses to be tested (see Section 4.2), and Part B would be considered to meet its primary objective if both null hypotheses are rejected. The same analysis methods described for Part A will be used for the primary immunogenicity objective for Part B. The primary immunogenicity objective (against ancestral SARS-CoV-2 mRNA-1273) is considered met if non-inferiority is demonstrated based on both GMR and SRR difference.

**Part C**

Each dose level of mRNA-1273.617.2 (50 and 100 µg) will be assessed with respect to mRNA-1273 primary series historical control separately. For the primary objective on immune response for a single booster dose of 100 µg mRNA-1273.617.2, there are 2 null hypotheses to be tested for each arm (see Section 4.2). The same analysis methods described for Part A will be used for the primary immunogenicity objective for Part C. The primary immunogenicity objective (against the variant strain) is considered met if non-inferiority is demonstrated based on both GMR and SRR difference.

**Part D**

Each dose level of mRNA-1273.213 (50 and 100 µg) will be assessed with respect to mRNA-1273 primary series historical control separately. For the primary objective on immune response for a single booster dose of 100 µg mRNA-1273.617.2, there are 4 null hypotheses to be tested for each arm (see Section 4.2). The same analysis methods described for Part A will be used for the primary immunogenicity objective for Part D. The primary immunogenicity objective (against the variant strain) is considered met if non-inferiority is demonstrated based on both GMR and SRR difference.

**Part F Cohort 1**

Primary analysis set for immunogenicity objectives will be based on PPSI-Neg.

For the primary objective on immune response for a first booster dose of 50 µg mRNA-1273.529, see Section 4.2. An analysis of covariance (ANCOVA) model will be performed to assess the difference in immune response between mRNA-1273.529 and mRNA-1273 (P201B 50 µg booster after 100 µg primary series) as the first booster dose. For immune response against the B.1.1.529 strain, in the ANCOVA model, antibody titers at Day 29 post-booster against the B.1.1.529 strain will be a dependent variable, and a group variable (mRNA-1273.529 and mRNA-1273) will be the fixed effect, adjusting for age groups (< 65, ≥ 65 years) and pre-booster antibody titer level if applicable; the model may also be adjusted for other characteristics.

The GMT will be estimated by the GLSM from the model and its corresponding 95% will be provided for each group. The GMR (ratio of GMTs) for mRNA-1273.529 with respect to mRNA-1273 will be estimated by the ratio of GLSM from the model and the corresponding 95% CIs will be provided. The 95% CI for GMR will be used to assess the between group difference in immune response against the B.1.1.529 strain for mRNA-1273.529 at Day 29 compared to the mRNA-1273.

The number and percentage (rate) of participants achieving seroresponse at Day 29 will be summarized with 95% CI calculated using the Clopper-Pearson method for each group.

The difference of SRR between mRNA-1273.529 and mRNA-1273 will be calculated with 95% CI based on stratified Miettinen-Nurminen method adjusted for age group. The non-inferiority in SRR of mRNA-1273.529 compared to mRNA-1273 will be considered demonstrated if the lower bound of the 95% of the SRR difference is > -10% based on the non-inferiority margin of 10%.

The primary immunogenicity objective (against the variant B.1.1.529) is considered met if non-inferiority is demonstrated based on GMR and SRR difference, specifically:

- If the lower bound of the 95% CI of the GMT ratio between mRNA1273.529 against the variant (B.1.1.529) at Day 29 as compared to 50 µg mRNA1273 against B.1.1.529 at Day 29 is > 0.667 based on the non-inferiority margin of 1.5.
- the lower bound of the 95% CI of the SRR difference (50 µg mRNA-1273.529 against the variant B.1.1.529 at Day 29 - 50 µg mRNA-1273 against B.1.1.529) is > -10%.
- If non-inferiority is demonstrated, the lower bound of 95% CI of the GMT ratio will be compared to 1, if it's greater than 1, then superiority is demonstrated.

A supportive analysis for the primary immunogenicity endpoints may also be performed using Per-Protocol Set for Immunogenicity. For immune response against the B.1.1.529 strain, in the ANCOVA model, antibody titers at Day 29 post-booster against the B.1.1.529 strain will be a dependent variable, and a group variable (mRNA-1273.529 and mRNA-1273) will be the fixed effect, adjusting for age groups (< 65, ≥ 65 years), pre-booster SARS-CoV-2 status, and pre-booster titer if applicable; the model may also be adjusted for other characteristics. The difference of SRR between mRNA-1273.529 and mRNA-1273 will be calculated with 95% CI based on stratified Miettinen-Nurminen method adjusted for pre-booster SARS-CoV-2 status and age group.

**Part F Cohort 2**

Primary analysis set for immunogenicity objectives will be based on PPSI-Neg.

50 µg mRNA-1273.529 as the second booster dose will be compared to 50 µg mRNA-1273 as the second booster dose. For the primary objective on immune response for a second booster dose of 50 µg mRNA-1273.529, see Section 4.2. The same analysis

methods described for Part F Cohort 1 will be used for the primary immunogenicity objective for Part F Cohort 2.

The primary immunogenicity objective (against the B.1.1.529) is considered met if non-inferiority is demonstrated based on GMR and SRR difference, specifically:

- The lower bound of the 95% CI of the GMT ratio between mRNA1273.529 against the variant (B.1.1.529) at Day 29 as compared to 50 µg mRNA-1273 against B.1.1.529 at Day 29 is > 0.667 based on the non-inferiority margin of 1.5.
- The lower bound of the 95% CI of the SRR difference (50 µg mRNA-1273.529 against the variant B.1.1.529 at Day 29 - 50 µg mRNA-1273 against B.1.1.529 at Day 29) is >-10%.
- If non-inferiority is demonstrated (based on GMT ratio and SRR difference), the lower bound of 95% CI of GMT ratio will be compared to 1, if it's greater than 1, then superiority is demonstrated.

Analyses for the primary immunogenicity endpoints will also be performed using Per-Protocol Set for Immunogenicity.

**Part G**

Primary analysis set for immunogenicity objectives will be based on PPSI-Neg.

50 µg mRNA-1273.214 as the second booster dose will be compared to 50 µg mRNA-1273 as the second booster dose (active control arm in Part F, Cohort 2). For the primary objective on immune response for a second booster dose of 50 µg mRNA-1273.214, hypotheses are to be tested at Day 29 and Day 91 (see Section 4.2). For the primary objective of immune response, an alpha of 0.05 (two-sided) will be allocated to the two time points. Day 29 and Day 91 will each have an alpha of 0.025 (two-sided) for hypotheses testing. The analyses method described in Part F Cohort 1 will be used for Part G.

The primary immunogenicity objective is considered met if non-inferiority against B.1.1.529 based on GMR and SRR difference and non-inferiority against the ancestral SARS-CoV-2 based on GMR are demonstrated based on GMR at Day 29 or Day 91.

Day 29: alpha = 0.025 (2-sided)

- The lower bound of the 97.5% CI of the GMT ratio between mRNA-1273.214 against the variant (B.1.1.529) at Day 29 as compared to 50 µg mRNA-1273 against B.1.1.529 at Day 29 is > 0.667 based on the non-inferiority margin of 1.5.
- The lower bound of the 97.5% CI of the SRR difference (50 µg mRNA-1273.214 against the variant B.1.1.529 at Day 29 - 50 µg mRNA-1273 against B.1.1.529 at Day 29) is >-10%.
- The lower bound of the 97.5% CI of the GMT ratio between mRNA-1273.214 against ancestral SARS-CoV-2 at Day 29 as compared to 50 µg mRNA-1273 against ancestral SARS-CoV-2 at Day 29 is > 0.667 based on the non-inferiority margin of 1.5.
- If non-inferiority is demonstrated for both B.1.1.529 (based on GMR and SRR) and ancestral SARS-CoV-2 (based on GMR), the lower bound of 97.5% CI of GMR will be compared to 1, if it's greater than 1, then superiority against B.1.1.529 is demonstrated.

Analyses for the primary immunogenicity endpoints will also be performed using Per-Protocol Set for Immunogenicity.

Day 91: alpha=0.025 (2-sided)

Hypotheses testing at Day 91 will be performed in the same manner, first test two non-inferiority hypotheses (two against the B.1.1.529 strain and one against ancestral SARS-CoV-2) at alpha of 0.025 level (two-sided). Once non-inferiority is demonstrated for both B.1.1529 and ancestral SARS-CoV-2, then superiority testing against the B.1.1.529 at alpha of 0.025 level (two-sided) will be performed.

For the key secondary objective, hypotheses to be tested (Day 29 and Day 91 each with alpha level of 0.025, 2-sided) are described in Section 4.2. If the lower bound of the 97.5% CI of the SRR difference (50 µg mRNA-1273.214 against the ancestral SARS-CoV-2 - 50 µg mRNA-1273 against ancestral SARS-CoV-2) is >-10% at Day 29 or Day 91, then key secondary objective will be considered met.

In the event that an early assessment of the 1273.214 data is needed due to public health concerns, a two-staged approach will be used. Specifically, a subset of participants' (ie, 50 first enrolled participants) serum samples will first be tested against ancestral SARS-CoV-2 and various VOCs. For the Day 29 and Day 91 immunogenicity analyses, all participants' immune data will be used in the formal analysis to evaluate the primary immunogenicity objective.

**Part H**

Primary analysis set for immunogenicity objectives will be based on PP Set for Immunogenicity, SARS-CoV-2 negative.

50 µg mRNA-1273.222 as the second booster dose will be compared to 50 µg mRNA-1273 as the second booster dose (active control arm in Part F, Cohort 2).

Study Part H was enrolled in August 2022 and Part F 50 µg mRNA-1273 was enrolled in February 2022, baseline and demographic characteristics will be examined between the two study parts before comparing immunogenicity data.

An analysis of covariance (ANCOVA) model will be performed to assess the difference in immune response between mRNA-1273.222 and mRNA-1273. In the ANCOVA model, antibody titers at Day 29 post-booster against the Omicron BA. 4/5 strain will be a dependent variable, and a group variable (mRNA-1273.222 and mRNA-1273) will be the fixed effect, adjusting for age, pre-booster antibody titer level if applicable; the model may also be adjusted for other characteristics. Age group (≥65, <65 years) or age as continuous variable may be considered in the ANCOVA model.

The GMT will be estimated by the GLSM from the model and its corresponding 95% will be provided for each group. The GMR (ratio of GMTs) for mRNA-1273.222 with respect to mRNA-1273 will be estimated by the ratio of GLSM from the model and the corresponding 95% CIs will be provided. The 95% CI for GMR will be used to assess the between group difference in immune response against the BA. 4/5 strain for mRNA-1273.222 at Day 29 compared to the mRNA-1273.

The number and percentage (rate) of participants achieving seroresponse at Day 29 will be summarized with 95% CI calculated using the Clopper-Pearson method for each group. The difference of SRR between mRNA-1273.222 and mRNA-1273 will be calculated with 95% CI based on stratified Miettinen-Nurminen (MN) method adjusted for age group. The difference of SRR and it’s 95% CI using unstratified MN method may also be provided as a sensitivity analysis.

A supportive analysis for the primary immunogenicity endpoints may also be performed using Per-Protocol Set for Immunogenicity. For immune response against the BA.4/5 strain, in the ANCOVA model, antibody titers at Day 29 post-booster against the BA.4/5 strain will be a dependent variable, and a group variable (mRNA-1273.222 and mRNA-1273) will be the fixed effect, adjusting for age, pre-booster SARS-CoV-2 status, pre-booster titer if applicable; the model may also be adjusted for other characteristics. Age group (≥65, <65 years) or age as continuous variable may be considered in the ANCOVA model. The difference of SRR between mRNA-1273.222 and mRNA-1273 will be calculated with 95% CI based on stratified Miettinen-Nurminen method adjusted for pre-booster SARS-CoV-2 status and age group.

The primary immunogenicity objective (against BA.4/5 and ancestral SARS-CoV-2 D614G) is considered met if non-inferiority is demonstrated based on GMR and SRR difference, specifically:

- The lower bound of the 95% CI of the GMT ratio between mRNA1273.222 against Omicron BA.4/5 at Day 29 as compared to 50 µg mRNA-1273 against Omicron BA.4/5 at Day 29 is > 0.667 based on the non-inferiority margin of 1.5.
- 50 µg mRNA 1273.222, as a second booster dose, against Omicron BA.4/5 is non-inferior to the second booster dose of (50 µg) mRNA 1273 against Omicron BA.4/5 based on the difference in SRR at Day 29 based on a non-inferiority boundary, if:
    - the lower bound of the 95% CI of the SRR difference (50 µg mRNA-1273.222 against Omicron BA.4/5 at Day 29 - 50 µg mRNA-1273 against Omicron BA.4/5 at Day 29) is >-5%, then the non-inferiority of mRNA-1273.222 against Omicron BA. 4/5 compared to that of mRNA-1273 is demonstrated based on a non-inferiority margin of 5%.
    - the lower bound of the 95% CI of the SRR difference (50 µg mRNA-1273.222 against Omicron BA.4/5 at Day 29 - 50 µg mRNA-1273 against Omicron BA.4/5 at Day 29) is >-10% but ≤-5%, then non-inferiority of mRNA-1273.222 against Omicron BA. 4/5 compared to that of mRNA-1273 is demonstrated based on a non-inferiority margin of 10%.
- The lower bound of the 95% CI of the GMT ratio between mRNA1273.222 against the ancestral SARS-CoV-2 D614G at Day 29 as compared to 50 µg mRNA-1273

against the ancestral SARS-CoV-2 D614G at Day 29 is > 0.667 based on the non-inferiority margin of 1.5.

- The lower bound of the 95% CI of the SRR difference (50 µg mRNA-1273.222 against the ancestral SARS-CoV-2 D614G at Day 29 - 50 µg mRNA-1273 against ancestral SARS-CoV-2 D614G at Day 29) is >-10%.
- If non-inferiority is demonstrated (against Omicron BA. 4/5 and ancestral SARS-CoV-2 D614G based on GMT ratio and SRR difference with a NI margin of 10%), the lower bound of 95% CI of GMT ratio will be compared to 1, if it's greater than 1, then superiority is demonstrated.

**Part J**

No formal hypothesis testing will be performed for Part J and all safety and immunogenicity analyses will be descriptive.

The primary immunogenicity objective will be assessed using the Per-Protocol Set for Immunogenicity. Descriptive comparisons on immunogenicity response between the mRNA-1273.815 and mRNA-1273.231 treatment arms will be performed.

An ANCOVA model will be used to compare antibody response between the two treatment arms at Day 15 and Day 29. Antibody titers (against SARS-CoV-2 variants) will be included in the model as a dependent variable and treatment group variable (mRNA-1273.815, mRNA-1273.231) will be included as fixed effect. The model may adjust for other covariates as appropriate (e.g. pre-booster SARS-CoV-2 status etc.). Geometric mean ratio and the corresponding 95% CI will be estimated from the model.

For assays with established LLOQ, SRR will be derived for the antibody response, SRR difference at Day 15 and Day 29 will be compared and 95% CI will be provided using Miettinen-Nurminen method.

For each study part, reverse cumulative distribution plots and box plots of titers or levels against will be generated for each antibody of interest.

##### 6.4.4. Secondary Analysis of Antibody-Mediated Immunogenicity Endpoints

The same analysis methods described for the primary analysis of Part A.1 will be used to evaluate each booster arm against variant strain or ancestral SARS-CoV-2 (Part A.1, B, C,

D, F Cohorts 1 and 2, and G) as compared with 100 µg mRNA-1273 primary series against ancestral SARS-CoV-2, specifically:

- To compare immune response to circulating variants of concern after a single booster dose of 50 µg or 100 µg mRNA-1273.211 (Part A.1) with 100 µg mRNA-1273 primary series against ancestral SARS-CoV-2.
- To compare immune response to circulating variants of concern after a single booster dose of 100 µg mRNA-1273 (Part B) with 100 µg mRNA-1273 primary series against ancestral SARS-CoV-2.
- To compare immune response to ancestral SARS-CoV-2 and circulating variants of concern after a single booster dose of mRNA-1273.617.2 (Part C) with 100 µg mRNA-1273 primary series against ancestral SARS-CoV-2.
- To compare immune response to ancestral SARS-CoV-2 and circulating variants of concern after a single booster dose of 50 µg or 100 µg mRNA-1273.213 (Part D) with 100 µg mRNA-1273 primary series against ancestral SARS-CoV-2.
- To compare immune response to a first booster dose of 50 µg mRNA-1273.529 (Part F Cohort 1) against Omicron variant with 100 µg mRNA-1273 primary series against ancestral SARS-CoV-2.
- To compare immune response to a second booster dose of 50 µg mRNA-1273.529 (Part F Cohort 2) against Omicron variant with 100 µg mRNA-1273 primary series against ancestral SARS-CoV-2.
- To compare immune response to a second booster dose of 50 µg mRNA-1273.214 (Part G) against Omicron variant with 100 µg mRNA-1273 primary series against ancestral SARS-CoV-2.

For Part E, immune response to ancestral SARS-CoV-2 and circulating variants for concern after a single booster dose of 100 µg mRNA-1273.213 will be summarized descriptively.

The mixed effect model repeated measure (MMRM) will be used to analyze all post-booster measures for between booster comparisons (Part F Cohort 1, Part F Cohort 2, Part G, and Part H) when analyzing immunogenicity data at Month 6 and Month 12 (if applicable), the model will include treatment group, analysis visit, treatment by visit interaction, and adjusting for age groups and pre-booster titer levels, and pre-booster

SARS-CoV-2 status when applicable (i.e. using per-protocol immunogenicity set). An unstructured covariance structure will be used to model the within-participant errors. A Kenward-Roger approximation will be used for the denominator degrees of freedom. If the model does not converge, a compound symmetry covariance structure will be used. The GMT will be estimated from the model and its corresponding 95% CI will be provided for each group at each post-boost timepoint. The GMR (ratio of GMTs) will be estimated from the model and the corresponding 95% CI will be provided at each post-boost timepoint.

##### 6.4.5. Exploratory Analysis of Antibody-Mediated Immunogenicity Endpoints

For each booster arm, the following evaluations will be performed at each time point at which blood samples are collected for immunogenicity (unless otherwise specified).

- In relation to the immune response of each booster arm to other SARS-CoV-2 viral variants, the GMT, geometric mean fold rise (GMFR) and seroresponse rate will be calculated at the time points where the immune response is assessed for such variants.
- For each antibody of interest, the GMT or level with corresponding 95% CI at each time point will be provided. The 95% CIs will be calculated based on the t-distribution of the log-transformed values then back-transformed to the original scale for presentation. The following descriptive statistics will also be provided at each time point: number of participants (n), median, minimum, and maximum. Additionally, reverse cumulative distribution plots and box plots of titers or levels will be generated for each antibody of interest.
- For each antibody of interest GMFR of post-baseline titers or levels over baseline with their corresponding 95% CIs at each post-baseline time point will be provided. The 95% CIs will be calculated based on the t-distribution of the log-transformed values then back-transformed to the original scale for presentation. The following descriptive statistics will also be provided at each time point: number of participants (n), median, minimum, and maximum. Additionally, reverse cumulative distribution plots and box plots of titers or levels will be generated for each antibody of interest.
- For each antibody of interest, the proportion of subjects with fold-rise ≥ 2, fold-rise ≥ 3, and fold-rise ≥ 4 from baseline at each post-injection time point will be tabulated with 95% CI calculated using the Clopper-Pearson method.

- Seroresponse rate of each booster arm against the ancestral SARS-CoV-2 and variants, defined as the percentage of participants achieving seroresponse against the ancestral SARS-CoV-2 strain and variants respectively, will be provided with the 95% CI calculated using the Clopper-Pearson method.

The following exploratory analyses may be performed to compare booster regimens using the same analysis methods described in Section 6.4.3 Part A.1.

- GMT ratio and SRR difference of 50 µg mRNA-1273.211 (Part A.1) compared to 100 µg mRNA-1273.211 (Part A.1) against the same viral strains.
- GMT ratio and SRR difference of 50 or 100 µg mRNA-1273.211 (Part A.1) as a booster dose against ancestral SARS-CoV-2 and variants compared to a booster dose of 100 µg mRNA-1273 (Part B) against the ancestral SARS-CoV-2 and variants.
- GMT ratio and SRR difference of 50 µg mRNA-1273.617.2 (Part C) compared to 100 µg mRNA-1273.617.2 (Part C) against the same viral strains.
- GMT ratio and SRR difference of 50 µg or 100 µg mRNA-1273.617.2 (Part C) as a booster dose against ancestral SARS-CoV-2 and variants compared to a booster dose of 100 µg mRNA-1273 (Part B) against the ancestral SARS-CoV-2 and variants.
- GMT ratio and SRR difference of 50 µg or 100 µg mRNA-1273.617.2 (Part C) as a booster dose against ancestral SARS-CoV-2 and variants compared to a booster dose of 50 or 100 µg mRNA-1273.211 (Part A.1) against the ancestral SARS-CoV-2 and variants.
- GMT ratio and SRR difference of 50 µg mRNA-1273.213 (Part D) compared to 100 µg mRNA-1273.213 (Part D) against the same viral strains.
- GMT ratio and SRR difference of 50 µg or 100 µg mRNA-1273.213 (Part D) as a booster dose against ancestral SARS-CoV-2 and variants compared to a booster dose of 50 or 100 µg mRNA-1273.211 (Part A.1) against the ancestral SARS-CoV-2 and variants.
- GMT ratio and SRR difference of 50 µg or 100 µg mRNA-1273.213 (Part D) as a booster dose against ancestral SARS-CoV-2 and variants compared to a booster

dose of 100 µg mRNA-1273 (Part B) against the ancestral SARS-CoV-2 and variants.

- GMT ratio and SRR difference of 50 µg or 100 µg mRNA-1273.213 (Part D) as a booster dose against ancestral SARS-CoV-2 and variants compared to a booster dose of 50 µg or 100 µg mRNA-1273.617.2 (Part C) against the ancestral SARS-CoV-2 and variants.

##### 6.4.6. Sensitivity Analysis

Sensitivity analysis for the immunogenicity endpoints may be performed with the same methods described above based on the PP Immunogenicity Sensitivity Set.

Sensitivity analysis may be performed for the primary immunogenicity endpoints by excluding all records after SARS-CoV-2 infection.

Additional sensitivity analysis may be performed to assess robustness of the primary immunogenicity analysis results for GMR if more than 10% immunogenicity data are missing at Day 29 or Day 91 (Part G) in the Per-Protocol Set for immunogenicity Set. Multiple imputation will be used to impute for missing antibody titer data. Following steps outline multiple imputation process and subsequent analysis.

Step 1: We will assume antibody titer data follows a log normal distribution.

- Markov Chain Monte Carlo (MCMC) will be used for imputation, imputation model will include treatment group, pre-booster baseline titer, age group, Day 29/ Day 91 (when applicable) titer data in log scale (with a base of 10). Each missing data point will be filled with a set of imputed values.

Step 2: Generate a set of datasets with imputed antibody titer values.

Step 3: Each complete dataset will be analyzed using ANCOVA model with treatment group as a fixed effect, adjusting for pre-booster baseline tier, and age group. Each model will estimate GMR (between treatment group comparison) and standard error in Log 10 scale.

Step 4: The results from these analyses will be combined into a single estimate using Rubin’s (1987) method, the combined estimate will be transformed back to original scale for presentation.

###### 6.4.7. Seroresponse

Seroresponse is defined as ≥ 4*LLOQ for those with baseline <LLOQ; ≥ 4-foldrise for those with baseline ≥ LLOQ.

Seroresponse will be derived based on two types of baselines:

1) Pre-vaccination (Pre-Dose 1 of the primary series)
2) Pre-booster baseline

Both definitions will be used when comparing seroresponse rate for all study parts. Seroresponse based on change (fold rise) from pre-dose 1 of the primary series would be considered the primary approach of seroresponse.

For subjects without pre-Dose 1 antibody titer information, seroresponse is defined as ≥ 4*LLOQ for subjects with negative SARS-CoV-2 status at their pre-dose 1 of primary series, and these subjects antibody titer will be deemed <LLOQ at pre-dose 1 of primary series.

- For subjects who are without SARS-CoV-2 status information at pre-dose 1 of primary series, their pre-booster SARS-CoV-2 status will be used to impute their SARS-CoV-2 status at their pre-dose 1 of primary series.

##### 6.5. Efficacy Analysis

Vaccine efficacy will not be formally assessed in this trial but active surveillance for COVID-19 and SARS-CoV-2 infection through weekly contact and blood draws (see SoE, table in the protocol), will be performed.

Pre-booster SARS-CoV-2 status is described in Section 6.1. Pre-booster SARS-CoV-2 status, the serology test results based on Roche Elecsys assay pre-booster, the RT-PCR test results pre-booster will be summarized.

In this study, the serology test results based on Roche Elecsys assay and the RT-PCR test results will be summarized by visit.

The primary analysis population to assess incidence of symptomatic SARS-CoV-2 infection (COVID-19), asymptomatic SARS-CoV-2 infection, and SARS-CoV-2 infection is PP Set for Efficacy, unless otherwise specified. mITT or FAS may be used for supportive analyses. All results will be summarized by study arm for each study part.

##### 6.5.1. Endpoint Definition/Derivation

###### 6.5.1.1. Derivation of SARS-CoV-2 Infection

SARS-CoV-2 infection is a combination of COVID-19 and asymptomatic SARS-CoV-2 infection. SARS-CoV-2 infection will be defined in participants by either:

- Participants with negative SARS-CoV-2 status at baseline and that becomes serology positive (as measured by *Roche Elecsys*) post baseline, OR
- Positive RT-PCR post baseline.

During the analysis, documented infection is counted starting 14 days after the dose of IP.

The date of documented infection will be the earlier of:

- Date of positive post-baseline RT-PCR result, or
- Date of positive serology test result based on bAb specific to SARS-CoV-2 nucleocapsid

The time to the first SARS-CoV-2 infection will be calculated as:

Time to the 1st SARS-CoV-2 infection = Date of the 1st documented infection – Date of injection + 1.

Cases will be counted starting 14 days after the injection, i.e. date of documented infection – Date of the injection ≥ 14.

SARS-CoV-2 infection cases will also be summarized based on tests performed at least 14 days after the dose of IP.

###### 6.5.1.2. Derivation of Asymptomatic SARS-CoV-2 Infection

This is an exploratory efficacy endpoint: the incidence of asymptomatic SARS-CoV-2 infection measured by RT-PCR of nasal swabs and/or serology tests obtained at post-baseline study visits counted starting 14 days after the injection.

Asymptomatic SARS-CoV-2 infection is identified by absence of symptoms and infections as detected by RT-PCR or serology tests. Specifically:

- Absent of COVID-19 symptoms

  AND at least one from below:

  - Participants with negative SARS-CoV-2 status at baseline and becomes positive (serology test result based on bAb specific to SARS-CoV-2 nucleocapsid protein) post baseline, when blood samples for immunogenicity are collected, or

  - Positive RT-PCR test at scheduled or unscheduled/illness visits

The date of documented asymptomatic infection is the earlier date of positive serology test result based on bAb specific to SARS-CoV-2 nucleocapsid due to infection, or positive RT-PCR , with absence of symptoms.

The time to the asymptomatic SARS-CoV-2 infection will be calculated as:

Time to the asymptomatic SARS-CoV-2 infection = Date of asymptomatic SARS-CoV-2 infection – Date of injection + 1.

###### 6.5.1.3.Derivation of Symptomatic SARS-CoV-2 Infection (COVID-19)

This is an exploratory efficacy endpoint: the incidence of the first occurrence of symptomatic SARS-CoV-2 infection measured by RT-PCR of nasal swabs counted starting 14 days after the injection. Surveillance for COVID-19 symptoms will be conducted via weekly contact and blood draw. Subjects reporting COVID-19 symptoms will be arranged an illness visit to collect an NP swab.

Two definitions of symptomatic SARS-CoV-2 Infection, COVID-19, will be evaluated:

1. Primary case definition per the P301 (COVE) study: Cases are defined as participants meeting clinical criteria based on both symptoms for COVID-19 and positive RT-PCR test results as described in Table 3-1.
2. Secondary case definition based on CDC criteria: Cases are defined as participants with symptomatic disease based on the criteria defined by the CDC (https://www.cdc.gov/coronavirus/2019-ncov/symptoms-testing/symptoms.html).

**Table 3-1. Derivation of primary case definition of COVID-19**

| **COVID-19 (per the P301 COVE study)** | |
|---|---|
| Post-baseline PCR results | Positive, **AND** |
| Systemic Symptoms | at least TWO of the following **systemic symptoms**: Fever (≥ 38ºC/≥ 100.4º F), chills, muscle and/or body aches (not related to exercise), headache, sore throat, new loss of taste/smell; **OR** |
| Respiratory Symptoms | at least **ONE** of the following **respiratory** signs/symptoms: cough, shortness of breath and/or difficulty breathing, OR clinical or radiographical evidence of pneumonia. |

**Table 3-2. Derivation for secondary case definition of COVID-19**

| **COVID-19 (CDC criteria)** | |
|---|---|
| Post-baseline PCR results | Positive, **AND** |
| Systemic and Respiratory Symptoms | at least ONE of the following **systemic or respiratory symptoms**: Fever (≥ 38ºC/≥ 100.4º F), chills, cough, shortness of breath and/or difficulty breathing, fatigue, muscle and/or body aches (not related to exercise), headache, new loss of taste/smell, sore throat, congestion, runny nose, nausea, vomiting, or diarrhea. |

The date of documented COVID-19 (case) will be the later date of eligible symptom and date of positive PCR test. Specifically, the date of documented COVID-19 will be the later date of the following two dates (date of positive PCR test, and the date of eligible symptom(s)), and the two dates should be within 14 days of each other.

The time to the first occurrence of COVID-19 will be calculated as:

Time to the 1st occurrence of COVID-19 = Date of documented COVID-19 – Date of injection + 1.

Cases will be counted starting 14 days after the injection, i.e. date of documented COVID-19 - Date of the injection ≥ 14.

##### 6.5.2. Analysis Method

The number and percentage of subjects who had each type of event (ie, an asymptomatic or a symptomatic SARS-CoV-2 infection) will be summarized in the PP Set for Efficacy.

The incidence rate of each type of event will be calculated as the number of cases divided by the total person-time. The 95% CI of the incidence rate will be calculated using the exact method (Poisson distribution) and adjusted by person-time.

Person-time is defined as the total time from injection date to the date of event, last date of study participation, censoring time, or efficacy data cutoff date, whichever is earlier.

##### 6.5.3. Sensitivity Analysis

Sensitivity analysis for the efficacy endpoints may be performed with the same methods described above based on the FAS or mITT Set and with cases counted starting at different time points.

##### 6.5.4. SARS-CoV-2 Exposure and Symptoms

SARS-CoV-2 reported exposure history and symptoms assessment will be assessed during the study.

The number and percentage of subjects who had close contact with a person with SARS-CoV-2 infection, reasons for exposure, subjects with any symptoms of potential

COVID-19, and subjects with each symptom will be presented by visit as defined in Section 6.1. Descriptive statistics will be provided for length of exposure in days.

In addition, the following listings will be provided for subjects infected by SARS-CoV-2:

- Serum bAb level against SARS-CoV-2
- Serum nAb titer against SARS-CoV-2

#### 6.6. Interim Analysis

The interim analysis will be conducted on safety and immunogenicity data collected through Day 15 or Day 29. The interim analysis may be performed either after all subjects in Part A.1, Part A.2, Part B, Part C, Part D, Part F, Part G, Part H or Part J have completed their Day 15 or Day 29 visit assessments and/or subsequent timepoint visits (eg, Day 91 for Parts F, G, H and J) or combined after the last subject of each study part (Parts A.1, A.2, B, C, D, F, G, H, or J) dose arm, or pre-specified subset of dose arm has completed their Day 15 or Day 29 visit assessments.

#### 6.7. Data Safety Monitoring Board

Not applicable.

#### 6.8. Final Analysis

The final analysis of all endpoints will be performed after all participants have completed all planned study procedures. Results of this analysis will be presented in a final CSR, including individual listings. The final CSR will include full analyses of all safety and immunogenicity through Day 366 (Month 12) for Parts A.1. B, C, D, F, and G and through Day 181 (Month 6) for Parts A.2, H and J.

### 7. References

Department of Health and Human Services (DHHS), Food and Drug Administration, Center for Biologics Evaluation and Research (US). Guidance for industry: Toxicity grading scale for healthy adult and adolescent volunteers enrolled in preventative vaccine clinical trials. September 2007 [cited 2019 Apr 10] [10 screens].

Available from:
https://www.fda.gov/downloads/BiologicsBloodVaccines/GuidanceComplianceRegulatoryInformation/Guidances/Vaccines/ucm091977.pdf.List of Appendices

IND 19745.291 Moderna COVID-19 Vaccine_CBER Comments

Rubin, D. B. (1987), Multiple Imputation for Nonresponse in Surveys, New York: John Wiley & Sons.

World Health Organization (WHO). Coronavirus disease 2019 (COVID-19) Weekly Epidemiological Update [Internet]. Geneva, Switzerland: WHO; 2021 May 9 [cited 2021 May 12]. Available from: https://www.who.int/publications/m/item/weekly-epidemiological-update-on-covid-19---11-may-2021.

### 8. List of Appendices

#### 8.1. Appendix A. Standards for Safety and Immunogenicity Variable Display in TFLs

**Continuous Variables**: The precision for continuous variables will be based on the precision of the data itself. The mean and median will be presented to one decimal place more than the original results; the SD will be presented to two decimal places more than the original results; the minimum and maximum will be presented to the same precision as the original results.

Antibody titer GMTs, and GMFRs will be presented to 1 decimal place, GMT ratio will be presented to three decimal places.

**Categorical Variables**: Percentages will be presented to 1 decimal place.

#### 8.2. Appendix B. Analysis Visit Windows for Safety and Immunogenicity Analysis

Safety and Immunogenicity Analysis will be summarized using the following analysis visit window for post injection assessments:

Step 1: If the safety and immunogenicity assessments are collected at scheduled visit, i.e. nominal scheduled visit, the data collected at scheduled visit will be used.

Step 2: If the safety and immunogenicity assessments are not collected at the scheduled visit, assessments collected at unscheduled visit will be used using the analysis visit windows described in Table 4 below.

If a subject has multiple assessments within the same analysis visit, the following rule will be used:

- If multiple assessments occur within a given analysis visit, the assessment closest to the target study day will be used.
- If there are 2 or more assessments equal distance to the target study day, the last assessment will be used.

**Table 4. Visit Window for Parts A.1, B, C, D, and E**

| Visit | Target Study Day | Visit Window in Study Day |
|---|---|---|
| **Nasopharyngeal Swabs for SARS-CoV-2** | | |
| Day 1 | 1 (Date of Injection) | 1, Pre-first-dose |
| Day 29 (Month 1) | 29 | [2, 105] |
| Day 181 (Month 6) | 181 | [106, 274] |
| Day 366 (Month 12) | 366 | ≥275 |
| **Vital Signs** | | |
| Day 1 | 1 (Date of First Injection) | ≤1, Pre-first-dose |
| Day 1 | 1 (Date of First Injection) | 1, Post-first-dose |
| Day 29 (Month 1) | 29 | [2, 105] |
| Day 181 (Month 6) | 181 | [106, 274] |
| Day 366 (Month 12) | 366 | ≥275 |
| **Immunogenicity** | | |
| Day 1 | 1 (Date of First Injection) | 1, Pre-first-dose |
| Day 15 | 15 | [2,22] |
| Day 29 (Month 1) | 29 | [23, 105] |
| Day 181 (Month 6) | 181 | [106, 274] |
| Day 366 (Month 12) | 366 | ≥275 |

**Table 5. Visit Window for Parts A.2, F, G, H and J**

| Visit | Target Study Day | Visit Window in Study Day |
|---|---|---|
| **Nasopharyngeal Swabs for SARS-CoV-2** | | |
| Day 1 | 1 (Date of Injection) | 1, Pre-first-dose |
| Day 29 (Month 1) | 29 | [2, 60] |
| Day 91 (Month 3) | 91 | [61, 136] |
| Day 181 (Month 6) | 181 | [137, 274] |
| Day 366 (Month 12)* | 366 | ≥275 |
| **Vital Signs** | | |
| Day 1 | 1 (Date of First Injection) | ≤1, Pre-first-dose |
| Day 1 | 1 (Date of First Injection) | 1, Post-first-dose |
| Day 29 (Month 1) | 29 | [2, 60] |
| Day 91 (Month 3) | 91 | [61, 136] |
| Day 181 (Month 6) | 181 | [137, 274] |
| Day 366 (Month 12)* | 366 | ≥275 |
| **Immunogenicity** | | |
| Day 1 | 1 (Date of First Injection) | 1, Pre-first-dose |
| Day 15 | 15 | [2,22] |
| Day 29 (Month 1) | 29 | [23, 60] |
| Day 91 (Month 3) | 91 | [61, 136] |
| Day 181 (Month 6) | 181 | [137, 274] |
| Day 366 (Month 12)* | 366 | ≥275 |

*Part A.2, Part H and Part J will not have Day 366 visit.

#### 8.3. Appendix C. Imputation Rules for Missing Prior/Concomitant Medications and Non-Study Vaccinations

Imputation rules for missing or partial medication start/stop dates are defined below:

1. Missing or partial medication start date:

- If only Day is missing, use the first day of the month, unless:
  - o The medication end date is after the date of the injection or is missing AND the start month and year of the medication coincide with the start month and year of the injection. In this case, use the date of the injection
- If Day and Month are both missing, use the first day of the year, unless:
  - o The medication end date is after the date of the injection or is missing AND the start year of the medication coincide with the start year of the injection. In this case, use the date of the injection
- If Day, Month and Year are all missing, the date will not be imputed, but the medication will be treated as though it began prior to the injection for purposes of determining if status as prior or concomitant.

2. Missing or partial medication stop date:

- If only Day is missing, use the earliest date of (last day of the month, study completion, discontinuation from the study, or death).
- If Day and Month are both missing, use the earliest date of (last day of the year, study completion, discontinuation from the study, or death).
- If Day, Month and Year are all missing, the date will not be imputed, but the medication will be flagged as a continuing medication.

##### 8.4. Appendix D. Imputation Rules for Missing AE dates

Imputation rules for missing or partial AE start dates and stop dates are defined below:

1. Missing or partial AE start date:

- If only Day is missing, use the first day of the month, unless:

    - The AE end date is after the date of the injection or is missing AND the start month and year of the AE coincide with the start month and year of the injection. In this case, use the date and time of the injection, even if time is collected.
- If Day and Month are both missing, use the first day of the year, unless:
    - The AE end date is after the date of the injection or is missing AND the start year of the AE coincides with the start year of the injection. In this case, use the date of the injection
- If Day, Month and Year are all missing, the date will not be imputed. However, if the AE end date is prior to the date of the injection, then the AE will be considered a pre-treatment AE. Otherwise, the AE will be considered treatment-emergent.

2. Missing or partial AE end dates will not be imputed.

#### 8.5. Appendix E. Immunogenicity Endpoints by Study Part

| **Endpoint** | **Study Part** | **Booster** | **Comparison group*** | **GMT or SRR Comparison Based on Antibody Titer** |
|---|---|---|---|---|
| Primary | A.1 | 50 µg or 100 µg mRNA-1273.211 | 100 µg mRNA-1273 historical control | booster against ancestral SARS-CoV-2 (D29 post-booster) vs. primary series against ancestral SARS-CoV-2 (D57)** |
| | | 50 µg or 100 µg mRNA-1273.211 | 100 µg mRNA-1273 historical control | booster against B.1.351 (D29) vs. primary series against ancestral SARS-CoV-2 (D57) |
| | A.2 | 50 µg mRNA-1273.214 | 50 µg mRNA-1273.211 (Part A.1) | within participant booster comparison against ancestral SARS-CoV-2 and SARS-CoV-2 variants (D29 and D181) |

| Endpoint | Study Part | Booster | Comparison group* | GMT or SRR Comparison Based on Antibody Titer |
|---|---|---|---|---|
| | B | 100 µg mRNA-1273 | 100 µg mRNA-1273 historical control | booster against ancestral SARS-CoV-2 (D29) vs. primary series against ancestral SARS-CoV-2 (D57) |
| | C | 50 µg or 100 µg mRNA-1273.617.2 | 100 µg mRNA-1273 historical control | booster against B.1.617.2 (D29) vs. primary series against ancestral SARS-CoV-2 (D57) |
| | D | 50 µg or 100 µg mRNA-1273.213 | 100 µg mRNA-1273 historical control | booster against B.1.617.2 (D29) vs. primary series against ancestral SARS-CoV-2 (D57) |
| | | 50 µg or 100 µg mRNA-1273.213 | 100 µg mRNA-1273 historical control | booster against B.1.351 (D29) vs. primary series against ancestral SARS-CoV-2 (D57) |
| | F, Cohort 1 | 50 µg mRNA-1273.529 | 50 µg mRNA-1273 historical booster control | booster against B.1.1.529 (D29) vs. booster control against B.1.1.529 (D29) |
| | F, Cohort 2 | 50 µg mRNA-1273.529 | 50 µg mRNA-1273 | between booster comparison against B.1.1.529 (D29) |
| | G | 50 µg mRNA-1273.214 | 50 µg mRNA-1273 (Part F Cohort 2) | between booster comparison against B.1.1.529 (D29 and D91) |
| | | 50 µg mRNA-1273.214 | 50 µg mRNA-1273 (Part F Cohort 2) | between booster comparison against ancestral SARS-CoV-2 (D29 and D91) |
| | H | 50 µg mRNA-1273.222 | 50 µg mRNA-1273 (Part F Cohort 2) | between booster comparison against BA.4/5 (D29) |
| | H | 50 µg mRNA-1273.222 | 50 µg mRNA-1273 (Part F Cohort 2) | between booster comparison against ancestral SARS-CoV-2 (D29) |
| | J | 50 µg mRNA-1273.231 | 50 µg mRNA-1273.815 | between booster comparison against SARS-CoV-2 Omicron BA.4/BA.5 (D15 and D29) |
| | | 50 µg mRNA-1273.231 | 50 µg mRNA-1273.815 | between booster comparison against BQ.1.1 (D15 and D29) |

| Endpoint | Study Part | Booster | Comparison group* | GMT or SRR Comparison Based on Antibody Titer |
|---|---|---|---|---|
| | | 50 µg mRNA-1273.231 | 50 µg mRNA-1273.815 | between booster comparison against XBB.1.5 (D15 and D29) |
| Secondary | A.1 | 50 µg or 100 ug mRNA-1273.211 | 100 µg mRNA-1273 historical control | booster against variant (D29) vs. primary series against ancestral SARS-CoV-2 (D57) |
| | A.2 | 50 µg mRNA-1273.214 | 50 µg mRNA-1273.211 (Part A.1) | within participant booster comparison against ancestral SARS-CoV-2 and SARS-CoV-2 variants (multiple time points) |
| | B | 100 µg mRNA-1273 | 100 µg mRNA-1273 historical control | booster against variant (D29) vs. primary series against ancestral SARS-CoV-2 (D57) |
| | C | 50 µg or 100 ug mRNA-1273.617.2 | 100 µg mRNA-1273 historical control | booster against ancestral SARS-CoV-2 and variant (D29) vs. primary series against ancestral SARS-CoV-2 (D57) |
| | D | 50 µg or 100 ug mRNA-1273.213 | 100 µg mRNA-1273 historical control | booster against ancestral SARS-CoV-2 and variant (D29) vs. primary series against ancestral SARS-CoV-2 (D57) |
| | F, Cohort 1 | 50 µg mRNA-1273.529 | 50 µg mRNA-1273 historical booster control | booster vs. booster control against B.1.1.529 (D29) |
| | | 50 µg mRNA-1273.529 | 50 µg mRNA-1273 historical booster control | booster vs. booster control against B.1.1.529 (all timepoints) |
| | | 50 µg mRNA-1273.529 | 50 µg mRNA-1273 historical booster control | booster vs. booster control against ancestral SARS-CoV-2/other variants (all timepoints) |
| | | 50 µg mRNA-1273.529 | 100 µg mRNA-1273 historical control | booster against B.1.1.529 (D29) vs. primary series against ancestral SARS-CoV-2 (D57) |

| Endpoint | Study Part | Booster | Comparison group* | GMT or SRR Comparison Based on Antibody Titer |
|---|---|---|---|---|
| | F, Cohort 2 | 50 µg mRNA-1273.529 | 50 µg mRNA-1273 | between booster comparison against B.1.1.529 (D29) |
| | | 50 µg mRNA-1273.529 | 50 µg mRNA-1273 | between booster comparison against B.1.1.529 (all timepoints) |
| | | 50 µg mRNA-1273.529 | 50 µg mRNA-1273 | between booster comparison against ancestral SARS-CoV-2/other variants (all timepoints) |
| | | 50 µg mRNA-1273.529 | 100 µg mRNA-1273 historical control | booster against B.1.1.529 (D29) vs. primary series against ancestral SARS-CoV-2 (D57) |
| | G | 50 µg mRNA-1273.214 | 50 µg mRNA-1273 (Part F Cohort 2) | between booster comparison against ancestral SARS-CoV-2 (D29 and D91) |
| | | 50 µg mRNA-1273.214 | 50 µg mRNA-1273 (Part F Cohort 2) | between booster comparison against B.1.1.529 (all timepoints) |
| | | 50 µg mRNA-1273.214 | 50 µg mRNA-1273 (Part F Cohort 2) | between booster comparison against ancestral SARS-CoV-2/other variants (all timepoints) |
| | | 50 µg mRNA-1273.214 | 100 µg mRNA-1273 historical control | booster against B.1.1.529 (D29) vs. primary series against ancestral SARS-CoV-2 (D57) |
| | H | 50 µg mRNA-1273.222 | 50 µg mRNA-1273 (Part F Cohort 2) | between booster comparison against ancestral SARS-CoV-2/other variants (all timepoints) |
| Exploratory | A.1 | 50 ug mRNA-1273.211 | 100 µg mRNA-1273.211 | between booster comparison against same strains (ancestral SARS-CoV-2 and variants) |
| | | 50 ug or 100 µg mRNA-1273.211 | 100 µg mRNA-1273 historical control | booster against variant (D29) vs. mRNA-1273 against variant (D57) |

| Endpoint | Study Part | Booster | Comparison group* | GMT or SRR Comparison Based on Antibody Titer |
|---|---|---|---|---|
| | B | 100 ug mRNA-1273 | 50 ug or 100 ug mRNA-1273.211 | between booster comparison against same strains (ancestral SARS-CoV-2 and variants) |
| | | 100 ug mRNA-1273 | 100 µg mRNA-1273 historical control | booster against variant (D29) vs. mRNA-1273 against variant (D57) |
| | C | 50 µg mRNA-1273.617.2 | 100 µg mRNA-1273. 617.2 | between booster comparison against same strains (ancestral SARS-CoV-2 and variants) |
| | | 50 µg or 100 µg mRNA-1273.617.2 | 50 µg or 100 µg mRNA-1273.211 | between booster comparison against same strains (ancestral SARS-CoV-2 and variants) |
| | | 50 µg or 100 µg mRNA-1273.617.2 | 100 µg mRNA-1273 | between booster comparison against same strains (ancestral SARS-CoV-2 and variants) |
| | | 50 µg or 100 µg mRNA-1273.617.2 | 100 µg mRNA-1273 historical control | booster against variant (D29) vs. primary series against variant (D57) |
| | D | 50 ug mRNA-1273.213 | 100 ug mRNA-1273.213 | between booster comparison against same strains (ancestral SARS-CoV-2 and variants) |
| | | 50 ug or 100 ug mRNA-1273.213 | 50 µg or 100 µg mRNA-1273.211 | between booster comparison against same strains (ancestral SARS-CoV-2 and variants) |
| | | 50 ug or 100 ug mRNA-1273.213 | 100 µg mRNA-1273 | between booster comparison against same strains (ancestral SARS-CoV-2 and variants) |
| | | 50 ug or 100 ug mRNA-1273.213 | 50 µg or 100 µg mRNA-1273.617.2 | between booster comparison against same strains (ancestral SARS-CoV-2 and variants) |

| Endpoint | Study Part | Booster | Comparison group* | GMT or SRR Comparison Based on Antibody Titer |
|---|---|---|---|---|
| | | 50 ug or 100 ug mRNA-1273.213 | 100 µg mRNA-1273 historical control | booster against variant (D29) vs. primary series against variant (D57) |
| | J | 50 µg mRNA-1273.231 | 50 µg mRNA-1273.815 | between booster comparison against same strains (SARS-CoV-2 variants) |

*Historical control arm from study mRNA-1273-P301 (100 µg mRNA-1273). Historical booster control arm from study mRNA-1273-P201 (50 µg mRNA-1273).
**D57 refers to post dose 1 of primary series.

#### 8.6. Appendix F. Summary of Major Changes in Previous Versions of SAP

<u>Summary of major changes in SAP Version 2.0</u>

| Section | Brief Description of Changes | Rationale |
|---|---|---|
| 2.1 (Primary Objectives), 2.2 (Secondary Objectives), 2.3 (Exploratory Objectives) | Added objectives for Part C, Part D, and Part E. | Updated to align with protocol amendment 3. |
| 2.1 (Primary Objectives), 6.4.3 (Primary Analysis of Antibody-Mediated Immunogenicity Endpoints) | Removed GMT ratio ≥1 requirement for 50 µg dose arm. | The point estimator of GMT ratio ≥1 for the 50 µg booster dose of the mRNA-1273 vaccines was considered prior to interim analyses of the 50 µg mRNA-1273 booster dose interim analyses from another mRNA-1273 clinical study (P201). These results demonstrated non-inferior immune responses, after the 50 µg booster dose, compared to the mRNA-1273 primary |

| Section | Brief Description of Changes | Rationale |
|---|---|---|
| | | series and supported the emergency use authorization of the 50 µg mRNA-1273 booster dose. Therefore, the additional point estimator criterion for the 50 µg dosed is now removed from the statistical analysis plan of the P205 study. |
| 3.1 (Primary Endpoints), 3.2 (Secondary Endpoints), 3.3 (Exploratory Endpoints) | Added endpoints for Part C, Part D, and Part E. | Updated to align with protocol amendment 3. |
| 4.1 (Overall Study Design), 4.3 (Sample Size and Power) | Added new arm and study parts. | Updated to align with protocol amendment 3. |
| 4.2 (Statistical Hypotheses), 6.4.3 (Primary Analysis of Antibody-Mediated Immunogenicity Endpoints) | For study parts with both, 50 µg and 100 µg doses, removed testing sequence. | Results from another mRNA-1273 clinical study (P201) demonstrated non-inferior immune responses, after the 50 µg mRNA-1273 booster dose, compared to the mRNA-1273 primary series and supported the emergency use authorization of the 50 µg mRNA-1273 booster dose. Therefore, the 50 µg booster dose can be evaluated independently from the 100 µg dose, without multiplicity adjustment for hypotheses |

| Section | Brief Description of Changes | Rationale |
|---|---|---|
| | | testing for these two dose arms. |
| 6.4.3 (Primary Analysis of Antibody-Mediated Immunogenicity Endpoints), 6.4.5 (Exploratory Analysis of Antibody-Mediated Immunogenicity Endpoints) | Added analysis for Part C, Part D, and Part E. | Updated to align with protocol amendment 3. |
| 6.4.7 (Seroresponse) | Added seroresponse definition based on pre-dose 1 of primary series. | Added a seroresponse definition to calculate seroresponse rate based on both, pre-dose 1 of primary series and pre-booster dose antibody titers. |

<u>Summary of major changes in SAP Version 3.0</u>

| Section | Brief Description of Changes | Rationale |
|---|---|---|
| 2.1 (Primary Objectives), 2.2 (Secondary Objectives), 2.3 (Exploratory Objectives) | Added objectives for Part F, Part G, and Part A.2. | Updated according to protocol amendment 7. |
| 3.1 (Primary Endpoints), 3.2 (Secondary Endpoints), 3.3 (Exploratory Endpoints) | Added endpoints for Part F, Part G, and Part A.2. | Updated according to protocol amendment 7. |
| 4.1 (Overall Study Design), 4.2 (Statistical Hypotheses), 4.3 (Sample Size and Power) | Added details for Part F, Part G, and Part A.2. | Updated according to protocol amendment 7. |

| | | |
|---|---|---|
| 6.4.3 (Primary Analysis of Antibody-Mediated Immunogenicity Endpoints), 6.4.5 (Exploratory Analysis of Antibody-Mediated Immunogenicity Endpoints) | Added analysis for Part F, Part G, and Part A.2. | Updated according to protocol amendment 7. |
| 6.4.6 (Sensitivity Analysis) | Added sensitivity analyses 1): To assess impact of positive SARS-COV-2 infection that occurs during study to antibody titer data<br><br>2): Added Multiple imputation method to impute for missing antibody titer for primary endpoint if percentage of missing value at analysis visit is more than 10% | 1) Natural immunity can also increase antibody titer level, excluding SARS-CoV-2 infection up to the analysis visit to assess treatment effect without the impact of infection on immune response<br>2) Per FDA recent feedback (IND 19745.291) |
| 6.4.7 Seroresponse | Clarify for Part F cohort 2, Part G, seroresponse will be based on pre-vaccination baseline. | |

Summary of major changes in SAP Version 4.0

| Section | Brief Description of Changes | Rationale |
|---|---|---|

| | | |
|---|---|---|
| 6.1 (General consideration) | If a participant received off-study COVID-19 vaccine, the participant's immune response and effectiveness data will be censored at the earlier date of date of off-study COVID-19 vaccine, database lock/data snapshot or data cutoff date, or date of last observed in study | Once a participant receive additional off-study COVID-19 booster dose, this participant immunogenicity and infection data is no longer a pure response of on-study COVID-19 booster dose, records post additional COVID-19 booster dose need to be excluded. |

Summary of major changes in SAP Version 5.0

| Section | Brief Description of Changes | Rationale |
|---|---|---|
| 2.1 (Primary Objectives), 2.2 (Secondary Objectives), 2.3 (Exploratory Objectives) | Added objectives for Part H. | Updated according to protocol amendment 9. |
| 3.1 (Primary Endpoints), 3.2 (Secondary Endpoints), 3.3 (Exploratory Endpoints) | Added endpoints for Part H. | Updated according to protocol amendment 9. |
| 4.1 (Overall Study Design), 4.2 (Statistical Hypotheses), 4.3 (Sample Size and Power) | Added details for Part H. | Updated according to protocol amendment 9. |
| 6.4.3 (Primary Analysis of Antibody-Mediated | Revised the GMR non-inferiority boundary for all | Updated according to protocol amendment 9. |

| | | |
|---|---|---|
| Immunogenicity Endpoints) | study parts, from ≥ 0.67 to > 0.667. | |
| 6.4.3 (Primary Analysis of Antibody-Mediated Immunogenicity Endpoints), 6.4.4 (Secondary Analysis of Antibody-Mediated Immunogenicity Endpoints) | Added analysis for Part H. | Updated according to protocol amendment 9. |
| 6.4.7 Seroresponse | Added evaluation of seroresponse will be based on both definitions for all study parts. | Updated according to protocol amendment 9. |
