## Supplementary Appendix for "Safety and Immunogenicity of XBB.1.5-Containing mRNA Vaccines"

**Supplement**

**List of Investigators**

| Investigator | Institution | Location |
| --- | --- | --- |
| Adam Brosz | Meridian Clinical Research | Grand Island, Nebraska |
| Laurence Chu | Benchmark Research | Austin, Texas |
| Michael Cotugno | Benchmark Research | Metairie, Louisiana |
| Brandon Essink | Meridian Clinical Research | Omaha, Nebraska |
| Carlos Fierro | Johnson County Clin-Trials | Lenexa, Kansas |
| Carl Griffin | Lynn Health Science Institute | Oklahoma City, Oklahoma |
| Greg Hachigian | Benchmark Research | Sacramento, California |
| Charles Harper. Keith Vrbicky, | Meridian Clinical Research | Norfolk, Nebraska |
| Jordan Whatley | Meridian Clinical Research | Baton Rouge, Louisiana |

**Immunogenicity Assays**

***Lentivirus-Based SARS-CoV-2 Spike-Pseudotyped Virus Neutralization Assay***

SARS-CoV-2 neutralizing antibodies (nAb) in samples were assessed using a SARS-CoV-2 Spike (S)-Pseudotyped Virus Neutralization Assay (PsVNA) in 293/ACE2 cells.[1] The PsVNA quantifies nAb using lentivirus particles that express full-length spike proteins on their surface, and contain a firefly luciferase reporter gene for quantitative measurements of infection in transduced 293T cells expressing high levels of ACE2 (293T/ACE2 cells) by relative luminescence units (RLU). Serial dilution of antibodies was used to produce a dose-response curve. Neutralization was measured as the serum dilution at which RLU was reduced by 50% (ID50) relative to mean RLU in virus control wells (cells + virus but no sample) after subtraction of mean RLU in cell control wells (cells only). Positive controls were included on each assay plate in order to follow stability over time. The assay is validated for the prototypic D614G variant and the omicron BA.5/BA.5 sublineage but has not been validated for the omicron BQ.1.1 and XBB.1 sublineages.

The spike-pseudotyped viruses were derived from ancestral Wuhan-Hu-1 with the following amino acid substitutions: (prototype [D614G]; omicron subvariants BA.4 and BA.5, designated BA.4/BA.5 for identical spike sequences between BA.4 and BA.5 [T19I, L24S, ΔP25, ΔP26, ΔA27, G142D, V213G, G339D, S371F, S373P, S375F, T376A, D405N, R408S, K417N, N440K, L452R, S477N, T478K, E484A, F486V, Q498R, N501Y, Y505H, D614G, H655Y, N679K, P681G, N764K, D796Y, Q954H, N969K]); omicron subvariant BQ.1.1 [T19I, L24S, ΔP25, ΔP26, ΔA27, H69-, V70-, G142D, V213G, G339D, R346T, S371F, S373P, S375F, T376A, D405N, R408S, K417N, N440K, K444T, L452R, N460K, S477N, T478K, E484A, F486V, Q498R, N501Y, Y505H, D614G, H655Y, N679K, P681H, N764K, D796Y, Q954H, N969K]; omicron subvariant XBB.1.5 [T19I, L24S, ΔP25, ΔP26, ΔA27, V83A, G142D, Y145Q, ΔH146, Q183E, V213E, G252V, G339H, R346T, L368I, S371F, S373P, S375F, T376A, D405N, R408S, K417N, N440K, V445P, G446S, N460K, S477N, T478K, E484A, F486P, F490S, Q498R, N501Y, Y505H, D614G, H655Y, N679K, P681H, N764K, D796Y, Q954H, N969K]; omicron subvariant XBB.1.16 [T19I, L24S, ΔP25, ΔP26, ΔA27, V83A, G142D, Y145Q, ΔH146, E180V, Q183E, V213E, G252V, G339H, R346T, L368I, S371F, S373P, S375F, T376A, D405N, R408S, K417N, N440K, V445P, G446S, N460K, S477N, T478R, E484A, F486P, F490S, Q498R, N501Y, Y505H, D614G, H655Y, N679K, P681H, N764K, D796Y, Q954H, N969K].

***VSV-based SARS-CoV-2 Pseudovirus Neutralization Assay***

Codon-optimized full-length spike genes (Wuhan-1 with D614G, BA.4/BA.5, XBB.1.5, XBB.1.16, XBB.2.3.2, EG.5.1, FL.1.5.1, BA.2.86) were cloned into a pCAGGS vector. Spike genes contained the following mutations: (a) BA.4/BA.5: T19I, L24- P25- P26-, A27S H69- V70-, G142D, V213G, G339D, S371F, S373P, S375F, T376A, D405N, R408S, K417N, N440K, L452R, S477N, T478K, E484A, F486V, Q498R, N501Y, Y505H, D614G, H655Y, N679K, P681H, N764K, D796Y, Q954H, N969K; (b) XBB.1.5: T19I, L24-, P25-, P26-, A27S, V83A, G142D, Y144-, H146Q, Q183E, V213E, G252V, G339H, R346T, L368I, S371F, S373P, S375F, T376A, D405N, R408S, K417N, N440K, V445P, G446S, N460K, S477N, T478K, E484A, F486P, F490S, Q498R, N501Y, Y505H, D614G, H655Y, N679K, P681H, N764K, D796Y, Q954H, N969K; (c) XBB.1.16: T19I, L24-, P25-, P26-, A27S, V83A, G142D, Y144-, H146Q, E180V, Q183E, V213E, G252V, G339H, R346T, L368I, S371F, S373P, S375F, T376A, D405N, R408S, K417N, N440K, V445P, G446S, N460K, S477N, T478R, E484A, F486P, F490S, Q498R, N501Y, Y505H, D614G, H655Y, N679K, P681H, N764K, D796Y, Q954H, N969K; (d) XBB.2.3.2: T19I, L24-, P25-, P26-, A27S, V83A, G142D, Y144-, H146Q, Q183E, G184V, V213E, D253G, G339H, R346T, L368I, S371F, S373P, S375F, T376A, D405N, R408S, K417N, N440K, V445P, G446S, N460K, S477N, T478K, E484A, F486P, F490S, Q498R, N501Y, Y505H, P521S, D614G, H655Y, N679K, P681H, N764K, D796Y, Q954H, N969K; (e) EG.5.1: T19I L24- P25- P26- A27S Q52H V83A G142D Y144- H146Q Q183E V213E G252V G339H R346T L368I S371F S373P S375F T376A D405N R408S K417N N440K V445P G446S F456L N460K S477N T478K E484A F486P F490S Q498R N501Y Y505H D614G H655Y N679K P681H N764K D796Y Q954H N969K; (f) FL.1.5.1: T19I L24- P25- P26- A27S V83A G142D Y144- H146Q Q183E V213E G252V G339H R346T L368I S371F S373P S375F T376A D405N R408S K417N N440K V445P G446S F456L N460K S477N T478R E484A F486P F490S Q498R N501Y Y505H D614G H655Y N679K P681H A701V N764K D796Y Q954H N969K; BA.2.86: ins16MPLF T19I R21T L24- P25- P26- A27S S50L H69- V70- V127F G142D Y144- F157S R158G N211- L212I V213G L216F H245N A264D I332V G339H K356T S371F S373P S375F T376A R403K D405N R408S K417N N440K V445H G446S N450D L452W N460K S477N T478K N481K V483- E484K F486P Q498R N501Y Y505H E554K A570V D614G P621S H655Y N679K P681R N764K D796Y S939F Q954H N969K P1143L. To generate VSVΔG-based SARS-CoV-2 pseudovirus, BHK-21/WI-2 cells were transfected with the spike expression plasmid and infected by VSVΔG-firefly-luciferase as previously described (Whitt 2010 Journal of Virological Methods; PMID 20709108). Vero E6 cells (ATCC, CRL-1586) were used as target cells for the neutralization assay and maintained in DMEM supplemented with 10% fetal bovine serum. To perform neutralization assay, serum samples were heat-inactivated for 45 min at 56ºC, and serial dilutions were made in DMEM supplemented with 10% FBS. The diluted serum samples or culture medium (serving as virus only control) were mixed with VSVΔG-based SARS-CoV-2 pseudovirus and incubated at 37ºC for 45 min. The inoculum virus or virus-serum mix was subsequently used to infect Vero E6 cells for 18 h at 37ºC. At 18 h post infection, an equal volume of One-Glo reagent (Promega; E6120) was added to culture medium for readout using BMG PHERAstar FSX plate reader. The percentage of neutralization was calculated based on relative light units of the virus control, and subsequently analyzed using four parameter logistic curve (GraphPad Prism 8.0).

### Figure S1. Distribution of Neutralizing Antibody Titers Against XBB.1.5, XBB.1.16, BA.4/BA.5 and BQ.1.1 Variants and Ancestral SARS-CoV-2 (D614G)

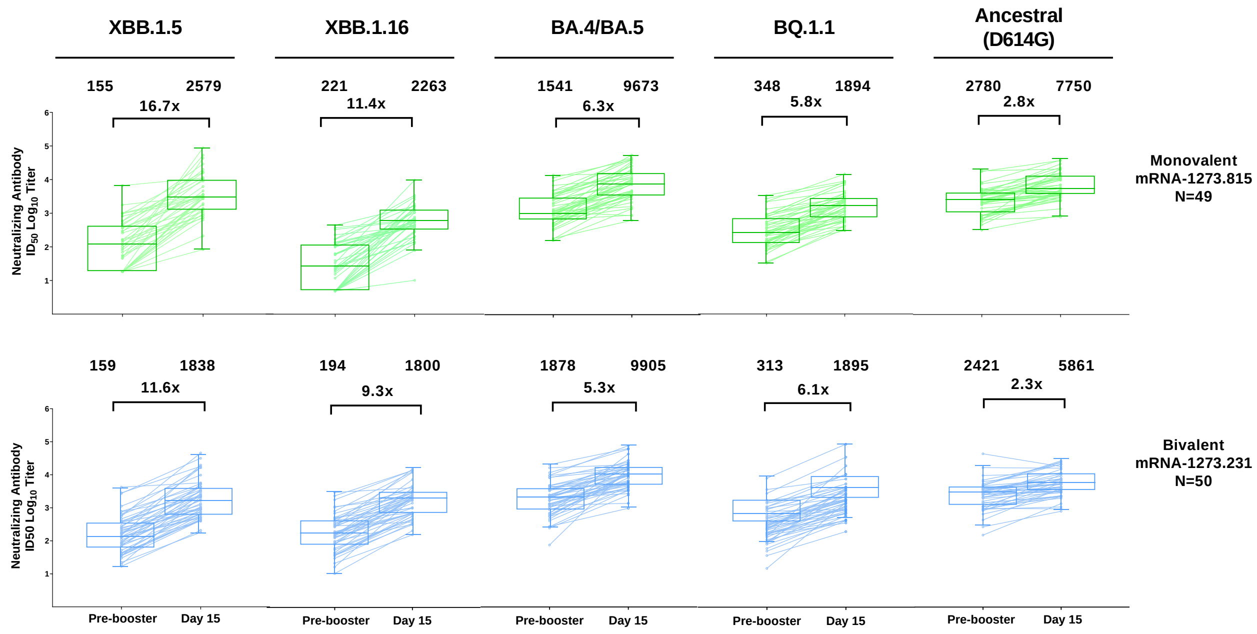


**Figure S1.** Distribution of neutralizing antibody titers (ID50 $\log_{10}$) in Figure 1 detected in the pseudovirus assay against omicron XBB.1.5, XBB.1.16, BA.4/BA.5, BQ.1.1 and ancestral SARS-CoV-2 (D614G) is shown for serum samples collected before the booster dose of 50-µg of monovalent mRNA-1273.815 or 50-µg of bivalent mRNA-1273.231, and at 15 days after the booster dose (Day 15) in the per-protocol set for immunogenicity (with and without prior SARS-CoV-2 infection). The circles are values from individual serum samples. Pseudovirus neutralizing antibody assay lower limits of quantification (LLOQ) are 18.5 (1.3 $\log_{10}$) for ancestral SARS-CoV-2 (D614G) and 36.7 (1.6 $\log_{10}$) for omicron BA.4/BA.5; upper limits of quantification (ULOQ) are 45,118 (4.7 $\log_{10}$) for ancestral SARS-CoV-2 (D614G) and 13,705 (4.1 $\log_{10}$) for omicron BA.4/BA.5. The Limits of Detection are 10 for XBB.1.5, XBB.1.16 and BQ.1.1. Boxes and horizontal bars denote interquartile ranges (IQR) and median endpoint titers; whisker endpoints are the maximum and minimum values below or above the median ±1.5 times the IQR.

**Table S1. Neutralizing Antibodies Titers and Geometric-fold Rises After 3rd Booster Dose of XBB.1.5-Containing Monovalent and Bivalent Vaccines Against XBB.1.5, XBB.1.16, BA.4/BA.5 and BQ.1.1 Variants and Ancestral SARS-CoV-2 (D614G), Per-protocol Immunogenicity Set**

| | Monovalent mRNA-1273.815 (XBB.1.5) 50 µg | Bivalent mRNA-1273.231 (XBB.1.5 + BA.4/5) 50 µg |
|---|---|---|
| | N=49 | N=50 |
| **XBB.1.5** | | |
| Pre-booster, n | 49 | 50 |
| GM Level (95% CI) | 154.7 (106.8-224.1) | 158.8 (109.9-229.3) |
| Day 15, n | 49 | 50 |
| GMT (95% CI) | 2579.0 (1809.1-3676.7) | 1838.1 (1265.9-2668.9) |
| GMFR (95% CI) | 16.7 (12.8-21.7) | 11.6 (8.7-15.4) |
| **XBB.1.16** | | |
| Pre-booster, n | 47 | 50 |
| GM Level (95% CI) | 221.0 (153.9-317.3) | 193.9 (134.1-280.3) |
| Day 15, n | 47 | 50 |
| GM Level (95% CI) | 2262.6 (1570.1-3260.6) | 1799.9 (1297.2-2497.5) |
| GMFR (95% CI) | 11.4 (8.5-15.4) | 9.3 (7.0-12.3) |
| **BA.4/BA.5** | | |
| Pre-booster, n | 49 | 50 |
| GM Level (95% CI) | 1540.7 (1127.2-2105.8) | 1878.1 (1350.9-2611.1) |
| Day 15, n | 49 | 50 |
| GM Level (95% CI) | 9673.4 (6965.6-13433.8) | 9904.8 (7610.8-12890.1) |
| GMFR (95% CI) | 6.3 (4.8, 8.2) | 5.3 (3.9-7.1) |
| **BQ.1.1** | | |
| Pre-booster, n | 48 | 50 |
| GM Level (95% CI) | 347.5 (249.5-483.9) | 312.8 (221.4-441.9) |
| Day 15, n | 48 | 50 |
| GM Level (95% CI) | 1894.1 (1383.2, 2593.6) | 1895.4 (1348.2-2664.7) |
| GMFR (95% CI) | 5.8 (4.7-7.3) | 6.1 (4.6-7.9) |
| **Ancestral SARS-CoV-2 (D614G)** | | |
| Pre-booster, n | 49 | 50 |
| GM Level (95% CI) | 2780.3 (2146.5-3601.3) | 2421.3 (1788.4-3278.1) |
| Day 15, n | 49 | 49 |
| GM Level (95% CI) | 7749.7 (5943.7-10104.3) | 5860.9 (4558.0-7536.3) |
| GMFR (95% CI) | 2.8 (2.2-3.5) | 2.3 (1.9-2.8) |

n = number of observations at corresponding timepoint; GMT = geometric mean titer; 95% CI = 95% confidence interval; GMFR = geometric mean fold rise. 95% CI is calculated based on the t-distribution of the log-transformed values or the difference in the log-transformed values for GM value and GM fold-rise, respectively, then back transformed to the original scale for presentation. GMFR is GM Level at Day 15 relative to GM Level at pre-booster. Antibody values reported as below the lower limit of quantification (LLOQ: 36.7 for BA.4/BA.5; 18.5 for ancestral SARS-CoV-2 (D614G)) are replaced by 0.5 x LLOQ. Values greater than the upper limit of quantification (ULOQ: 13,705 for BA.4/BA.5; 45,188 for Ancestral SARS-CoV-2 (D614G)) are replaced by the ULOQ if actual values are not available. Antibody values reported as below the lower limit of detection (LOD: 10 for XBB.1.5; 10 for XBB.1.16; 10 for BQ.1.1) are replaced by 0.5 x LOD.

**Table S2. Neutralizing Antibodies Titers and Geometric-fold Rises After 3rd Booster Dose of XBB.1.5-Containing Monovalent and Bivalent Vaccines Against XBB.1.5, XBB.1.16, BA.4/BA.5 and BQ.1.1 Variants and Ancestral SARS-CoV-2 (D614G) by Pre-booster SARS-CoV-2 Status**

| | No Prior Infection | | Prior Infection | |
|---|---|---|---|---|
| | Monovalent mRNA-1273.815 (XBB.1.5) 50 µg | Bivalent mRNA-1273.231 (XBB.1.5 + BA.4/5) 50 µg | Monovalent mRNA-1273.815 (XBB.1.5) 50 µg | Bivalent mRNA-1273.231 (XBB.1.5 + BA.4/5) 50 µg |
| | N=16 | N=11 | N=33 | N=39 |
| **XBB.1.5** | | | | |
| Pre-booster, n | 16 | 11 | 33 | 39 |
| GM Level (95% CI) | 81.7 (40.7-164.1) | 65.2 (33.8-125.6) | 210.8 (138.8-320.3) | 204.1 (135.3-307.9) |
| Day 15, n | 16 | 11 | 33 | 39 |
| GMT (95% CI) | 1639.3 (742.9-3617.4) | 1256.9 (537.5-2939.4) | 3212.8 (2215.2-4659.5) | 2046.1 (1333.2-3140.1) |
| GMFR (95% CI) | 20.1 (12.5-32.1) | 19.3 (8.5-43.8) | 15.2 (10.9-21.2) | 10.0 (7.5-13.5) |
| **XBB.1.16** | | | | |
| Pre-booster, n | 14 | 11 | 33 | 39 |
| GM Level (95% CI) | 133.7 (67.9-263.2) | 71.6 (33.9-150.8) | 273.5 (178.0-420.1) | 256.9 (173.3-380.7) |
| Day 15, n | 16 | 11 | 33 | 39 |
| GM Level (95% CI) | 1409.5 (646.1-3074.8) | 1452.8 (618.3-3413.6) | 2846.2 (1912.7-4235.4) | 1912.0 (1326.8-2755.2) |
| GMFR (95% CI) | 14.2 (8.5-24.0) | 20.3 (9.6-43.0) | 10.4 (7.2-15.1) | 7.4 (5.6-9.8) |
| **BA.4/BA.5** | | | | |
| Pre-booster, n | 16 | 11 | 33 | 39 |
| GM Level (95% CI) | 691.3 (438.9-1088.7) | 844.4 (405.9-1756.9) | 2272.3 (1606.0-3215.0) | 2353.1 (1653.9-3347.9) |
| Day 15, n | 16 | 11 | 33 | 39 |
| GM Level (95% CI) | 6254.4 (3482.3-11233.1) | 9269.3 (6282.1-13676.8) | 11951.0 (8035.2-17775.2) | 10091.8 (7276.3-13996.6) |
| GMFR (95% CI) | 9.0 (5.9-13.9) | 11.0 (4.5-27.0) | 5.3 (3.7-7.4) | 4.3 (3.3-5.6) |
| **BQ.1.1** | | | | |
| Pre-booster, n | 15 | 11 | 33 | 39 |
| GM Level (95% CI) | 247.7 (125.7-488.2) | 142.2 (71.7-281.9) | 405.3 (275.9-595.3) | 390.7 (266.1-573.5) |
| Day 15, n | 16 | 11 | 33 | 39 |
| GM Level (95% CI) | 1438.3 (125.7-488.2) | 1376.1 (877.5-2157.8) | 2164.5 (1554.6-3013.6) | 2074.6 (1360.0-3164.5) |
| GMFR (95% CI) | 7.1 (4.6-11.1) | 9.7 (4.5-20.9) | 5.3 (4.1-7.0) | 5.3 (4.0-7.0) |
| **Ancestral SARS-CoV-2 (D614G)** | | | | |
| Pre-booster, n | 16 | 11 | 33 | 39 |
| GM Level (95% CI) | 2179.9 (1205.3-3942.6) | 1772.2 (820.7-3827.2) | 3128.4 (2380.6-4111.1) | 2644.0 (1886.7-3705.3) |
| Day 15, n | 16 | 10 | 33 | 39 |
| GM Level (95% CI) | 7743.5 (4611.2-13003.4) | 6226.5 (3849.4-10071.4) | 7752.7 (5605.8-10721.9) | 5770.7 (4272.7-7793.9) |
| GMFR (95% CI) | 3.6 (2.1-5.9) | 2.8 (1.6-4.7) | 2.5 (2.0-3.1) | 2.2 (1.7-2.7) |

n = number of observations at corresponding timepoint; GM = geometric mean; 95% CI = 95% confidence interval; GMFR = geometric mean fold rise. 95% CI is calculated based on the t-distribution of the log-transformed values or the difference in the log-transformed values for GM value and GM fold-rise, respectively, then back transformed to the original scale for presentation. GMFR is GM Level at Day 15 relative to GM Level at pre-booster. Antibody values reported as below the lower limit of quantification (LLOQ: 36.7 for BA.4/BA.5; 18.5 for ancestral SARS-CoV-2 [D614G]) are replaced by 0.5 x LLOQ. Values greater than the upper limit of quantification (ULOQ: 13,705 for BA.4/BA.5; 45,188 for Ancestral SARS-CoV-2 [D614G]) are replaced by the ULOQ if actual values are not available. Antibody values reported as below the lower limit of detection (LOD: 10 for XBB.1.5; 10 for XBB.1.16; 10 for BQ.1.1) are replaced by 0.5 x LOD.

**Table S3. Solicited Local and Systemic Adverse Reactions (Solicited Safety Set)**

| Solicited Adverse Reaction | Monovalent mRNA-1273.815 (XBB.1.5) 50 µg (N=50) | Bivalent mRNA-1273.231 (XBB.1.5 + BA.4/5) 50 µg (N=51) |
|---|---|---|
| Solicited Adverse Reactions - N1 | 50 | 51 |
| Any Solicited | 38 (76.0) | 45 (88.2) |
| Grade 1 | 23 (46.0) | 28 (54.9) |
| Grade 2 | 14 (28.0) | 11 (21.6) |
| Grade 3 | 1 (2.0) | 6 (11.8) |
| Solicited Local Adverse Reactions - N1 | 50 | 51 |
| Any Local | 34 (68.0) | 43 (84.3) |
| Grade 1 | 26 (52.0) | 33 (64.7) |
| Grade 2 | 8 (16.0) | 7 (13.7) |
| Grade 3 | 0 | 3 (5.9) |
| Pain - N1 | 50 | 51 |
| Any | 34 (68.0) | 42 (82.4) |
| Grade 1 | 28 (56.0) | 33 (64.7) |
| Grade 2 | 6 (12.0) | 7 (13.7) |
| Grade 3 | 0 | 2 (3.9) |
| Erythema (Redness) - N1 | 50 | 51 |
| Any | 2 (4.0) | 1 (2.0) |
| Grade 1 | 1 (2.0) | 0 |
| Grade 2 | 1 (2.0) | 0 |
| Grade 3 | 0 | 1 (2.0) |
| Swelling (Hardness) - N1 | 50 | 51 |
| Any | 5 (10.0) | 6 (11.8) |
| Grade 1 | 2 (4.0) | 3 (5.9) |
| Grade 2 | 3 (6.0) | 2 (3.9) |
| Grade 3 | 0 | 1 (2.0) |
| Axillary Swelling or Tenderness - N1 | 50 | 51 |
| Any | 8 (16.0) | 14 (27.5) |
| Grade 1 | 8 (16.0) | 12 (23.5) |
| Grade 2 | 0 | 2 (3.9) |
| Grade 3 | 0 | 0 |
| Solicited Systemic Adverse Reactions - N1 | 50 | 51 |
| Any Systemic | 29 (58.0) | 33 (64.7) |
| Grade 1 | 16 (32.0) | 17 (33.3) |
| Grade 2 | 12 (24.0) | 12 (23.5) |
| Grade 3 | 1 (2.0) | 4 (7.8) |
| Fever - N1 | 50 | 51 |
| Any | 3 (6.0) | 1 (2.0) |
| Grade 1 | 1 (2.0) | 1 (2.0) |
| Grade 2 | 1 (2.0) | 0 |
| Grade 3 | 1 (2.0) | 0 |
| Headache - N1 | 50 | 51 |
| Any | 17 (34.0) | 23 (45.1) |
| Grade 1 | 14 (28.0) | 17 (33.3) |
| Grade 2 | 3 (6.0) | 5 (9.8) |
| Grade 3 | 0 | 1 (2.0) |
| Fatigue - N1 | 50 | 51 |
| Any | 22 (44.0) | 25 (49.0) |
| Grade 1 | 12 (24.0) | 14 (27.5) |
| Grade 2 | 10 (20.0) | 9 (17.6) |
| Grade 3 | 0 | 2 (3.9) |
| Myalgia - N1 | 50 | 51 |
| Any | 19 (38.0) | 19 (37.3) |
| Grade 1 | 14 (28.0) | 11 (21.6) |

| | | |
|---|---|---|
| Grade 2 | 5 (10.0) | 6 (11.8) |
| Grade 3 | 0 | 2 (3.9) |
| Arthralgia - N1 | 50 | 51 |
| Any | 14 (28.0) | 15 (29.4) |
| Grade 1 | 12 (24.0) | 8 (15.7) |
| Grade 2 | 2 (4.0) | 6 (11.8) |
| Grade 3 | 0 | 1 (2.0) |
| Nausea/Vomiting - N1 | 50 | 51 |
| Any | 4 (8.0) | 5 (9.8) |
| Grade 1 | 4 (8.0) | 3 (5.9) |
| Grade 2 | 0 | 2 (3.9) |
| Grade 3 | 0 | 0 |
| Chills - N1 | 50 | 51 |
| Any | 7 (14.0) | 12 (23.5) |
| Grade 1 | 5 (10.0) | 9 (17.6) |
| Grade 2 | 2 (4.0) | 3 (5.9) |
| Grade 3 | 0 | 0 |

N1=Number of exposed participants with data for the event. Any = Grade 1 or higher. Percentages are based on the number of exposed participants who submitted any data for the event (N1). Toxicity grade for Erythema (Redness) is defined as: G1 = 25 — 50 mm; G2 = 51 — 100 mm; G3 = > 100 mm. Toxicity grade for Fever is defined as: G1 = 38 — 38.4 C; G2 = 38.5 — 38.9 C; G3 = 39 — 40 C; G4 = > 40 C.

**Table S4. Unsolicited Treatment-emergent Adverse Reactions (Safety Set)**

| n (%) | Monovalent mRNA-1273.815 (XBB.1.5) 50 µg (N=50) | Bivalent mRNA-1273.231 (XBB.1.5 + BA.4/BA.5) 50 µg (N=51) |
|---|---|---|
| Unsolicited AEs Regardless of Relationship to Study Vaccination | | |
| All | 5 (10.0) | 7 (13.7) |
| Serious | 0 | 0 |
| Fatal | 0 | 0 |
| Medically-Attended | 4 (8.0) | 4 (7.8) |
| Unsolicited AEs Related to Study Vaccination | | |
| All | 1 (2.0) | 2 (3.9) |
| Serious | 0 | 0 |
| Fatal | 0 | 0 |
| Medically-Attended | 1 (2.0) | 0 |

AEs = adverse reactions. AE=adverse event. An adverse event was defined as any event not present prior to study vaccination or any event already present that worsened in intensity or frequency after vaccination. Percentages are based on the number of participants in the safety set.
